# A Worldwide Study of White Matter Microstructural Alterations in People Living with Parkinson’s Disease

**DOI:** 10.1101/2024.01.16.24301235

**Authors:** Conor Owens-Walton, Talia M. Nir, Sarah Al-Bachari, Sonia Ambrogi, Tim J. Anderson, Ítalo Karmann Aventurato, Fernando Cendes, Yao-Liang Chen, Valentina Ciullo, Phil Cook, John C. Dalrymple-Alford, Michiel F. Dirkx, Jason Druzgal, Hedley C. A. Emsley, Rachel Guimarães, Hamied A. Haroon, Rick C. Helmich, Michele T. Hu, Martin E. Johansson, Ho Bin Kim, Johannes C. Klein, Max Laansma, Katherine E. Lawrence, Christine Lochner, Clare Mackay, Corey McMillan, Tracy R. Melzer, Leila Nabulsi, Ben Newman, Peter Opriessnig, Laura M. Parkes, Clelia Pellicano, Fabrizio Piras, Federica Piras, Lukas Pirpamer, Toni L. Pitcher, Kathleen L. Poston, Annerine Roos, Lucas Scárdua Silva, Reinhold Schmidt, Petra Schwingenschuh, Marian Shahid, Gianfranco Spalletta, Dan J. Stein, Sophia I. Thomopoulos, Duygu Tosun, Chih-Chien Tsai, Odile A. van den Heuvel, Eva van Heese, Daniela Vecchio, Julio E. Villalón-Reina, Chris Vriend, Jiun-Jie Wang, Yih-Ru Wu, Clarissa Lin Yasuda, Paul M. Thompson, Neda Jahanshad, Ysbrand van der Werf

## Abstract

**Background:** The progression of Parkinson’s disease (PD) is associated with microstructural alterations in neural pathways, contributing to both motor and cognitive decline. However, conflicting findings have emerged due to the use of heterogeneous methods in small studies, particularly regarding the involvement of white matter (WM) tracts. Here we performed the largest diffusion MRI study of PD to date, integrating data from 17 cohorts worldwide, to identify stage-specific profiles of WM differences.

**Methods:** Diffusion-weighted MRI data from 1,654 participants diagnosed with PD (age range: 20-89 years; 33% female) and 885 controls (age range: 19-84 years; 47% female) were analyzed using the ENIGMA-DTI protocol to evaluate regional microstructure in 21 white matter regions. Skeletonized maps of diffusion tensor imaging fractional anisotropy (FA) and mean diffusivity (MD) were analyzed and compared between Hoehn and Yahr (HY) disease groups and controls to reveal the profile of white matter differences at different stages.

**Results:** We found an enhanced, more widespread pattern of microstructural differences with each stage of PD, with eventually lower FA and higher MD in almost all regions of interest (ROIs): Cohen’s *d* effect sizes reached *d*=-1.01 for FA differences in the fornix by PD HY Stage 4/5. The early PD signature in HY stages 1 and 2 included higher FA and lower MD across the entire white matter skeleton, in a direction opposite to that typical of other neurodegenerative diseases. FA and MD were associated with clinical metrics of motor and non-motor clinical dysfunction.

**Conclusion:** While overridden by degenerative changes in the later stages of PD, early PD is associated with paradoxically higher FA in PD, which is consistent with early compensatory changes associated with the disorder.

## Introduction

Parkinson’s disease (PD) is the second most common neurodegenerative disorder, impacting millions of people worldwide, contributing to significant disability and death in people diagnosed with the condition (Dorsey et al., 2018; Feigin et al., 2019). PD is characterized by motor symptoms of bradykinesia, tremor at rest or postural rigidity (Postuma et al., 2015). Non-motor symptoms include neuropsychiatric and cognitive impairment, disruptions of sleep-wake cycle regulation, autonomic dysfunction, sensory disturbances and pain (Schapira et al., 2017). Our recently published study - the largest neuroimaging study of PD - mapped cortical gray matter thinning and subcortical volume deficits across PD disease progression stages, revealing a pattern of cortical thinning, closely tracking Braak stages of Lewy body pathology in PD (Laansma et al., 2021). Diffusion MRI (dMRI) is primarily used to investigate pathophysiological processes that impact white matter in the brain, enabling an investigation of the brain circuits that are impacted in the condition. The most common dMRI model used to study white matter alterations in PD is diffusion tensor imaging (DTI) which characterizes the three dimensional diffusion of water as a function of spatial location: the diffusion tensor describes the magnitude, orientation and degree of diffusion anisotropy (Alexander et al., 2007). The current study investigates the two most commonly used DTI parameters of fractional anisotropy (FA) and mean diffusivity (MD). Typically, high FA and low MD are associated with structurally intact WM pathways, while the opposite is found in damaged WM.

Reviews and literature-based meta-analyses of individual studies report a heterogeneous, multifocal pattern of DTI alterations in PD, with higher FA and/or lower MD found in motor pathways soon after the disease is diagnosed, but lower FA and/or higher MD in frontal, temporal and callosal regions at advanced disease stages (Atkinson-Clement, C et al., 2017; Wei et al., 2021; Zhang & Burock, 2020). Individual studies often have smaller sample sizes comprised of individuals from similar demographics, and may include participants from various disease stages, making it difficult to assess whether the findings generalize to other populations. Here, we aim to overcome these limitations by conducting a coordinated analysis of dMRI across many international studies, using standardized and validated image analysis and quality control protocols. Using the largest sample size generated in this field of research, we set out to investigate how microstructural brain measures are impacted at different stages along the PD disease spectrum, and how WM alterations are associated with clinical symptoms. We hypothesized that participants with PD would show more pronounced DTI WM alterations at advanced disease stages. At the onset of PD, male patients typically show more bradykinesia and rigidity, while female patients tend to display greater signs of tremor (Haaxma et al., 2007). Due to the slower progression of PD in females (Marras & Saunders-Pullman, 2014), along with research suggesting that males demonstrate greater local WM disruptions (Tremblay et al., 2020), we also hypothesized that female PD participants would have less severe alterations in DTI measures than their male counterparts. Finally, we hypothesized that lower microstructural integrity would be associated with poorer motor and cognitive function.

## Methods

### Study Participants

To enhance statistical power and generalizability of the findings, we coordinated a worldwide, pooled, multisite analysis of data from 17 cohorts from Africa, Asia, Europe, Oceania, and both North and South America (**Supplementary Figure 1**). In total, dMRI scans from 1,654 participants with PD and 885 controls were analyzed (age range: 20-89 years; 38% female).

Information on how each cohort diagnosed PD, as well as inclusion and exclusion criteria, is available in **Supplementary Methods 1.1**. Fourteen of the seventeen cohorts collected scans and clinical information from age-matched control participants. Demographic and clinical characteristics of each cohort are summarized in **Table 1**; the distribution of age across sites is shown in **Figure 1**.

**Table 1:**
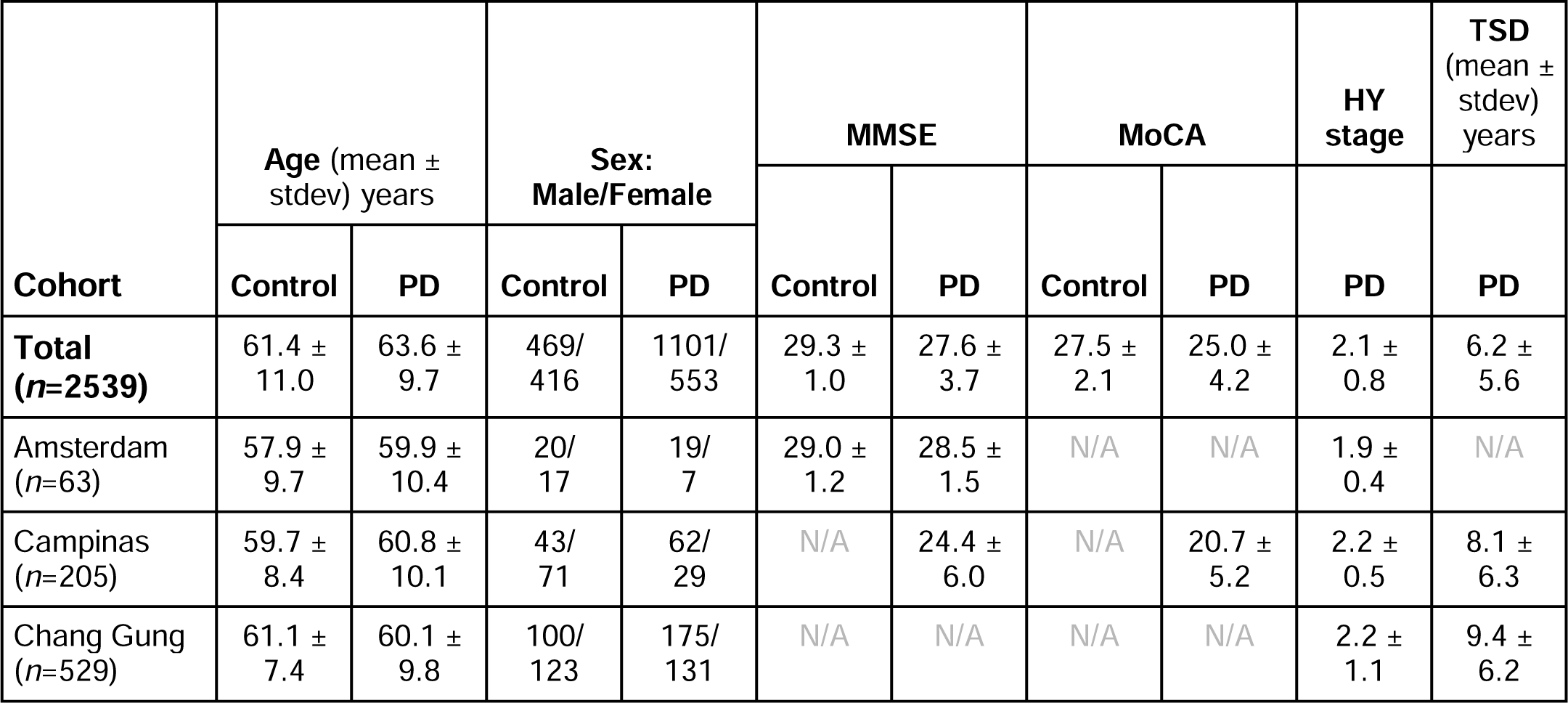
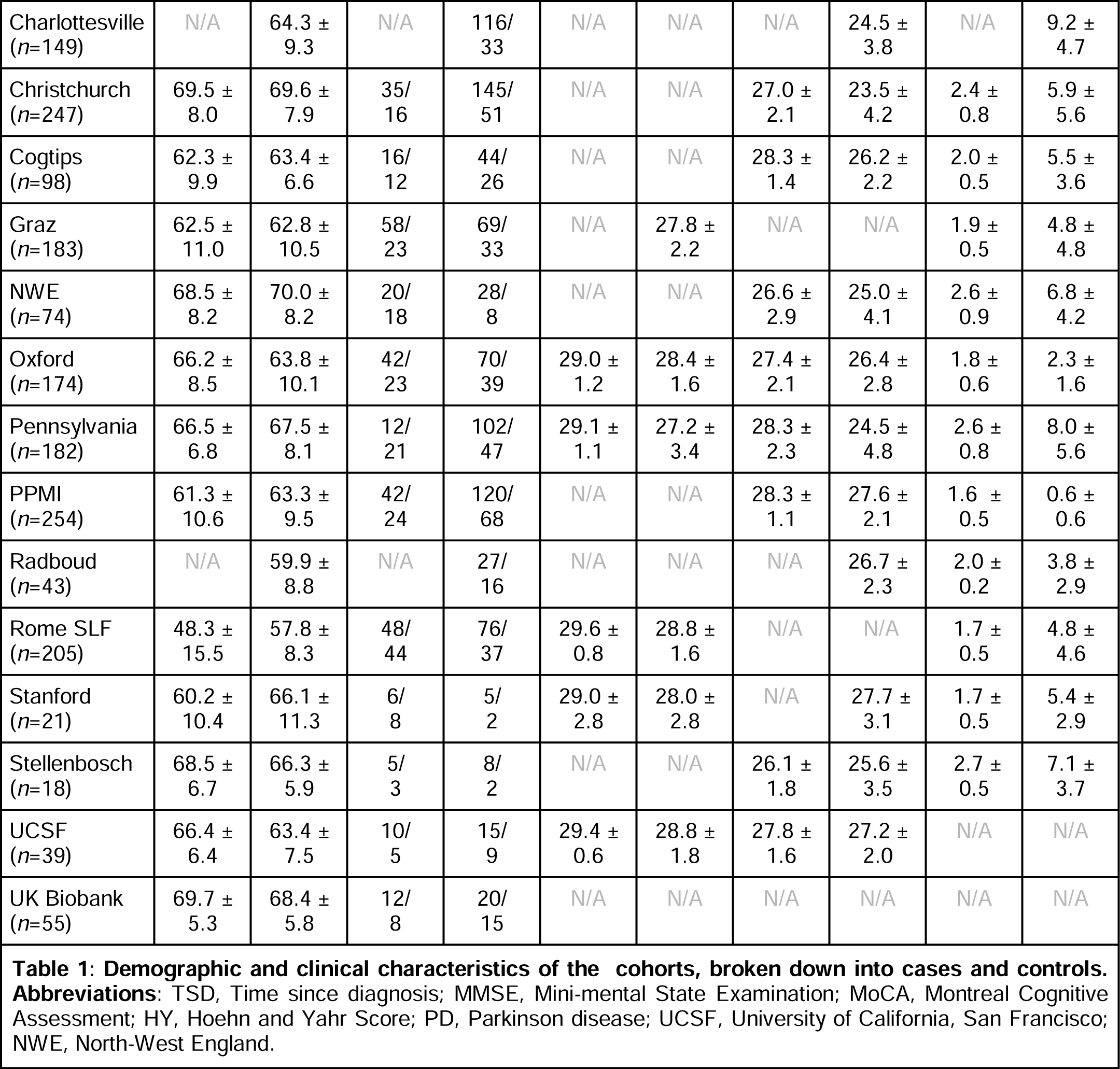
ENIGMA-PD Sample Characteristics by Site.

**Figure 1:**
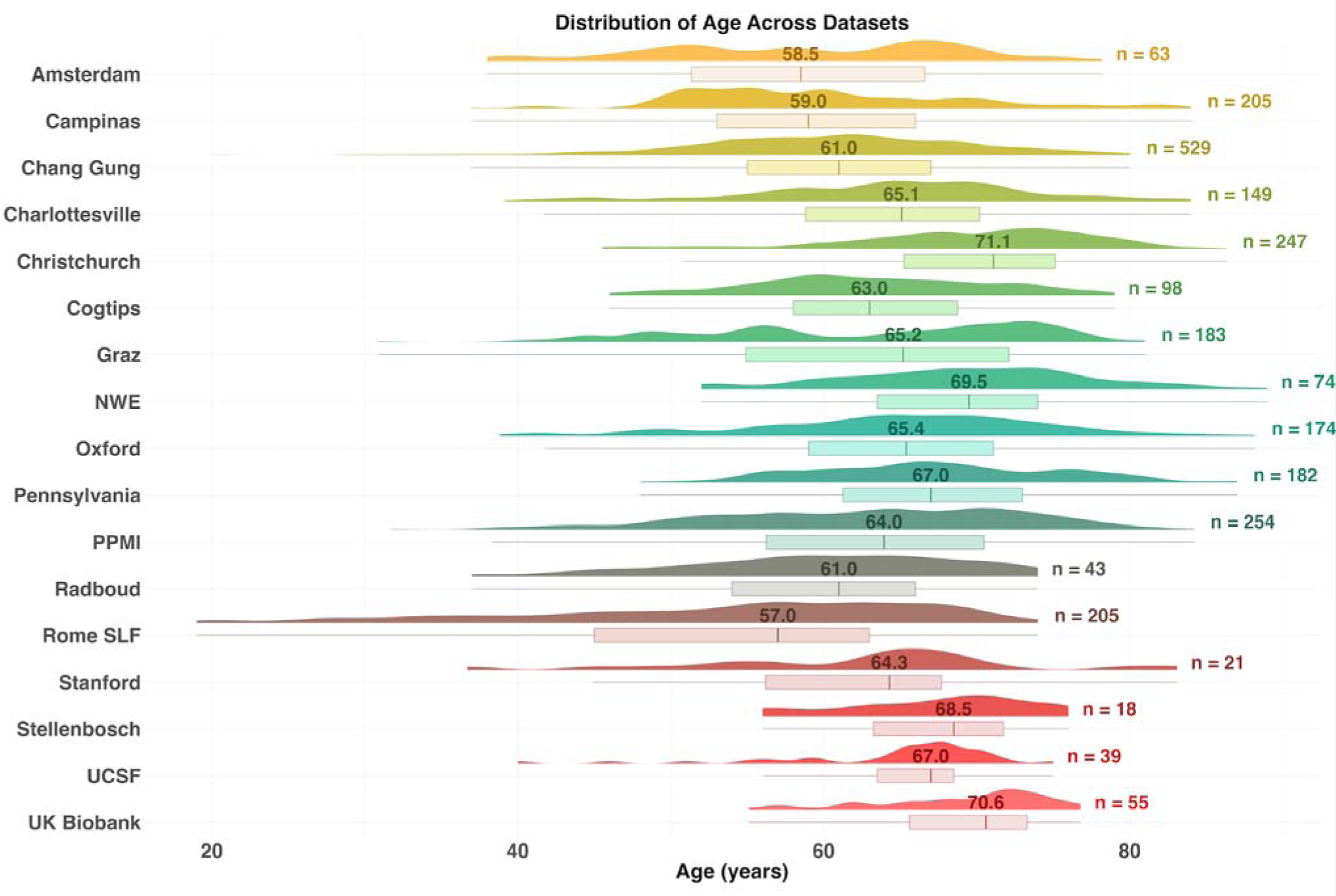
Age distribution for the 17 datasets we analyzed. The number above the boxplot is the median age for each sample and to the right is the total number subjects.

Clinical assessments included the Hoehn and Yahr (HY) stage (Hoehn & Yahr, 1967), Mini-Mental State Examination (Folstein et al., 1975), Montreal Cognitive Assessment (Nasreddine et al., 2005), and the UPDRS Part-III test (Fahn & Elton, 1987). Participants were assessed with either the original UPDRS Part III (original UPDRS-III) or revised Movement Disorder Society UPDRS Part III scores. We therefore used a validated formula to convert original UPDRS-III scores to predicted MDS-UPDRS-III scores (MDS-UPDRS-III) **(Supplementary Methods 1.2**). To harmonize HY staging across cohorts, HY1.5 and HY2.5 increments were reclassified as HY2. HY5 scores were grouped with HY4 to maintain a sufficiently large sample for the analysis. These groupings align with our consortium’s published international study using standard anatomical MRI (Laansma et al., 2021). All participants provided written informed consent; each local study was approved by institutional review boards, and all research was performed in accordance with the World Medical Association’s Declaration of Helsinki.

Demographic and clinical characteristics of each HY disease stage group are shown in **Table 2**.

**Table 2:** Characteristics of the PD HY Cohorts, Stratified by HY Disease Stage.

| PD HY Stage | Age | Sex: M/F | MMSE | MoCA | AAO | DD | MDS-UPDRS -III (ON) | MDS-UPDRS -III (OFF) |
| --- | --- | --- | --- | --- | --- | --- | --- | --- |
| All HY (n=1312) | 63.5 ±<br>9.8 | 858/454 | 27.5 ±<br>3.8 | 25.1 ±<br>4.2 | 57.8 ±<br>10.9 | 5.8 ±<br>5.6 | 27.5 ±<br>14.4 | 26.8 ±<br>15.4 |
| HY1<br>(n=275) | 59.3 ±<br>10.0 | 170/105 | 27.9 ±<br>3.9 | 26.5 ±<br>3.2 | 56.5 ±<br>10.4 | 2.8 ±<br>2.8 | 14.4 ±<br>7.2 | 15.6 ±<br>8.2 |
| HY2<br>(n=742) | 63.0 ±<br>9.2 | 504/238 | 27.6 ±<br>3.7 | 25.6 ±<br>3.7 | 59.0 ±<br>10.3 | 5.0 ±<br>4.8 | 27.8 ±<br>11.4 | 26.0 ±<br>10.7 |
| HY3<br>(n=220) | 66.2 ±<br>10.1 | 142/78 | 27.0 ±<br>3.4 | 23.9 ±<br>4.0 | 56.5 ±<br>12.6 | 9.5 ±<br>6.5 | 39.4 ±<br>15.8 | 31.6 ±<br>13.5 |
| HY4/5<br>(n=75) | 67.1 ±<br>9.6 | 42/33 | 24.5 ±<br>5.1 | 18.2 ±<br>6.4 | 54.1 ±<br>11.8 | 13.2 ±<br>5.9 | 57.5 ±<br>13.5 | 56.8 ±<br>16.6 |

### MRI Acquisition

All cohorts’ scanner descriptions and acquisition protocols are provided in **Supplementary Methods Table 1.3**. Prior work by the ENIGMA Worldwide Consortium in epilepsy, brain trauma, PTSD, major depression, bipolar disorder, obsessive-compulsive disorder, and schizophrenia (Dennis et al., 2021; Favre et al., 2019; Hatton et al., 2020; Kelly et al., 2018, 2018; Piras et al., 2021; Van Velzen et al., 2020) has shown that DTI measures can be merged across scanning protocols, provided that standardized processing procedures are used as detailed below alongside statistical adjustments.

### Image Processing

Each cohort conducted dMRI processing locally, or sent unprocessed imaging data to the central site for analysis after appropriate data transfer agreements were approved by the relevant institutions. Preprocessing steps included image denoising, Gibbs deringing correction, eddy current correction, echo-planar imaging induced distortion and B1 field inhomogeneity correction. All data was then processed using the standardized ENIGMA-DTI pipeline (Jahanshad et al., 2013). A tensor estimation step was performed for each participant, creating FA, MD, axial diffusivity (AD) and radial diffusivity (RD) maps. FA images were visually checked to ensure that the principal eigenvector direction was anatomically plausible. DTI FA maps were non-linearly registered to the ENIGMA-DTI template and alignment was visually checked. Tract-based spatial-statistics (TBSS) (Smith et al., 2006) was then used to skeletonize FA, MD, AD and RD maps. Mean DTI measures were extracted from 21 WM tracts based on the Johns Hopkins University atlas of WM labels (herein referred to as regions of interest (ROI) (Hua et al., 2008). An overall average value was also derived for the whole brain WM skeleton (entire WM) assessing the mean of left and right DTI measures for bilateral ROIs. The final step in the ENIGMA-DTI pipeline produces ROI data for each participant; this was then sent to the central analysis site for data curation and statistical analyses. The main text of this paper presents results from FA and MD only - the two most widely reported DTI measures in the literature. AD and RD results are presented in the **Supplementary Results**.

### Between-group Differences in White Matter Microstructural Measures

#### Stratification by HY Stage

To investigate DTI alterations at different stages of PD, mixed effects linear regression models were fitted to evaluate regional DTI differences at each HY subgroup with controls. Fixed effect covariates included age, sex, the square of the mean-centered age, and the mean-centered interaction of age with sex. To account for differences in dMRI acquisitions across contributing groups, ‘cohort’ was used as a random-effect covariate, as in prior multi-site morphometric ENIGMA studies (Radua et al., 2020).

Effect sizes were estimated as Cohen’s *d* values as described in **Supplementary Methods 1.4.** To adjust for multiple comparisons, we used the false discovery rate (FDR) procedure to correct raw *p*-values, with a *q*=0.05 (Benjamini & Hochberg, 1995). Statistical analyses were run using R software (v4.0.3) with *lme4* (v1.1.30) and MASS (v7.3.53) packages. To control the experiment-wise false positive rate and limit false positive inferences across the whole study, we considered the FA and MD analyses as primary, while the clinical association tests were treated as *post hoc* secondary analyses.

#### Matched Control Samples

To ensure that findings were not driven by age or sex differences in case-control sample comparisons, all HY PD subgroup analyses were repeated using subsamples of age- and sex-matched controls, generated using 1:1 nearest-neighbor matching with the *MatchIt* package (v4.5.0) in R.

#### Associations Between Effect Sizes

To evaluate the similarity between results generated using ‘Stratification by HY Stage’ and ‘Matched Control Samples’ designs, we tested correlations between the Cohen’s *d* values generated for each ROI across these two analyses.

#### Entire PD Cohort and Controls

Mixed effects linear regression models were also fitted to evaluate regional DTI differences between the overall PD cohort and controls.

#### Sex-by-Diagnosis Interactions

To examine sex differences in the effect of PD on DTI measures, sex-by-diagnosis interactions were also investigated, in groups stratified by HY PD stage. These analyses used mixed-effects linear regression models, with fixed-effect covariates including age, sex, the square of mean-centered measure of age, and the mean-centered interaction of age with sex, with ‘cohort’ modeled as a random effect.

#### Random-effects Meta-Analysis: PD and Controls

Differences in DTI measures between PD participants and controls were also assessed using random-effects meta-analysis (RE-Meta), to ensure that pooled analysis findings were not driven by any single site (**Supplementary Results 2.6**). Within each of the 15 out of 17 cohorts that had both PD participants and controls, linear regressions were used, adjusting for age, sex, the square of the mean-centered age, and the mean-centered interaction of age with sex. Cohen’s *d* effect sizes and standard errors were then analyzed for each ROI across site via meta-analysis using the *Metafor* package (v3.8.1) in R.

### Associations Between Clinical Assessments and Diffusion Measures in PD

We tested for associations between regional DTI measures and (1) MoCA scores, and (2) MDS-UPDRS-III scores in the entire PD cohort (MDS-UPDRS-III scores were only considered when the participant was in the functional OFF-state for their Parkinsonian medication at the time of clinical assessment). Fixed-effect covariates included disease duration, age, sex, the square of mean-centered measure of age and the mean-centered age interaction with sex, with ‘cohort’ used as the random-effect covariate. Effect sizes were estimated using partial correlation coefficient *r* values (**Supplementary Methods 1.4**).

## Results

There were significant differences in age, MMSE and MoCA between PD HY subgroups and controls, and in the proportions of males and females across PD HY subgroups and controls. There were also significant differences in these measures when comparing only the PD HY subgroup participants (**Supplementary Methods 1.5**). Control participants were significantly younger than the entire PD cohort by 2.2 years. In particular, the Rome cohort collected data from controls who were significantly younger than PD participants (*p*<0.001). As expected, the entire PD cohort had lower scores on the MMSE and MoCA tests than controls. There was also a significant difference in the proportions of males and females across the two groups (**Supplementary Methods 1.6)**. Two cohorts, Charlottesville and Radboud, did not collect control data. Because of these age differences, below we report results comparing PD patients to all controls, as well as to only age- and sex-matched controls.

### Between-group Differences in White Matter Microstructural Measures

#### FA: Stratification by HY Stage

Pronounced patterns of FA differences emerged when stratifying PD participants according to HY stage. HY1 participants (*n*=275) had *higher* FA across the entire WM skeleton (*d*=0.30) as well as 4 out of 21 ROIs, compared to controls (*n*=885), with effect sizes of *d*=0.18-0.19 (all *p*_FDR_<0.05). Implicated regions included the anterior *corona radiata*, anterior and retrolenticular parts of the internal capsule, and the genu of the corpus callosum. HY2 PD participants (*n*=742) had *lower* FA at the *fornix* (*d*=-0.26) compared to controls, while HY3 PD participants (*n*=220) had *lower* FA across the entire WM skeleton (*d*=-0.24), as well as 9 out of 21 ROIs:the largest effect was found for the *fornix* (*d*=-0.33). Implicated regions include the anterior and posterior *corona radiata*, posterior thalamic radiation, genu of the corpus callosum, external capsule, *fornix*/*stria terminalis*, superior longitudinal fasciculus and the sagittal *stratum*. HY4/5 PD participants (*n*=75) had *lower* FA across the entire WM skeleton (*d*=-0.74), as well as 20 out of 21 ROIs relative to controls (*n*=75), with the largest effect in the *fornix* (*d*=-1.01) and remaining values increasing in magnitude from *d*=-0.26. The only ROI not implicated in HY4/5 was the cingulum (hippocampal portion) (**Figure 2; Supplementary Results 2.1.1**).

**Figure 2:**
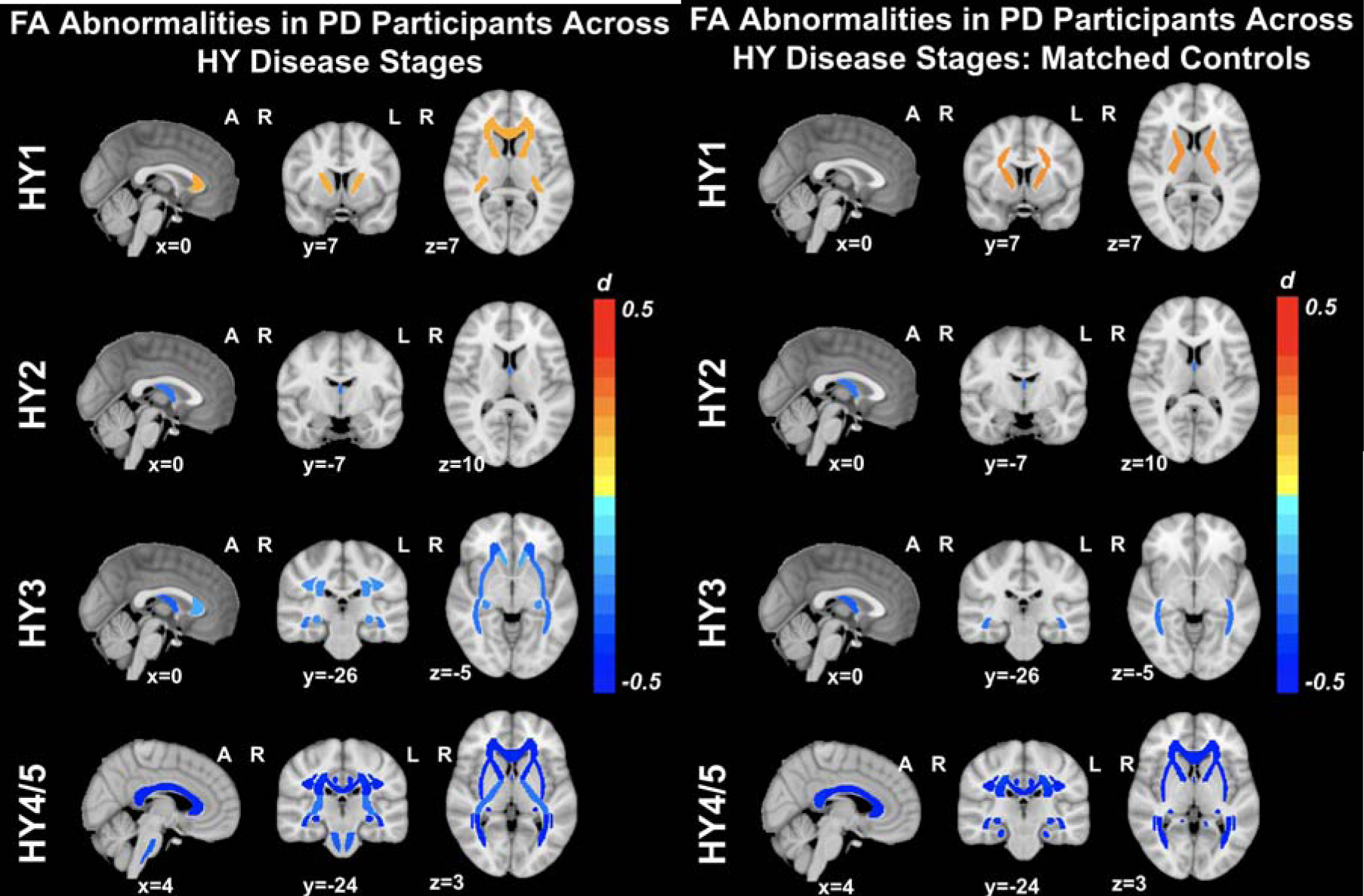
Microstructural differences in FA. Between-group analyses comparing FA across PD H subgroups and controls. Results when compared to all controls on left, and when compared to only age- and sex-matched controls on the right. **Abbreviations**: A, anterior; L, left; R, right;

#### FA: Matched Control Samples

HY1 PD participants displayed *higher* FA across the entire WM skeleton (*d*=0.27) as well as 3 out of 21 ROIs, relative to matched controls (*n*=275), with effect sizes *d*=0.23-0.24. Implicated regions included the anterior *corona radiata* and the anterior and posterior limb of the internal capsule. HY2 PD participants demonstrated significantly *lower* FA in the fornix (*d*=-0.27), relative to matched controls (*n*=742). HY3 PD participants displayed *lower* FA in the fornix *(d*=-0.31) and the sagittal *stratum* (*d*=-0.29), relative to matched controls (*n*=220). HY4/5 PD participants displayed widespread significant differences in FA across the entire WM skeleton (*d*=-0.56) as well as 18 out of 21 ROIs, relative to matched controls (*n*=75), with effect sizes ranging from *d*=-0.38 to -1.09. ROIs not implicated include the retrolenticular and posterior limb of the internal capsule and the corticospinal tract (**Figure 2; Supplementary Results 2.1.2**).

#### MD: Stratification by HY Stage

HY1 PD participants (*n*=275) displayed *lower* MD across the entire WM skeleton (*d*=-0.19) as well as 5 out of 21 ROIs, relative to controls (*n*=885), with effect sizes ranging from *d*=-0.18 to *d*=-0.27. Implicated regions include the anterior and retrolenticular limbs of the internal capsule, the fornix/*stria terminalis*, cingulum (hippocampal portion) and the sagittal *stratum*. HY2 PD participants displayed *lower* MD at the fornix/*stria terminalis* (*d*=-0.22), as well as 2 out of 21 ROIs relative to controls, with effect sizes ranging from *d*=-0.15 to -0.19. Implicated regions include the retrolenticular limb of the internal capsule and the cingulum (hippocampal portion). HY2 participants also displayed *greater* MD at the fornix (*d*=0.15). There were no significant MD differences in HY3 PD participants (*n*=220) relative to controls. HY4/5 PD participants (*n*=75) displayed significantly *higher* MD in the fornix (*d*=0.69) as well as 6 out of 21 ROIs, relative to controls, with effect sizes ranging from *d*=0.33 to 0.46. Implicated regions include the anterior and superior *corona radiata*, external capsule, genu and body of the corpus callosum and the superior fronto-occipital fasciculus. HY4/5 PD participants also displayed *lower* MD in the cingulum (hippocampal portion) (*d*=-0.32) relative to controls (**Figure 3; Supplementary Results 2.1.3**).

**Figure 3:**
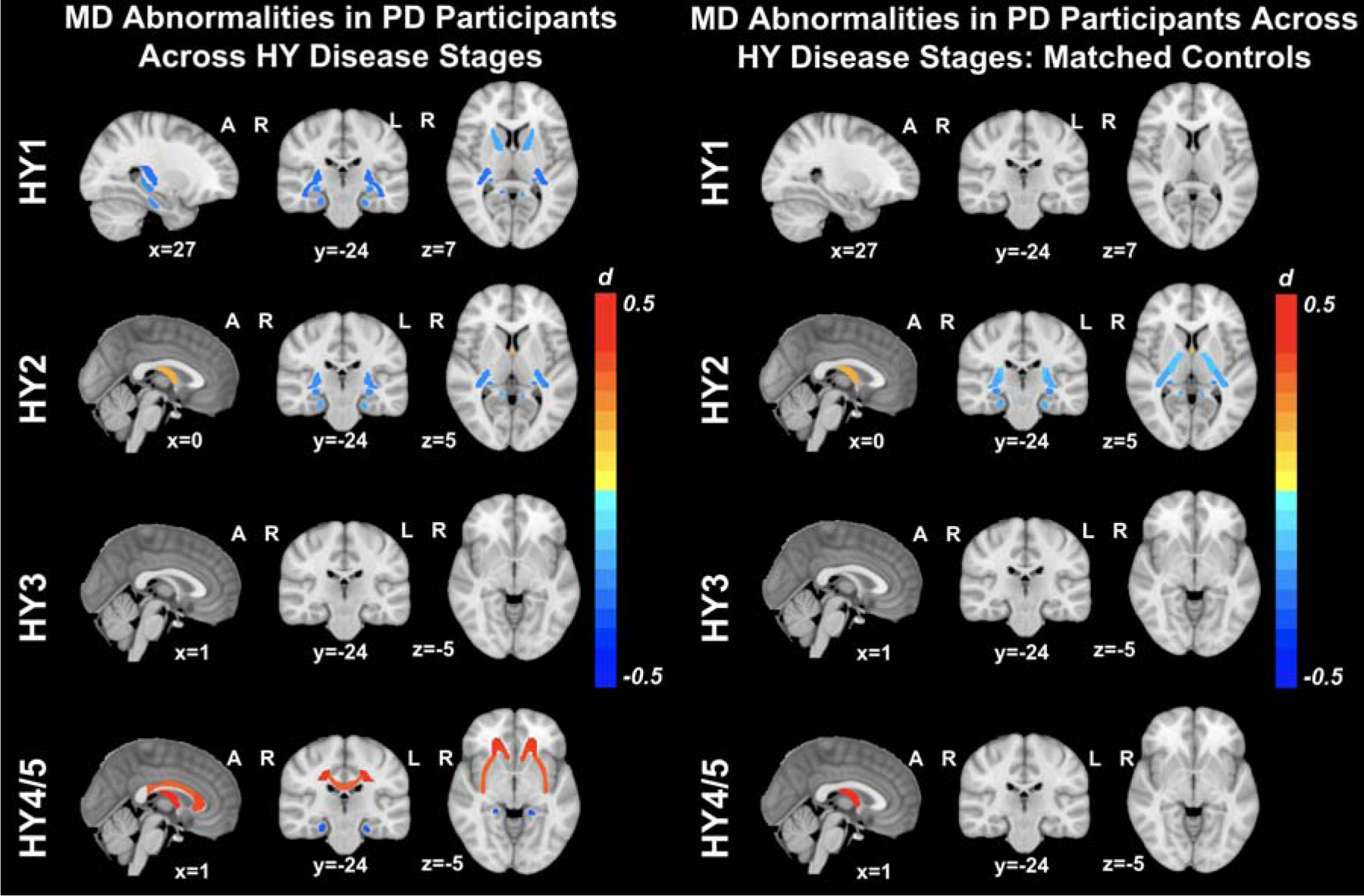
Microstructural differences in MD. Between-group analyses comparing MD across PD H subgroups and controls. Results when compared to all controls on left, and when compared to only age- an sex-matched controls on the right. **Abbreviations**: A, anterior; L, left; R, right;

#### MD: Matched Control Samples

There were no significant differences in MD in HY1 PD participants relative to matched controls (*n*=275). HY2 PD participants demonstrated significantly *lower* MD at the fornix/*stria terminalis* (*d*=-0.22), as well as 3 out of 21 ROIs, relative to matched controls (*n*=742), with effect sizes ranging from *d*=-0.14 to -0.19. Implicated regions include the posterior limb and the retrolenticular parts of the internal capsule as well as the cingulum (hippocampal portion). These participants also showed *higher* MD at the fornix (*d*=0.15). There were no significant MD differences in HY3 PD participants relative to matched controls (*n*=275), while HY4/5 PD participants displayed significantly *higher* MD at the fornix (*d*=0.72) relative to matched controls (*n*=75) (**Figure 3; Supplementary Results 2.1.4**).

**Figure 3:**
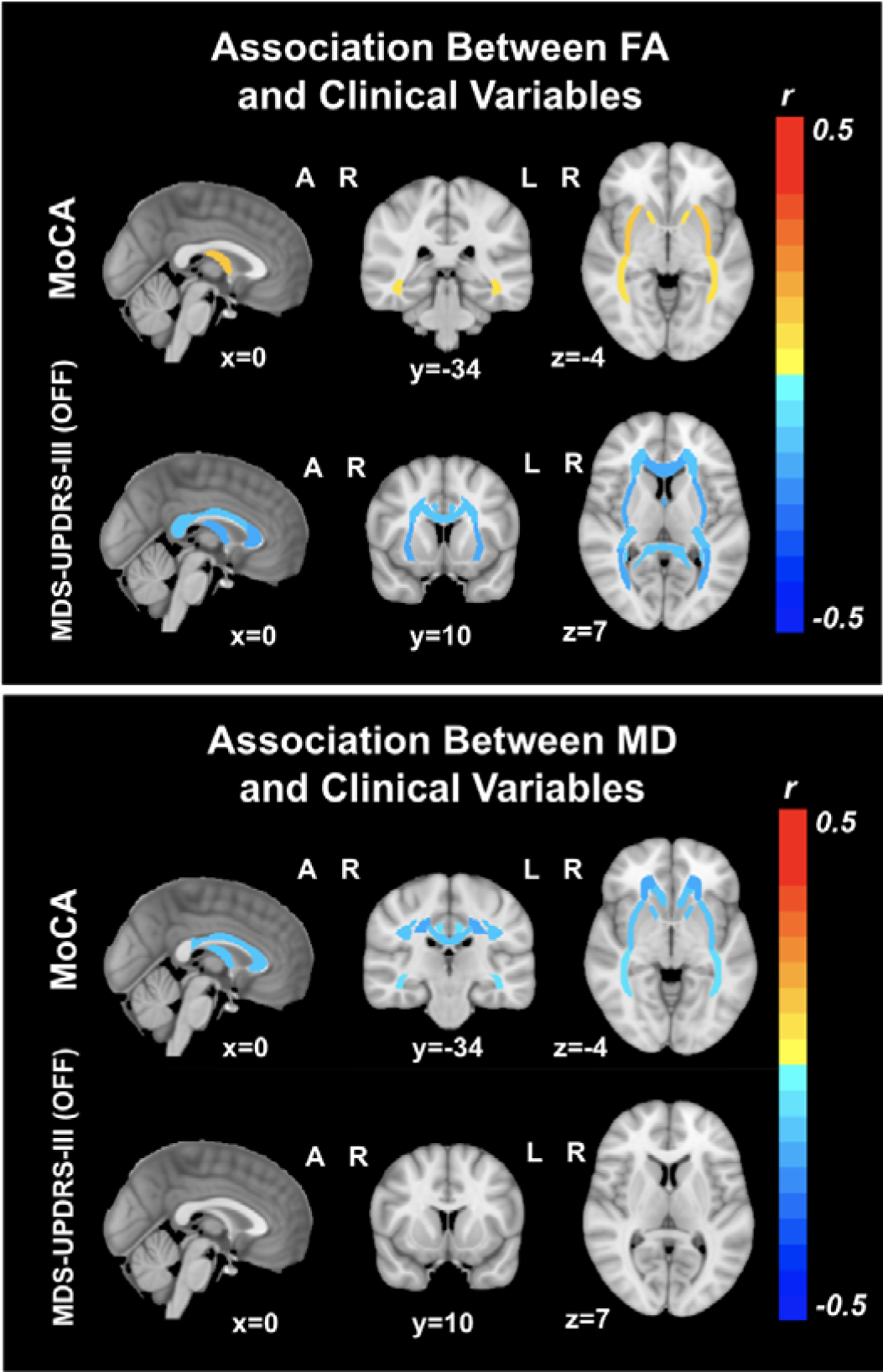
Associations between DTI measures and MoCA and MDS-UPDRS-III (OFF) Score Abbreviations: A, anterior; L, left; R, right; MoCA, Montreal Cognitive Assessment; MDS-UPDRS-I Movement Disorders Sponsored Revision of the Unified Parkinson’s Disease Rating Scale part-III

#### Overall Aggregation of All PD Participants and Controls: FA and MD

If participants from all HY stages were combined, the overall PD group (*n*=1,654) showed significantly *lower* FA (*d*=-0.24) and *higher* MD (*d*=0.17) at the fornix, relative to controls (*n*=885). The overall PD group also showed lower MD at the cingulum (hippocampal portion) (*d*=-0.16) relative to controls (**Supplementary Results 2.2**).

#### Random-effects Meta-Analysis: PD and Controls

Random-effects meta-analysis revealed no significant differences in FA or MD between entire PD cohort and controls across the individual sites (**Supplementary Results 2.3**).

#### Interaction Effects between Sex and Diagnosis

We found no significant interactions between sex and diagnosis when comparing FA and MD across each PD HY stage and matched controls.

#### Associations Between Effect Sizes

A number of results found when performing HY Stage analyses did not remain significant when performing the same analyses with age- and sex-matched controls. To investigate this incongruence, we examined the correlation between the effect sizes generated at the two levels of analysis. Effect sizes generated for each respective ROI across analyses were strongly positively correlated, with coefficients greater than 0.8 found for each ROI (**Supplementary Results 2.4**).

### Associations between Diffusion Measures and Clinical Variables

#### FA/MD and Montreal Cognitive Assessment Scores

In PD participants with MoCA data (*n*=907), we found significant positive associations between these scores and FA across the entire WM skeleton (*r*=0.11), as well as 5 out of 21 ROIs, with effect sizes ranging from *r*=0.09 to 0.13. Implicated regions include the anterior limb of the internal capsule, external capsule, fornix, posterior thalamic radiation and the sagittal stratum. Significant *negative* associations with MD were found across the entire WM skeleton (*r*=-0.13), as well as 12 out of 21 ROIs: effect sizes ranged from *r*=0.07 to 0.17. implicated regions include the anterior and superior *corona radiata*, anterior limb of the internal capsule, external capsule, fornix, genu and body of the corpus callosum, cingulate gyrus, superior frontoLoccipital and longitudinal fasciculi, sagittal stratum and the uncinate fasciculus (**Supplementary Results 2.5.1**)

#### FA/MD and MDS-UPDRS Part-III (OFF) Scores

In PD participants with MDS-UPDRS-III (OFF) scores (*n*=540), we found significant *negative* associations between these scores and FA across the entire WM skeleton as well as 14 out of 21 ROIs, with effect sizes ranging from *r*=0.10 to 0.15. Implicated regions include the anterior, superior and posterior *corona radiata*, retrolenticular part of the internal capsule, external capsule, fornix, posterior thalamic radiation, genu, body and splenium of the corpus callosum, cingulate gyrus, sagittal stratum, tapetum and the uncinate fasciculus. No significant associations were detected between MD and MDS-UPDRS-III (OFF) scores (**Supplementary Results 2.5.2**).

## Discussion

### Summary of Findings

In this worldwide study of white matter microstructure in PD, the largest of its kind, we compared brain diffusion MRI data from 1,654 PD participants to 885 control participants from 17 cohorts across Africa, Asia, Europe, North and South America, and Oceania. We found widespread WM microstructural differences in people with PD that appear as higher FA and lower MD at early disease stages, with a reversal of this pattern at advanced disease stages. We also found significant relationships between FA and MD and cognitive and motor function, with poorer clinical performance associated with lower FA and higher MD.

### Case-Control Analyses

After stratifying participants by HY stage, a pattern of DTI differences emerged that was linked to disease progression. The direction of effects earlier in the disease appeared opposite to those later in the disease. These stage-dependent effects were generally obscured when all PD participants were aggregated.

We found significantly *higher* FA in HY1 participants across the entire WM skeleton, *lower* FA of the fornix in HY2 participants, and *lower* FA across the entire WM skeleton in participants at HY stages 3 and 4/5. In the HY1 PD participants, the largest effect sizes were found for the FA increases in the internal capsule and anterior corona radiata. These structures are key components of the cerebello-thalamo-cortical and basal ganglia-cortical loops and likely play an important role in the pathophysiology of tremor in PD. It has been suggested that tremor signals originating in the basal ganglia ascend via the ventrolateral nucleus of the thalamus through the internal capsule to the corona radiata where they radiate out to the cortex (Abusrair et al., 2022). One study found that increased FA is associated with tremor-dominant, relative to postural instability-gait disturbance PD participants, suggesting an important relationship between this motor phenotype and DTI abnormalities in PD (Wen et al., 2018). The fornix also shows among the largest effect sizes in PD; FA was significantly reduced in HY2, HY3 and HY4/5 participants. The fornix is a major hippocampal output structure that traverses longitudinally from the mesial temporal lobes to the diencephalon and basal forebrain, and it plays a key role in cognition and episodic memory recall (Douet & Chang, 2015). Here we show that this structure is greatly impacted in PD, with a significant correlation between FA and cognitive performance.

We identified significantly *lower* MD across the entire WM skeleton at the HY1 stage, while in HY2, this result was confined to the internal capsule, fornix/*stria terminalis* and the cingulum (hippocampal portion). In HY3, there were no significant differences in MD, while HY4/5 participants showed significantly *higher* MD at widespread regions of interest. Some of the significant MD findings were not observed when comparing PD cohorts to age- and sex-matched controls, with only the most robust effect sizes remaining significant. In our correlational analyses, the effect sizes generated at the matched and unmatched levels of analysis had the same direction and were highly correlated, suggesting that the lack of significant results when comparing MD in HY3 and HY4/5 groups with age- and sex-matched controls are due to differences in sample size and statistical power, as well as removing confounding effects. Similar to our results for the FA analyses, in HY1 and HY2 participants large effect sizes were found for ROIs in the internal capsule, further highlighting this region as an important neuroanatomical structure in PD. In HY4/5 participants, we also found significantly increased MD in the fornix, and MD of the fornix was inversely correlated with performance on the MoCA cognitive assessment.

When all PD participants were combined irrespective of HY disease stage, the full PD cohort showed lower FA and higher MD at the fornix, and lower MD at the cingulum (hippocampal portion) compared to controls, but no other significant effects.

The contrast in findings between 1) comparing all cases to controls and 2) performing stage-stratified analyses may explain why a number of prior TBSS studies in PD have found few, if any, significant differences in FA or MD (Acosta-Cabronero et al., 2017; Inguanzo et al., 2021; L. Ji et al., 2015; Kamagata et al., 2013; Quattrone et al., 2019; Worker et al., 2014), while other studies that have found lower FA or higher MD in PD (N.-K. Chen et al., 2018; Cousineau et al., 2017; Georgiopoulos et al., 2017; Mole et al., 2016; Skidmore et al., 2015; Vercruysse et al., 2015). Stage-dependent effects may account for these opposing findings in the literature, which were resolved by the current design using the large ENIGMA-PD sample.

Diffusion anisotropy in the WM, including the DTI-based FA metric, is influenced by differences in fiber density, degree of myelination, diameter of fibers, presence of ‘crossing’ or ‘kissing’ fibers, and the density of neuroglial cells (Pierpaoli et al., 1996). Several PD-related processes have been linked to changes to anisotropy and diffusivity (Lenfeldt et al., 2015). Higher FA may indicate a compensatory response to altered activity arising from dopaminergic depletion of the *substantia nigra* and connected neuronal pathways (Brotchie & Fitzer-Attas, 2009; Mole et al., 2016). Such a process could promote activity-driven myelination whereby neuronal reserve is recruited and strengthened in response to the increased demand (Hanganu et al., 2018). At the onset of PD symptoms, over two thirds of dopaminergic neurons in the lateral ventral *substantia nigra* may already be lost (Fearnley & Lees, 1991). Still, many participants in the early stages of PD can maintain normal levels of functional performance in a range of domains, potentially through the recruitment of compensatory mechanisms (Cabeza et al., 2018; Hanganu et al., 2018; Palop et al., 2006). This theory is supported by recent work demonstrating that lower clinical severity in PD is associated with stronger upregulation of activity in parietal and premotor cortex during movements made under mild cognitive load (Johansson et al., 2023). Further supporting this *compensatory* hypothesis, experimental animal models of PD have found increased anisotropy and decreased diffusivity in the degenerating nigrostriatal pathways after injection of the 6-OHDA neurotoxin (Van Camp et al., 2009). Other compensatory mechanisms that may contribute to higher anisotropy or lower diffusivity could include increases in axonal density, potentially due to axonal sprouting (Arkadir et al., 2014; Kamagata et al., 2013). Supporting this, a PD study using advanced diffusion MRI data found greater apparent fiber and WM tract-density metrics early in PD compared to controls (Cousineau et al., 2017). Increased anisotropy may reflect neuroinflammation, whereby the infiltration of gliotic cells to certain brain areas may contribute to a more anisotropic diffusion of water molecules (Van Camp et al., 2009). This may partly explain our findings as neuroinflammation plays a crucial role in the etiology of PD (McGeer & McGeer, 2008). Another possibility might be that genetic risk factors for PD may contribute to these DTI alterations. Members of our team recently reported that polygenic risk for PD is associated with greater cortical surface area, potentially relating to increases in neural progenitor cells (Abbasi et al., 2022). Subcortical white matter contains populations of glial progenitors (Nunes et al., 2003), and genetic polymorphisms associated with PD may impact the development of these cells in a way that increases the number or density of white matter cells in PD.

In contrast, several studies have also reported lower FA (B. Chen et al., 2017; Diez-Cirarda et al., 2015; Guan et al., 2019; Guimarães et al., 2018; Inguanzo et al., 2021; G.-J. Ji et al., 2019; L. Ji et al., 2015; Li et al., 2018), and higher MD in PD compared to controls (Guan et al., 2019; Kim et al., 2013; Melzer et al., 2013; Minett et al., 2018), as we found at later disease stages. These findings are more typically associated with neurodegeneration, where brain tissue is constraining water diffusion to a lesser degree, due to neurite loss or diminished membrane integrity causing an increase in the extracellular space (Alexander et al., 2011; Sykova, 2004). Considering the neuropathology of PD in particular, the accumulation of Lewy neurites in neuronal axons impacts the functional integrity and survival of neurons by inhibiting axonal transport (Morfini et al., 2009; Perlson et al., 2010). Further, alpha-synuclein, a key component of Lewy neurites, inhibits neurite outgrowth and branching (Koch et al., 2015). Comorbid small-vessel ischemia and WM hyperintensities may also contribute to lower anisotropy and higher diffusivity in WM tracts in PD (Bohnen & Albin, 2011). Our group has shown that neurodegeneration, in the form of deficits in subcortical and cortical grey matter tissue, occurs at greater levels at advanced disease stages (Laansma et al., 2021), and it does so in a way that parallels the topological spread of Lewy pathology across the brain which becomes more severe as the disease advances (Braak et al., 2003).

### Interaction Effects between Sex and Diagnosis

Females have a lower incidence of PD, generally slower disease progression, and a more mild motor phenotype, indicating likely hormonal, environmental, genetic load and gene-environment interactions (Cerri et al., 2019; Haaxma et al., 2007; Marras & Saunders-Pullman, 2014). Given these factors, we expected to find a significant interaction effect between sex and PD diagnosis in this work; however, we found no evidence to support this hypothesis.

### Associations Between Diffusion measures with Clinical Variables

We found widespread *positive* associations between MoCA scores and FA and *negative* associations between MoCA scores and MD across the brain. The relationship between lower MoCA scores (more impairment) and higher MD, is consistent with prior reports in PD linking DTI alterations to level of cognitive function (Gallagher et al., 2013; Hattori et al., 2012; Koshimori et al., 2015; Melzer et al., 2013; Zheng et al., 2014). While DTI-derived diffusivity alterations have been associated with cognitive impairment in PD (Zhang & Burock, 2020), here we also show anisotropy effects related to cognitive impairment. The large sample size of our study provided the power to uncover these small but significant effects in FA, with effect sizes ranging from *r*=0.09 to 0.11, that would not necessarily be detectable in studies with small sample sizes. We found *negative* associations between MDS-UPDRS-III (OFF) scores and FA across widespread regions of interest across the brain, with no significant associations found with MD at any regions of interest. Our results contrast with prior reports in smaller samples that were unable to detect associations between FA and measures of motor impairment (L. Ji et al., 2015; Melzer et al., 2013; Minett et al., 2018). Our results provide evidence that microstructural differences in WM may underlie the cognitive impairment and motor disturbances in PD, suggesting that cumulative effects of the PD may result in greater microstructural damage to WM at later disease stages, characteristic of the changes associated with neurodegeneration (Alexander et al., 2011; Sykova, 2004). Higher scores on the MDS-UPDRS-III (OFF) are indicative of greater motor disability in people with PD, and our results suggest that greater motor disability is mainly associated with lower FA, more so than differences in diffusivity.

### Limitations and Future Directions

A limitation of the current work is that we can only speculate about the potential compensatory mechanisms underlying early DTI alterations in PD, and given the cross sectional nature of the study, we could not to map disease progression on an individual level or in the same subjects. Longitudinal studies that include PD participants from across the disease spectrum (including prodromal PD participants) are needed to better understand how DTI metrics change with the progression of the disorder. A second limitation of the current work is that datasets from each site had already been collected, using different clinical protocols, limiting our ability to investigate how DTI differences relate to specific symptoms of the disorder. While a limitation, this is also a strength of our work as it means our findings have been derived from diverse populations from around the world. A final limitation of this work is that we assess large white matter tracts as a whole and do not evaluate the effects of cerebrovascular factors that may impact local white matter regions. Tractography-based analyses incorporating white matter hyperintensities will be performed in the future, as well as mapping how these factors contribute to cognitive impairment.

## Conclusion

Our research demonstrates that people with Parkinson’s disease present with a pattern of DTI alterations involving higher anisotropy and lower diffusivity at early disease stages. By contrast, later-stage participants exhibit differences more typically associated with neurodegeneration, including lower anisotropy and higher diffusivity.

## Data Availability

Open source datasets that support the findings of this study include PPMI and UK Biobank. Data for remaining cohorts are not openly available, but researchers are invited to register interest with the ENIGMA-PD Working Group in order to formally request site data through secondary proposals.

## References

Abbasi, N., Tremblay, C., Rajimehr, R., Yu, E., Markello, R. D., Shafiei, G., Khatibi, N., study, E.-P., Jahanshad, N., & Thompson, P. M. (2022). Neuroanatomical correlates of polygenic risk for Parkinson’s Disease. medRxiv, 2022.01. 17.22269262.

Abusrair, A. H., Elsekaily, W., & Bohlega, S. (2022). Tremor in Parkinson’s Disease: From Pathophysiology to Advanced Therapies. Tremor and Other Hyperkinetic Movements, 12.

Acosta-Cabronero, J., Cardenas-Blanco, A., Betts, M. J., Butryn, M., Valdes-Herrera, J. P., Galazky, I., & Nestor, P. J. (2017). The whole-brain pattern of magnetic susceptibility perturbations in Parkinson’s disease. Brain, 140(1), 118–131.

Alexander, A. L., Hurley, S. A., Samsonov, A. A., Adluru, N., Hosseinbor, A. P., Mossahebi, P., Tromp, D. P., Zakszewski, E., & Field, A. S. (2011). Characterization of cerebral white matter properties using quantitative magnetic resonance imaging stains. Brain Connectivity, 1(6), 423–446.

Alexander, A. L., Lee, J. E., Lazar, M., & Field, A. S. (2007). Diffusion tensor imaging of the brain. Neurotherapeutics, 4(3), 316–329.

Arkadir, D., Bergman, H., & Fahn, S. (2014). Redundant dopaminergic activity may enable compensatory axonal sprouting in Parkinson disease. Neurology, 82(12), 1093–1098.

Atkinson-Clement, C, Pinto, S, Eusebio, A, & Coulon O. (2017). Diffusion tensor imaging in Parkinson’s disease: Review and meta-analysis. 2213–1582.

Benjamini, Y., & Hochberg, Y. (1995). Controlling the false discovery rate: A practical and powerful approach to multiple testing. Journal of the Royal Statistical Society: Series B (Methodological) 57(1), 289–300.

Bohnen, N. I., & Albin, R. L. (2011). White matter lesions in Parkinson disease. Nature Reviews Neurology, 7(4), 229–236.

Braak, H., Del Tredici, K., Rüb, U., De Vos, R. A., Steur, E. N. J., & Braak, E. (2003). Staging of brain pathology related to sporadic Parkinson’s disease. Neurobiology of Aging, 24(2), 197–211.

Brotchie, J., & Fitzer-Attas, C. (2009). Mechanisms compensating for dopamine loss in early Parkinson disease. Neurology, 72(7 Supplement 2), S32–S38.

Cabeza, R., Albert, M., Belleville, S., Craik, F. I., Duarte, A., Grady, C. L., Lindenberger, U., Nyberg, L., Park, D. C., & Reuter-Lorenz, P. A. (2018). Maintenance, reserve and compensation: The cognitive neuroscience of healthy ageing. Nature Reviews Neuroscience, 19(11), 701–710.

Cerri, S., Mus, L., & Blandini, F. (2019). Parkinson’s disease in women and men: What’s the difference? Journal of Parkinson’s Disease, 9(3), 501–515.

Chen, B., Fan, G., Sun, W., Shang, X., Shi, S., Wang, S., Lv, G., & Wu, C. (2017). Usefulness of diffusion-tensor MRI in the diagnosis of Parkinson variant of multiple system atrophy and Parkinson’s disease: A valuable tool to differentiate between them? Clinical Radiology, 72(7), 610. e9-610. e15.

Chen, N.-K., Chou, Y.-H., Sundman, M., Hickey, P., Kasoff, W. S., Bernstein, A., Trouard, T. P., Lin, T., Rapcsak, S. Z., & Sherman, S. J. (2018). Alteration of diffusion-tensor magnetic resonance imaging measures in brain regions involved in early stages of parkinson’s disease. Brain Connectivity, 8(6), 343–349.

Cousineau, M., Jodoin, P.-M., Garyfallidis, E., Côté, M.-A., Morency, F. C., Rozanski, V., Grand’Maison, M., Bedell, B. J., & Descoteaux, M. (2017). A test-retest study on Parkinson’s PPMI dataset yields statistically significant white matter fascicles. NeuroImage: Clinical, 16, 222–233.

Dennis, E. L., Disner, S. G., Fani, N., Salminen, L. E., Logue, M., Clarke, E. K., Haswell, C. C., Averill, C. L., Baugh, L. A., & Bomyea, J. (2021). Altered white matter microstructural organization in posttraumatic stress disorder across 3047 adults: Results from the PGC-ENIGMA PTSD consortium. Molecular Psychiatry, 26(8), 4315–4330.

Diez-Cirarda, M., Ojeda, N., Pena, J., Cabrera-Zubizarreta, A., Gómez-Beldarrain, M. Á., Gómez-Esteban, J. C., & Ibarretxe-Bilbao, N. (2015). Neuroanatomical correlates of theory of mind deficit in Parkinson’s disease: A multimodal imaging study. PLoS One, 10(11), e0142234.

Dorsey, E. R., Elbaz, A., Nichols, E., Abd-Allah, F., Abdelalim, A., Adsuar, J. C., Ansha, M. G., Brayne, C., Choi, J.-Y. J., Collado-Mateo, D., Dahodwala, N., Do, H. P., Edessa, D., Endres, M., Fereshtehnejad, S.-M., Foreman, K. J., Gankpe, F. G., Gupta, R., Hankey, G. J., … Murray, C. J. L. (2018). Global, regional, and national burden of Parkinson’s disease, 1990–2016: A systematic analysis for the Global Burden of Disease Study 2016. The Lancet Neurology, 17(11), 939–953. 10.1016/S14744422(18)30295-3

Douet, V., & Chang, L. (2015). Fornix as an imaging marker for episodic memory deficits in healthy aging and in various neurological disorders. Frontiers in Aging Neuroscience, 6, 343.

Fahn, S., & Elton, R. L. (1987). UPDRS program members. Unified Parkinsons disease rating scale. Recent Developments in Parkinson’s Disease, 2, 153–163.

Favre, P., Pauling, M., Stout, J., Hozer, F., Sarrazin, S., Abé, C., Alda, M., Alloza, C., Alonso-Lana, S., & Andreassen, O. A. (2019). Widespread white matter microstructural abnormalities in bipolar disorder: Evidence from mega-and meta-analyses across 3033 individuals. Neuropsychopharmacology, 44(13), 2285–2293.

Fearnley, J. M., & Lees, A. J. (1991). Ageing and Parkinson’s disease: Substantia nigra regional selectivity. Brain, 114(5), 2283–2301.

Feigin, V. L., Nichols, E., Alam, T., Bannick, M. S., Beghi, E., Blake, N., Culpepper, W. J., Dorsey, E. R., Elbaz, A., & Ellenbogen, R. G. (2019). Global, regional, and national burden of neurological disorders, 1990–2016: A systematic analysis for the Global Burden of Disease Study 2016. The Lancet Neurology, 18(5), 459–480.

Folstein, M. F., Folstein, S. E., & McHugh, P. R. (1975). “Mini-mental state”: A practical method for grading the cognitive state of patients for the clinician. Journal of Psychiatric Research, 12(3), 189–198.

Gallagher, C., Bell, B., Bendlin, B., Palotti, M., Okonkwo, O., Sodhi, A., Wong, R., Buyan-Dent, L., Johnson, S., & Wilette, A. (2013). White matter microstructural integrity and executive function in Parkinson’s disease. Journal of the International Neuropsychological Society, 19(3), 349–354.

Georgiopoulos, C., Warntjes, M., Dizdar, N., Zachrisson, H., Engström, M., Haller, S., & Larsson, E.-M. (2017). Olfactory impairment in Parkinson’s disease studied with diffusion tensor and magnetization transfer imaging. Journal of Parkinson’s Disease, 7(2), 301–311.

Guan, X., Huang, P., Zeng, Q., Liu, C., Wei, H., Xuan, M., Gu, Q., Xu, X., Wang, N., & Yu, X. (2019). Quantitative susceptibility mapping as a biomarker for evaluating white matter alterations in Parkinson’s disease. Brain Imaging and Behavior, 13, 220–231.

Guimarães, R. P., Campos, B. M., de Rezende, T. J., Piovesana, L., Azevedo, P. C., Amato-Filho, A. C., Cendes, F., & D’Abreu, A. (2018). Is diffusion tensor imaging a good biomarker for early Parkinson’s disease? Frontiers in Neurology, 9, 626.

Haaxma, C. A., Bloem, B. R., Borm, G. F., Oyen, W. J., Leenders, K. L., Eshuis, S., Booij, J., Dluzen, D. E., & Horstink, M. W. (2007). Gender differences in Parkinson’s disease. Journal of Neurology, Neurosurgery & Psychiatry, 78(8), 819–824.

Hanganu, A., Houde, J.-C., Fonov, V. S., Degroot, C., MejiaLConstain, B., Lafontaine, A.-L., Soland, V., Chouinard, S., Collins, L. D., & Descoteaux, M. (2018). White matter degeneration profile in the cognitive corticoLsubcortical tracts in Parkinson’s disease. Movement Disorders, 33(7), 1139–1150.

Hatton, S. N., Huynh, K. H., Bonilha, L., Abela, E., Alhusaini, S., Altmann, A., Alvim, M. K. M., Balachandra, A. R., Bartolini, E., Bender, B., Bernasconi, N., Bernasconi, A., Bernhardt, B., Bargallo, N., Caldairou, B., Caligiuri, M. E., Carr, S. J. A., Cavalleri, G. L., Cendes, F., … McDonald, C. R. (2020). White matter abnormalities across different epilepsy syndromes in adults: An ENIGMA-Epilepsy study. Brain, 143(8), 2454–2473. 10.1093/brain/awaa200

Hattori, T., Orimo, S., Aoki, S., Ito, K., Abe, O., Amano, A., Sato, R., Sakai, K., & Mizusawa, H. (2012). Cognitive status correlates with white matter alteration in Parkinson’s disease. Human Brain Mapping, 33(3), 727–739.

Hoehn, M. M., & Yahr, M. D. (1967). Parkinsonism: Onset, progression and mortality. Neurology, 17(5), 427–442. 10.1212/wnl.17.5.427

Hua, K., Zhang, J., Wakana, S., Jiang, H., Li, X., Reich, D. S., Calabresi, P. A., Pekar, J. J., van Zijl, P. C., & Mori, S. (2008). Tract probability maps in stereotaxic spaces: Analyses of white matter anatomy and tract-specific quantification. Neuroimage, 39(1), 336–347.

Inguanzo, A., Sala-Llonch, R., Segura, B., Erostarbe, H., Abós, A., Campabadal, A., Uribe, C., Baggio, H. C., Compta, Y., & Marti, M. J. (2021). Hierarchical cluster analysis of multimodal imaging data identifies brain atrophy and cognitive patterns in Parkinson’s disease. Parkinsonism & Related Disorders, 82, 16–23.

Jahanshad, N., Kochunov, P. V., Sprooten, E., Mandl, R. C., Nichols, T. E., Almasy, L., Blangero, J., Brouwer, R. M., Curran, J. E., de Zubicaray, G. I., Duggirala, R., Fox, P. T., Hong, L. E., Landman, B. A., Martin, N. G., McMahon, K. L., Medland, S. E., Mitchell, B. D., Olvera, R. L., … Glahn, D. C. (2013). Multi-site genetic analysis of diffusion images and voxelwise heritability analysis: A pilot project of the ENIGMA–DTI working group. NeuroImage, 81, 455–469. 10.1016/j.neuroimage.2013.04.061

Ji, G.-J., Ren, C., Li, Y., Sun, J., Liu, T., Gao, Y., Xue, D., Shen, L., Cheng, W., & Zhu, C. (2019). Regional and network properties of white matter function in Parkinson’s disease. Human Brain Mapping, 40(4), 1253–1263.

Ji, L., Wang, Y., Zhu, D., Liu, W., & Shi, J. (2015). White matter differences between multiple system atrophy (parkinsonian type) and Parkinson’s disease: A diffusion tensor image study. Neuroscience, 305, 109–116.

Johansson, M. E., Toni, I., Kessels, R. P. C., Bloem, B. R., & Helmich, R. C. (2023). Clinical severity in Parkinson’s disease is determined by decline in cortical compensation. Brain, awad325. 10.1093/brain/awad325

Kamagata, K., Motoi, Y., Tomiyama, H., Abe, O., Ito, K., Shimoji, K., Suzuki, M., Hori, M., Nakanishi, A., & Sano, T. (2013). Relationship between cognitive impairment and white-matter alteration in Parkinson’s disease with dementia: Tract-based spatial statistics and tract-specific analysis. European Radiology, 23, 1946–1955.

Kelly, S., Jahanshad, N., Zalesky, A., Kochunov, P., Agartz, I., Alloza, C., Andreassen, O. A., Arango, C., Banaj, N., & Bouix, S. (2018). Widespread white matter microstructural differences in schizophrenia across 4322 individuals: Results from the ENIGMA Schizophrenia DTI Working Group. Molecular Psychiatry, 23(5), 1261–1269.

Kim, H. J., Kim, S. J., Kim, H. S., Choi, C. G., Kim, N., Han, S., Jang, E. H., Chung, S. J., & Lee, C. S. (2013). Alterations of mean diffusivity in brain white matter and deep gray matter in Parkinson’s disease. Neuroscience Letters, 550, 64–68.

Koch, J. C., Bitow, F., Haack, J., d’Hedouville, Z., Zhang, J. N., Tönges, L., Michel, U., Oliveira, L. M. A., Jovin, T. M., & Liman, J. (2015). Alpha-Synuclein affects neurite morphology, autophagy, vesicle transport and axonal degeneration in CNS neurons. Cell Death & Disease, 6(7), e1811–e1811.

Koshimori, Y., Segura, B., Christopher, L., Lobaugh, N., Duff-Canning, S., Mizrahi, R., Hamani, C., Lang, A. E., Aminian, K., & Houle, S. (2015). Imaging changes associated with cognitive abnormalities in Parkinson’s disease. Brain Structure and Function, 220, 2249–2261.

Laansma, M. A., Bright, J. K., Al-Bachari, S., Anderson, T. J., Ard, T., Assogna, F., Baquero, K. A., Berendse, H. W., Blair, J., Cendes, F., Dalrymple-Alford, J. C., de Bie, R. M. A., Debove, I., Dirkx, M. F., Druzgal, J., Emsley, H. C. A., Garraux, G., Guimarães, R. P., Gutman, B. A., … The ENIGMA-Parkinson’s Study. (2021). International Multicenter Analysis of Brain Structure Across Clinical Stages of Parkinson’s Disease. Movement Disorders, 36(11), 2583–2594. 10.1002/mds.28706

Lenfeldt, N., Larsson, A., Nyberg, L., Birgander, R., & Forsgren, L. (2015). Fractional anisotropy in the substantia nigra in Parkinson’s disease: A complex picture. European Journal of Neurology, 22(10), 1408–1414.

Li, X.-R., Ren, Y.-D., Cao, B., & Huang, X.-L. (2018). Analysis of white matter characteristics with tract-based spatial statistics according to diffusion tensor imaging in early Parkinson’s disease. Neuroscience Letters, 675, 127–132.

Marras, C., & Saunders-Pullman, R. (2014). The complexities of hormonal influences and risk of Parkinson’s disease. Movement Disorders: Official Journal of the Movement Disorder Society, 29(7), 845.

McGeer, P. L., & McGeer, E. G. (2008). Glial reactions in Parkinson’s disease. Movement Disorders: Official Journal of the Movement Disorder Society, 23(4), 474–483.

Melzer, T. R., Watts, R., MacAskill, M. R., Pitcher, T. L., Livingston, L., Keenan, R. J., Dalrymple-Alford, J. C., & Anderson, T. J. (2013). White matter microstructure deteriorates across cognitive stages in Parkinson disease. Neurology, 80(20), 1841– 1849.

Minett, T., Su, L., Mak, E., Williams, G., Firbank, M., Lawson, R. A., Yarnall, A. J., Duncan, G. W., Owen, A. M., & Khoo, T. K. (2018). Longitudinal diffusion tensor imaging changes in early Parkinson’s disease: ICICLE-PD study. Journal of Neurology, 265, 1528–1539.

Mole, J. P., Subramanian, L., Bracht, T., Morris, H., Metzler-Baddeley, C., & Linden, D. E. J. (2016). Increased fractional anisotropy in the motor tracts of Parkinson’s disease suggests compensatory neuroplasticity or selective neurodegeneration. European Radiology, 26(10), 3327–3335. 10.1007/s00330-015-4178-1

Morfini, G. A., Burns, M., Binder, L. I., Kanaan, N. M., LaPointe, N., Bosco, D. A., Brown, R. H., Brown, H., Tiwari, A., & Hayward, L. (2009). Axonal transport defects in neurodegenerative diseases. Journal of Neuroscience, 29(41), 12776–12786.

Nasreddine, Z. S., Phillips, N. A., Bédirian, V., Charbonneau, S., Whitehead, V., Collin, I., Cummings, J. L., & Chertkow, H. (2005). The Montreal Cognitive Assessment, MoCA: a brief screening tool for mild cognitive impairment. Journal of the American Geriatrics Society, 53(4), 695–699.

Nunes, M. C., Roy, N. S., Keyoung, H. M., Goodman, R. R., McKhann, G., Jiang, L., Kang, J., Nedergaard, M., & Goldman, S. A. (2003). Identification and isolation of multipotential neural progenitor cells from the subcortical white matter of the adult human brain. Nature Medicine, 9(4), 439–447.

Palop, J. J., Chin, J., & Mucke, L. (2006). A network dysfunction perspective on neurodegenerative diseases. Nature, 443(7113), 768–773.

Perlson, E., Maday, S., Fu, M., Moughamian, A. J., & Holzbaur, E. L. (2010). Retrograde axonal transport: Pathways to cell death? Trends in Neurosciences, 33(7), 335–344.

Pierpaoli, C., Jezzard, P., Basser, P. J., Barnett, A., & Di Chiro, G. (1996). Diffusion tensor MR imaging of the human brain. Radiology, 201(3), 637–648.

Piras, F., Piras, F., Abe, Y., Agarwal, S. M., Anticevic, A., Ameis, S., Arnold, P., Banaj, N., Bargalló, N., & Batistuzzo, M. C. (2021). White matter microstructure and its relation to clinical features of obsessive–compulsive disorder: Findings from the ENIGMA OCD Working Group. Translational Psychiatry, 11(1), 173.

Postuma, R. B., Berg, D., Stern, M., Poewe, W., Olanow, C. W., Oertel, W., Obeso, J., Marek, K., Litvan, I., Lang, A. E., Halliday, G., Goetz, C. G., Gasser, T., Dubois, B., Chan, P., Bloem, B. R., Adler, C. H., & Deuschl, G. (2015). MDS clinical diagnostic criteria for Parkinson’s disease. Movement Disorders, 30(12), 1591–1601. 10.1002/mds.26424

Quattrone, A., Caligiuri, M. E., Morelli, M., Nigro, S., Vescio, B., Arabia, G., Nicoletti, G., Nisticò, R., Salsone, M., & Novellino, F. (2019). Imaging counterpart of postural instability and vertical ocular dysfunction in patients with PSP: a multimodal MRI study. Parkinsonism & Related Disorders, 63, 124–130.

Radua, J., Vieta, E., Shinohara, R., Kochunov, P., Quidé, Y., Green, M. J., Weickert, C. S., Weickert, T., Bruggemann, J., & Kircher, T. (2020). Increased power by harmonizing structural MRI site differences with the ComBat batch adjustment method in ENIGMA. NeuroImage, 218, 116956.

Schapira, A. H. V., Chaudhuri, K. R., & Jenner, P. (2017). Non-motor features of Parkinson disease. Nature Reviews Neuroscience, 18(7), 435–450. 10.1038/nrn.2017.62

Skidmore, F. M., Spetsieris, P. G., Anthony, T., Cutter, G. R., von Deneen, K. M., Liu, Y., White, K. D., Heilman, K. M., Myers, J., & Standaert, D. G. (2015). A full-brain, bootstrapped analysis of diffusion tensor imaging robustly differentiates Parkinson disease from healthy controls. Neuroinformatics, 13, 7–18.

Smith, S. M., Jenkinson, M., Johansen-Berg, H., Rueckert, D., Nichols, T. E., Mackay, C. E., Watkins, K. E., Ciccarelli, O., Cader, M. Z., & Matthews, P. M. (2006). Tract-based spatial statistics: Voxelwise analysis of multi-subject diffusion data. Neuroimage, 31(4), 1487–1505.

Sykova, E. (2004). Extrasynaptic volume transmission and diffusion parameters of the extracellular space. Neuroscience, 129(4), 861–876.

Tremblay, C., Abbasi, N., Zeighami, Y., Yau, Y., Dadar, M., Rahayel, S., & Dagher, A. (2020). Sex effects on brain structure in de novo Parkinson’s disease: A multimodal neuroimaging study. Brain, 143(10), 3052–3066.

Van Camp, N., Blockx, I., Verhoye, M., Casteels, C., Coun, F., Leemans, A., Sijbers, J., Baekelandt, V., Van Laere, K., & Van der Linden, A. (2009). Diffusion tensor imaging in a rat model of Parkinson’s disease after lesioning of the nigrostriatal tract. NMR in Biomedicine: An International Journal Devoted to the Development and Application of Magnetic Resonance In Vivo, 22(7), 697–706.

Van Velzen, L. S., Kelly, S., Isaev, D., Aleman, A., Aftanas, L. I., Bauer, J., Baune, B. T., Brak, I. V., Carballedo, A., & Connolly, C. G. (2020). White matter disturbances in major depressive disorder: A coordinated analysis across 20 international cohorts in the ENIGMA MDD working group. Molecular Psychiatry, 25(7), 1511–1525.

Vercruysse, S., Leunissen, I., Vervoort, G., Vandenberghe, W., Swinnen, S., & Nieuwboer, A. (2015). Microstructural changes in white matter associated with freezing of gait in Parkinson’s disease. Movement Disorders, 30(4), 567–576.

Wei, X., Luo, C., Li, Q., Hu, N., Xiao, Y., Liu, N., Lui, S., & Gong, Q. (2021). White matter abnormalities in patients with Parkinson’s disease: A meta-analysis of diffusion tensor imaging using tract-based spatial statistics. Frontiers in Aging Neuroscience, 12, 610962.

Wen, M.-C., Heng, H. S., Lu, Z., Xu, Z., Chan, L. L., Tan, E. K., & Tan, L. C. (2018). Differential white matter regional alterations in motor subtypes of early drug-naive Parkinson’s disease patients. Neurorehabilitation and Neural Repair, 32(2), 129–141.

Worker, A., Blain, C., Jarosz, J., Chaudhuri, K. R., Barker, G. J., Williams, S. C., Brown, R. G., Leigh, P. N., Dell’Acqua, F., & Simmons, A. (2014). Diffusion tensor imaging of Parkinson’s disease, multiple system atrophy and progressive supranuclear palsy: A tract-based spatial statistics study. PloS One, 9(11), e112638.

Zhang, Y., & Burock, M. A. (2020). Diffusion Tensor Imaging in Parkinson’s Disease and Parkinsonian Syndrome: A Systematic Review. Frontiers in Neurology, 11. 10.3389/fneur.2020.531993

Zheng, Z., Shemmassian, S., Wijekoon, C., Kim, W., Bookheimer, S. Y., & Pouratian, N. (2014). DTI correlates of distinct cognitive impairments in Parkinson’s disease. Human Brain Mapping, 35(4), 1325–1333.

