## Supplementary Material for "A Worldwide Study of White Matter Microstructural Alterations in People Living with Parkinson’s Disease"

### **Supplementary Material - White matter abnormalities in Parkinson’s disease: An ENIGMA-Parkinson’s Disease TBSS Study**

### **1. Supplementary Methods**

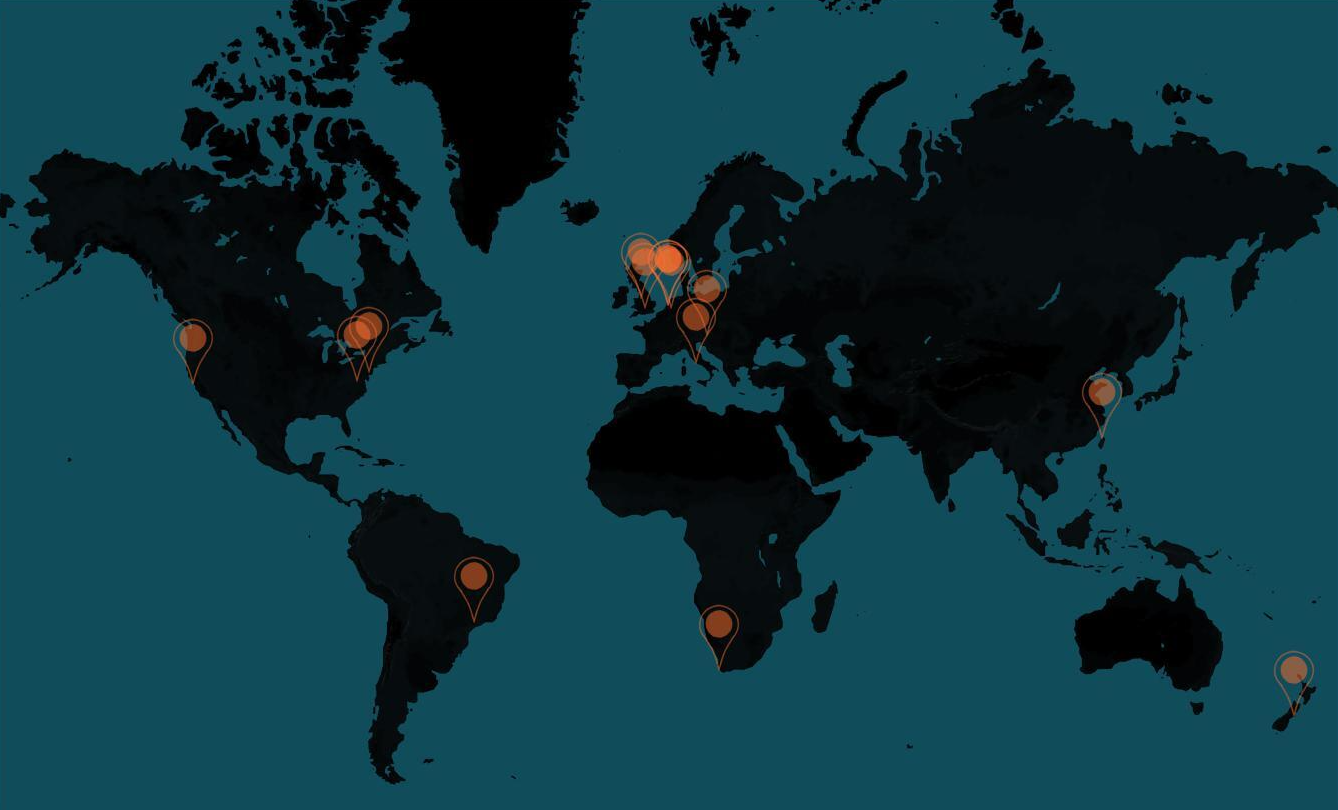

**Supplementary Figure 1: World Map showing location of sites included in this study.** Data were also included that came from the PPMI (https://www.ppmi-info.org/) and UK Biobank (https://www.ukbiobank.ac.uk/) which are large multi-site studies.

#### *1.1 Inclusion and Exclusion Criteria*

| **Site** | **Diagnostic Criteria** | **Inclusion Criteria** | | **Exclusion Criteria** | |
| --- | --- | --- | --- | --- | --- |
|  |  | **Controls** | **PD** | **Controls** | **PD** |
| Amsterdam | UKBB | Sex and age-matched control | Patients seen at the movement disorders outpatient clinic. | - | - |
| Campinas | UKBB | - | Idiopathic PD, taking antiparkinsonian medications, age > 30 years. | - | Clinically significant musculoskeletal, cardiovascular, respiratory or other neurological disease. |
| Chang Gung | NINDS | Aged between 50- 90. | Diagnosis of probable PD, ability to tolerate treatment discontinuation for 12 hours. | Major physical illnesses, psychiatric disorders, known brain abnormalities, history of intracranial surgery, pharmacotherapy for more than ten years or treatment with drugs able to cross the blood- brain-barrier (other than those used to treat PD). | Major physical illnesses, psychiatric disorders, known brain abnormalities, history of intracranial surgery, pharmacotherapy for more than ten years or treatment with drugs able to cross the blood- brain-barrier (other than those used to treat PD). |
| Christchurch | UKBB | - | Met the UK Parkinson's Society criteria for PD, motor symptoms present for at least 1 year at study entry. |  | Atypical parkinsonian disorder, history of moderate/severe head injury, stroke, early-life learning disability, major psychiatric or medical illness in the previous 6 months, poor English (precluding testing). |
| Cogtips | UKBB | Sex, age, and education-matched. | Subjective cognitive complaints (PD-CFRS > 3), HY stage < 4. | Neurological disease, indication of dementia (MoCA < 22), indication of psychotic (SAPS-PD) or depressive disorder (BDI > 18), drugs and/or alcohol abuse, inability to undergo neuropsychological assessment, traumatic brain injury, tumor or vascular abnormalities. | Dementia (SAGE <14 or MoCA < 22), drugs or alcohol abuse (CAGE AID > 1), depressive symptoms (BDI > 18), impulse control disorder (ICD criteria interview), psychotic symptoms (SAPS-PD criteria), tumors and significant vascular abnormalities. |
| Graz | QSBB | No history of previous stroke or dementia and a normal neurologic examination. | Clinical diagnosis of PD. | - | MMSE <24, secondary parkinsonism, atypical parkinsonian diseases, a history of neuroleptic drugs, structural abnormalities on routine MRI scans or a history of previous stroke. |
| NWE | UKBB | Age-matched to PD group and without a history of idiopathic PD or clinical CVD, or any other significant neurological condition. | PD diagnosis without known clinical cardiovascular disease or dementia. No other significant neurological conditions. | - | - |
| Oxford | UKBB | - | PD diagnosis within the past 3.5 years. Full details of criteria are available at: Szewczyk-Krolikowski K et. al. (2013). | Controls without blood relatives with PD. | No atypical features to suggest an alternative diagnosis. No secondary parkinsonism due to head trauma or medication use, atypical parkinsonism syndromes (multiple system atrophy, progressive supra nuclear palsy, corticobasal degeneration, dementia with Lewy bodies), documented postural BP drop on standardized measurement or significant urinary symptoms. |
| PPMI | MDS | https://www.ppmi-info.org/sites/default/files/docs/PA2 PPMI Clinical%20Protocol Final 01Feb2021.pdf | https://www.ppmi-info.org/sites/default/files/docs/PA2 PPMI Clinical%20Protocol Final 01Feb2021.pdf | https://www.ppmi-info.org/sites/default/files/docs/PA2 PPMI Clinical%20Protocol Final 01Feb2021.pdf | https://www.ppmi-info.org/sites/default/files/docs/PA2 PPMI Clinical%20Protocol Final 01Feb2021.pdf |
| Radboud | UKBB | Same age/gender balance as PD patients | Idiopathic PD, UPDRS tremor-score > 2, dopaminergic therapy with a clear clinical response of non-tremor symptoms (bradykinesia, rigidity), HY stage 1-3. | Neurological or psychiatric disease, cognitive impairment (MMSE < 26), medication associated with elongated QT-time, pregnancy, age < 25 years. | Neurological or psychiatric comorbidity, severe head tremor or dyskinesias, cognitive impairment (MMSE < 26), co-medication associated with elongated QT-time, pregnancy, age < 25 years. |
| Rome SLF | MDS | Vision and hearing sufficient for compliance with testing procedures, laboratory values within normal reference intervals, neuropsychological domain scores above normal cognitive level cutoff scores, corrected for age and educational level. | Diagnosis of idiopathic, MMSE score > 26, no dementia. | Dementia or MCI diagnosis, confirmed by a comprehensive neuropsychological battery,  MMSE score<26, presence of major non-stabilized medical illnesses, known or suspected history of alcoholism, drug dependence and abuse, head trauma, and mental disorders (apart from mood or anxiety disorders). | Presence of major non-stabilized medical illnesses, known or suspected history of alcoholism, drug dependence and abuse, head trauma, and mental disorders (apart from mood or anxiety disorders), history of neurological diseases other than idiopathic PD, unclear history of chronic dopaminergic treatment responsiveness. |
| Stanford | UKB | Normal neurological exam and normal neuropsychiatric battery (within 1.5 SD of age- and education- adjusted norms). | > 20% improvement on MDS-UPDRS part III ON medication compared to OFF medication. | - | - |
| Stellenbosch | MDS | No current or lifetime history of any DSM-5 psychiatric disorder | Diagnosis of PD by neurologist , age > 40 years and <= than 75 years.  Exclusion: Any significant medical/physical illness (other than PD), participants with metal prostheses, cardiac pacemakers or metal clips likely to interfere with ability to acquire MR image | Neurological conditions that would preclude completion of neurocognitive tasks, current or lifetime daily psychotropic medication use. | - |
| UCSF | MDS | - | Ages 40 to 85; Diagnosis of Parkinson’s disease and mild to moderate symptoms defined by a Hoehn and Yahr score of 1.0 to 3.0; Diagnosis of a movement disorder with Parkinsonian symptoms, including but not limited to multiple system atrophy. | Any contra-indication for undergoing MRI, Abnormal MRI findings. History of encephalitis, multiple sclerosis, other CNS infection, epilepsy, or primary CNS disease besides PD, loss of consciousness for more than 2 minutes; clinical diagnosis of dementia, as indicated by clinical interview. Inability to give informed consent. | Any contra-indication for undergoing MRI, Abnormal MRI findings. History of encephalitis, multiple sclerosis, other CNS infection, epilepsy, or primary CNS disease besides PD, loss of consciousness for more than 2 minutes; clinical diagnosis of dementia, as indicated by clinical interview. Inability to give informed consent. |
| UKBiobank |  | Age 40-69 years at recruitment. Capacity to consent. Lived within 20-25 miles of one of the assessment centres. | Age 40-69 years at recruitment. Capacity to consent. Lived within 20-25 miles of one of the assessment centres. | - | - |
| Pennsylvania | UKBB | >40 years of age, MMSE > 27, a negative self-reported history of neurological or psychiatric condition, and MRI safe (e.g., no metal, claustrophobia). | Clinical diagnosis of PD. | - | - |
| Charlottesville | Neurologist | - | PD diagnosis with a motor symptom that is not (or inconsistently) responsive to oral medication. | - | - |
| **Supplementary Table 1:** Inclusion and exclusion criteria used to enroll participants at each site. **Abbreviations:** UKB, United Kingdom Brain Bank; MDS, Movement Disorder Society clinical diagnostic criteria for Parkinson's disease | | | | | |

#### *1.2 Study participants*

To address the issue of participants being assessed with either original UPDRS Part III (original UPDRS-III) or newer Movement Disorder Society UPDRS Part III scores (MDS-UPDRS-III), we used a validated formula to convert original UPDRS-III scores to predicted MDS-UPDRS-III scores. This is done by first taking the participant’s original UPDRS-III score, multiplying that value by a weighting factor given the participant’s HY stage, with the product then summed with an intercept factor, rounded to the nearest integer. For HY1 and 2, the MDS-UPDRS Part-III predicted score is calculated (UPDRS-III × 1.2) + 2.3. For HY3, the MDS-UPDRS-III predicted score is calculated (UPDRS-III × 1.2) + 1.0, and for HY4/5 the MDS-UPDRS-III predicted score is calculated (UPDRS-III × 1.1) + 7.5. Research has shown that true MDS-UPDRS Part-III scores and those derived from these formulas are highly correlated (Goetz, Stebbins, and Tilley 2012).

#### *1.3* MRI Acquisition

| **Site** | **Scanner Manufacturer (Model)** | **(T)** | **Acq.** | **Slices** | **Voxel**  **sizes (mm3)** | **Voxel**  **vols (mm3)** | **Dirs** | ***b*** | **Shells** | **b=0 scans** | **TE (ms)** | **TR (ms)** |
| --- | --- | --- | --- | --- | --- | --- | --- | --- | --- | --- | --- | --- |
| Amsterdam | GE Signa HDxT | 3T | Axial | 49-51 | 2 x 2 x 2.4 | 9.60 | 30 | 1000 | 1 | 5 | 85 | 14000 |
| Campinas | Philips Achieva | 3T | Axial | 70 | 2 x 2 x 2 | 8.00 | 32 | 1000 | 1 | 1 | 61 | 8500 |
| Chang Gung | Siemens TrioTim | 3T | Axial | 64 | 2 x 2 x 2 | 8.00 | 64 | 1000 | 1 | - | 96 | 8299 |
| Christchurch | GE HDxt | 3T | Axial | 48 | 1.875 x 1.875 x 3 | 10.55 | 28 | 1000 | 1 | 4 | 86 | 13000 |
| Cogtips | General Electric | 3T | Axial | 56 | 2.5 x 2.5 x 2.5 | 15.63 | 24 | 1000 | 1 | 7 | 81 | 7350 |
| Graz | Siemens TrioTim | 3T | Axial | 50 | 1.95 x 1.95 x 2.5 | 9.51 | 12 | 1000 | 1 | 1 | 95 | 6700 |
| NWE | Philips Achieva | 3T | Axial | 30 | 1.8 x 1.8 x 5 | 16.15 | 7 | 1000 | 1 | 1 | 68 | 3000 |
| Oxford | Siemens Trio Tim | 3T | Axial | 60 | 2 x 2 x 2 | 8.00 | 60 | 1000 | 1 | 5 | 94 | 9300 |
| PPMI | Siemens TrioTim | 3T | - | 72 | 2 x 2 x 2 | 8.00 | 65 | 1000 | 1 | 1 | 90 | 8000 |
| Radboud | Siemens Skyra | 3T | - | 64 | 2.2 x 2.2 x 2.2 | 10.65 | 68 | 1000 | 1 | 8 | 89 | 8200 |
| Rome | Siemens Allegra | 3T | Axial | 80 | 1.8 x 1.8 x 1.8 | 5.83 | 30 | 1000 | 1 | 2 | 89 | 8500 |
| Stanford | GE | 3T | - | 63 | 0.85 x 0.85 x 2 | 1.45 | 26 | 1000 | 1 | 1 | 0.002 | 0.006 |
| Stellenbosch | Siemens Skyra | 3T | T > C8.0 > S1.8 | 70 | 2 x 2 x 2 | 8.00 | 30 | 1000 | 1 | 1 | 92 | 9800 |
| UCSF | Siemens Skyra | 3T | Sagittal | 61 | 2 x 2 x 2 | 8.00 | 65 | 1000 | 1 | 1 | 0.073 | 2300 |
| UK Biobank | Siemens Skyra | 3T | Axial | 72 | 2 x 2 x 2 | 8.00 | 50 | 1000 | 1 | 5 | 92 | 3600 |
| UPenn | Siemens TrioTim | 3T | - | 57-70 | 1.8 x 1.8 x 1.8 | 5.83 | 35 | 1000 | 1 | 5 | 0.082 | 8100 |
| UVA | Siemens TrioTim; Skyra | 3T | Axial | 30-55 | 1.8 x 1.8 x 1.8 | 5.83 | 60-80 | 1000 | 1 | 1-4 | 0.069-0.01 | - |
| **Supplementary Table 2:** Diffusion MR imaging acquisition parameters for each site. **Abbreviations:** Dirs, gradient directions; | | | | | | | | | | | | |

#### 1.4 Effect Sizes

For between-group effects, we used a partial Cohen’s *d* effect sizes calculated for each regions-of-interest based on the estimated marginal means (after adjusting for covariates),

$d = \frac{t(n_{1} + n_{2})}{\sqrt{n_{1}n_{2}} \sqrt{df}}$

where *t* is t-value from the between-group comparison, *n*_1_ and *n*_2_ are the numbers of observations in each group, and *df* is the degrees of freedom. Values were interpreted according to the following criteria: small *d* = 0.20–0.49; medium *d* = 0.50–0.79; large *d* ≥ 0.8 (Cohen, 1988). For between-group analyses throughout the text and figures, positive effect size values correspond to PD patients having higher values than controls, whereas negative effect size values correspond to PD participants having lower values relative to controls. We also show the precision of these effect size estimates using confidence intervals calculated as 95% CI = *d* ± 1.96 × *se*, where *d* represents the effect size and *se* is the asymptotic standard error for the effect size (Nakagawa & Cuthill, 2007). For within-group investigations of the association between DTI metrics and clinical variables, Pearson partial correlation coefficients

$r = \frac{t}{\sqrt{t^{2} + df}}$

are used to represent the directionality and strength of the association, where *t* is the value of the t-test and *df* is the degrees of freedom.

#### 1.5 Demographics: HY Subgroups and Controls

Oneway ANOVAs comparing the HY subgroups and controls demonstrated significant differences across the groups in:

- *Age*: [F(5, 2197) = 24.4, *p*<0.001)]
- *MMSE*: [F(5, 652) = 27.61, *p*<0.001)]
- *MoCA*: [F(5, 1070) = 72.0, *p*<0.001)]

A Chi-square test for independence was used to compare proportions of males and females between the HY subgroups and controls. We found:

- *Sex:* A significant difference in the proportions of males and females across the groups [𝑋^2^(1) = 40.28, *p*<0.001].

Oneway ANOVAs comparing just HY subgroups demonstrated significant differences across the groups in:

- *Disease duration*: [F(4,1243) = 133.9, *p*<0.001)]
- *Age- at-onset*: [F(4, 1251) = , *p*<0.001]
- *MDS-UPDRS-III (ON)*: [F(4,702) = 132.92, *p*<0.001)]
- *MDS-UPDRS (OFF)*: [F(4, 575) = 179.9, *p*<0.001)]

Post-hoc analyses using Tukey’s Test with a family-wise error rate of 0.05 revealed the following significant between-group differences in demographic characteristics:

- *Age*: Controls were significantly older than HY1 participants, and significantly younger than HY2, HY3 and HY4 participants. HY1 participants were significantly younger than HY2, HY3 and HY4 participants, while HY2 participants were significantly younger than HY3 participants (*p*<0.05).
- *MMSE*: Controls had significantly higher MMSE scores than HY1, HY2, HY3 and HY4/5 participants (*p*<0.05).
- *MoCA*: Controls had significantly higher MoCA scores than HY1, HY2, HY3 and HY4/5 participants. HY1 participants had significantly higher MoCA scores than HY3 and HY4/5 scores. HY2 had significantly higher MoCA scors than HY2 and HY4/5, while HY3 had significantly higher MoCA scores than HY4/5 participants (*p*<0.05).
- *DURILL*: HY1 participants had significantly shorter disease duration than HY2, HY3 and HY4/5 participants. HY2 participants had significantly shorter disease duration than HY3 and HY4/5 participants, and HY3 participants had significantly shorter disease duration than HY4/5 participants (*p*<0.05).
- AAO: HY1 participants had an earlier age- at-onset than HY2 participants, while HY2 participants had a later age- at-onset than HY3 and HY4/5 participants (*p*<0.05).
- *MDS-UPDRS-III (ON)*: HY1 had significantly lower MDS-UPDRS-III ON scores than HY2, HY3 and HY4/5 participants. HY2 had significantly lower MDS-UPDRS-III ON scores than HY3 and HY4/5, and HY3 had significantly lower scores than HY4/5 participants (*p*<0.05).
- *MDS-UPDRS-III (OFF)*: HY1 had significantly lower MDS-UPDRS-III OFF scores than HY2, HY3 and HY4/5 participants. HY2 had significantly lower MDS-UPDRS-III OFF scores than HY3 and HY4/5, and HY3 had significantly lower scores than HY4/5 participants (*p*<0.05).

Post-hoc chi-square tests for independence were used to compare proportions of males and females between HY subgroups and controls. We found:

- Controls & HY1: Significant difference in the proportions of males and females across Controls and HY1 participants [𝑋^2^(1) = 6.25, *p*=0.012].
- Controls & HY2: Significant difference in the proportions of males and females across Controls and HY2 participants [𝑋^2^(1) = 36.81, *p*<0.001].
- Controls & HY3: Significant difference in the proportions of males and females across Controls and HY3 participants [𝑋^2^(1) = 9.05, *p*=0.003].
- Controls & HY4/5: No significant difference in the proportions of males and females across controls and HY4/5 participants [𝑋^2^(1) = 0.14, *p*=0.704].
- HY1 & HY2: No significant difference in the proportions of males and females across HY1 and HY2 participants [𝑋^2^(1) = 3.08, *p*=0.079].
- HY1 & HY3: No significant difference in the proportions of males and females across HY1 and HY3 participants [𝑋^2^(1) = 0.28, *p*=0.595].
- HY1 & HY4/5: No significant difference in the proportions of males and females across HY1 and HY4/5 participants [𝑋^2^(1) = 0.61, *p*=0.435].
- HY2 & HY3: No significant difference in the proportions of males and females across HY2 and HY3 participants [𝑋^2^(1) = 0.73, *p*=0.392].
- HY2 & HY4/5: Significant difference in the proportions of males and females across HY2 and HY4/5 participants [𝑋^2^(1) = 3.85, *p*=0.0498].
- HY3 & HY4/5: No significant difference in the proportions of males and females across HY3 and HY4/5 participants [𝑋^2^(1) = 1.40, *p*=0.238].

#### 1.6 Demographics: Total PD and Controls

Independent samples *t*-tests were used to compare the mean age, MMSE and MoCA scores between the Total PD group and controls. We found that:

- *Age*: Control participants were younger than the Total PD cohort [t(2537)=-5.2, *p*<0.001]
- *MMSE*: Total PD cohort had lower scores on the MMSE [t(689) = -6.92, *p*<0.001]
- *MoCA*: Total PD cohort had lower scores on the MoCA [t(1207) = -9.12, *p*<0.001]

A Chi-square test for independence was used to compare proportions of males and females between the Total PD group and controls. We found:

- *Sex:* A significant difference in the proportions of males and females across the two groups [𝑋^2^(1) = 44.41, *p*<0.001].

### **2. Supplementary Results**

#### 2.1 Between-group Differences in White Matter Microstructural Metrics

##### 2.1.1 FA: Stratification by HY Stage

| **HY1 & Controls** | ***p*-value** | **Degrees of Freedom** | **Partial Cohen’s *d*** | **Standard Error** | **Lower CI** | **Upper CI** | ***p*_FDR_** |
| --- | --- | --- | --- | --- | --- | --- | --- |
| **Entire WM FA** | 0.00 | 1139 | 0.304 | 0.06 | 0.19 | 0.42 | 3.07 𝗑 10^-4^ |
| **ACR FA** | 0.01 | 1139 | 0.181 | 0.06 | 0.07 | 0.30 | 4.22 𝗑 10^-2^ |
| **SCR FA** | 0.04 | 1139 | 0.143 | 0.06 | 0.03 | 0.26 | 7.52 𝗑 10^-2^ |
| **PCR FA** | 0.15 | 1139 | 0.101 | 0.06 | -0.01 | 0.22 | 1.93 𝗑 10^-1^ |
| **ALIC FA** | 0.01 | 1139 | 0.188 | 0.06 | 0.07 | 0.30 | 4.22 𝗑 10^-2^ |
| **PLIC FA** | 0.02 | 1139 | 0.168 | 0.06 | 0.05 | 0.28 | 5.07 𝗑 10^-2^ |
| **RLIC FA** | 0.01 | 1139 | 0.194 | 0.06 | 0.08 | 0.31 | 4.22 𝗑 10^-2^ |
| **EC FA** | 0.04 | 1139 | 0.147 | 0.06 | 0.03 | 0.26 | 7.52 𝗑 10^-2^ |
| **FX FA** | 0.30 | 1139 | -0.072 | 0.06 | -0.19 | 0.04 | 3.16 𝗑 10^-1^ |
| **FXST FA** | 0.30 | 1139 | 0.072 | 0.06 | -0.04 | 0.19 | 3.16 𝗑 10^-1^ |
| **PTR FA** | 0.14 | 1139 | 0.104 | 0.06 | -0.01 | 0.22 | 1.93 𝗑 10^-1^ |
| **GCC FA** | 0.01 | 1139 | 0.186 | 0.06 | 0.07 | 0.30 | 4.22 𝗑 10^-2^ |
| **BCC FA** | 0.03 | 1139 | 0.151 | 0.06 | 0.04 | 0.27 | 7.40 𝗑 10^-2^ |
| **SCC FA** | 0.04 | 1139 | 0.145 | 0.06 | 0.03 | 0.26 | 7.52 𝗑 10^-2^ |
| **CGC FA** | 0.62 | 1139 | 0.035 | 0.06 | -0.08 | 0.15 | 6.18 𝗑 10^-1^ |
| **CGH FA** | 0.18 | 1139 | 0.093 | 0.06 | -0.02 | 0.21 | 2.09 𝗑 10^-1^ |
| **CST FA** | 0.02 | 1139 | 0.157 | 0.06 | 0.04 | 0.27 | 6.60 𝗑 10^-2^ |
| **SFO FA** | 0.05 | 1139 | 0.138 | 0.06 | 0.02 | 0.25 | 8.04 𝗑 10^-2^ |
| **SLF FA** | 0.07 | 1139 | 0.128 | 0.06 | 0.01 | 0.24 | 1.06 𝗑 10^-1^ |
| **SS FA** | 0.02 | 1139 | 0.168 | 0.06 | 0.05 | 0.28 | 5.07 𝗑 10^-2^ |
| **TAP FA** | 0.16 | 1139 | 0.097 | 0.06 | -0.02 | 0.21 | 1.99 𝗑 10^-1^ |
| **UNC FA** | 0.15 | 1139 | 0.101 | 0.06 | -0.01 | 0.22 | 1.93 𝗑 10^-1^ |

| **HY2 & Controls** | ***p*-value** | **Degrees of Freedom** | **Partial Cohen’s *d*** | **Standard Error** | **Lower CI** | **Upper CI** | ***p*_FDR_** |
| --- | --- | --- | --- | --- | --- | --- | --- |
| **Entire WM FA** | 0.74 | 1606 | 0.016 | 0.05 | -0.08 | 0.11 | 9.03 𝗑 10^-1^ |
| **ACR FA** | 0.49 | 1606 | -0.035 | 0.05 | -0.13 | 0.06 | 9.03 𝗑 10^-1^ |
| **SCR FA** | 0.07 | 1606 | 0.092 | 0.05 | 0.00 | 0.19 | 7.19 𝗑 10^-1^ |
| **PCR FA** | 0.87 | 1606 | 0.008 | 0.05 | -0.09 | 0.11 | 9.07 𝗑 10^-1^ |
| **ALIC FA** | 0.68 | 1606 | 0.021 | 0.05 | -0.08 | 0.12 | 9.03 𝗑 10^-1^ |
| **PLIC FA** | 0.18 | 1606 | 0.068 | 0.05 | -0.03 | 0.16 | 9.03 𝗑 10^-1^ |
| **RLIC FA** | 0.49 | 1606 | 0.034 | 0.05 | -0.06 | 0.13 | 9.03 𝗑 10^-1^ |
| **EC FA** | 0.82 | 1606 | -0.011 | 0.05 | -0.11 | 0.09 | 9.03 𝗑 10^-1^ |
| **FX FA** | 0.00 | 1606 | -0.263 | 0.05 | -0.36 | -0.17 | 3.80 𝗑 10^-6^ |
| **FXST FA** | 0.33 | 1606 | -0.049 | 0.05 | -0.15 | 0.05 | 9.03 𝗑 10^-1^ |
| **PTR FA** | 0.56 | 1606 | -0.029 | 0.05 | -0.13 | 0.07 | 9.03 𝗑 10^-1^ |
| **GCC FA** | 0.71 | 1606 | -0.019 | 0.05 | -0.12 | 0.08 | 9.03 𝗑 10^-1^ |
| **BCC FA** | 0.92 | 1606 | 0.005 | 0.05 | -0.09 | 0.10 | 9.17 𝗑 10^-1^ |
| **SCC FA** | 0.34 | 1606 | 0.048 | 0.05 | -0.05 | 0.15 | 9.03 𝗑 10^-1^ |
| **CGC FA** | 0.26 | 1606 | -0.057 | 0.05 | -0.15 | 0.04 | 9.03 𝗑 10^-1^ |
| **CGH FA** | 0.58 | 1606 | -0.028 | 0.05 | -0.13 | 0.07 | 9.03 𝗑 10^-1^ |
| **CST FA** | 0.25 | 1606 | 0.058 | 0.05 | -0.04 | 0.16 | 9.03 𝗑 10^-1^ |
| **SFO FA** | 0.41 | 1606 | 0.041 | 0.05 | -0.06 | 0.14 | 9.03 𝗑 10^-1^ |
| **SLF FA** | 0.73 | 1606 | 0.017 | 0.05 | -0.08 | 0.11 | 9.03 𝗑 10^-1^ |
| **SS FA** | 0.82 | 1606 | -0.012 | 0.05 | -0.11 | 0.09 | 9.03 𝗑 10^-1^ |
| **TAP FA** | 0.77 | 1606 | 0.014 | 0.05 | -0.08 | 0.11 | 9.03 𝗑 10^-1^ |
| **UNC FA** | 0.28 | 1606 | -0.054 | 0.05 | -0.15 | 0.04 | 9.03 𝗑 10^-1^ |

| **HY3 & Controls** | ***p*-value** | **Degrees of Freedom** | **Partial Cohen’s *d*** | **Standard Error** | **Lower CI** | **Upper CI** | ***p*_FDR_** |
| --- | --- | --- | --- | --- | --- | --- | --- |
| **Entire WM FA** | 0.00 | 1085 | -0.240 | 0.06 | -0.36 | -0.12 | 5.94 𝗑 10^-3^ |
| **ACR FA** | 0.00 | 1085 | -0.326 | 0.06 | -0.44 | -0.21 | 2.17 𝗑 10^-4^ |
| **SCR FA** | 0.18 | 1085 | -0.103 | 0.06 | -0.22 | 0.02 | 2.43 𝗑 10^-1^ |
| **PCR FA** | 0.01 | 1085 | -0.202 | 0.06 | -0.32 | -0.08 | 1.95 𝗑 10^-2^ |
| **ALIC FA** | 0.12 | 1085 | -0.117 | 0.06 | -0.24 | 0.00 | 2.21 𝗑 10^-1^ |
| **PLIC FA** | 0.60 | 1085 | 0.039 | 0.06 | -0.08 | 0.16 | 6.37 𝗑 10^-1^ |
| **RLIC FA** | 0.14 | 1085 | -0.112 | 0.06 | -0.23 | 0.01 | 2.21 𝗑 10^-1^ |
| **EC FA** | 0.00 | 1085 | -0.260 | 0.06 | -0.38 | -0.14 | 3.86 𝗑 10^-3^ |
| **FX FA** | 0.00 | 1085 | -0.337 | 0.06 | -0.46 | -0.22 | 2.17 𝗑 10^-4^ |
| **FXST FA** | 0.00 | 1085 | -0.236 | 0.06 | -0.35 | -0.12 | 6.22 𝗑 10^-3^ |
| **PTR FA** | 0.00 | 1085 | -0.215 | 0.06 | -0.33 | -0.10 | 1.32 𝗑 10^-2^ |
| **GCC FA** | 0.02 | 1085 | -0.181 | 0.06 | -0.30 | -0.06 | 3.83 𝗑 10^-2^ |
| **BCC FA** | 0.27 | 1085 | -0.084 | 0.06 | -0.20 | 0.03 | 3.47 𝗑 10^-1^ |
| **SCC FA** | 0.29 | 1085 | -0.081 | 0.06 | -0.20 | 0.04 | 3.52 𝗑 10^-1^ |
| **CGC FA** | 0.05 | 1085 | -0.149 | 0.06 | -0.27 | -0.03 | 1.00 𝗑 10^-1^ |
| **CGH FA** | 0.16 | 1085 | -0.106 | 0.06 | -0.22 | 0.01 | 2.37 𝗑 10^-1^ |
| **CST FA** | 0.94 | 1085 | -0.006 | 0.06 | -0.12 | 0.11 | 9.37 𝗑 10^-1^ |
| **SFO FA** | 0.14 | 1085 | -0.113 | 0.06 | -0.23 | 0.00 | 2.21 𝗑 10^-1^ |
| **SLF FA** | 0.00 | 1085 | -0.242 | 0.06 | -0.36 | -0.12 | 5.94 𝗑 10^-3^ |
| **SS FA** | 0.00 | 1085 | -0.258 | 0.06 | -0.38 | -0.14 | 3.86 𝗑 10^-3^ |
| **TAP FA** | 0.61 | 1085 | -0.039 | 0.06 | -0.16 | 0.08 | 6.37 𝗑 10^-1^ |
| **UNC FA** | 0.34 | 1085 | -0.073 | 0.06 | -0.19 | 0.05 | 3.93 𝗑 10^-1^ |

| **HY4/5 & Controls** | ***p*-value** | **Degrees of Freedom** | **Partial Cohen’s *d*** | **Standard Error** | **Lower CI** | **Upper CI** | ***p*_FDR_** |
| --- | --- | --- | --- | --- | --- | --- | --- |
| **Entire WM FA** | 0.00 | 940 | -0.743 | 0.07 | -0.87 | -0.61 | 1.04 𝗑 10^-8^ |
| **ACR FA** | 0.00 | 940 | -0.575 | 0.07 | -0.70 | -0.45 | 4.65 𝗑 10^-6^ |
| **SCR FA** | 0.00 | 940 | -0.402 | 0.07 | -0.53 | -0.27 | 1.18 𝗑 10^-3^ |
| **PCR FA** | 0.00 | 940 | -0.482 | 0.07 | -0.61 | -0.35 | 1.14 𝗑 10^-4^ |
| **ALIC FA** | 0.00 | 940 | -0.606 | 0.07 | -0.74 | -0.48 | 1.61 𝗑 10^-6^ |
| **PLIC FA** | 0.02 | 940 | -0.289 | 0.06 | -0.42 | -0.16 | 1.95 𝗑 10^-2^ |
| **RLIC FA** | 0.03 | 940 | -0.268 | 0.06 | -0.40 | -0.14 | 2.89 𝗑 10^-2^ |
| **EC FA** | 0.00 | 940 | -0.788 | 0.07 | -0.92 | -0.66 | 1.62 𝗑 10^-9^ |
| **FX FA** | 0.00 | 940 | -1.013 | 0.07 | -1.15 | -0.88 | 6.15 𝗑 10^-15^ |
| **FXST FA** | 0.00 | 940 | -0.555 | 0.07 | -0.68 | -0.43 | 9.53 𝗑 10^-6^ |
| **PTR FA** | 0.00 | 940 | -0.691 | 0.07 | -0.82 | -0.56 | 6.80 𝗑 10^-8^ |
| **GCC FA** | 0.00 | 940 | -0.690 | 0.07 | -0.82 | -0.56 | 6.80 𝗑 10^-8^ |
| **BCC FA** | 0.00 | 940 | -0.669 | 0.07 | -0.80 | -0.54 | 1.48 𝗑 10^-7^ |
| **SCC FA** | 0.00 | 940 | -0.511 | 0.07 | -0.64 | -0.38 | 4.53 𝗑 10^-5^ |
| **CGC FA** | 0.00 | 940 | -0.718 | 0.07 | -0.85 | -0.59 | 2.70 𝗑 10^-8^ |
| **CGH FA** | 0.07 | 940 | -0.224 | 0.06 | -0.35 | -0.10 | 6.58 𝗑 10^-2^ |
| **CST FA** | 0.00 | 940 | -0.345 | 0.07 | -0.47 | -0.22 | 5.29 𝗑 10^-3^ |
| **SFO FA** | 0.00 | 940 | -0.586 | 0.07 | -0.72 | -0.46 | 3.32 𝗑 10^-6^ |
| **SLF FA** | 0.00 | 940 | -0.607 | 0.07 | -0.74 | -0.48 | 1.61 𝗑 10^-6^ |
| **SS FA** | 0.00 | 940 | -0.443 | 0.07 | -0.57 | -0.31 | 3.70 𝗑 10^-4^ |
| **TAP FA** | 0.00 | 940 | -0.450 | 0.07 | -0.58 | -0.32 | 3.09 𝗑 10^-4^ |
| **UNC FA** | 0.00 | 940 | -0.624 | 0.07 | -0.75 | -0.49 | 9.46 𝗑 10^-7^ |

##### 2.1.2 FA: Stratification by HY Stage - Matched Control Samples

| **HY1 & Matched Controls** | ***p*-value** | **Degrees of Freedom** | **Partial Cohen’s *d*** | **Standard Error** | **Lower CI** | **Upper CI** | ***p*_FDR_** |
| --- | --- | --- | --- | --- | --- | --- | --- |
| **Entire WM FA** | 0.00 | 529 | 0.271 | 0.09 | 0.10 | 0.44 | 4.27 𝗑 10^-2^ |
| **ACR FA** | 0.19 | 529 | 0.114 | 0.09 | -0.05 | 0.28 | 3.59 𝗑 10^-1^ |
| **SCR FA** | 0.01 | 529 | 0.241 | 0.09 | 0.07 | 0.41 | 4.28 𝗑 10^-2^ |
| **PCR FA** | 0.11 | 529 | 0.141 | 0.09 | -0.03 | 0.31 | 2.92 𝗑 10^-1^ |
| **ALIC FA** | 0.01 | 529 | 0.243 | 0.09 | 0.08 | 0.41 | 4.28 𝗑 10^-2^ |
| **PLIC FA** | 0.01 | 529 | 0.231 | 0.09 | 0.06 | 0.40 | 4.55 𝗑 10^-2^ |
| **RLIC FA** | 0.02 | 529 | 0.204 | 0.09 | 0.04 | 0.37 | 8.60 𝗑 10^-2^ |
| **EC FA** | 0.06 | 529 | 0.163 | 0.09 | 0.00 | 0.33 | 1.91 𝗑 10^-1^ |
| **FX FA** | 0.15 | 529 | -0.125 | 0.09 | -0.29 | 0.04 | 3.36 𝗑 10^-1^ |
| **FXST FA** | 0.94 | 529 | -0.007 | 0.09 | -0.17 | 0.16 | 9.83 𝗑 10^-1^ |
| **PTR FA** | 0.48 | 529 | 0.062 | 0.09 | -0.11 | 0.23 | 5.44 𝗑 10^-1^ |
| **GCC FA** | 0.21 | 529 | 0.109 | 0.09 | -0.06 | 0.28 | 3.59 𝗑 10^-1^ |
| **BCC FA** | 0.28 | 529 | 0.095 | 0.09 | -0.07 | 0.26 | 4.35 𝗑 10^-1^ |
| **SCC FA** | 0.49 | 529 | 0.059 | 0.09 | -0.11 | 0.23 | 5.44 𝗑 10^-1^ |
| **CGC FA** | 1.00 | 529 | 0.000 | 0.09 | -0.17 | 0.17 | 9.98 𝗑 10^-1^ |
| **CGH FA** | 0.37 | 529 | 0.078 | 0.09 | -0.09 | 0.25 | 5.11 𝗑 10^-1^ |
| **CST FA** | 0.42 | 529 | 0.070 | 0.09 | -0.10 | 0.24 | 5.15 𝗑 10^-1^ |
| **SFO FA** | 0.15 | 529 | 0.127 | 0.09 | -0.04 | 0.29 | 3.36 𝗑 10^-1^ |
| **SLF FA** | 0.33 | 529 | 0.084 | 0.09 | -0.08 | 0.25 | 4.89 𝗑 10^-1^ |
| **SS FA** | 0.05 | 529 | 0.169 | 0.09 | 0.00 | 0.34 | 1.91 𝗑 10^-1^ |
| **TAP FA** | 0.20 | 529 | 0.112 | 0.09 | -0.06 | 0.28 | 3.59 𝗑 10^-1^ |
| **UNC FA** | 0.42 | 529 | 0.070 | 0.09 | -0.10 | 0.24 | 5.15 𝗑 10^-1^ |

| **HY2 & Matched Controls** | ***p*-value** | **Degrees of Freedom** | **Partial Cohen’s *d*** | **Standard Error** | **Lower CI** | **Upper CI** | ***p*_FDR_** |
| --- | --- | --- | --- | --- | --- | --- | --- |
| **Entire WM FA** | 0.85 | 1463 | 0.010 | 0.05 | -0.09 | 0.11 | 9.84 𝗑 10^-1^ |
| **ACR FA** | 0.46 | 1463 | -0.039 | 0.05 | -0.14 | 0.06 | 9.43 𝗑 10^-1^ |
| **SCR FA** | 0.08 | 1463 | 0.093 | 0.05 | -0.01 | 0.20 | 8.25 𝗑 10^-1^ |
| **PCR FA** | 0.99 | 1463 | -0.001 | 0.05 | -0.10 | 0.10 | 9.90 𝗑 10^-1^ |
| **ALIC FA** | 0.85 | 1463 | 0.010 | 0.05 | -0.09 | 0.11 | 9.84 𝗑 10^-1^ |
| **PLIC FA** | 0.16 | 1463 | 0.074 | 0.05 | -0.03 | 0.18 | 9.43 𝗑 10^-1^ |
| **RLIC FA** | 0.67 | 1463 | 0.022 | 0.05 | -0.08 | 0.12 | 9.84 𝗑 10^-1^ |
| **EC FA** | 0.86 | 1463 | -0.009 | 0.05 | -0.11 | 0.09 | 9.84 𝗑 10^-1^ |
| **FX FA** | 0.00 | 1463 | -0.279 | 0.05 | -0.38 | -0.18 | 2.32 𝗑 10^-6^ |
| **FXST FA** | 0.32 | 1463 | -0.052 | 0.05 | -0.15 | 0.05 | 9.43 𝗑 10^-1^ |
| **PTR FA** | 0.39 | 1463 | -0.045 | 0.05 | -0.15 | 0.06 | 9.43 𝗑 10^-1^ |
| **GCC FA** | 0.68 | 1463 | -0.021 | 0.05 | -0.12 | 0.08 | 9.84 𝗑 10^-1^ |
| **BCC FA** | 0.89 | 1463 | -0.007 | 0.05 | -0.11 | 0.09 | 9.84 𝗑 10^-1^ |
| **SCC FA** | 0.40 | 1463 | 0.044 | 0.05 | -0.06 | 0.15 | 9.43 𝗑 10^-1^ |
| **CGC FA** | 0.25 | 1463 | -0.061 | 0.05 | -0.16 | 0.04 | 9.43 𝗑 10^-1^ |
| **CGH FA** | 0.64 | 1463 | -0.024 | 0.05 | -0.13 | 0.08 | 9.84 𝗑 10^-1^ |
| **CST FA** | 0.36 | 1463 | 0.047 | 0.05 | -0.05 | 0.15 | 9.43 𝗑 10^-1^ |
| **SFO FA** | 0.47 | 1463 | 0.038 | 0.05 | -0.06 | 0.14 | 9.43 𝗑 10^-1^ |
| **SLF FA** | 0.82 | 1463 | 0.012 | 0.05 | -0.09 | 0.11 | 9.84 𝗑 10^-1^ |
| **SS FA** | 0.80 | 1463 | -0.013 | 0.05 | -0.12 | 0.09 | 9.84 𝗑 10^-1^ |
| **TAP FA** | 0.96 | 1463 | -0.002 | 0.05 | -0.10 | 0.10 | 9.90 𝗑 10^-1^ |
| **UNC FA** | 0.24 | 1463 | -0.061 | 0.05 | -0.16 | 0.04 | 9.43 𝗑 10^-1^ |

| **HY3 & Matched Controls** | ***p*-value** | **Degrees of Freedom** | **Partial Cohen’s *d*** | **Standard Error** | **Lower CI** | **Upper CI** | ***p*_FDR_** |
| --- | --- | --- | --- | --- | --- | --- | --- |
| **Entire WM FA** | 0.04 | 420 | -0.203 | 0.10 | -0.39 | -0.02 | 1.40 𝗑 10^-1^ |
| **ACR FA** | 0.01 | 420 | -0.265 | 0.10 | -0.45 | -0.08 | 5.10 𝗑 10^-2^ |
| **SCR FA** | 0.58 | 420 | -0.054 | 0.10 | -0.24 | 0.13 | 7.52 𝗑 10^-1^ |
| **PCR FA** | 0.25 | 420 | -0.113 | 0.10 | -0.30 | 0.07 | 4.99 𝗑 10^-1^ |
| **ALIC FA** | 0.57 | 420 | -0.055 | 0.10 | -0.24 | 0.13 | 7.52 𝗑 10^-1^ |
| **PLIC FA** | 0.41 | 420 | 0.080 | 0.10 | -0.11 | 0.27 | 6.05 𝗑 10^-1^ |
| **RLIC FA** | 0.75 | 420 | -0.031 | 0.10 | -0.22 | 0.16 | 8.69 𝗑 10^-1^ |
| **EC FA** | 0.02 | 420 | -0.226 | 0.10 | -0.41 | -0.04 | 9.29 𝗑 10^-2^ |
| **FX FA** | 0.00 | 420 | -0.319 | 0.10 | -0.51 | -0.13 | 2.61 𝗑 10^-2^ |
| **FXST FA** | 0.02 | 420 | -0.230 | 0.10 | -0.42 | -0.04 | 9.29 𝗑 10^-2^ |
| **PTR FA** | 0.06 | 420 | -0.187 | 0.10 | -0.37 | 0.00 | 1.74 𝗑 10^-1^ |
| **GCC FA** | 0.12 | 420 | -0.154 | 0.10 | -0.34 | 0.03 | 2.83 𝗑 10^-1^ |
| **BCC FA** | 0.36 | 420 | -0.090 | 0.10 | -0.28 | 0.10 | 6.05 𝗑 10^-1^ |
| **SCC FA** | 0.65 | 420 | -0.045 | 0.10 | -0.23 | 0.14 | 7.89 𝗑 10^-1^ |
| **CGC FA** | 0.13 | 420 | -0.147 | 0.10 | -0.33 | 0.04 | 2.90 𝗑 10^-1^ |
| **CGH FA** | 0.36 | 420 | -0.089 | 0.10 | -0.28 | 0.10 | 6.05 𝗑 10^-1^ |
| **CST FA** | 0.91 | 420 | 0.012 | 0.10 | -0.18 | 0.20 | 9.44 𝗑 10^-1^ |
| **SFO FA** | 0.39 | 420 | -0.083 | 0.10 | -0.27 | 0.10 | 6.05 𝗑 10^-1^ |
| **SLF FA** | 0.06 | 420 | -0.182 | 0.10 | -0.37 | 0.01 | 1.74 𝗑 10^-1^ |
| **SS FA** | 0.00 | 420 | -0.290 | 0.10 | -0.48 | -0.10 | 3.46 𝗑 10^-2^ |
| **TAP FA** | 0.90 | 420 | -0.012 | 0.10 | -0.20 | 0.18 | 9.44 𝗑 10^-1^ |
| **UNC FA** | 0.94 | 420 | 0.007 | 0.10 | -0.18 | 0.19 | 9.44 𝗑 10^-1^ |

| **HY4/5 & Matched Controls** | ***p*-value** | **Degrees of Freedom** | **Partial Cohen’s *d*** | **Standard Error** | **Lower CI** | **Upper CI** | ***p*_FDR_** |
| --- | --- | --- | --- | --- | --- | --- | --- |
| **Entire WM FA** | 0.00 | 131 | -0.569 | 0.17 | -0.90 | -0.24 | 6.36 𝗑 10^-3^ |
| **ACR FA** | 0.00 | 131 | -0.522 | 0.17 | -0.85 | -0.19 | 8.24 𝗑 10^-3^ |
| **SCR FA** | 0.03 | 131 | -0.386 | 0.17 | -0.71 | -0.06 | 3.33 𝗑 10^-2^ |
| **PCR FA** | 0.01 | 131 | -0.449 | 0.17 | -0.77 | -0.12 | 1.79 𝗑 10^-2^ |
| **ALIC FA** | 0.00 | 131 | -0.507 | 0.17 | -0.83 | -0.18 | 9.61 𝗑 10^-3^ |
| **PLIC FA** | 0.31 | 131 | -0.180 | 0.16 | -0.50 | 0.14 | 3.05 𝗑 10^-1^ |
| **RLIC FA** | 0.09 | 131 | -0.303 | 0.17 | -0.63 | 0.02 | 9.42 𝗑 10^-2^ |
| **EC FA** | 0.00 | 131 | -0.650 | 0.17 | -0.98 | -0.32 | 3.24 𝗑 10^-3^ |
| **FX FA** | 0.00 | 131 | -1.097 | 0.18 | -1.44 | -0.75 | 1.02 𝗑 10^-7^ |
| **FXST FA** | 0.02 | 131 | -0.400 | 0.17 | -0.73 | -0.07 | 2.89 𝗑 10^-2^ |
| **PTR FA** | 0.00 | 131 | -0.535 | 0.17 | -0.86 | -0.21 | 7.40 𝗑 10^-3^ |
| **GCC FA** | 0.00 | 131 | -0.541 | 0.17 | -0.87 | -0.21 | 7.40 𝗑 10^-3^ |
| **BCC FA** | 0.01 | 131 | -0.499 | 0.17 | -0.83 | -0.17 | 1.00 𝗑 10^-2^ |
| **SCC FA** | 0.02 | 131 | -0.427 | 0.17 | -0.75 | -0.10 | 2.18 𝗑 10^-2^ |
| **CGC FA** | 0.00 | 131 | -0.617 | 0.17 | -0.95 | -0.29 | 4.22 𝗑 10^-3^ |
| **CGH FA** | 0.02 | 131 | -0.411 | 0.17 | -0.74 | -0.09 | 2.60 𝗑 10^-2^ |
| **CST FA** | 0.26 | 131 | -0.196 | 0.16 | -0.52 | 0.13 | 2.76 𝗑 10^-1^ |
| **SFO FA** | 0.00 | 131 | -0.574 | 0.17 | -0.90 | -0.25 | 6.36 𝗑 10^-3^ |
| **SLF FA** | 0.01 | 131 | -0.476 | 0.17 | -0.80 | -0.15 | 1.35 𝗑 10^-2^ |
| **SS FA** | 0.01 | 131 | -0.440 | 0.17 | -0.77 | -0.11 | 1.90 𝗑 10^-2^ |
| **TAP FA** | 0.01 | 131 | -0.450 | 0.17 | -0.78 | -0.12 | 1.79 𝗑 10^-2^ |
| **UNC FA** | 0.00 | 131 | -0.541 | 0.17 | -0.87 | -0.21 | 7.40 𝗑 10^-3^ |

##### 2.1.3 MD: Stratification by HY Stage

| **HY1 & Controls** | ***p*-value** | **Degrees of Freedom** | **Partial Cohen’s *d*** | **Standard Error** | **Lower CI** | **Upper CI** | ***p*_FDR_** |
| --- | --- | --- | --- | --- | --- | --- | --- |
| **Entire WM MD** | 0.00 | 1139 | -0.198 | 0.06 | -0.31 | -0.08 | 1.98 𝗑 10^-2^ |
| **ACR MD** | 0.08 | 1139 | -0.122 | 0.06 | -0.24 | -0.01 | 1.45 𝗑 10^-1^ |
| **SCR MD** | 0.57 | 1139 | -0.039 | 0.06 | -0.15 | 0.08 | 6.02 𝗑 10^-1^ |
| **PCR MD** | 0.22 | 1139 | -0.085 | 0.06 | -0.20 | 0.03 | 3.05 𝗑 10^-1^ |
| **ALIC MD** | 0.01 | 1139 | -0.182 | 0.06 | -0.30 | -0.07 | 3.35 𝗑 10^-2^ |
| **PLIC MD** | 0.02 | 1139 | -0.157 | 0.06 | -0.27 | -0.04 | 7.82 𝗑 10^-2^ |
| **RLIC MD** | 0.00 | 1139 | -0.272 | 0.06 | -0.39 | -0.16 | 2.26 𝗑 10^-3^ |
| **EC MD** | 0.07 | 1139 | -0.128 | 0.06 | -0.24 | -0.01 | 1.35 𝗑 10^-1^ |
| **FX MD** | 0.51 | 1139 | -0.046 | 0.06 | -0.16 | 0.07 | 6.02 𝗑 10^-1^ |
| **FXST MD** | 0.00 | 1139 | -0.220 | 0.06 | -0.34 | -0.10 | 1.02 𝗑 10^-2^ |
| **PTR MD** | 0.10 | 1139 | -0.115 | 0.06 | -0.23 | 0.00 | 1.57 𝗑 10^-1^ |
| **GCC MD** | 0.05 | 1139 | -0.135 | 0.06 | -0.25 | -0.02 | 1.17 𝗑 10^-1^ |
| **BCC MD** | 0.34 | 1139 | 0.067 | 0.06 | -0.05 | 0.18 | 4.36 𝗑 10^-1^ |
| **SCC MD** | 0.77 | 1139 | 0.020 | 0.06 | -0.10 | 0.14 | 7.74 𝗑 10^-1^ |
| **CGC MD** | 0.04 | 1139 | -0.147 | 0.06 | -0.26 | -0.03 | 9.72 𝗑 10^-2^ |
| **CGH MD** | 0.00 | 1139 | -0.217 | 0.06 | -0.33 | -0.10 | 1.02 𝗑 10^-2^ |
| **CST MD** | 0.05 | 1139 | -0.140 | 0.06 | -0.25 | -0.02 | 1.11 𝗑 10^-1^ |
| **SFO MD** | 0.57 | 1139 | -0.039 | 0.06 | -0.15 | 0.08 | 6.02 𝗑 10^-1^ |
| **SLF MD** | 0.11 | 1139 | -0.112 | 0.06 | -0.23 | 0.00 | 1.58 𝗑 10^-1^ |
| **SS MD** | 0.00 | 1139 | -0.253 | 0.06 | -0.37 | -0.14 | 3.32 𝗑 10^-3^ |
| **TAP MD** | 0.54 | 1139 | -0.043 | 0.06 | -0.16 | 0.07 | 6.02 𝗑 10^-1^ |
| **UNC MD** | 0.09 | 1139 | -0.119 | 0.06 | -0.23 | 0.00 | 1.50 𝗑 10^-1^ |

| **HY2 & Controls** | ***p*-value** | **Degrees of Freedom** | **Partial Cohen’s *d*** | **Standard Error** | **Lower CI** | **Upper CI** | ***p*_FDR_** |
| --- | --- | --- | --- | --- | --- | --- | --- |
| **Entire WM MD** | 0.58 | 1606 | -0.028 | 0.05 | -0.12 | 0.07 | 8.53 𝗑 10^-1^ |
| **ACR MD** | 0.87 | 1606 | 0.008 | 0.05 | -0.09 | 0.11 | 9.38 𝗑 10^-1^ |
| **SCR MD** | 0.83 | 1606 | -0.011 | 0.05 | -0.11 | 0.09 | 9.38 𝗑 10^-1^ |
| **PCR MD** | 0.88 | 1606 | 0.008 | 0.05 | -0.09 | 0.11 | 9.38 𝗑 10^-1^ |
| **ALIC MD** | 0.02 | 1606 | -0.119 | 0.05 | -0.22 | -0.02 | 6.41 𝗑 10^-2^ |
| **PLIC MD** | 0.01 | 1606 | -0.126 | 0.05 | -0.22 | -0.03 | 5.20 𝗑 10^-2^ |
| **RLIC MD** | 0.00 | 1606 | -0.195 | 0.05 | -0.29 | -0.10 | 1.10 𝗑 10^-3^ |
| **EC MD** | 0.53 | 1606 | -0.032 | 0.05 | -0.13 | 0.07 | 8.29 𝗑 10^-1^ |
| **FX MD** | 0.00 | 1606 | 0.152 | 0.05 | 0.05 | 0.25 | 1.39 𝗑 10^-2^ |
| **FXST MD** | 0.00 | 1606 | -0.221 | 0.05 | -0.32 | -0.12 | 2.43 𝗑 10^-4^ |
| **PTR MD** | 0.27 | 1606 | -0.055 | 0.05 | -0.15 | 0.04 | 5.43 𝗑 10^-1^ |
| **GCC MD** | 0.90 | 1606 | -0.007 | 0.05 | -0.10 | 0.09 | 9.38 𝗑 10^-1^ |
| **BCC MD** | 0.70 | 1606 | 0.020 | 0.05 | -0.08 | 0.12 | 9.38 𝗑 10^-1^ |
| **SCC MD** | 0.98 | 1606 | -0.002 | 0.05 | -0.10 | 0.10 | 9.76 𝗑 10^-1^ |
| **CGC MD** | 0.09 | 1606 | -0.086 | 0.05 | -0.18 | 0.01 | 2.34 𝗑 10^-1^ |
| **CGH MD** | 0.00 | 1606 | -0.187 | 0.05 | -0.28 | -0.09 | 1.40 𝗑 10^-3^ |
| **CST MD** | 0.17 | 1606 | -0.069 | 0.05 | -0.17 | 0.03 | 4.15 𝗑 10^-1^ |
| **SFO MD** | 0.25 | 1606 | -0.058 | 0.05 | -0.15 | 0.04 | 5.43 𝗑 10^-1^ |
| **SLF MD** | 0.51 | 1606 | -0.033 | 0.05 | -0.13 | 0.06 | 8.29 𝗑 10^-1^ |
| **SS MD** | 0.05 | 1606 | -0.100 | 0.05 | -0.20 | 0.00 | 1.47 𝗑 10^-1^ |
| **TAP MD** | 0.49 | 1606 | 0.034 | 0.05 | -0.06 | 0.13 | 8.29 𝗑 10^-1^ |
| **UNC MD** | 0.82 | 1606 | 0.012 | 0.05 | -0.09 | 0.11 | 9.38 𝗑 10^-1^ |

| **HY3 & Controls** | ***p*-value** | **Degrees of Freedom** | **Partial Cohen’s *d*** | **Standard Error** | **Lower CI** | **Upper CI** | ***p*_FDR_** |
| --- | --- | --- | --- | --- | --- | --- | --- |
| **Entire WM MD** | 0.26 | 1085 | 0.086 | 0.06 | -0.03 | 0.20 | 4.72 𝗑 10^-1^ |
| **ACR MD** | 0.00 | 1085 | 0.216 | 0.06 | 0.10 | 0.33 | 5.13 𝗑 10^-2^ |
| **SCR MD** | 0.06 | 1085 | 0.145 | 0.06 | 0.03 | 0.26 | 1.79 𝗑 10^-1^ |
| **PCR MD** | 0.05 | 1085 | 0.150 | 0.06 | 0.03 | 0.27 | 1.79 𝗑 10^-1^ |
| **ALIC MD** | 0.45 | 1085 | -0.058 | 0.06 | -0.18 | 0.06 | 5.82 𝗑 10^-1^ |
| **PLIC MD** | 0.20 | 1085 | -0.097 | 0.06 | -0.21 | 0.02 | 4.07 𝗑 10^-1^ |
| **RLIC MD** | 0.31 | 1085 | -0.077 | 0.06 | -0.19 | 0.04 | 5.28 𝗑 10^-1^ |
| **EC MD** | 0.16 | 1085 | 0.107 | 0.06 | -0.01 | 0.23 | 3.91 𝗑 10^-1^ |
| **FX MD** | 0.00 | 1085 | 0.222 | 0.06 | 0.10 | 0.34 | 5.13 𝗑 10^-2^ |
| **FXST MD** | 0.95 | 1085 | -0.005 | 0.06 | -0.12 | 0.11 | 9.84 𝗑 10^-1^ |
| **PTR MD** | 0.78 | 1085 | 0.021 | 0.06 | -0.10 | 0.14 | 9.00 𝗑 10^-1^ |
| **GCC MD** | 0.35 | 1085 | 0.071 | 0.06 | -0.05 | 0.19 | 5.53 𝗑 10^-1^ |
| **BCC MD** | 0.19 | 1085 | 0.099 | 0.06 | -0.02 | 0.22 | 4.07 𝗑 10^-1^ |
| **SCC MD** | 0.98 | 1085 | 0.002 | 0.06 | -0.12 | 0.12 | 9.84 𝗑 10^-1^ |
| **CGC MD** | 0.67 | 1085 | -0.032 | 0.06 | -0.15 | 0.09 | 8.18 𝗑 10^-1^ |
| **CGH MD** | 0.04 | 1085 | -0.160 | 0.06 | -0.28 | -0.04 | 1.79 𝗑 10^-1^ |
| **CST MD** | 0.91 | 1085 | 0.009 | 0.06 | -0.11 | 0.13 | 9.84 𝗑 10^-1^ |
| **SFO MD** | 0.04 | 1085 | 0.156 | 0.06 | 0.04 | 0.27 | 1.79 𝗑 10^-1^ |
| **SLF MD** | 0.16 | 1085 | 0.107 | 0.06 | -0.01 | 0.23 | 3.91 𝗑 10^-1^ |
| **SS MD** | 0.45 | 1085 | 0.058 | 0.06 | -0.06 | 0.18 | 5.82 𝗑 10^-1^ |
| **TAP MD** | 0.05 | 1085 | 0.150 | 0.06 | 0.03 | 0.27 | 1.79 𝗑 10^-1^ |
| **UNC MD** | 0.42 | 1085 | 0.061 | 0.06 | -0.06 | 0.18 | 5.82 𝗑 10^-1^ |

| **HY4/5 & Controls** | ***p*-value** | **Degrees of Freedom** | **Partial Cohen’s *d*** | **Standard Error** | **Lower CI** | **Upper CI** | ***p*_FDR_** |
| --- | --- | --- | --- | --- | --- | --- | --- |
| **Entire WM MD** | 0.07 | 940 | 0.222 | 0.06 | 0.10 | 0.35 | 1.36 𝗑 10^-1^ |
| **ACR MD** | 0.00 | 940 | 0.410 | 0.07 | 0.28 | 0.54 | 4.20 𝗑 10^-3^ |
| **SCR MD** | 0.00 | 940 | 0.413 | 0.07 | 0.28 | 0.54 | 4.20 𝗑 10^-3^ |
| **PCR MD** | 0.06 | 940 | 0.232 | 0.06 | 0.11 | 0.36 | 1.24 𝗑 10^-1^ |
| **ALIC MD** | 0.51 | 940 | 0.079 | 0.06 | -0.05 | 0.21 | 5.81 𝗑 10^-1^ |
| **PLIC MD** | 0.04 | 940 | -0.253 | 0.06 | -0.38 | -0.13 | 9.29 𝗑 10^-2^ |
| **RLIC MD** | 0.08 | 940 | -0.213 | 0.06 | -0.34 | -0.09 | 1.48 𝗑 10^-1^ |
| **EC MD** | 0.01 | 940 | 0.331 | 0.07 | 0.20 | 0.46 | 2.41 𝗑 10^-2^ |
| **FX MD** | 0.00 | 940 | 0.689 | 0.07 | 0.56 | 0.82 | 4.11 𝗑 10^-7^ |
| **FXST MD** | 0.98 | 940 | 0.003 | 0.06 | -0.12 | 0.13 | 9.78 𝗑 10^-1^ |
| **PTR MD** | 0.43 | 940 | -0.096 | 0.06 | -0.22 | 0.03 | 5.27 𝗑 10^-1^ |
| **GCC MD** | 0.00 | 940 | 0.380 | 0.07 | 0.25 | 0.51 | 8.03 𝗑 10^-3^ |
| **BCC MD** | 0.01 | 940 | 0.307 | 0.06 | 0.18 | 0.43 | 3.19 𝗑 10^-2^ |
| **SCC MD** | 0.75 | 940 | -0.039 | 0.06 | -0.17 | 0.09 | 7.86 𝗑 10^-1^ |
| **CGC MD** | 0.31 | 940 | -0.124 | 0.06 | -0.25 | 0.00 | 4.22 𝗑 10^-1^ |
| **CGH MD** | 0.01 | 940 | -0.325 | 0.07 | -0.45 | -0.20 | 2.41 𝗑 10^-2^ |
| **CST MD** | 0.10 | 940 | -0.202 | 0.06 | -0.33 | -0.08 | 1.63 𝗑 10^-1^ |
| **SFO MD** | 0.00 | 940 | 0.459 | 0.07 | 0.33 | 0.59 | 1.84 𝗑 10^-3^ |
| **SLF MD** | 0.42 | 940 | 0.097 | 0.06 | -0.03 | 0.22 | 5.27 𝗑 10^-1^ |
| **SS MD** | 0.15 | 940 | -0.175 | 0.06 | -0.30 | -0.05 | 2.21 𝗑 10^-1^ |
| **TAP MD** | 0.12 | 940 | 0.188 | 0.06 | 0.06 | 0.32 | 1.91 𝗑 10^-1^ |
| **UNC MD** | 0.53 | 940 | 0.077 | 0.06 | -0.05 | 0.20 | 5.81 𝗑 10^-1^ |

##### 2.1.4 MD: Stratification by HY Stage - Matched Control Samples

| **HY1 & Matched Controls** | ***p*-value** | **Degrees of Freedom** | **Partial Cohen’s *d*** | **Standard Error** | **Lower CI** | **Upper CI** | ***p*_FDR_** |
| --- | --- | --- | --- | --- | --- | --- | --- |
| **Entire WM MD** | 0.16 | 529 | -0.123 | 0.09 | -0.29 | 0.04 | 4.92 𝗑 10^-1^ |
| **ACR MD** | 0.44 | 529 | -0.067 | 0.09 | -0.23 | 0.10 | 7.03 𝗑 10^-1^ |
| **SCR MD** | 0.95 | 529 | -0.006 | 0.09 | -0.17 | 0.16 | 9.48 𝗑 10^-1^ |
| **PCR MD** | 0.64 | 529 | -0.040 | 0.09 | -0.21 | 0.13 | 7.43 𝗑 10^-1^ |
| **ALIC MD** | 0.06 | 529 | -0.162 | 0.09 | -0.33 | 0.01 | 3.49 𝗑 10^-1^ |
| **PLIC MD** | 0.18 | 529 | -0.117 | 0.09 | -0.28 | 0.05 | 4.92 𝗑 10^-1^ |
| **RLIC MD** | 0.01 | 529 | -0.217 | 0.09 | -0.39 | -0.05 | 2.00 𝗑 10^-1^ |
| **EC MD** | 0.25 | 529 | -0.099 | 0.09 | -0.27 | 0.07 | 5.60 𝗑 10^-1^ |
| **FX MD** | 0.64 | 529 | 0.041 | 0.09 | -0.13 | 0.21 | 7.43 𝗑 10^-1^ |
| **FXST MD** | 0.06 | 529 | -0.163 | 0.09 | -0.33 | 0.00 | 3.49 𝗑 10^-1^ |
| **PTR MD** | 0.41 | 529 | -0.072 | 0.09 | -0.24 | 0.10 | 7.03 𝗑 10^-1^ |
| **GCC MD** | 0.14 | 529 | -0.128 | 0.09 | -0.30 | 0.04 | 4.92 𝗑 10^-1^ |
| **BCC MD** | 0.57 | 529 | 0.050 | 0.09 | -0.12 | 0.22 | 7.35 𝗑 10^-1^ |
| **SCC MD** | 0.50 | 529 | 0.059 | 0.09 | -0.11 | 0.23 | 7.03 𝗑 10^-1^ |
| **CGC MD** | 0.37 | 529 | -0.079 | 0.09 | -0.25 | 0.09 | 7.03 𝗑 10^-1^ |
| **CGH MD** | 0.11 | 529 | -0.141 | 0.09 | -0.31 | 0.03 | 4.64 𝗑 10^-1^ |
| **CST MD** | 0.21 | 529 | -0.108 | 0.09 | -0.28 | 0.06 | 5.23 𝗑 10^-1^ |
| **SFO MD** | 0.94 | 529 | 0.006 | 0.09 | -0.16 | 0.17 | 9.48 𝗑 10^-1^ |
| **SLF MD** | 0.51 | 529 | -0.057 | 0.09 | -0.22 | 0.11 | 7.03 𝗑 10^-1^ |
| **SS MD** | 0.02 | 529 | -0.206 | 0.09 | -0.37 | -0.04 | 2.00 𝗑 10^-1^ |
| **TAP MD** | 0.73 | 529 | -0.030 | 0.09 | -0.20 | 0.14 | 8.03 𝗑 10^-1^ |
| **UNC MD** | 0.49 | 529 | -0.060 | 0.09 | -0.23 | 0.11 | 7.03 𝗑 10^-1^ |

| **HY2 & Matched Controls** | ***p*-value** | **Degrees of Freedom** | **Partial Cohen’s *d*** | **Standard Error** | **Lower CI** | **Upper CI** | ***p*_FDR_** |
| --- | --- | --- | --- | --- | --- | --- | --- |
| **Entire WM MD** | 0.66 | 1463 | -0.023 | 0.05 | -0.12 | 0.08 | 9.07 𝗑 10^-1^ |
| **ACR MD** | 0.81 | 1463 | 0.012 | 0.05 | -0.09 | 0.11 | 9.28 𝗑 10^-1^ |
| **SCR MD** | 0.89 | 1463 | -0.008 | 0.05 | -0.11 | 0.09 | 9.28 𝗑 10^-1^ |
| **PCR MD** | 0.70 | 1463 | 0.020 | 0.05 | -0.08 | 0.12 | 9.12 𝗑 10^-1^ |
| **ALIC MD** | 0.02 | 1463 | -0.125 | 0.05 | -0.23 | -0.02 | 6.08 𝗑 10^-2^ |
| **PLIC MD** | 0.01 | 1463 | -0.140 | 0.05 | -0.24 | -0.04 | 3.22 𝗑 10^-2^ |
| **RLIC MD** | 0.00 | 1463 | -0.190 | 0.05 | -0.29 | -0.09 | 2.11 𝗑 10^-3^ |
| **EC MD** | 0.48 | 1463 | -0.037 | 0.05 | -0.14 | 0.07 | 8.19 𝗑 10^-1^ |
| **FX MD** | 0.00 | 1463 | 0.154 | 0.05 | 0.05 | 0.26 | 1.83 𝗑 10^-2^ |
| **FXST MD** | 0.00 | 1463 | -0.222 | 0.05 | -0.32 | -0.12 | 5.23 𝗑 10^-4^ |
| **PTR MD** | 0.44 | 1463 | -0.040 | 0.05 | -0.14 | 0.06 | 8.07 𝗑 10^-1^ |
| **GCC MD** | 0.95 | 1463 | -0.003 | 0.05 | -0.10 | 0.10 | 9.53 𝗑 10^-1^ |
| **BCC MD** | 0.56 | 1463 | 0.031 | 0.05 | -0.07 | 0.13 | 8.74 𝗑 10^-1^ |
| **SCC MD** | 0.86 | 1463 | 0.009 | 0.05 | -0.09 | 0.11 | 9.28 𝗑 10^-1^ |
| **CGC MD** | 0.13 | 1463 | -0.078 | 0.05 | -0.18 | 0.02 | 3.70 𝗑 10^-1^ |
| **CGH MD** | 0.00 | 1463 | -0.192 | 0.05 | -0.29 | -0.09 | 2.11 𝗑 10^-3^ |
| **CST MD** | 0.19 | 1463 | -0.069 | 0.05 | -0.17 | 0.03 | 4.55 𝗑 10^-1^ |
| **SFO MD** | 0.36 | 1463 | -0.048 | 0.05 | -0.15 | 0.05 | 7.12 𝗑 10^-1^ |
| **SLF MD** | 0.64 | 1463 | -0.024 | 0.05 | -0.13 | 0.08 | 9.07 𝗑 10^-1^ |
| **SS MD** | 0.06 | 1463 | -0.097 | 0.05 | -0.20 | 0.01 | 2.03 𝗑 10^-1^ |
| **TAP MD** | 0.28 | 1463 | 0.056 | 0.05 | -0.05 | 0.16 | 6.27 𝗑 10^-1^ |
| **UNC MD** | 0.79 | 1463 | 0.014 | 0.05 | -0.09 | 0.12 | 9.28 𝗑 10^-1^ |

| **HY3 & Matched Controls** | ***p*-value** | **Degrees of Freedom** | **Partial Cohen’s *d*** | **Standard Error** | **Lower CI** | **Upper CI** | ***p*_FDR_** |
| --- | --- | --- | --- | --- | --- | --- | --- |
| **Entire WM MD** | 0.79 | 420 | 0.026 | 0.10 | -0.16 | 0.21 | 8.70 𝗑 10^-1^ |
| **ACR MD** | 0.11 | 420 | 0.156 | 0.10 | -0.03 | 0.34 | 6.03 𝗑 10^-1^ |
| **SCR MD** | 0.59 | 420 | 0.052 | 0.10 | -0.14 | 0.24 | 8.70 𝗑 10^-1^ |
| **PCR MD** | 0.56 | 420 | 0.056 | 0.10 | -0.13 | 0.24 | 8.70 𝗑 10^-1^ |
| **ALIC MD** | 0.25 | 420 | -0.112 | 0.10 | -0.30 | 0.08 | 7.93 𝗑 10^-1^ |
| **PLIC MD** | 0.23 | 420 | -0.116 | 0.10 | -0.30 | 0.07 | 7.93 𝗑 10^-1^ |
| **RLIC MD** | 0.09 | 420 | -0.168 | 0.10 | -0.36 | 0.02 | 6.03 𝗑 10^-1^ |
| **EC MD** | 0.85 | 420 | 0.018 | 0.10 | -0.17 | 0.21 | 8.70 𝗑 10^-1^ |
| **FX MD** | 0.03 | 420 | 0.216 | 0.10 | 0.03 | 0.40 | 5.97 𝗑 10^-1^ |
| **FXST MD** | 0.56 | 420 | -0.058 | 0.10 | -0.24 | 0.13 | 8.70 𝗑 10^-1^ |
| **PTR MD** | 0.53 | 420 | -0.061 | 0.10 | -0.25 | 0.13 | 8.70 𝗑 10^-1^ |
| **GCC MD** | 0.67 | 420 | 0.042 | 0.10 | -0.15 | 0.23 | 8.70 𝗑 10^-1^ |
| **BCC MD** | 0.42 | 420 | 0.080 | 0.10 | -0.11 | 0.27 | 8.70 𝗑 10^-1^ |
| **SCC MD** | 0.82 | 420 | -0.022 | 0.10 | -0.21 | 0.17 | 8.70 𝗑 10^-1^ |
| **CGC MD** | 0.35 | 420 | -0.091 | 0.10 | -0.28 | 0.10 | 8.70 𝗑 10^-1^ |
| **CGH MD** | 0.05 | 420 | -0.188 | 0.10 | -0.38 | 0.00 | 5.97 𝗑 10^-1^ |
| **CST MD** | 0.75 | 420 | 0.031 | 0.10 | -0.16 | 0.22 | 8.70 𝗑 10^-1^ |
| **SFO MD** | 0.19 | 420 | 0.129 | 0.10 | -0.06 | 0.32 | 7.93 𝗑 10^-1^ |
| **SLF MD** | 0.83 | 420 | 0.021 | 0.10 | -0.17 | 0.21 | 8.70 𝗑 10^-1^ |
| **SS MD** | 0.87 | 420 | -0.016 | 0.10 | -0.20 | 0.17 | 8.70 𝗑 10^-1^ |
| **TAP MD** | 0.47 | 420 | 0.070 | 0.10 | -0.12 | 0.26 | 8.70 𝗑 10^-1^ |
| **UNC MD** | 0.84 | 420 | -0.020 | 0.10 | -0.21 | 0.17 | 8.70 𝗑 10^-1^ |

| **HY4/5 & Matched Controls** | ***p*-value** | **Degrees of Freedom** | **Partial Cohen’s *d*** | **Standard Error** | **Lower CI** | **Upper CI** | ***p*_FDR_** |
| --- | --- | --- | --- | --- | --- | --- | --- |
| **Entire WM MD** | 0.48 | 131 | 0.122 | 0.16 | -0.20 | 0.44 | 6.67 𝗑 10^-1^ |
| **ACR MD** | 0.06 | 131 | 0.330 | 0.17 | 0.01 | 0.65 | 4.34 𝗑 10^-1^ |
| **SCR MD** | 0.04 | 131 | 0.362 | 0.17 | 0.04 | 0.69 | 4.34 𝗑 10^-1^ |
| **PCR MD** | 0.18 | 131 | 0.237 | 0.16 | -0.09 | 0.56 | 4.86 𝗑 10^-1^ |
| **ALIC MD** | 0.48 | 131 | 0.122 | 0.16 | -0.20 | 0.45 | 6.67 𝗑 10^-1^ |
| **PLIC MD** | 0.48 | 131 | -0.123 | 0.16 | -0.45 | 0.20 | 6.67 𝗑 10^-1^ |
| **RLIC MD** | 0.27 | 131 | -0.194 | 0.16 | -0.52 | 0.13 | 5.40 𝗑 10^-1^ |
| **EC MD** | 0.22 | 131 | 0.214 | 0.16 | -0.11 | 0.54 | 4.89 𝗑 10^-1^ |
| **FX MD** | 0.00 | 131 | 0.723 | 0.17 | 0.39 | 1.06 | 1.38 𝗑 10^-3^ |
| **FXST MD** | 0.96 | 131 | -0.009 | 0.16 | -0.33 | 0.31 | 9.61 𝗑 10^-1^ |
| **PTR MD** | 0.85 | 131 | -0.034 | 0.16 | -0.36 | 0.29 | 8.86 𝗑 10^-1^ |
| **GCC MD** | 0.11 | 131 | 0.279 | 0.17 | -0.04 | 0.60 | 4.76 𝗑 10^-1^ |
| **BCC MD** | 0.47 | 131 | 0.127 | 0.16 | -0.20 | 0.45 | 6.67 𝗑 10^-1^ |
| **SCC MD** | 0.77 | 131 | -0.052 | 0.16 | -0.37 | 0.27 | 8.57 𝗑 10^-1^ |
| **CGC MD** | 0.63 | 131 | -0.083 | 0.16 | -0.41 | 0.24 | 8.20 𝗑 10^-1^ |
| **CGH MD** | 0.14 | 131 | -0.257 | 0.17 | -0.58 | 0.07 | 4.76 𝗑 10^-1^ |
| **CST MD** | 0.15 | 131 | -0.252 | 0.17 | -0.58 | 0.07 | 4.76 𝗑 10^-1^ |
| **SFO MD** | 0.08 | 131 | 0.309 | 0.17 | -0.01 | 0.63 | 4.34 𝗑 10^-1^ |
| **SLF MD** | 0.73 | 131 | 0.061 | 0.16 | -0.26 | 0.38 | 8.57 𝗑 10^-1^ |
| **SS MD** | 0.34 | 131 | -0.167 | 0.16 | -0.49 | 0.16 | 6.23 𝗑 10^-1^ |
| **TAP MD** | 0.22 | 131 | 0.217 | 0.16 | -0.11 | 0.54 | 4.89 𝗑 10^-1^ |
| **UNC MD** | 0.78 | 131 | 0.049 | 0.16 | -0.27 | 0.37 | 8.57 𝗑 10^-1^ |

##### 2.1.5 AD: Stratification by HY Stage

| **HY1 & Controls** | ***p*-value** | **Degrees of Freedom** | **Partial Cohen’s *d*** | **Standard Error** | **Lower CI** | **Upper CI** | ***p*_FDR_** |
| --- | --- | --- | --- | --- | --- | --- | --- |
| **Entire WM AD** | 0.09 | 1139 | -0.119 | 0.059 | -0.23 | 0.00 | 1.95 𝗑 10^-1^ |
| **ACR AD** | 0.55 | 1139 | -0.042 | 0.059 | -0.16 | 0.07 | 7.09 𝗑 10^-1^ |
| **SCR AD** | 0.65 | 1139 | 0.031 | 0.059 | -0.08 | 0.15 | 7.57 𝗑 10^-1^ |
| **PCR AD** | 0.36 | 1139 | -0.064 | 0.059 | -0.18 | 0.05 | 4.93 𝗑 10^-1^ |
| **ALIC AD** | 0.02 | 1139 | -0.160 | 0.059 | -0.27 | -0.04 | 8.14 𝗑 10^-2^ |
| **PLIC AD** | 0.25 | 1139 | -0.080 | 0.059 | -0.19 | 0.04 | 4.29 𝗑 10^-1^ |
| **RLIC AD** | 0.00 | 1139 | -0.251 | 0.059 | -0.37 | -0.14 | 4.49 𝗑 10^-3^ |
| **EC AD** | 0.21 | 1139 | -0.087 | 0.059 | -0.20 | 0.03 | 3.86 𝗑 10^-1^ |
| **FX AD** | 0.08 | 1139 | -0.124 | 0.059 | -0.24 | -0.01 | 1.86 𝗑 10^-1^ |
| **FXST AD** | 0.00 | 1139 | -0.213 | 0.059 | -0.33 | -0.10 | 1.18 𝗑 10^-2^ |
| **PTR AD** | 0.30 | 1139 | -0.073 | 0.059 | -0.19 | 0.04 | 4.35 𝗑 10^-1^ |
| **GCC AD** | 0.62 | 1139 | -0.035 | 0.059 | -0.15 | 0.08 | 7.53 𝗑 10^-1^ |
| **BCC AD** | 0.00 | 1139 | 0.210- | 0.059 | 0.09 | 0.33 | 1.18 𝗑 10^-2^ |
| **SCC AD** | 0.06 | 1139 | 0.132 | 0.059 | 0.02 | 0.25 | 1.60 𝗑 10^-1^ |
| **CGC AD** | 0.05 | 1139 | -0.135 | 0.059 | -0.25 | -0.02 | 1.60 𝗑 10^-1^ |
| **CGH AD** | 0.00 | 1139 | -0.247 | 0.059 | -0.36 | -0.13 | 4.49 𝗑 10^-3^ |
| **CST AD** | 0.74 | 1139 | -0.023 | 0.059 | -0.14 | 0.09 | 8.19 𝗑 10^-1^ |
| **SFO AD** | 0.86 | 1139 | 0.012 | 0.059 | -0.10 | 0.13 | 8.59 𝗑 10^-1^ |
| **SLF AD** | 0.16 | 1139 | -0.097 | 0.059 | -0.21 | 0.02 | 3.25 𝗑 10^-1^ |
| **SS AD** | 0.00 | 1139 | -0.233 | 0.059 | -0.35 | -0.12 | 6.29 𝗑 10^-3^ |
| **TAP AD** | 0.84 | 1139 | 0.014 | 0.059 | -0.10 | 0.13 | 8.59 𝗑 10^-1^ |
| **UNC AD** | 0.28 | 1139 | -0.075 | 0.059 | -0.19 | 0.04 | 4.35 𝗑 10^-1^ |

| **HY2 & Controls** | ***p*-value** | **Degrees of Freedom** | **Partial Cohen’s *d*** | **Standard Error** | **Lower CI** | **Upper CI** | ***p*_FDR_** |
| --- | --- | --- | --- | --- | --- | --- | --- |
| **Entire WM AD** | 0.11 | 1606 | -0.079 | 0.050 | -0.18 | 0.02 | 2.77 𝗑 10^-1^ |
| **ACR AD** | 0.50 | 1606 | -0.034 | 0.050 | -0.13 | 0.06 | 5.74 𝗑 10^-1^ |
| **SCR AD** | 0.44 | 1606 | 0.039 | 0.050 | -0.06 | 0.14 | 5.40 𝗑 10^-1^ |
| **PCR AD** | 0.88 | 1606 | -0.007 | 0.050 | -0.10 | 0.09 | 8.84 𝗑 10^-1^ |
| **ALIC AD** | 0.00 | 1606 | -0.162 | 0.050 | -0.26 | -0.06 | 6.85 𝗑 10^-3^ |
| **PLIC AD** | 0.08 | 1606 | -0.087 | 0.050 | -0.18 | 0.01 | 2.29 𝗑 10^-1^ |
| **RLIC AD** | 0.00 | 1606 | -0.235 | 0.050 | -0.33 | -0.14 | 2.16 𝗑 10^-5^ |
| **EC AD** | 0.37 | 1606 | -0.045 | 0.050 | -0.14 | 0.05 | 5.40 𝗑 10^-1^ |
| **FX AD** | 0.17 | 1606 | 0.069 | 0.050 | -0.03 | 0.17 | 3.69 𝗑 10^-1^ |
| **FXST AD** | 0.00 | 1606 | -0.274 | 0.050 | -0.37 | -0.18 | 5.85 𝗑 10^-7^ |
| **PTR AD** | 0.02 | 1606 | -0.119 | 0.050 | -0.22 | -0.02 | 5.47 𝗑 10^-2^ |
| **GCC AD** | 0.63 | 1606 | -0.024 | 0.050 | -0.12 | 0.07 | 6.88 𝗑 10^-1^ |
| **BCC AD** | 0.79 | 1606 | 0.014 | 0.050 | -0.08 | 0.11 | 8.24 𝗑 10^-1^ |
| **SCC AD** | 0.44 | 1606 | 0.039 | 0.050 | -0.06 | 0.14 | 5.40 𝗑 10^-1^ |
| **CGC AD** | 0.01 | 1606 | -0.133 | 0.050 | -0.23 | -0.04 | 2.86 𝗑 10^-2^ |
| **CGH AD** | 0.00 | 1606 | -0.279 | 0.050 | -0.38 | -0.18 | 5.85 𝗑 10^-7^ |
| **CST AD** | 0.24 | 1606 | -0.059 | 0.050 | -0.16 | 0.04 | 3.77 𝗑 10^-1^ |
| **SFO AD** | 0.21 | 1606 | -0.063 | 0.050 | -0.16 | 0.03 | 3.69 𝗑 10^-1^ |
| **SLF AD** | 0.22 | 1606 | -0.062 | 0.050 | -0.16 | 0.04 | 3.69 𝗑 10^-1^ |
| **SS AD** | 0.00 | 1606 | -0.157 | 0.050 | -0.25 | -0.06 | 7.87 𝗑 10^-3^ |
| **TAP AD** | 0.40 | 1606 | 0.042 | 0.050 | -0.06 | 0.14 | 5.40 𝗑 10^-1^ |
| **UNC AD** | 0.21 | 1606 | -0.062 | 0.050 | -0.16 | 0.04 | 3.69 𝗑 10^-1^ |

| **HY3 & Controls** | ***p*-value** | **Degrees of Freedom** | **Partial Cohen’s *d*** | **Standard Error** | **Lower CI** | **Upper CI** | ***p*_FDR_** |
| --- | --- | --- | --- | --- | --- | --- | --- |
| **Entire WM AD** | 0.18 | 1085 | -0.103 | 0.060 | -0.22 | 0.02 | 3.66 𝗑 10^-1^ |
| **ACR AD** | 0.68 | 1085 | 0.032 | 0.060 | -0.09 | 0.15 | 6.75 𝗑 10^-1^ |
| **SCR AD** | 0.48 | 1085 | 0.053 | 0.060 | -0.06 | 0.17 | 6.75 𝗑 10^-1^ |
| **PCR AD** | 0.65 | 1085 | 0.035 | 0.060 | -0.08 | 0.15 | 6.75 𝗑 10^-1^ |
| **ALIC AD** | 0.02 | 1085 | -0.181 | 0.060 | -0.30 | -0.06 | 1.28 𝗑 10^-1^ |
| **PLIC AD** | 0.37 | 1085 | -0.068 | 0.060 | -0.19 | 0.05 | 6.42 𝗑 10^-1^ |
| **RLIC AD** | 0.03 | 1085 | -0.168 | 0.060 | -0.29 | -0.05 | 1.48 𝗑 10^-1^ |
| **EC AD** | 0.67 | 1085 | -0.033 | 0.060 | -0.15 | 0.09 | 6.75 𝗑 10^-1^ |
| **FX AD** | 0.16 | 1085 | 0.108 | 0.060 | -0.01 | 0.23 | 3.66 𝗑 10^-1^ |
| **FXST AD** | 0.01 | 1085 | -0.189 | 0.060 | -0.31 | -0.07 | 1.28 𝗑 10^-1^ |
| **PTR AD** | 0.04 | 1085 | -0.155 | 0.060 | -0.27 | -0.04 | 1.83 𝗑 10^-1^ |
| **GCC AD** | 0.42 | 1085 | -0.061 | 0.060 | -0.18 | 0.06 | 6.61 𝗑 10^-1^ |
| **BCC AD** | 0.56 | 1085 | 0.045 | 0.060 | -0.07 | 0.16 | 6.75 𝗑 10^-1^ |
| **SCC AD** | 0.55 | 1085 | -0.045 | 0.060 | -0.16 | 0.07 | 6.75 𝗑 10^-1^ |
| **CGC AD** | 0.05 | 1085 | -0.147 | 0.060 | -0.27 | -0.03 | 1.83 𝗑 10^-1^ |
| **CGH AD** | 0.00 | 1085 | -0.277 | 0.061 | -0.40 | -0.16 | 6.24 𝗑 10^-3^ |
| **CST AD** | 0.64 | 1085 | 0.036 | 0.060 | -0.08 | 0.15 | 6.75 𝗑 10^-1^ |
| **SFO AD** | 0.18 | 1085 | 0.101 | 0.060 | -0.02 | 0.22 | 3.66 𝗑 10^-1^ |
| **SLF AD** | 0.38 | 1085 | -0.067 | 0.060 | -0.18 | 0.05 | 6.42 𝗑 10^-1^ |
| **SS AD** | 0.13 | 1085 | -0.114 | 0.060 | -0.23 | 0.00 | 3.66 𝗑 10^-1^ |
| **TAP AD** | 0.06 | 1085 | 0.144 | 0.060 | 0.03 | 0.26 | 1.83 𝗑 10^-1^ |
| **UNC AD** | 0.61 | 1085 | -0.038 | 0.060 | -0.16 | 0.08 | 6.75 𝗑 10^-1^ |

| **HY4/5 & Controls** | ***p*-value** | **Degrees of Freedom** | **Partial Cohen’s *d*** | **Standard Error** | **Lower CI** | **Upper CI** | ***p*_FDR_** |
| --- | --- | --- | --- | --- | --- | --- | --- |
| **Entire WM AD** | 0.00 | 940 | -0.354 | 0.065 | -0.48 | -0.23 | 6.69 𝗑 10^-3^ |
| **ACR AD** | 0.51 | 940 | 0.079 | 0.065 | -0.05 | 0.21 | 5.96 𝗑 10^-1^ |
| **SCR AD** | 0.17 | 940 | 0.168 | 0.065 | 0.04 | 0.30 | 2.44 𝗑 10^-1^ |
| **PCR AD** | 0.89 | 940 | -0.016 | 0.065 | -0.14 | 0.11 | 8.93 𝗑 10^-1^ |
| **ALIC AD** | 0.02 | 940 | -0.275 | 0.065 | -0.40 | -0.15 | 3.77 𝗑 10^-2^ |
| **PLIC AD** | 0.00 | 940 | -0.387 | 0.065 | -0.51 | -0.26 | 3.31 𝗑 10^-3^ |
| **RLIC AD** | 0.00 | 940 | -0.409 | 0.065 | -0.54 | -0.28 | 2.17 𝗑 10^-3^ |
| **EC AD** | 0.30 | 940 | -0.125 | 0.065 | -0.25 | 0.00 | 3.91 𝗑 10^-1^ |
| **FX AD** | 0.00 | 940 | 0.431 | 0.065 | 0.30 | 0.56 | 1.28 𝗑 10^-3^ |
| **FXST AD** | 0.00 | 940 | -0.440 | 0.065 | -0.57 | -0.31 | 1.15 𝗑 10^-3^ |
| **PTR AD** | 0.00 | 940 | -0.673 | 0.066 | -0.80 | -0.54 | 8.82 𝗑 10^-7^ |
| **GCC AD** | 0.77 | 940 | -0.035 | 0.065 | -0.16 | 0.09 | 8.10 𝗑 10^-1^ |
| **BCC AD** | 0.20 | 940 | -0.155 | 0.065 | -0.28 | -0.03 | 2.79 𝗑 10^-1^ |
| **SCC AD** | 0.01 | 940 | -0.321 | 0.065 | -0.45 | -0.19 | 1.41 𝗑 10^-2^ |
| **CGC AD** | 0.00 | 940 | -0.650 | 0.066 | -0.78 | -0.52 | 1.21 𝗑 10^-6^ |
| **CGH AD** | 0.00 | 940 | -0.617 | 0.066 | -0.75 | -0.49 | 3.45 𝗑 10^-6^ |
| **CST AD** | 0.00 | 940 | -0.402 | 0.065 | -0.53 | -0.27 | 2.36 𝗑 10^-3^ |
| **SFO AD** | 0.40 | 940 | 0.103 | 0.065 | -0.02 | 0.23 | 4.86 𝗑 10^-1^ |
| **SLF AD** | 0.00 | 940 | -0.370 | 0.065 | -0.50 | -0.24 | 4.84 𝗑 10^-3^ |
| **SS AD** | 0.00 | 940 | -0.596 | 0.066 | -0.73 | -0.47 | 6.07 𝗑 10^-6^ |
| **TAP AD** | 0.56 | 940 | -0.070 | 0.065 | -0.20 | 0.06 | 6.19 𝗑 10^-1^ |
| **UNC AD** | 0.00 | 940 | -0.458 | 0.065 | -0.59 | -0.33 | 7.76 𝗑 10^-4^ |

##### 2.1.6 AD: Stratification by HY Stage - Matched Control Samples

| **HY1 & Matched Controls** | ***p*-value** | **Degrees of Freedom** | **Partial Cohen’s *d*** | **Standard Error** | **Lower CI** | **Upper CI** | ***p*_FDR_** |
| --- | --- | --- | --- | --- | --- | --- | --- |
| **Entire WM AD** | 0.61 | 529 | -0.044 | 0.09 | -0.21 | 0.12 | 7.56 𝗑 10^-1^ |
| **ACR AD** | 0.89 | 529 | -0.012 | 0.09 | -0.18 | 0.16 | 8.92 𝗑 10^-1^ |
| **SCR AD** | 0.10 | 529 | 0.143 | 0.09 | -0.02 | 0.31 | 4.42 𝗑 10^-1^ |
| **PCR AD** | 0.78 | 529 | 0.024 | 0.09 | -0.14 | 0.19 | 8.59 𝗑 10^-1^ |
| **ALIC AD** | 0.26 | 529 | -0.099 | 0.09 | -0.27 | 0.07 | 6.52 𝗑 10^-1^ |
| **PLIC AD** | 0.85 | 529 | 0.016 | 0.09 | -0.15 | 0.18 | 8.92 𝗑 10^-1^ |
| **RLIC AD** | 0.04 | 529 | -0.176 | 0.09 | -0.34 | -0.01 | 3.43 𝗑 10^-1^ |
| **EC AD** | 0.61 | 529 | -0.045 | 0.09 | -0.21 | 0.12 | 7.56 𝗑 10^-1^ |
| **FX AD** | 0.62 | 529 | -0.043 | 0.09 | -0.21 | 0.12 | 7.56 𝗑 10^-1^ |
| **FXST AD** | 0.02 | 529 | -0.208 | 0.09 | -0.38 | -0.04 | 3.43 𝗑 10^-1^ |
| **PTR AD** | 0.54 | 529 | -0.053 | 0.09 | -0.22 | 0.11 | 7.56 𝗑 10^-1^ |
| **GCC AD** | 0.33 | 529 | -0.084 | 0.09 | -0.25 | 0.08 | 7.36 𝗑 10^-1^ |
| **BCC AD** | 0.12 | 529 | 0.135 | 0.09 | -0.03 | 0.30 | 4.42 𝗑 10^-1^ |
| **SCC AD** | 0.15 | 529 | 0.124 | 0.09 | -0.04 | 0.29 | 4.85 𝗑 10^-1^ |
| **CGC AD** | 0.27 | 529 | -0.097 | 0.09 | -0.26 | 0.07 | 6.52 𝗑 10^-1^ |
| **CGH AD** | 0.05 | 529 | -0.169 | 0.09 | -0.34 | 0.00 | 3.43 𝗑 10^-1^ |
| **CST AD** | 0.62 | 529 | -0.044 | 0.09 | -0.21 | 0.12 | 7.56 𝗑 10^-1^ |
| **SFO AD** | 0.44 | 529 | 0.067 | 0.09 | -0.10 | 0.23 | 7.56 𝗑 10^-1^ |
| **SLF AD** | 0.58 | 529 | -0.048 | 0.09 | -0.22 | 0.12 | 7.56 𝗑 10^-1^ |
| **SS AD** | 0.06 | 529 | -0.162 | 0.09 | -0.33 | 0.01 | 3.43 𝗑 10^-1^ |
| **TAP AD** | 0.58 | 529 | 0.048 | 0.09 | -0.12 | 0.22 | 7.56 𝗑 10^-1^ |
| **UNC AD** | 0.72 | 529 | -0.031 | 0.09 | -0.20 | 0.14 | 8.39 𝗑 10^-1^ |

| **HY2 & Matched Controls** | ***p*-value** | **Degrees of Freedom** | **Partial Cohen’s *d*** | **Standard Error** | **Lower CI** | **Upper CI** | ***p*_FDR_** |
| --- | --- | --- | --- | --- | --- | --- | --- |
| **Entire WM AD** | 0.11 | 1463 | -0.083 | 0.05 | -0.18 | 0.02 | 2.80 𝗑 10^-1^ |
| **ACR AD** | 0.55 | 1463 | -0.031 | 0.05 | -0.13 | 0.07 | 6.34 𝗑 10^-1^ |
| **SCR AD** | 0.42 | 1463 | 0.042 | 0.05 | -0.06 | 0.14 | 5.10 𝗑 10^-1^ |
| **PCR AD** | 0.98 | 1463 | 0.001 | 0.05 | -0.10 | 0.10 | 9.83 𝗑 10^-1^ |
| **ALIC AD** | 0.00 | 1463 | -0.179 | 0.05 | -0.28 | -0.08 | 3.48 𝗑 10^-3^ |
| **PLIC AD** | 0.06 | 1463 | -0.099 | 0.05 | -0.20 | 0.00 | 1.61 𝗑 10^-1^ |
| **RLIC AD** | 0.00 | 1463 | -0.240 | 0.05 | -0.34 | -0.14 | 3.67 𝗑 10^-5^ |
| **EC AD** | 0.30 | 1463 | -0.054 | 0.05 | -0.16 | 0.05 | 4.22 𝗑 10^-1^ |
| **FX AD** | 0.21 | 1463 | 0.066 | 0.05 | -0.04 | 0.17 | 3.90 𝗑 10^-1^ |
| **FXST AD** | 0.00 | 1463 | -0.281 | 0.05 | -0.38 | -0.18 | 1.01 𝗑 10^-6^ |
| **PTR AD** | 0.03 | 1463 | -0.113 | 0.05 | -0.21 | -0.01 | 9.65 𝗑 10^-2^ |
| **GCC AD** | 0.68 | 1463 | -0.021 | 0.05 | -0.12 | 0.08 | 7.53 𝗑 10^-1^ |
| **BCC AD** | 0.76 | 1463 | 0.016 | 0.05 | -0.09 | 0.12 | 8.01 𝗑 10^-1^ |
| **SCC AD** | 0.34 | 1463 | 0.050 | 0.05 | -0.05 | 0.15 | 4.42 𝗑 10^-1^ |
| **CGC AD** | 0.01 | 1463 | -0.130 | 0.05 | -0.23 | -0.03 | 4.89 𝗑 10^-2^ |
| **CGH AD** | 0.00 | 1463 | -0.287 | 0.05 | -0.39 | -0.18 | 1.01 𝗑 10^-6^ |
| **CST AD** | 0.20 | 1463 | -0.067 | 0.05 | -0.17 | 0.04 | 3.90 𝗑 10^-1^ |
| **SFO AD** | 0.31 | 1463 | -0.053 | 0.05 | -0.16 | 0.05 | 4.22 𝗑 10^-1^ |
| **SLF AD** | 0.29 | 1463 | -0.056 | 0.05 | -0.16 | 0.05 | 4.22 𝗑 10^-1^ |
| **SS AD** | 0.00 | 1463 | -0.159 | 0.05 | -0.26 | -0.06 | 1.07 𝗑 10^-2^ |
| **TAP AD** | 0.26 | 1463 | 0.059 | 0.05 | -0.04 | 0.16 | 4.22 𝗑 10^-1^ |
| **UNC AD** | 0.21 | 1463 | -0.065 | 0.05 | -0.17 | 0.04 | 3.90 𝗑 10^-1^ |

| **HY3 & Matched Controls** | ***p*-value** | **Degrees of Freedom** | **Partial Cohen’s *d*** | **Standard Error** | **Lower CI** | **Upper CI** | ***p*_FDR_** |
| --- | --- | --- | --- | --- | --- | --- | --- |
| **Entire WM AD** | 0.09 | 420 | -0.166 | 0.10 | -0.35 | 0.02 | 2.49 𝗑 10^-1^ |
| **ACR AD** | 0.99 | 420 | -0.001 | 0.10 | -0.19 | 0.19 | 9.91 𝗑 10^-1^ |
| **SCR AD** | 0.97 | 420 | -0.004 | 0.10 | -0.19 | 0.18 | 9.91 𝗑 10^-1^ |
| **PCR AD** | 0.79 | 420 | -0.027 | 0.10 | -0.21 | 0.16 | 9.09 𝗑 10^-1^ |
| **ALIC AD** | 0.05 | 420 | -0.192 | 0.10 | -0.38 | 0.00 | 1.57 𝗑 10^-1^ |
| **PLIC AD** | 0.60 | 420 | -0.052 | 0.10 | -0.24 | 0.14 | 7.66 𝗑 10^-1^ |
| **RLIC AD** | 0.02 | 420 | -0.221 | 0.10 | -0.41 | -0.03 | 1.07 𝗑 10^-1^ |
| **EC AD** | 0.18 | 420 | -0.132 | 0.10 | -0.32 | 0.06 | 3.87 𝗑 10^-1^ |
| **FX AD** | 0.32 | 420 | 0.098 | 0.10 | -0.09 | 0.28 | 6.10 𝗑 10^-1^ |
| **FXST AD** | 0.02 | 420 | -0.233 | 0.10 | -0.42 | -0.04 | 1.00 𝗑 10^-1^ |
| **PTR AD** | 0.01 | 420 | -0.245 | 0.10 | -0.43 | -0.06 | 1.00 𝗑 10^-1^ |
| **GCC AD** | 0.39 | 420 | -0.083 | 0.10 | -0.27 | 0.10 | 6.67 𝗑 10^-1^ |
| **BCC AD** | 0.92 | 420 | 0.010 | 0.10 | -0.18 | 0.20 | 9.91 𝗑 10^-1^ |
| **SCC AD** | 0.63 | 420 | -0.048 | 0.10 | -0.23 | 0.14 | 7.66 𝗑 10^-1^ |
| **CGC AD** | 0.03 | 420 | -0.209 | 0.10 | -0.40 | -0.02 | 1.20 𝗑 10^-1^ |
| **CGH AD** | 0.00 | 420 | -0.276 | 0.10 | -0.46 | -0.09 | 1.00 𝗑 10^-1^ |
| **CST AD** | 0.49 | 420 | 0.068 | 0.10 | -0.12 | 0.26 | 6.69 𝗑 10^-1^ |
| **SFO AD** | 0.33 | 420 | 0.095 | 0.10 | -0.09 | 0.28 | 6.10 𝗑 10^-1^ |
| **SLF AD** | 0.12 | 420 | -0.151 | 0.10 | -0.34 | 0.04 | 3.01 𝗑 10^-1^ |
| **SS AD** | 0.02 | 420 | -0.231 | 0.10 | -0.42 | -0.04 | 1.00 𝗑 10^-1^ |
| **TAP AD** | 0.48 | 420 | 0.069 | 0.10 | -0.12 | 0.26 | 6.69 𝗑 10^-1^ |
| **UNC AD** | 0.46 | 420 | -0.072 | 0.10 | -0.26 | 0.12 | 6.69 𝗑 10^-1^ |

| **HY4/5 & Matched Controls** | ***p*-value** | **Degrees of Freedom** | **Partial Cohen’s *d*** | **Standard Error** | **Lower CI** | **Upper CI** | ***p*_FDR_** |
| --- | --- | --- | --- | --- | --- | --- | --- |
| **Entire WM AD** | 0.09 | 131 | -0.297 | 0.17 | -0.62 | 0.03 | 1.89 𝗑 10^-1^ |
| **ACR AD** | 0.89 | 131 | 0.024 | 0.16 | -0.30 | 0.35 | 9.69 𝗑 10^-1^ |
| **SCR AD** | 0.42 | 131 | 0.142 | 0.16 | -0.18 | 0.46 | 5.41 𝗑 10^-1^ |
| **PCR AD** | 0.93 | 131 | -0.016 | 0.16 | -0.34 | 0.31 | 9.69 𝗑 10^-1^ |
| **ALIC AD** | 0.41 | 131 | -0.146 | 0.16 | -0.47 | 0.18 | 5.41 𝗑 10^-1^ |
| **PLIC AD** | 0.38 | 131 | -0.155 | 0.16 | -0.48 | 0.17 | 5.41 𝗑 10^-1^ |
| **RLIC AD** | 0.05 | 131 | -0.340 | 0.17 | -0.66 | -0.02 | 1.64 𝗑 10^-1^ |
| **EC AD** | 0.31 | 131 | -0.178 | 0.16 | -0.50 | 0.14 | 4.88 𝗑 10^-1^ |
| **FX AD** | 0.02 | 131 | 0.397 | 0.17 | 0.07 | 0.72 | 1.09 𝗑 10^-1^ |
| **FXST AD** | 0.07 | 131 | -0.323 | 0.17 | -0.65 | 0.00 | 1.64 𝗑 10^-1^ |
| **PTR AD** | 0.02 | 131 | -0.411 | 0.17 | -0.74 | -0.09 | 1.09 𝗑 10^-1^ |
| **GCC AD** | 0.78 | 131 | -0.049 | 0.16 | -0.37 | 0.27 | 9.50 𝗑 10^-1^ |
| **BCC AD** | 0.24 | 131 | -0.204 | 0.16 | -0.53 | 0.12 | 4.14 𝗑 10^-1^ |
| **SCC AD** | 0.10 | 131 | -0.289 | 0.17 | -0.61 | 0.03 | 1.89 𝗑 10^-1^ |
| **CGC AD** | 0.01 | 131 | -0.477 | 0.17 | -0.80 | -0.15 | 5.53 𝗑 10^-2^ |
| **CGH AD** | 0.00 | 131 | -0.613 | 0.17 | -0.94 | -0.28 | 1.37 𝗑 10^-2^ |
| **CST AD** | 0.04 | 131 | -0.370 | 0.17 | -0.70 | -0.05 | 1.32 𝗑 10^-1^ |
| **SFO AD** | 0.99 | 131 | 0.002 | 0.16 | -0.32 | 0.32 | 9.89 𝗑 10^-1^ |
| **SLF AD** | 0.10 | 131 | -0.287 | 0.17 | -0.61 | 0.04 | 1.89 𝗑 10^-1^ |
| **SS AD** | 0.01 | 131 | -0.474 | 0.17 | -0.80 | -0.15 | 5.53 𝗑 10^-2^ |
| **TAP AD** | 0.88 | 131 | -0.027 | 0.16 | -0.35 | 0.30 | 9.69 𝗑 10^-1^ |
| **UNC AD** | 0.06 | 131 | -0.328 | 0.17 | -0.65 | 0.00 | 1.64 𝗑 10^-1^ |

##### 2.1.7 RD: Stratification by HY Stage

| **HY1 & Controls** | ***p*-value** | **Degrees of Freedom** | **Partial Cohen’s *d*** | **Standard Error** | **Lower CI** | **Upper CI** | ***p*_FDR_** |
| --- | --- | --- | --- | --- | --- | --- | --- |
| **Entire WM RD** | 0.00 | 1139 | -0.218 | 0.059 | -0.33 | -0.10 | 1.38 𝗑 10^-1^ |
| **ACR RD** | 0.03 | 1139 | -0.151 | 0.059 | -0.27 | -0.04 | 8.44 𝗑 10^-2^ |
| **SCR RD** | 0.24 | 1139 | -0.082 | 0.059 | -0.20 | 0.03 | 3.22 𝗑 10^-1^ |
| **PCR RD** | 0.22 | 1139 | -0.086 | 0.059 | -0.20 | 0.03 | 3.22 𝗑 10^-1^ |
| **ALIC RD** | 0.03 | 1139 | -0.151 | 0.059 | -0.27 | -0.04 | 8.44 𝗑 10^-2^ |
| **PLIC RD** | 0.02 | 1139 | -0.159 | 0.059 | -0.27 | -0.04 | 8.40 𝗑 10^-2^ |
| **RLIC RD** | 0.00 | 1139 | -0.217 | 0.059 | -0.33 | -0.10 | 1.38 𝗑 10^-2^ |
| **EC RD** | 0.06 | 1139 | -0.134 | 0.059 | -0.25 | -0.02 | 1.11 𝗑 10^-1^ |
| **FX RD** | 0.90 | 1139 | -0.009 | 0.059 | -0.12 | 0.11 | 9.00 𝗑 10^-1^ |
| **FXST RD** | 0.04 | 1139 | -0.143 | 0.059 | -0.26 | -0.03 | 9.71 𝗑 10^-2^ |
| **PTR RD** | 0.08 | 1139 | -0.120 | 0.059 | -0.24 | 0.00 | 1.56 𝗑 10^-1^ |
| **GCC RD** | 0.02 | 1139 | -0.159 | 0.059 | -0.27 | -0.04 | 8.40 𝗑 10^-2^ |
| **BCC RD** | 0.53 | 1139 | -0.044 | 0.059 | -0.16 | 0.07 | 5.54 𝗑 10^-1^ |
| **SCC RD** | 0.26 | 1139 | -0.079 | 0.059 | -0.19 | 0.04 | 3.22 𝗑 10^-1^ |
| **CGC RD** | 0.26 | 1139 | -0.078 | 0.059 | -0.19 | 0.04 | 3.22 𝗑 10^-1^ |
| **CGH RD** | 0.04 | 1139 | -0.140 | 0.059 | -0.26 | -0.02 | 9.79 𝗑 10^-1^ |
| **CST RD** | 0.00 | 1139 | -0.198 | 0.059 | -0.31 | -0.08 | 2.54 𝗑 10^-2^ |
| **SFO RD** | 0.35 | 1139 | -0.065 | 0.059 | -0.18 | 0.05 | 3.88 𝗑 10^-1^ |
| **SLF RD** | 0.16 | 1139 | -0.099 | 0.059 | -0.21 | 0.02 | 2.44 𝗑 10^-1^ |
| **SS RD** | 0.00 | 1139 | -0.218 | 0.059 | -0.33 | -0.10 | 1.38 𝗑 10^-2^ |
| **TAP RD** | 0.29 | 1139 | -0.073 | 0.059 | -0.19 | 0.04 | 3.38 𝗑 10^-1^ |
| **UNC RD** | 0.10 | 1139 | -0.114 | 0.059 | -0.23 | 0.00 | 1.71 𝗑 10^-1^ |

| **HY2 & Controls** | ***p*-value** | **Degrees of Freedom** | **Partial Cohen’s *d*** | **Standard Error** | **Lower CI** | **Upper CI** | ***p*_FDR_** |
| --- | --- | --- | --- | --- | --- | --- | --- |
| **Entire WM RD** | 0.81 | 1606 | 0.012 | 0.050 | -0.09 | 0.11 | 9.67 𝗑 10^-1^ |
| **ACR RD** | 0.55 | 1606 | 0.030 | 0.050 | -0.07 | 0.13 | 9.67 𝗑 10^-1^ |
| **SCR RD** | 0.37 | 1606 | -0.044 | 0.050 | -0.14 | 0.05 | 7.82 𝗑 10^-1^ |
| **PCR RD** | 0.74 | 1606 | 0.017 | 0.050 | -0.08 | 0.11 | 9.67 𝗑 10^-1^ |
| **ALIC RD** | 0.29 | 1606 | -0.053 | 0.050 | -0.15 | 0.04 | 7.82 𝗑 10^-1^ |
| **PLIC RD** | 0.03 | 1606 | -0.110 | 0.050 | -0.21 | -0.01 | 2.57 𝗑 10^-1^ |
| **RLIC RD** | 0.04 | 1606 | -0.106 | 0.050 | -0.20 | -0.01 | 2.57 𝗑 10^-1^ |
| **EC RD** | 0.74 | 1606 | -0.016 | 0.050 | -0.11 | 0.08 | 9.67 𝗑 10^-1^ |
| **FX RD** | 0.00 | 1606 | 0.188 | 0.050 | 0.09 | 0.29 | 4.09 𝗑 10^-3^ |
| **FXST RD** | 0.13 | 1606 | -0.076 | 0.050 | -0.17 | 0.02 | 7.25 𝗑 10^-1^ |
| **PTR RD** | 0.93 | 1606 | -0.005 | 0.050 | -0.10 | 0.09 | 9.67 𝗑 10^-1^ |
| **GCC RD** | 0.84 | 1606 | 0.010 | 0.050 | -0.09 | 0.11 | 9.67 𝗑 10^-1^ |
| **BCC RD** | 0.62 | 1606 | 0.025 | 0.050 | -0.07 | 0.12 | 9.67 𝗑 10^-1^ |
| **SCC RD** | 0.88 | 1606 | -0.008 | 0.050 | -0.10 | 0.09 | 9.67 𝗑 10^-1^ |
| **CGC RD** | 0.97 | 1606 | 0.002 | 0.050 | -0.10 | 0.10 | 9.67 𝗑 10^-1^ |
| **CGH RD** | 0.27 | 1606 | -0.055 | 0.050 | -0.15 | 0.04 | 7.82 𝗑 10^-1^ |
| **CST RD** | 0.35 | 1606 | -0.047 | 0.050 | -0.14 | 0.05 | 7.82 𝗑 10^-1^ |
| **SFO RD** | 0.39 | 1606 | -0.043 | 0.050 | -0.14 | 0.05 | 7.82 𝗑 10^-1^ |
| **SLF RD** | 0.87 | 1606 | -0.008 | 0.050 | -0.11 | 0.09 | 9.67 𝗑 10^-1^ |
| **SS RD** | 0.37 | 1606 | -0.045 | 0.050 | -0.14 | 0.05 | 7.82 𝗑 10^-1^ |
| **TAP RD** | 0.63 | 1606 | 0.024 | 0.050 | -0.07 | 0.12 | 9.67 𝗑 10^-1^ |
| **UNC RD** | 0.20 | 1606 | 0.064 | 0.050 | -0.03 | 0.16 | 7.82 𝗑 10^-1^ |

| **HY3 & Controls** | ***p*-value** | **Degrees of Freedom** | **Partial Cohen’s *d*** | **Standard Error** | **Lower CI** | **Upper CI** | ***p*_FDR_** |
| --- | --- | --- | --- | --- | --- | --- | --- |
| **Entire WM RD** | 0.01 | 1085 | 0.195 | 0.060 | 0.08 | 0.31 | 4.64 𝗑 10^-2^ |
| **ACR RD** | 0.00 | 1085 | 0.289 | 0.061 | 0.17 | 0.41 | 3.33 𝗑 10^-3^ |
| **SCR RD** | 0.02 | 1085 | 0.174 | 0.060 | 0.06 | 0.29 | 7.07 𝗑 10^-2^ |
| **PCR RD** | 0.01 | 1085 | 0.200 | 0.060 | 0.08 | 0.32 | 4.64 𝗑 10-^2^ |
| **ALIC RD** | 0.38 | 1085 | 0.066 | 0.060 | -0.05 | 0.18 | 4.43 𝗑 10^-1^ |
| **PLIC RD** | 0.29 | 1085 | -0.080 | 0.060 | -0.20 | 0.04 | 3.57 𝗑 10^-1^ |
| **RLIC RD** | 0.83 | 1085 | 0.016 | 0.060 | -0.10 | 0.13 | 8.68 𝗑 10^-1^ |
| **EC RD** | 0.01 | 1085 | 0.186 | 0.060 | 0.07 | 0.30 | 5.29 𝗑 10^-2^ |
| **FX RD** | 0.00 | 1085 | 0.271 | 0.060 | 0.15 | 0.39 | 4.16 𝗑 10^-3^ |
| **FXST RD** | 0.05 | 1085 | 0.150 | 0.060 | 0.03 | 0.27 | 9.17 𝗑 10^-2^ |
| **PTR RD** | 0.09 | 1085 | 0.130 | 0.060 | 0.01 | 0.25 | 1.38 𝗑 10^-1^ |
| **GCC RD** | 0.05 | 1085 | 0.150 | 0.060 | 0.03 | 0.27 | 9.17 𝗑 10^-2^ |
| **BCC RD** | 0.15 | 1085 | 0.110 | 0.060 | -0.01 | 0.23 | 2.18 𝗑 10^-1^ |
| **SCC RD** | 0.23 | 1085 | 0.091 | 0.060 | -0.03 | 0.21 | 3.17 𝗑 10^-1^ |
| **CGC RD** | 0.25 | 1085 | 0.087 | 0.060 | -0.03 | 0.20 | 3.29 𝗑 10^-1^ |
| **CGH RD** | 0.69 | 1085 | -0.030 | 0.060 | -0.15 | 0.09 | 7.63 𝗑 10^-1^ |
| **CST RD** | 0.88 | 1085 | -0.012 | 0.060 | -0.13 | 0.11 | 8.76 𝗑 10^-1^ |
| **SFO RD** | 0.03 | 1085 | 0.169 | 0.060 | 0.05 | 0.29 | 7.28 𝗑 10^--2^ |
| **SLF RD** | 0.01 | 1085 | 0.195 | 0.060 | 0.08 | 0.31 | 4.64 𝗑 10^-2^ |
| **SS RD** | 0.04 | 1085 | 0.160 | 0.060 | 0.04 | 0.28 | 8.63 𝗑 10^-2^ |
| **TAP RD** | 0.08 | 1085 | 0.132 | 0.060 | 0.01 | 0.25 | 1.38 𝗑 10^-1^ |
| **UNC RD** | 0.05 | 1085 | 0.149 | 0.060 | 0.03 | 0.27 | 9.17 𝗑 10^-2^ |

| **HY4/5 & Controls** | ***p*-value** | **Degrees of Freedom** | **Partial Cohen’s *d*** | **Standard Error** | **Lower CI** | **Upper CI** | ***p*_FDR_** |
| --- | --- | --- | --- | --- | --- | --- | --- |
| **Entire WM RD** | 0.00 | 940 | 0.576 | 0.066 | 0.45 | 0.70 | 9.25 𝗑 10^-6^ |
| **ACR RD** | 0.00 | 940 | 0.545 | 0.066 | 0.42 | 0.67 | 2.62 𝗑 10^-5^ |
| **SCR RD** | 0.00 | 940 | 0.488 | 0.066 | 0.36 | 0.62 | 1.75 𝗑 10^-4^ |
| **PCR RD** | 0.00 | 940 | 0.356 | 0.065 | 0.23 | 0.48 | 5.08 𝗑 10^-^3 |
| **ALIC RD** | 0.00 | 940 | 0.397 | 0.065 | 0.27 | 0.52 | 2.00 𝗑 10^-3^ |
| **PLIC RD** | 0.77 | 940 | 0.036 | 0.065 | -0.09 | 0.16 | 8.34 𝗑 10^-1^ |
| **RLIC RD** | 0.88 | 940 | 0.018 | 0.065 | -0.11 | 0.14 | 8.83 𝗑 10^-1^ |
| **EC RD** | 0.00 | 940 | 0.587 | 0.066 | 0.46 | 0.72 | 8.78 𝗑 10^-6^ |
| **FX RD** | 0.00 | 940 | 0.778 | 0.067 | 0.65 | 0.91 | 5.36 𝗑 10^-9^ |
| **FXST RD** | 0.00 | 940 | 0.441 | 0.065 | 0.31 | 0.57 | 6.69 𝗑 10^-4^ |
| **PTR RD** | 0.01 | 940 | 0.318 | 0.065 | 0.19 | 0.45 | 1.16 𝗑 10^-2^ |
| **GCC RD** | 0.00 | 940 | 0.638 | 0.066 | 0.51 | 0.77 | 1.39 𝗑 10^-6^ |
| **BCC RD** | 0.00 | 940 | 0.577 | 0.066 | 0.45 | 0.71 | 9.25 𝗑 10^-6^ |
| **SCC RD** | 0.00 | 940 | 0.395 | 0.065 | 0.27 | 0.52 | 2.00 𝗑 10^-3^ |
| **CGC RD** | 0.00 | 940 | 0.437 | 0.065 | 0.31 | 0.57 | 6.76 𝗑 10^-4^ |
| **CGH RD** | 0.80 | 940 | 0.031 | 0.065 | -0.10 | 0.16 | 8.34 𝗑 10^-1^ |
| **CST RD** | 0.74 | 940 | 0.040 | 0.065 | -0.09 | 0.17 | 8.34 𝗑 10^-1^ |
| **SFO RD** | 0.00 | 940 | 0.651 | 0.066 | 0.52 | 0.78 | 1.20 𝗑 10^-6^ |
| **SLF RD** | 0.00 | 940 | 0.378 | 0.065 | 0.25 | 0.51 | 3.03 𝗑 10^-3^ |
| **SS RD** | 0.17 | 940 | 0.167 | 0.065 | 0.04 | 0.29 | 2.07 𝗑 10^-1^ |
| **TAP RD** | 0.01 | 940 | 0.324 | 0.065 | 0.20 | 0.45 | 1.06 𝗑 10^-2^ |
| **UNC RD** | 0.00 | 940 | 0.462 | 0.065 | 0.33 | 0.59 | 3.71 𝗑 10^-4^ |

##### 2.1.8 RD: Stratification by HY Stage - Matched Control Samples

| **HY1 & Matched Controls** | ***p*-value** | **Degrees of Freedom** | **Partial Cohen’s *d*** | **Standard Error** | **Lower CI** | **Upper CI** | ***p*_FDR_** |
| --- | --- | --- | --- | --- | --- | --- | --- |
| **Entire WM RD** | 0.08 | 529 | -0.150 | 0.09 | -0.32 | 0.02 | 3.71 𝗑 10^-1^ |
| **ACR RD** | 0.31 | 529 | -0.088 | 0.09 | -0.26 | 0.08 | 6.20 𝗑 10^-1^ |
| **SCR RD** | 0.18 | 529 | -0.117 | 0.09 | -0.28 | 0.05 | 4.96 𝗑 10^-1^ |
| **PCR RD** | 0.40 | 529 | -0.074 | 0.09 | -0.24 | 0.09 | 6.20 𝗑 10^-1^ |
| **ALIC RD** | 0.06 | 529 | -0.167 | 0.09 | -0.33 | 0.00 | 3.04 𝗑 10^-1^ |
| **PLIC RD** | 0.03 | 529 | -0.186 | 0.09 | -0.35 | -0.02 | 2.41 𝗑 10^-1^ |
| **RLIC RD** | 0.03 | 529 | -0.189 | 0.09 | -0.36 | -0.02 | 2.41 𝗑 10^-1^ |
| **EC RD** | 0.17 | 529 | -0.120 | 0.09 | -0.29 | 0.05 | 4.96 𝗑 10^-1^ |
| **FX RD** | 0.34 | 529 | 0.083 | 0.09 | -0.08 | 0.25 | 6.20 𝗑 10^-1^ |
| **FXST RD** | 0.49 | 529 | -0.060 | 0.09 | -0.23 | 0.11 | 6.37 𝗑 10^-1^ |
| **PTR RD** | 0.41 | 529 | -0.072 | 0.09 | -0.24 | 0.10 | 6.20 𝗑 10^-1^ |
| **GCC RD** | 0.23 | 529 | -0.105 | 0.09 | -0.27 | 0.06 | 5.56 𝗑 10^-1^ |
| **BCC RD** | 0.88 | 529 | -0.013 | 0.09 | -0.18 | 0.15 | 9.24 𝗑 10^-1^ |
| **SCC RD** | 0.93 | 529 | -0.008 | 0.09 | -0.18 | 0.16 | 9.31 𝗑 10^-1^ |
| **CGC RD** | 0.83 | 529 | -0.019 | 0.09 | -0.19 | 0.15 | 9.10 𝗑 10^-1^ |
| **CGH RD** | 0.42 | 529 | -0.070 | 0.09 | -0.24 | 0.10 | 6.20 𝗑 10^-1^ |
| **CST RD** | 0.12 | 529 | -0.135 | 0.09 | -0.30 | 0.03 | 4.45 𝗑 10^-1^ |
| **SFO RD** | 0.76 | 529 | -0.027 | 0.09 | -0.19 | 0.14 | 8.76 𝗑 10^-1^ |
| **SLF RD** | 0.57 | 529 | -0.050 | 0.09 | -0.22 | 0.12 | 6.93 𝗑 10^-1^ |
| **SS RD** | 0.03 | 529 | -0.193 | 0.09 | -0.36 | -0.03 | 2.41 𝗑 10^-1^ |
| **TAP RD** | 0.38 | 529 | -0.076 | 0.09 | -0.24 | 0.09 | 6.20 𝗑 10^-1^ |
| **UNC RD** | 0.48 | 529 | -0.062 | 0.09 | -0.23 | 0.11 | 6.37 𝗑 10^-1^ |

| **HY2 & Matched Controls** | ***p*-value** | **Degrees of Freedom** | **Partial Cohen’s *d*** | **Standard Error** | **Lower CI** | **Upper CI** | ***p*_FDR_** |
| --- | --- | --- | --- | --- | --- | --- | --- |
| **Entire WM RD** | 0.69 | 1463 | 0.021 | 0.05 | -0.08 | 0.12 | 9.36 𝗑 10^-1^ |
| **ACR RD** | 0.50 | 1463 | 0.035 | 0.05 | -0.07 | 0.14 | 7.93 𝗑 10^-1^ |
| **SCR RD** | 0.41 | 1463 | -0.043 | 0.05 | -0.14 | 0.06 | 7.93 𝗑 10^-1^ |
| **PCR RD** | 0.57 | 1463 | 0.030 | 0.05 | -0.07 | 0.13 | 8.37 𝗑 10^-1^ |
| **ALIC RD** | 0.37 | 1463 | -0.047 | 0.05 | -0.15 | 0.05 | 7.93 𝗑 10^-1^ |
| **PLIC RD** | 0.02 | 1463 | -0.118 | 0.05 | -0.22 | -0.02 | 2.67 𝗑 10^-1^ |
| **RLIC RD** | 0.07 | 1463 | -0.094 | 0.05 | -0.20 | 0.01 | 5.41 𝗑 10^-1^ |
| **EC RD** | 0.73 | 1463 | -0.018 | 0.05 | -0.12 | 0.08 | 9.36 𝗑 10^-1^ |
| **FX RD** | 0.00 | 1463 | 0.192 | 0.05 | 0.09 | 0.29 | 5.49 𝗑 10^-3^ |
| **FXST RD** | 0.16 | 1463 | -0.073 | 0.05 | -0.17 | 0.03 | 7.93 𝗑 10^-1^ |
| **PTR RD** | 0.81 | 1463 | 0.013 | 0.05 | -0.09 | 0.11 | 9.36 𝗑 10^-1^ |
| **GCC RD** | 0.80 | 1463 | 0.013 | 0.05 | -0.09 | 0.12 | 9.36 𝗑 10^-1^ |
| **BCC RD** | 0.45 | 1463 | 0.039 | 0.05 | -0.06 | 0.14 | 7.93 𝗑 10^-1^ |
| **SCC RD** | 1.00 | 1463 | 0.000 | 0.05 | -0.10 | 0.10 | 9.98 𝗑 10^-1^ |
| **CGC RD** | 0.87 | 1463 | 0.009 | 0.05 | -0.09 | 0.11 | 9.53 𝗑 10^-1^ |
| **CGH RD** | 0.32 | 1463 | -0.052 | 0.05 | -0.15 | 0.05 | 7.93 𝗑 10^-1^ |
| **CST RD** | 0.44 | 1463 | -0.041 | 0.05 | -0.14 | 0.06 | 7.93 𝗑 10^-1^ |
| **SFO RD** | 0.50 | 1463 | -0.036 | 0.05 | -0.14 | 0.07 | 7.93 𝗑 10^-1^ |
| **SLF RD** | 1.00 | 1463 | 0.000 | 0.05 | -0.10 | 0.10 | 9.98 𝗑 10^-1^ |
| **SS RD** | 0.44 | 1463 | -0.040 | 0.05 | -0.14 | 0.06 | 7.93 𝗑 10^-1^ |
| **TAP RD** | 0.37 | 1463 | 0.046 | 0.05 | -0.06 | 0.15 | 7.93 𝗑 10^-1^ |
| **UNC RD** | 0.19 | 1463 | 0.069 | 0.05 | -0.03 | 0.17 | 7.93 𝗑 10^-1^ |

| **HY3 & Matched Controls** | ***p*-value** | **Degrees of Freedom** | **Partial Cohen’s *d*** | **Standard Error** | **Lower CI** | **Upper CI** | ***p*_FDR_** |
| --- | --- | --- | --- | --- | --- | --- | --- |
| **Entire WM RD** | 0.14 | 420 | 0.143 | 0.10 | -0.04 | 0.33 | 5.93 𝗑 10^-1^ |
| **ACR RD** | 0.03 | 420 | 0.219 | 0.10 | 0.03 | 0.41 | 2.80 𝗑 10^-1^ |
| **SCR RD** | 0.39 | 420 | 0.083 | 0.10 | -0.10 | 0.27 | 6.16 𝗑 10^-1^ |
| **PCR RD** | 0.32 | 420 | 0.097 | 0.10 | -0.09 | 0.28 | 6.16 𝗑 10^-1^ |
| **ALIC RD** | 0.89 | 420 | -0.013 | 0.10 | -0.20 | 0.17 | 9.37 𝗑 10^-1^ |
| **PLIC RD** | 0.15 | 420 | -0.139 | 0.10 | -0.33 | 0.05 | 5.93 𝗑 10^-1^ |
| **RLIC RD** | 0.42 | 420 | -0.079 | 0.10 | -0.27 | 0.11 | 6.16 𝗑 10^-1^ |
| **EC RD** | 0.25 | 420 | 0.112 | 0.10 | -0.08 | 0.30 | 6.15 𝗑 10^-1^ |
| **FX RD** | 0.01 | 420 | 0.264 | 0.10 | 0.08 | 0.45 | 1.57 𝗑 10^-1^ |
| **FXST RD** | 0.35 | 420 | 0.091 | 0.10 | -0.10 | 0.28 | 6.16 𝗑 10^-1^ |
| **PTR RD** | 0.50 | 420 | 0.066 | 0.10 | -0.12 | 0.25 | 6.17 𝗑 10^-1^ |
| **GCC RD** | 0.22 | 420 | 0.121 | 0.10 | -0.07 | 0.31 | 5.93 𝗑 10^-1^ |
| **BCC RD** | 0.30 | 420 | 0.102 | 0.10 | -0.09 | 0.29 | 6.16 𝗑 10^-1^ |
| **SCC RD** | 0.58 | 420 | 0.054 | 0.10 | -0.13 | 0.24 | 6.60 𝗑 10^-1^ |
| **CGC RD** | 0.49 | 420 | 0.067 | 0.10 | -0.12 | 0.25 | 6.17 𝗑 10^-1^ |
| **CGH RD** | 0.42 | 420 | -0.079 | 0.10 | -0.27 | 0.11 | 6.16 𝗑 10^-1^ |
| **CST RD** | 0.95 | 420 | -0.006 | 0.10 | -0.19 | 0.18 | 9.54 𝗑 10^-1^ |
| **SFO RD** | 0.18 | 420 | 0.131 | 0.10 | -0.06 | 0.32 | 5.93 𝗑 10^-1^ |
| **SLF RD** | 0.21 | 420 | 0.122 | 0.10 | -0.07 | 0.31 | 5.93 𝗑 10^-1^ |
| **SS RD** | 0.19 | 420 | 0.129 | 0.10 | -0.06 | 0.32 | 5.93 𝗑 10^-1^ |
| **TAP RD** | 0.51 | 420 | 0.065 | 0.10 | -0.12 | 0.25 | 6.17 𝗑 10^-1^ |
| **UNC RD** | 0.60 | 420 | 0.051 | 0.10 | -0.14 | 0.24 | 6.60 𝗑 10^-1^ |

| **HY4/5 & Matched Controls** | ***p*-value** | **Degrees of Freedom** | **Partial Cohen’s *d*** | **Standard Error** | **Lower CI** | **Upper CI** | ***p*_FDR_** |
| --- | --- | --- | --- | --- | --- | --- | --- |
| **Entire WM RD** | 0.01 | 131 | 0.448 | 0.17 | 0.12 | 0.77 | 3.63 𝗑 10^-2^ |
| **ACR RD** | 0.01 | 131 | 0.471 | 0.17 | 0.14 | 0.80 | 3.58 𝗑 10^-2^ |
| **SCR RD** | 0.01 | 131 | 0.449 | 0.17 | 0.12 | 0.78 | 3.63 𝗑 10^-2^ |
| **PCR RD** | 0.04 | 131 | 0.362 | 0.17 | 0.04 | 0.69 | 8.05 𝗑 10^-2^ |
| **ALIC RD** | 0.04 | 131 | 0.364 | 0.17 | 0.04 | 0.69 | 8.05 𝗑 10^-2^ |
| **PLIC RD** | 0.96 | 131 | 0.008 | 0.16 | -0.31 | 0.33 | 9.61 𝗑 10^-1^ |
| **RLIC RD** | 0.83 | 131 | 0.037 | 0.16 | -0.29 | 0.36 | 8.73 𝗑 10^-1^ |
| **EC RD** | 0.01 | 131 | 0.470 | 0.17 | 0.14 | 0.80 | 3.58 𝗑 10^-2^ |
| **FX RD** | 0.00 | 131 | 0.852 | 0.17 | 0.52 | 1.19 | 6.80 𝗑 10^-5^ |
| **FXST RD** | 0.08 | 131 | 0.304 | 0.17 | -0.02 | 0.63 | 1.16 𝗑 10^-1^ |
| **PTR RD** | 0.09 | 131 | 0.294 | 0.17 | -0.03 | 0.62 | 1.23 𝗑 10^-1^ |
| **GCC RD** | 0.00 | 131 | 0.558 | 0.17 | 0.23 | 0.89 | 1.94 𝗑 10^-2^ |
| **BCC RD** | 0.02 | 131 | 0.423 | 0.17 | 0.10 | 0.75 | 4.27 𝗑 10^-2^ |
| **SCC RD** | 0.05 | 131 | 0.344 | 0.17 | 0.02 | 0.67 | 8.66 𝗑 10^-1^ |
| **CGC RD** | 0.02 | 131 | 0.421 | 0.17 | 0.09 | 0.75 | 4.27 𝗑 10^-2^ |
| **CGH RD** | 0.44 | 131 | 0.135 | 0.16 | -0.19 | 0.46 | 5.10 𝗑 10^-1^ |
| **CST RD** | 0.57 | 131 | -0.101 | 0.16 | -0.42 | 0.22 | 6.23 𝗑 10^-1^ |
| **SFO RD** | 0.01 | 131 | 0.493 | 0.17 | 0.17 | 0.82 | 3.58 𝗑 10^-2^ |
| **SLF RD** | 0.06 | 131 | 0.326 | 0.17 | 0.00 | 0.65 | 9.45 𝗑 10^-2^ |
| **SS RD** | 0.39 | 131 | 0.150 | 0.16 | -0.17 | 0.47 | 4.79 𝗑 10^-1^ |
| **TAP RD** | 0.05 | 131 | 0.344 | 0.17 | 0.02 | 0.67 | 8.66 𝗑 10-^2^ |
| **UNC RD** | 0.06 | 131 | 0.335 | 0.17 | 0.01 | 0.66 | 8.99 𝗑 10^-2^ |

#### 2.2 Between-group differences in DTI metrics: Total PD and Controls

##### 2.2.1 Data: Differences in FA between the Total PD group and Controls

| **Total PD & Controls** | ***p*-value** | **Degrees of Freedom** | **Partial Cohen’s *d*** | **Standard Error** | **Lower CI** | **Upper CI** | ***p*_FDR_** |
| --- | --- | --- | --- | --- | --- | --- | --- |
| **Entire WM FA** | 0.65 | 2517 | -0.019 | 0.04 | -0.10 | 0.06 | 8.07 𝗑 10^-1^ |
| **ACR FA** | 0.04 | 2517 | -0.086 | 0.04 | -0.16 | -0.01 | 2.96 𝗑 10^-1^ |
| **SCR FA** | 0.48 | 2517 | 0.029 | 0.04 | -0.05 | 0.11 | 7.62 𝗑 10^-1^ |
| **PCR FA** | 0.14 | 2517 | -0.061 | 0.04 | -0.14 | 0.02 | 4.49 𝗑 10^-1^ |
| **ALIC FA** | 0.92 | 2517 | -0.004 | 0.04 | -0.08 | 0.07 | 9.60 𝗑 10^-1^ |
| **PLIC FA** | 0.10 | 2517 | 0.068 | 0.04 | -0.01 | 0.15 | 4.49 𝗑 10^-1^ |
| **RLIC FA** | 0.63 | 2517 | 0.020 | 0.04 | -0.06 | 0.10 | 8.07 𝗑 10^-1^ |
| **EC FA** | 0.09 | 2517 | -0.070 | 0.04 | -0.15 | 0.01 | 4.49 𝗑 10^-1^ |
| **FX FA** | 0.00 | 2517 | -0.245 | 0.04 | -0.32 | -0.17 | 1.20 𝗑 10^-7^ |
| **FXST FA** | 0.21 | 2517 | -0.053 | 0.04 | -0.13 | 0.03 | 4.71 𝗑 10^-1^ |
| **PTR FA** | 0.03 | 2517 | -0.092 | 0.04 | -0.17 | -0.01 | 2.96 𝗑 10^-1^ |
| **GCC FA** | 0.21 | 2517 | -0.052 | 0.04 | -0.13 | 0.03 | 4.71 𝗑 10^-1^ |
| **BCC FA** | 0.99 | 2517 | 0.001 | 0.04 | -0.08 | 0.08 | 9.89 𝗑 10^-1^ |
| **SCC FA** | 0.77 | 2517 | 0.012 | 0.04 | -0.07 | 0.09 | 8.72 𝗑 10^-1^ |
| **CGC FA** | 0.13 | 2517 | -0.063 | 0.04 | -0.14 | 0.02 | 4.49 𝗑 10^-1^ |
| **CGH FA** | 0.63 | 2517 | 0.020 | 0.04 | -0.06 | 0.10 | 8.07 𝗑 10^-1^ |
| **CST FA** | 0.46 | 2517 | 0.031 | 0.04 | -0.05 | 0.11 | 7.62 𝗑 10^-1^ |
| **SFO FA** | 0.66 | 2517 | -0.018 | 0.04 | -0.10 | 0.06 | 8.07 𝗑 10^-1^ |
| **SLF FA** | 0.17 | 2517 | -0.058 | 0.04 | -0.14 | 0.02 | 4.55 𝗑 10^-1^ |
| **SS FA** | 0.27 | 2517 | -0.046 | 0.04 | -0.12 | 0.03 | 5.42 𝗑 10^-1^ |
| **TAP FA** | 0.79 | 2517 | -0.011 | 0.04 | -0.09 | 0.07 | 8.72 𝗑 10^-1^ |
| **UNC FA** | 0.42 | 2517 | -0.034 | 0.04 | -0.11 | 0.04 | 7.62 𝗑 10^-1^ |

##### 2.2.2 Data: Differences in MD between the Total PD group and Controls

| **Total PD & Controls** | ***p*-value** | **Degrees of Freedom** | **Partial Cohen’s *d*** | **Standard Error** | **Lower CI** | **Upper CI** | ***p*_FDR_** |
| --- | --- | --- | --- | --- | --- | --- | --- |
| **Entire WM MD** | 0.84 | 2517 | 0.008 | 0.04 | -0.07 | 0.09 | 8.39 𝗑 10^-1^ |
| **ACR MD** | 0.14 | 2517 | 0.062 | 0.04 | -0.02 | 0.14 | 3.50 𝗑 10^-1^ |
| **SCR MD** | 0.17 | 2517 | 0.057 | 0.04 | -0.02 | 0.14 | 3.65 𝗑 10^-1^ |
| **PCR MD** | 0.18 | 2517 | 0.056 | 0.04 | -0.02 | 0.13 | 3.65 𝗑 10^-1^ |
| **ALIC MD** | 0.14 | 2517 | -0.061 | 0.04 | -0.14 | 0.02 | 3.50 𝗑 10^-1^ |
| **PLIC MD** | 0.06 | 2517 | -0.078 | 0.04 | -0.16 | 0.00 | 2.70 𝗑 10^-1^ |
| **RLIC MD** | 0.01 | 2517 | -0.107 | 0.04 | -0.19 | -0.03 | 7.54 𝗑 10^-2^ |
| **EC MD** | 0.64 | 2517 | 0.020 | 0.04 | -0.06 | 0.10 | 7.42 𝗑 10^-1^ |
| **FX MD** | 0.00 | 2517 | 0.166 | 0.04 | 0.09 | 0.24 | 1.27 𝗑 10^-3^ |
| **FXST MD** | 0.03 | 2517 | -0.090 | 0.04 | -0.17 | -0.01 | 1.77 𝗑 10^-1^ |
| **PTR MD** | 0.67 | 2517 | -0.018 | 0.04 | -0.10 | 0.06 | 7.42 𝗑 10^-1^ |
| **GCC MD** | 0.60 | 2517 | 0.022 | 0.04 | -0.06 | 0.10 | 7.42 𝗑 10^-1^ |
| **BCC MD** | 0.30 | 2517 | 0.043 | 0.04 | -0.03 | 0.12 | 5.16 𝗑 10^-1^ |
| **SCC MD** | 0.64 | 2517 | 0.019 | 0.04 | -0.06 | 0.10 | 7.42 𝗑 10^-1^ |
| **CGC MD** | 0.34 | 2517 | -0.040 | 0.04 | -0.12 | 0.04 | 5.40 𝗑 10^-1^ |
| **CGH MD** | 0.00 | 2517 | -0.162 | 0.04 | -0.24 | -0.08 | 1.27 𝗑 10^-3^ |
| **CST MD** | 0.27 | 2517 | -0.047 | 0.04 | -0.12 | 0.03 | 4.87 𝗑 10^-1^ |
| **SFO MD** | 0.49 | 2517 | 0.029 | 0.04 | -0.05 | 0.11 | 6.80 𝗑 10^-1^ |
| **SLF MD** | 0.43 | 2517 | 0.033 | 0.04 | -0.04 | 0.11 | 6.27 𝗑 10^-1^ |
| **SS MD** | 0.14 | 2517 | -0.062 | 0.04 | -0.14 | 0.02 | 3.50 𝗑 10^-1^ |
| **TAP MD** | 0.11 | 2517 | 0.067 | 0.04 | -0.01 | 0.14 | 3.50 𝗑 10^-1^ |
| **UNC MD** | 0.81 | 2517 | 0.010 | 0.04 | -0.07 | 0.09 | 8.39 𝗑 10^-1^ |

##### 2.2.3 Data: Differences in AD between the Total PD group and Controls

| **Total PD & Controls** | ***p*-value** | **Degrees of Freedom** | **Partial Cohen’s *d*** | **Standard Error** | **Lower CI** | **Upper CI** | ***p*_FDR_** |
| --- | --- | --- | --- | --- | --- | --- | --- |
| **Entire WM AD** | 0.29 | 2517 | -0.044 | 0.040 | -0.12 | 0.03 | 4.96 𝗑 10^-1^ |
| **ACR AD** | 0.79 | 2517 | 0.011 | 0.040 | -0.07 | 0.09 | 8.24 𝗑 10^-1^ |
| **SCR AD** | 0.09 | 2517 | 0.070 | 0.040 | -0.01 | 0.15 | 2.03 𝗑 10^-1^ |
| **PCR AD** | 0.63 | 2517 | 0.020 | 0.040 | -0.06 | 0.10 | 7.90 𝗑 10^-1^ |
| **ALIC AD** | 0.01 | 2517 | -0.107 | 0.040 | -0.19 | -0.03 | 4.55 𝗑 10^-2^ |
| **PLIC AD** | 0.19 | 2517 | -0.055 | 0.040 | -0.13 | 0.02 | 3.72 𝗑 10^-1^ |
| **RLIC AD** | 0.00 | 2517 | -0.133 | 0.040 | -0.21 | -0.06 | 1.08 𝗑 10^-2^ |
| **EC AD** | 0.65 | 2517 | -0.019 | 0.040 | -0.10 | 0.06 | 7.90 𝗑 10^-1^ |
| **FX AD** | 0.02 | 2517 | 0.102 | 0.040 | 0.02 | 0.18 | 5.59 𝗑 10^-2^ |
| **FXST AD** | 0.00 | 2517 | -0.163 | 0.040 | -0.24 | -0.08 | 1.15 𝗑 10^-3^ |
| **PTR AD** | 0.02 | 2517 | -0.098 | 0.040 | -0.18 | -0.02 | 6.03 𝗑 10^-2^ |
| **GCC AD** | 0.76 | 2517 | -0.013 | 0.040 | -0.09 | 0.07 | 8.24 𝗑 10^-1^ |
| **BCC AD** | 0.22 | 2517 | 0.052 | 0.040 | -0.03 | 0.13 | 3.98 𝗑 10^-1^ |
| **SCC AD** | 0.34 | 2517 | 0.040 | 0.040 | -0.04 | 0.12 | 5.10 𝗑 10^-1^ |
| **CGC AD** | 0.03 | 2517 | -0.092 | 0.040 | -0.17 | -0.01 | 7.68 𝗑 10^-2^ |
| **CGH AD** | 0.00 | 2517 | -0.214 | 0.040 | -0.29 | -0.14 | 7.05 𝗑 10^-6^ |
| **CST AD** | 0.55 | 2517 | -0.025 | 0.040 | -0.10 | 0.05 | 7.59 𝗑 10^-1^ |
| **SFO AD** | 0.88 | 2517 | 0.006 | 0.040 | -0.07 | 0.08 | 8.84 𝗑 10^-1^ |
| **SLF AD** | 0.70 | 2517 | -0.016 | 0.040 | -0.09 | 0.06 | 8.10 𝗑 10^-1^ |
| **SS AD** | 0.00 | 2517 | -0.125 | 0.040 | -0.20 | -0.05 | 1.55 𝗑 10^-2^ |
| **TAP AD** | 0.09 | 2517 | 0.072 | 0.040 | -0.01 | 0.15 | 2.03 𝗑 10^-1^ |
| **UNC AD** | 0.35 | 2517 | -0.039 | 0.040 | -0.12 | 0.04 | 5.10 𝗑 10^-1^ |

##### 2.2.4 Data: Differences in RD between the Total PD group and Controls

| **Total PD & Controls** | ***p*-value** | **Degrees of Freedom** | **Partial Cohen’s *d*** | **Standard Error** | **Lower CI** | **Upper CI** | ***p*_FDR_** |
| --- | --- | --- | --- | --- | --- | --- | --- |
| **Entire WM RD** | 0.28 | 2517 | 0.045 | 0.04 | -0.03 | 0.12 | 4.84 𝗑 10^-1^ |
| **ACR RD** | 0.04 | 2517 | 0.085 | 0.04 | 0.01 | 0.16 | 4.05 𝗑 10^-1^ |
| **SCR RD** | 0.42 | 2517 | 0.034 | 0.04 | -0.04 | 0.11 | 5.71 𝗑 10^-1^ |
| **PCR RD** | 0.09 | 2517 | 0.071 | 0.04 | -0.01 | 0.15 | 4.05 𝗑 10^-1^ |
| **ALIC RD** | 0.89 | 2517 | -0.006 | 0.04 | -0.08 | 0.07 | 9.37 𝗑 10^-1^ |
| **PLIC RD** | 0.09 | 2517 | -0.071 | 0.04 | -0.15 | 0.01 | 4.05 𝗑 10^-1^ |
| **RLIC RD** | 0.20 | 2517 | -0.054 | 0.04 | -0.13 | 0.02 | 4.84 𝗑 10^-1^ |
| **EC RD** | 0.28 | 2517 | 0.045 | 0.04 | -0.03 | 0.12 | 4.84 𝗑 10^-1^ |
| **FX RD** | 0.00 | 2517 | 0.193 | 0.04 | 0.12 | 0.27 | 8.98 𝗑 10^-5^ |
| **FXST RD** | 0.94 | 2517 | 0.003 | 0.04 | -0.07 | 0.08 | 9.37 𝗑 10^-1^ |
| **PTR RD** | 0.32 | 2517 | 0.042 | 0.04 | -0.04 | 0.12 | 4.84 𝗑 10^-1^ |
| **GCC RD** | 0.24 | 2517 | 0.049 | 0.04 | -0.03 | 0.13 | 4.84 𝗑 10^-1^ |
| **BCC RD** | 0.48 | 2517 | 0.030 | 0.04 | -0.05 | 0.11 | 6.15 𝗑 10^-1^ |
| **SCC RD** | 0.60 | 2517 | 0.022 | 0.04 | -0.06 | 0.10 | 6.94 𝗑 10^-1^ |
| **CGC RD** | 0.55 | 2517 | 0.025 | 0.04 | -0.05 | 0.10 | 6.78 𝗑 10^-1^ |
| **CGH RD** | 0.09 | 2517 | -0.071 | 0.04 | -0.15 | 0.01 | 4.05 𝗑 10^-1^ |
| **CST RD** | 0.30 | 2517 | -0.043 | 0.04 | -0.12 | 0.03 | 4.84 𝗑 10^-1^ |
| **SFO RD** | 0.33 | 2517 | 0.041 | 0.04 | -0.04 | 0.12 | 4.84 𝗑 10^-1^ |
| **SLF RD** | 0.16 | 2517 | 0.059 | 0.04 | -0.02 | 0.14 | 4.84 𝗑 10^-1^ |
| **SS RD** | 0.92 | 2517 | -0.004 | 0.04 | -0.08 | 0.07 | 9.37 𝗑 10^-1^ |
| **TAP RD** | 0.20 | 2517 | 0.053 | 0.04 | -0.02 | 0.13 | 4.84 𝗑 10^-1^ |
| **UNC RD** | 0.23 | 2517 | 0.051 | 0.04 | -0.03 | 0.13 | 4.84 𝗑 10^-1^ |

#### 2.3 RE-Meta Analysis: Between-Group Differences in FA and MD

Random-effects meta analysis revealed no significant differences in FA, MD, AD or RD for any region-of-interest.

| 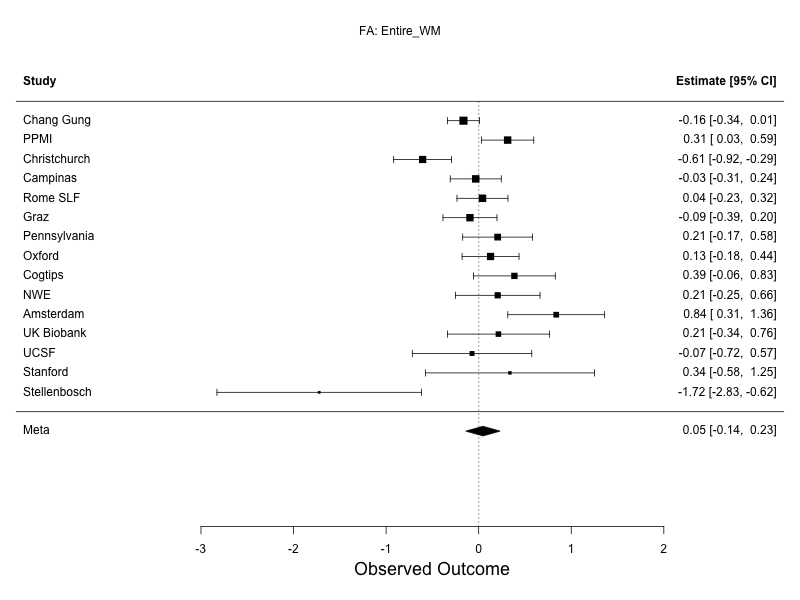 | 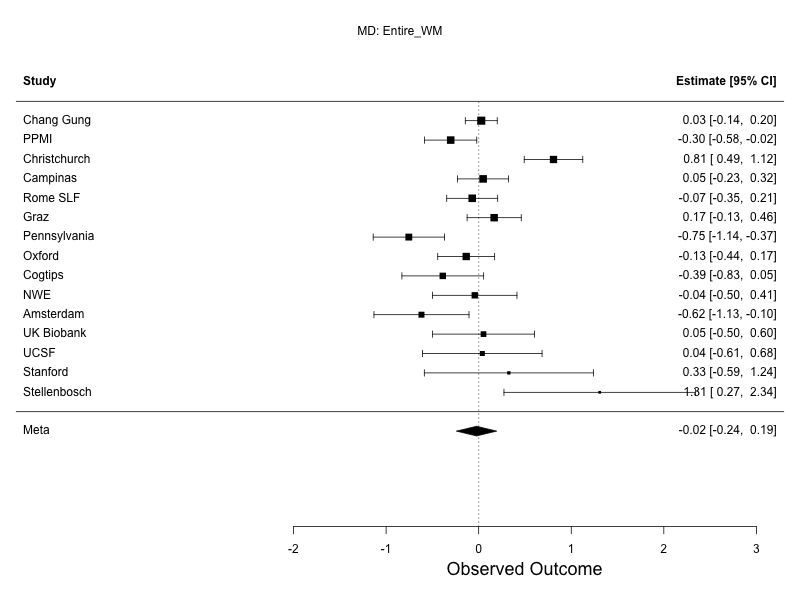 |
| --- | --- |
| 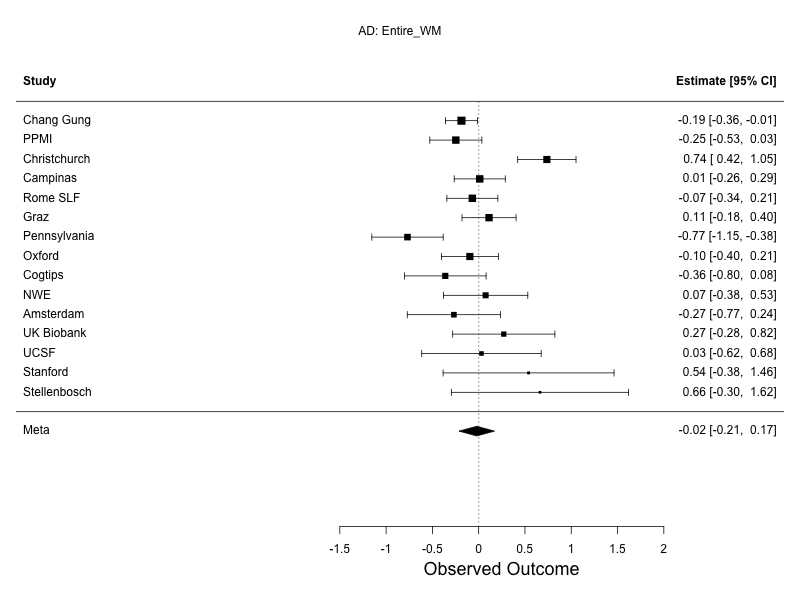 | 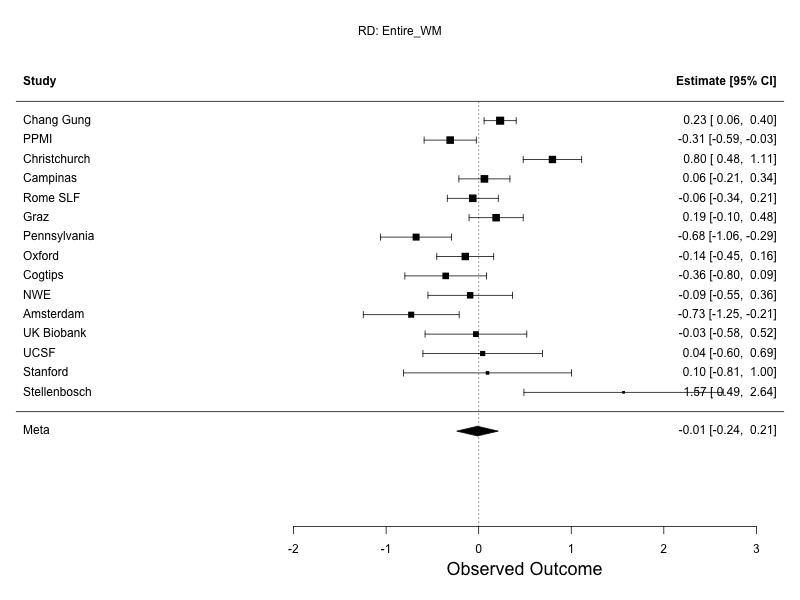 |
| **Supplementary Figure 2: Mixed effects meta analysis forest plots for the Entire WM ROI.** Differences in DTI measures between PD participants and controls, assessed using random-effects meta-analysis. Sites listed in descending order based on sample size. FA, top left; MD, top right; AD, bottom left; RD, bottom right. | |

| 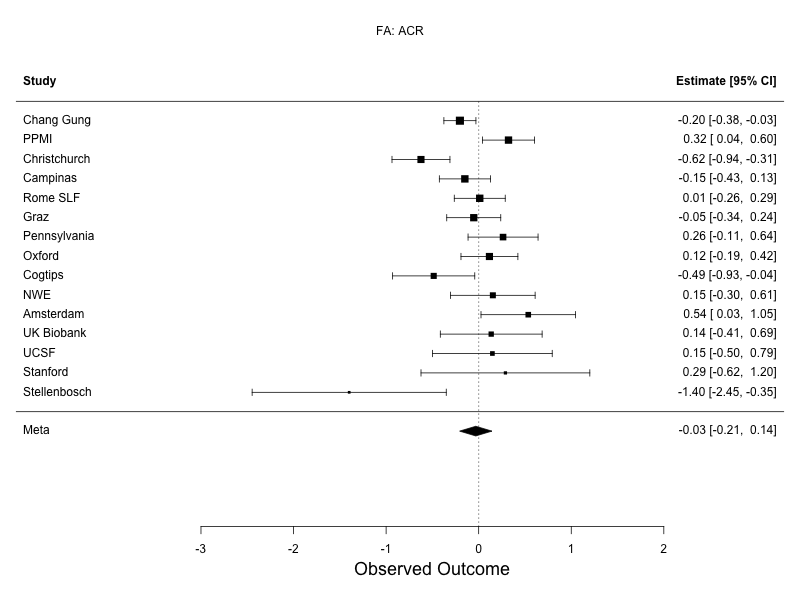 | 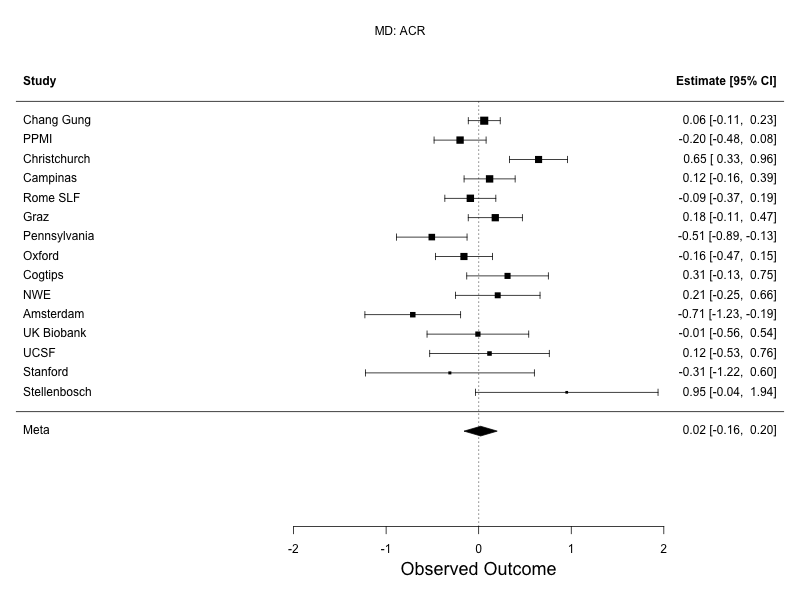 |
| --- | --- |
| 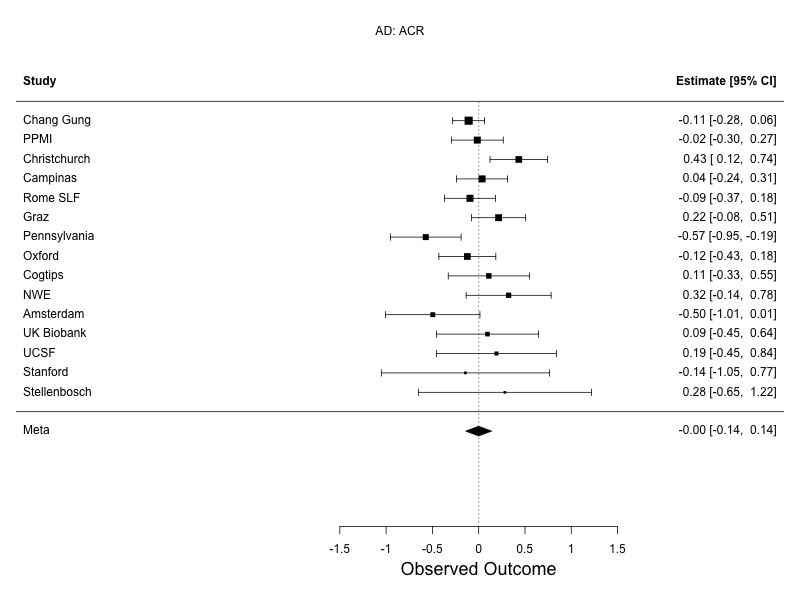 | 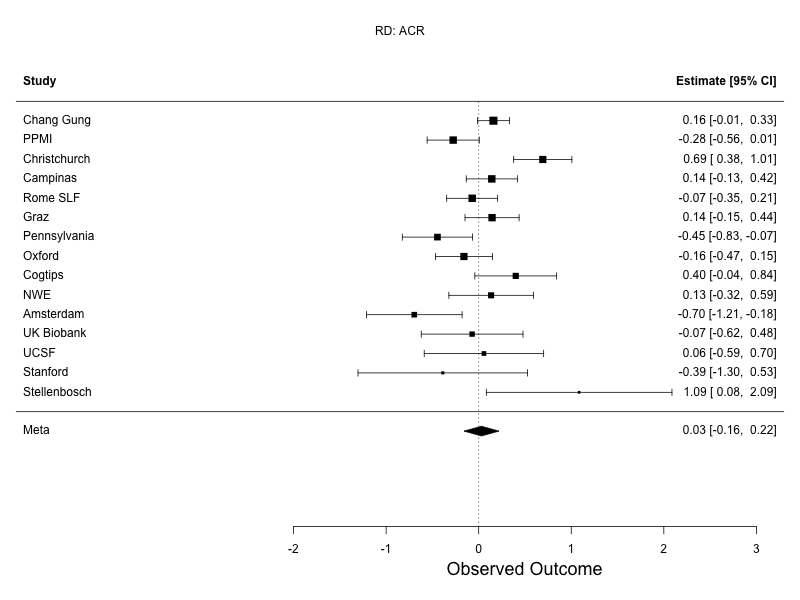 |
| **Supplementary Figure 3: Mixed effects meta analysis forest plots for the ACR.** Differences in DTI measures between PD participants and controls, assessed using random-effects meta-analysis. Sites listed in descending order based on sample size. FA, top left; MD, top right; AD, bottom left; RD, bottom right. | |

| 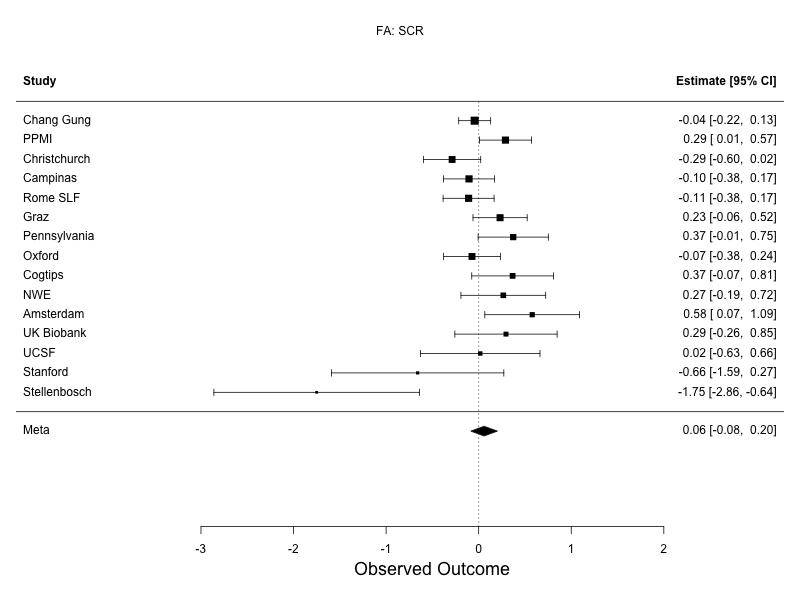 | 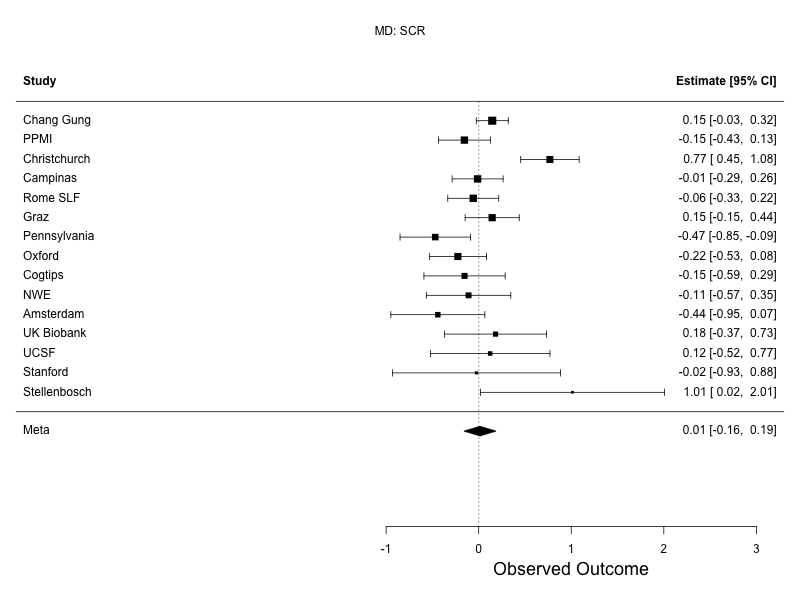 |
| --- | --- |
| 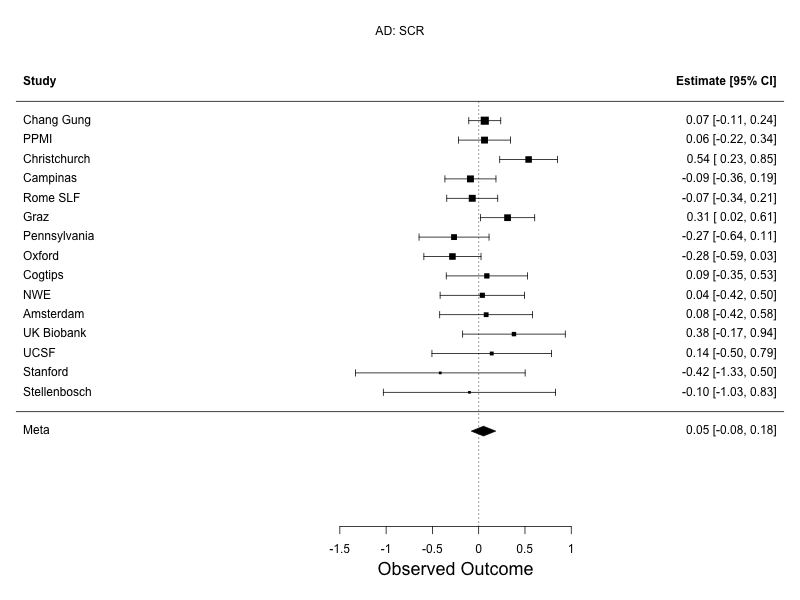 | 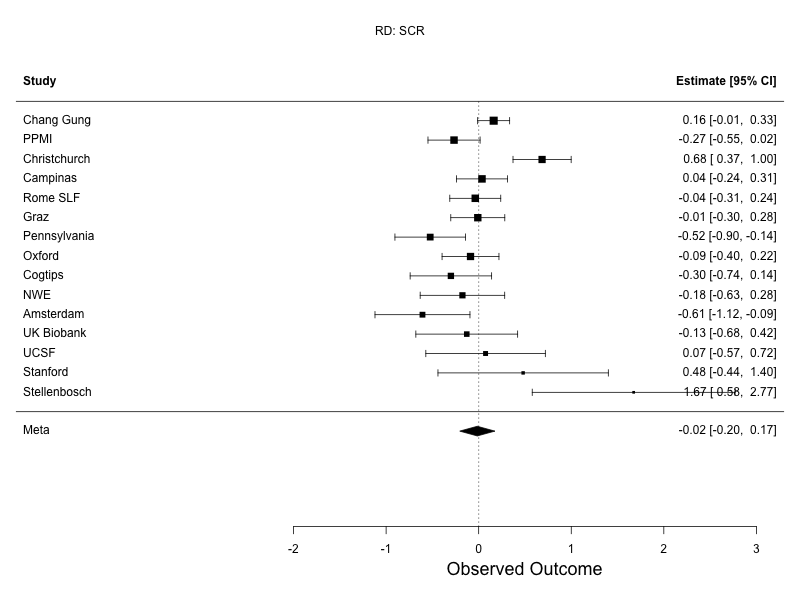 |
| **Supplementary Figure 4: Mixed effects meta analysis forest plots for the SCR.** Differences in DTI measures between PD participants and controls, assessed using random-effects meta-analysis. Sites listed in descending order based on sample size. FA, top left; MD, top right; AD, bottom left; RD, bottom right. | |

| 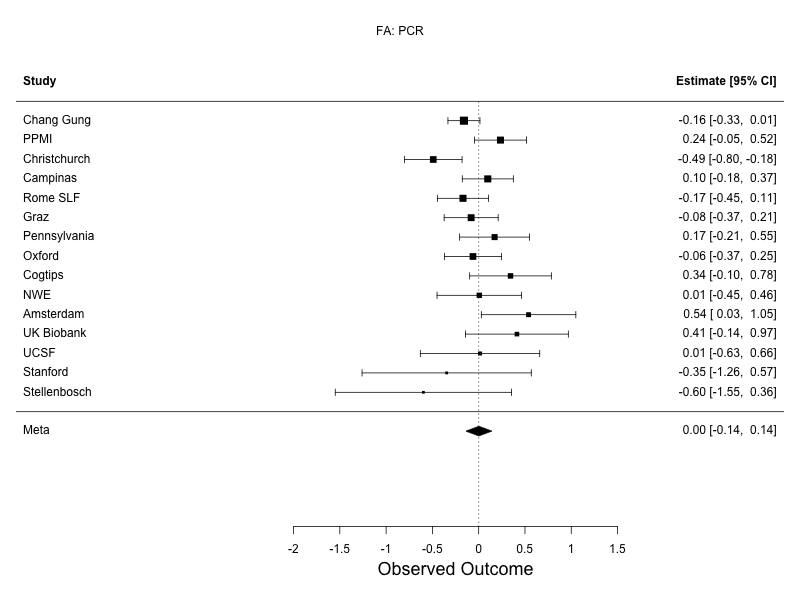 | 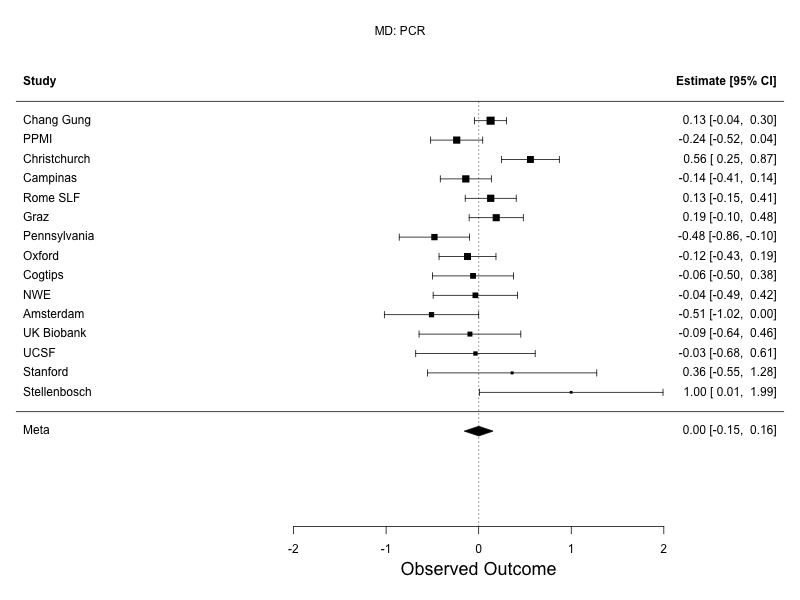 |
| --- | --- |
| 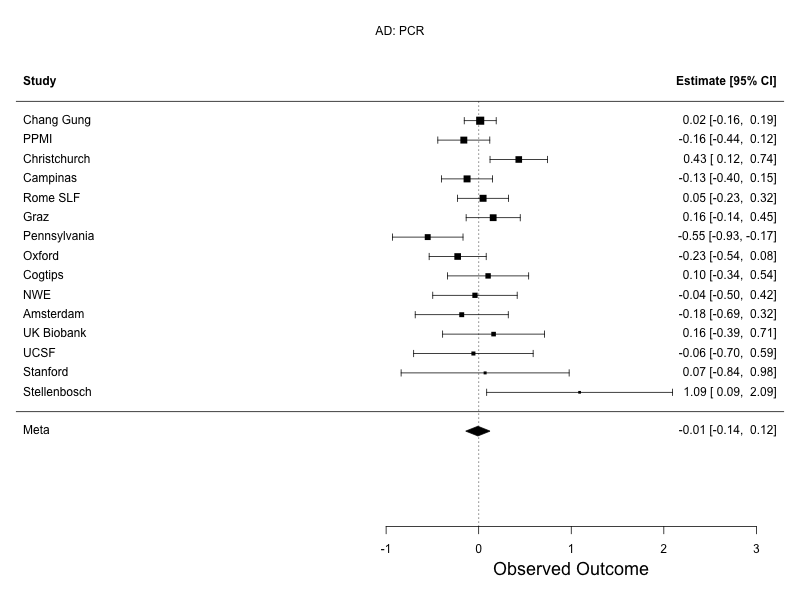 | 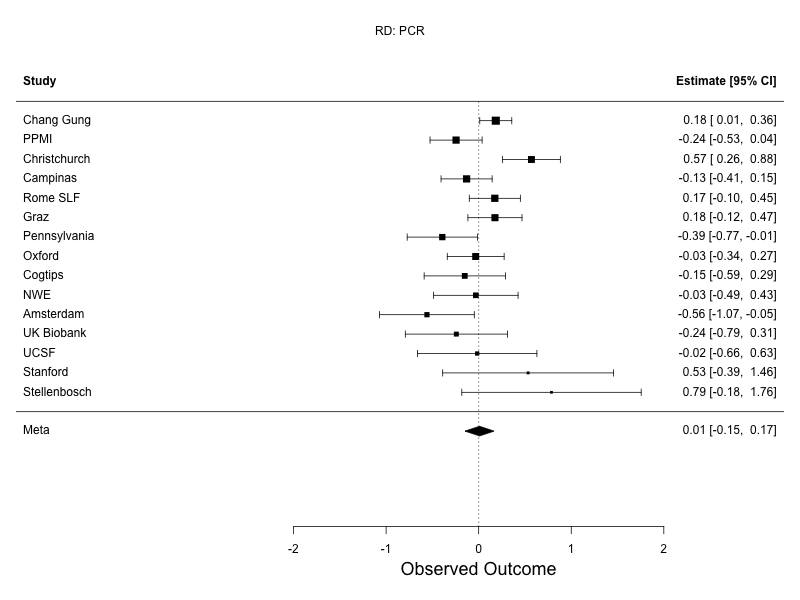 |
| **Supplementary Figure 5: Mixed effects meta analysis forest plots for the PCR.** Differences in DTI measures between PD participants and controls, assessed using random-effects meta-analysis. Sites listed in descending order based on sample size. FA, top left; MD, top right; AD, bottom left; RD, bottom right. | |

| 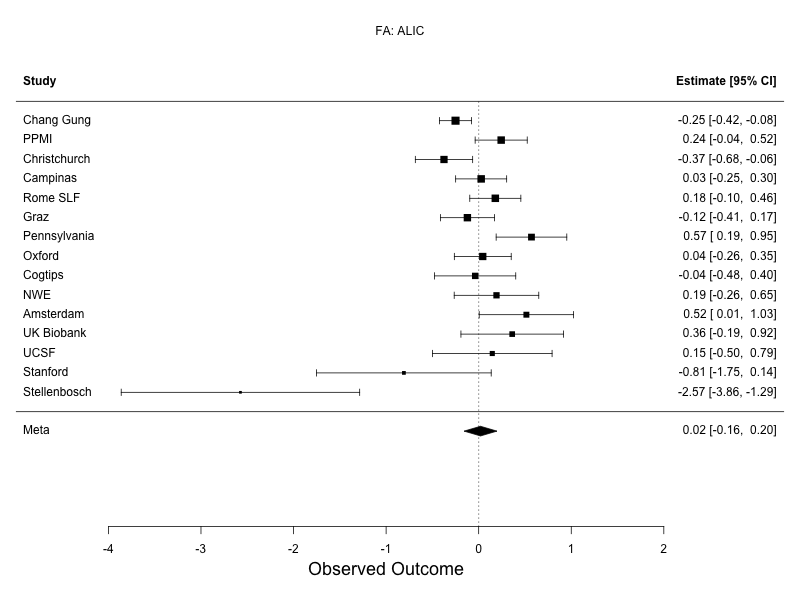 | 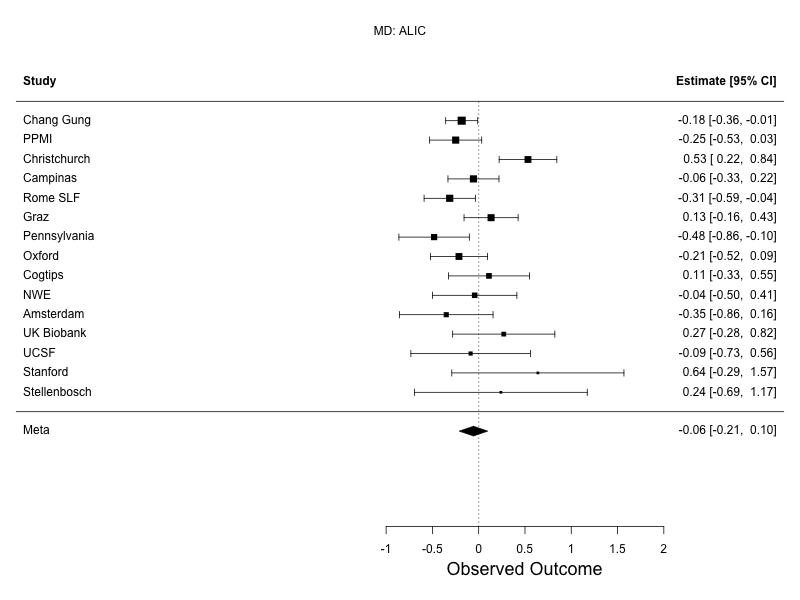 |
| --- | --- |
| 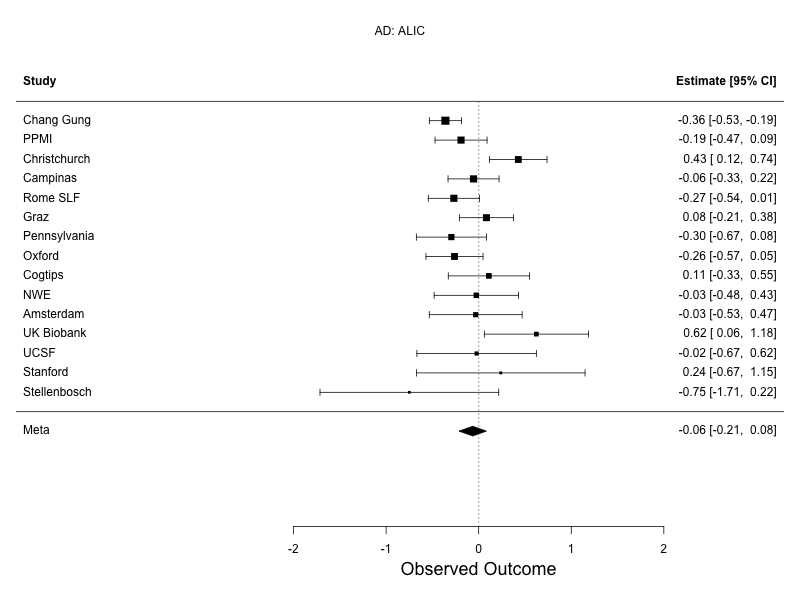 | 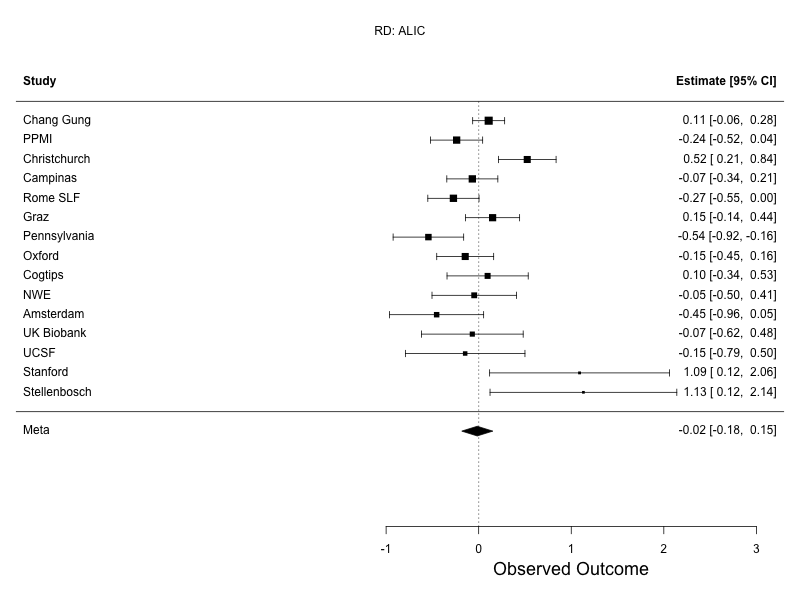 |
| **Supplementary Figure 6: Mixed effects meta analysis forest plots for the ALIC.** Differences in DTI measures between PD participants and controls, assessed using random-effects meta-analysis. Sites listed in descending order based on sample size. FA, top left; MD, top right; AD, bottom left; RD, bottom right. | |

| 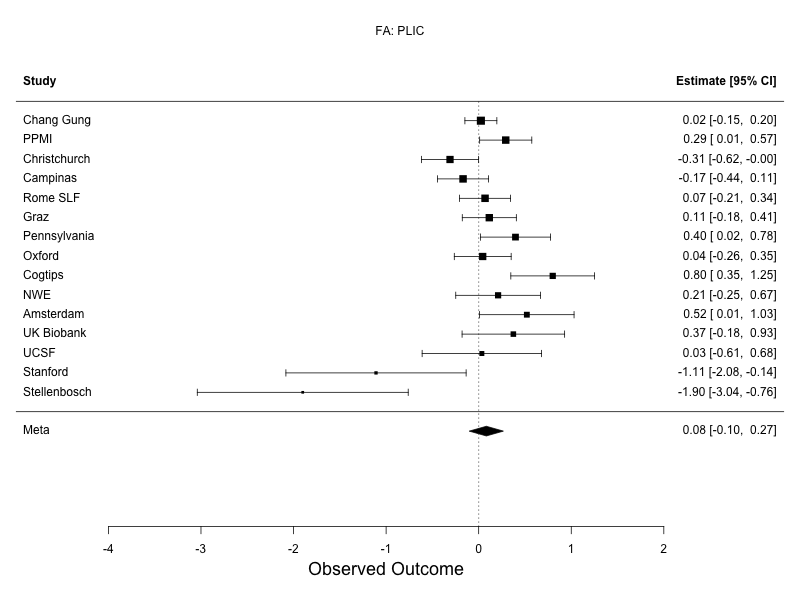 | 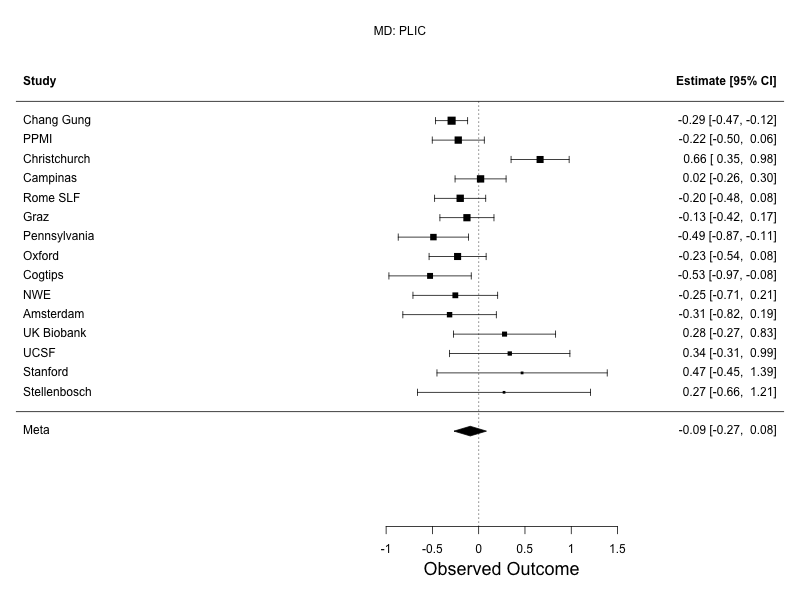 |
| --- | --- |
| 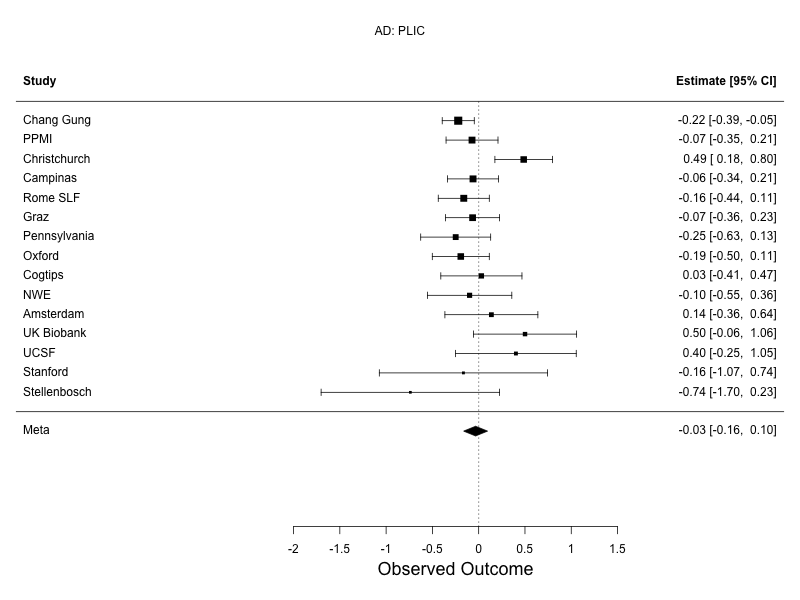 | 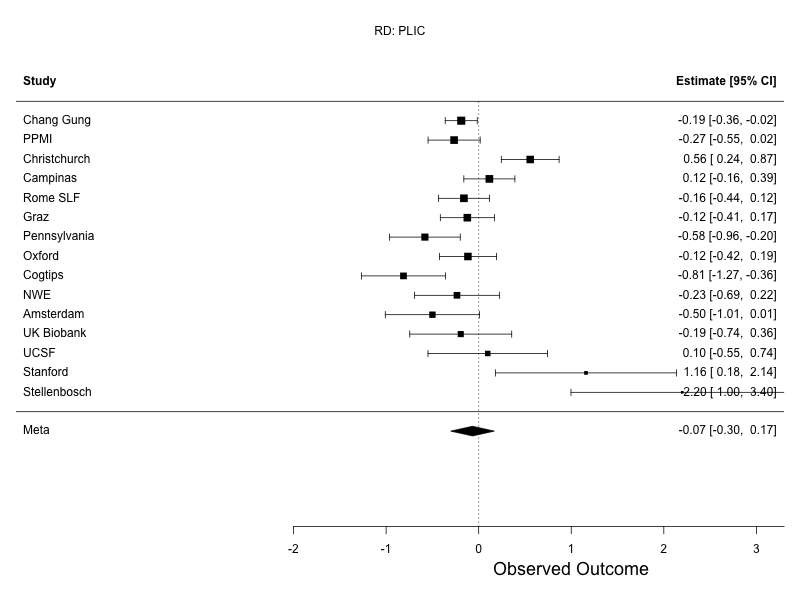 |
| **Supplementary Figure 7: Mixed effects meta analysis forest plots for the PLIC.** Differences in DTI measures between PD participants and controls, assessed using random-effects meta-analysis. Sites listed in descending order based on sample size. FA, top left; MD, top right; AD, bottom left; RD, bottom right. | |

| 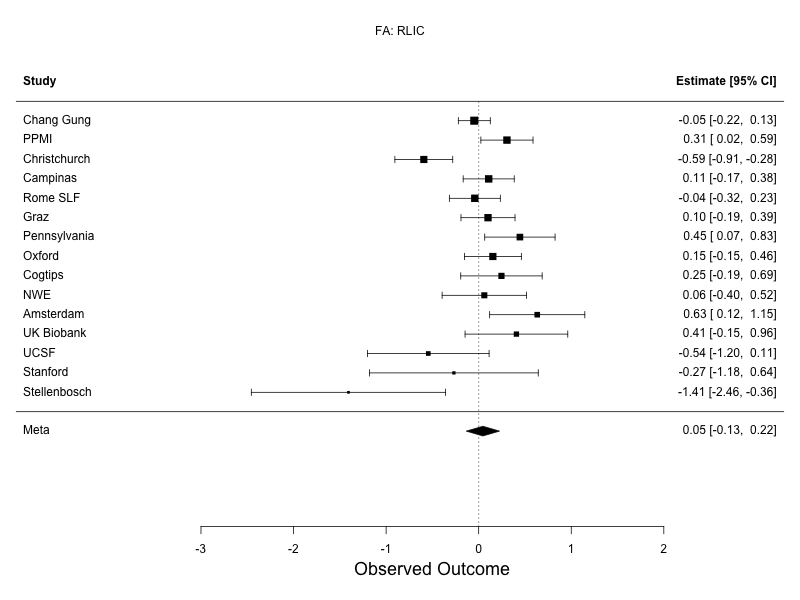 | 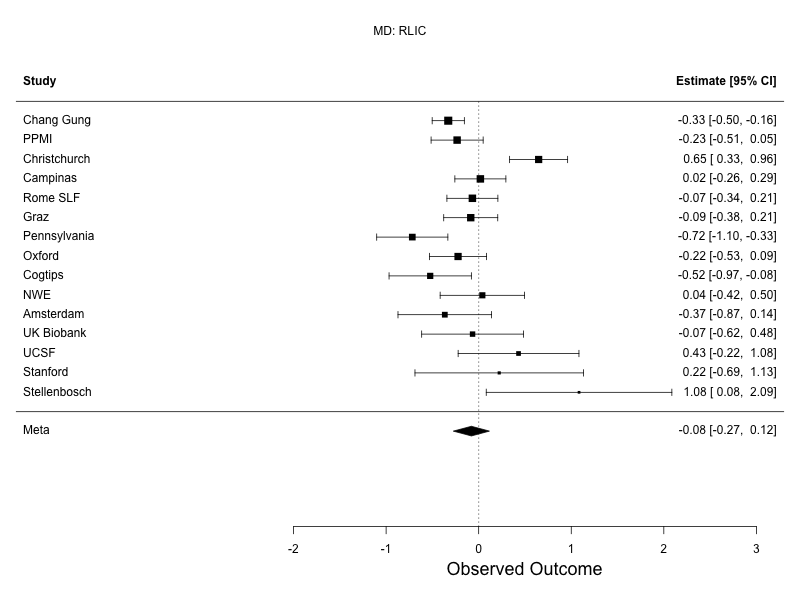 |
| --- | --- |
| 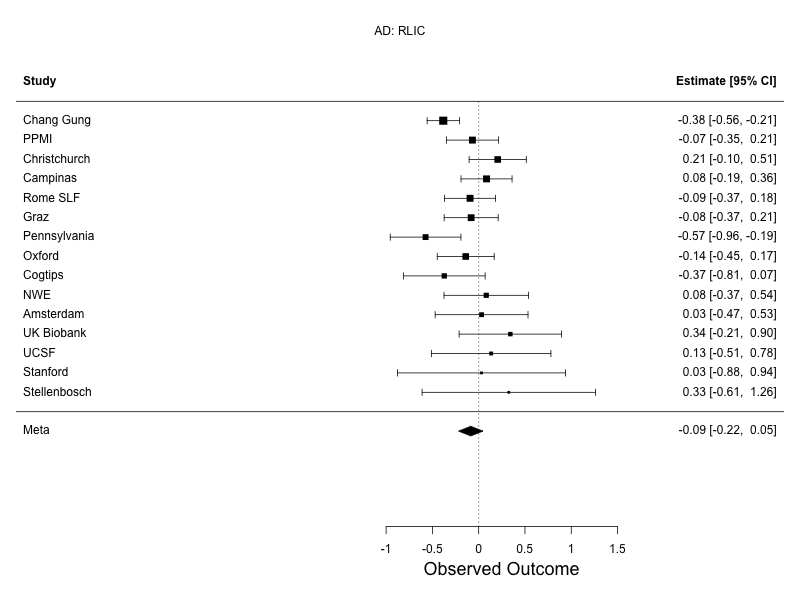 | 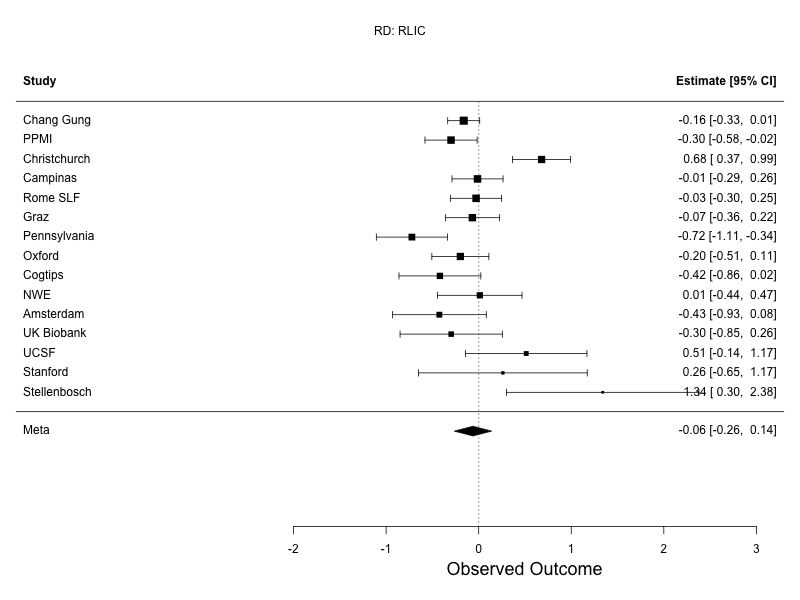 |
| **Supplementary Figure 8: Mixed effects meta analysis forest plots for the RLIC.** Differences in DTI measures between PD participants and controls, assessed using random-effects meta-analysis. Sites listed in descending order based on sample size. FA, top left; MD, top right; AD, bottom left; RD, bottom right. | |

| 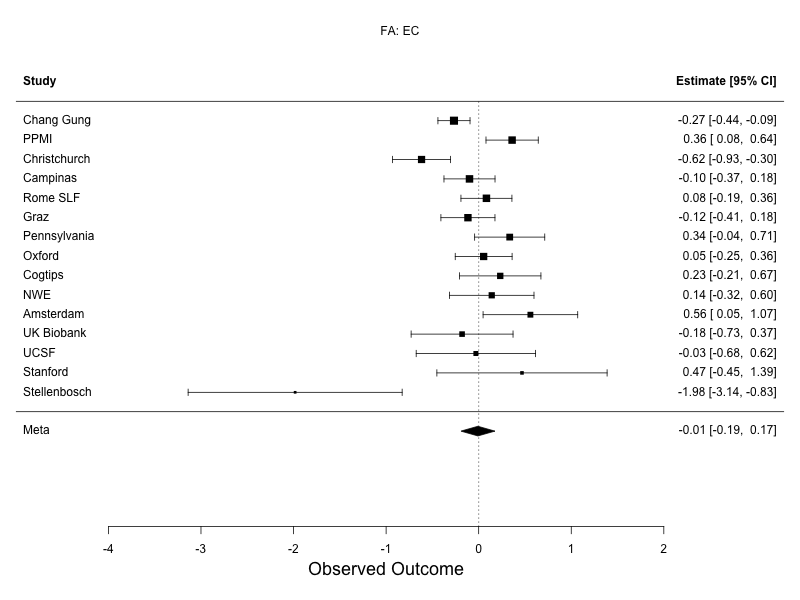 | 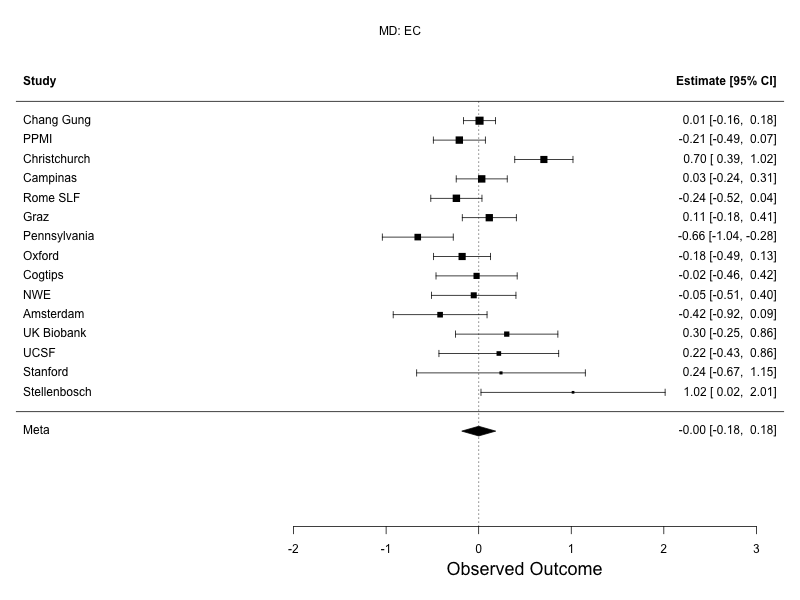 |
| --- | --- |
| 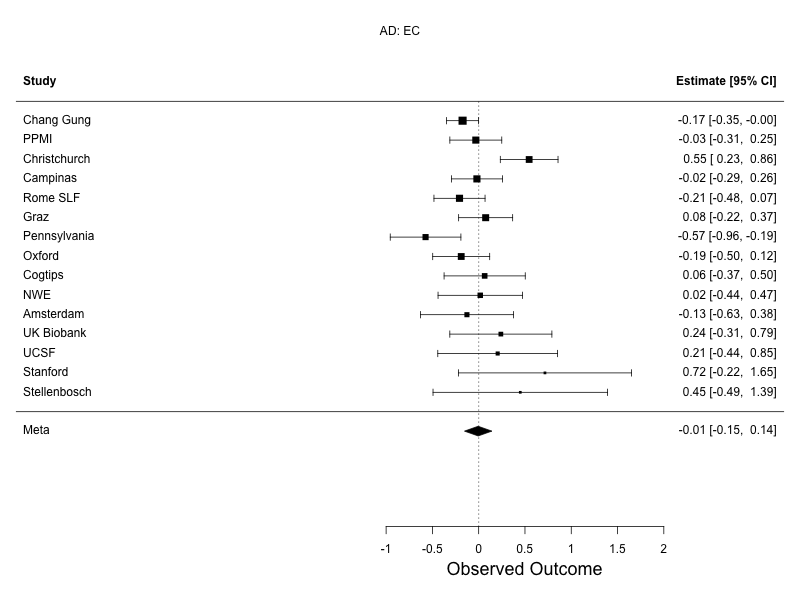 | 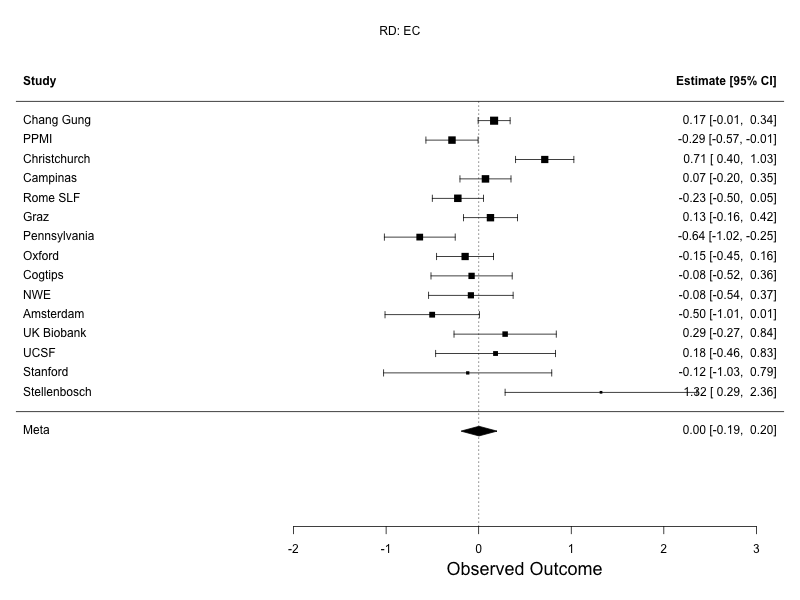 |
| **Supplementary Figure 9: Mixed effects meta analysis forest plots for the EC.** Differences in DTI measures between PD participants and controls, assessed using random-effects meta-analysis. Sites listed in descending order based on sample size. FA, top left; MD, top right; AD, bottom left; RD, bottom right. | |

| 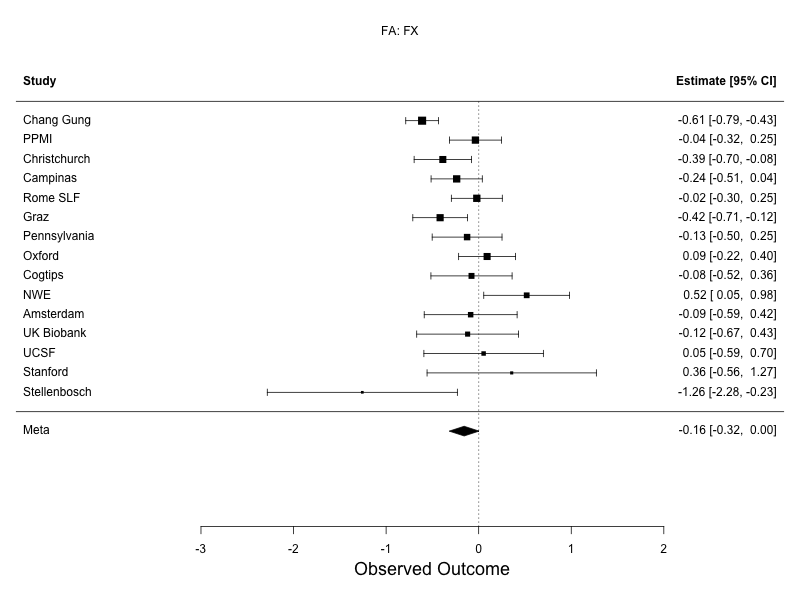 | 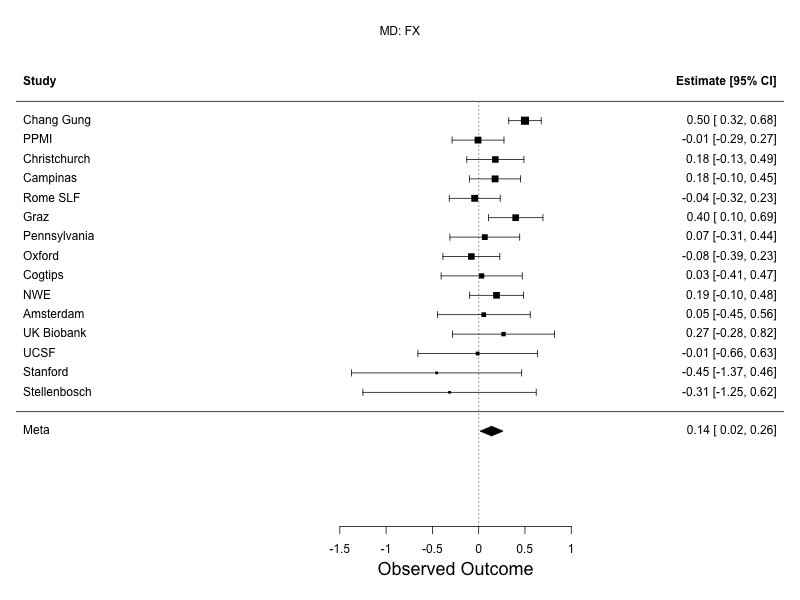 |
| --- | --- |
| 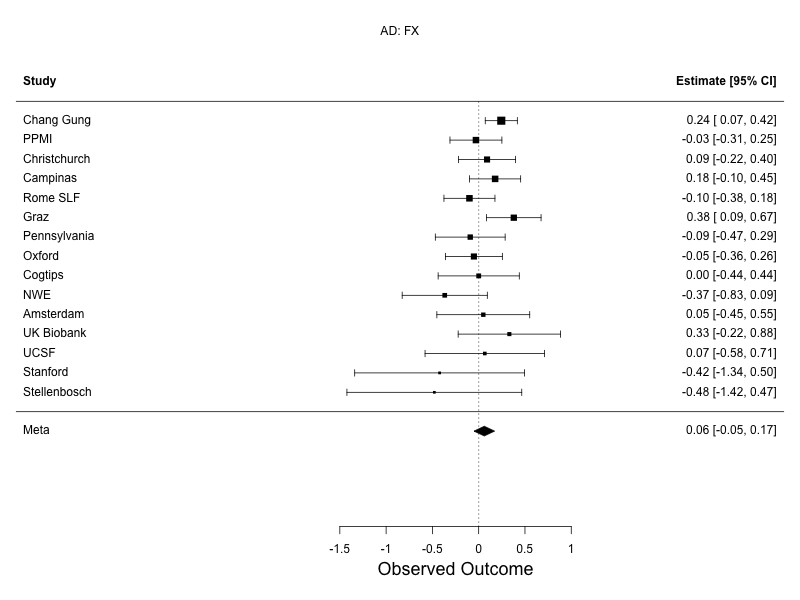 | 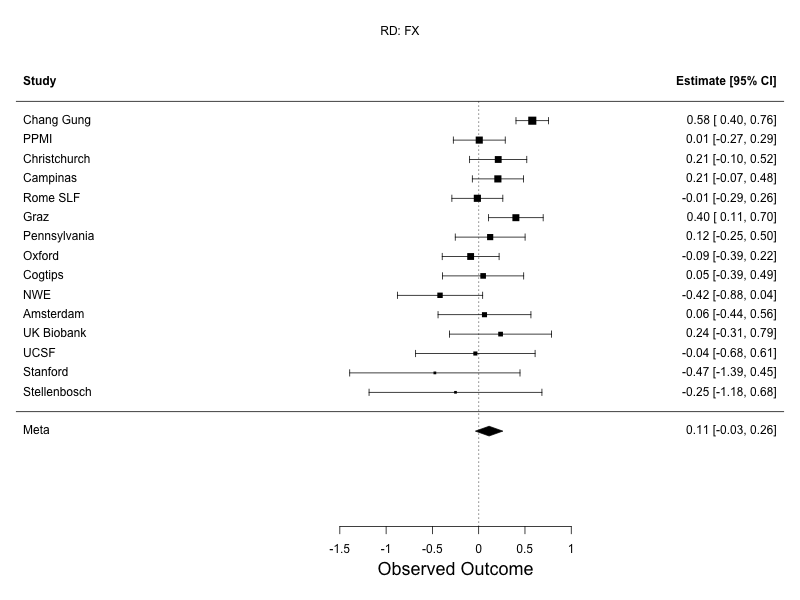 |
| **Supplementary Figure 10: Mixed effects meta analysis forest plots for the FX.** Differences in DTI measures between PD participants and controls, assessed using random-effects meta-analysis. Sites listed in descending order based on sample size. FA, top left; MD, top right; AD, bottom left; RD, bottom right. | |

| 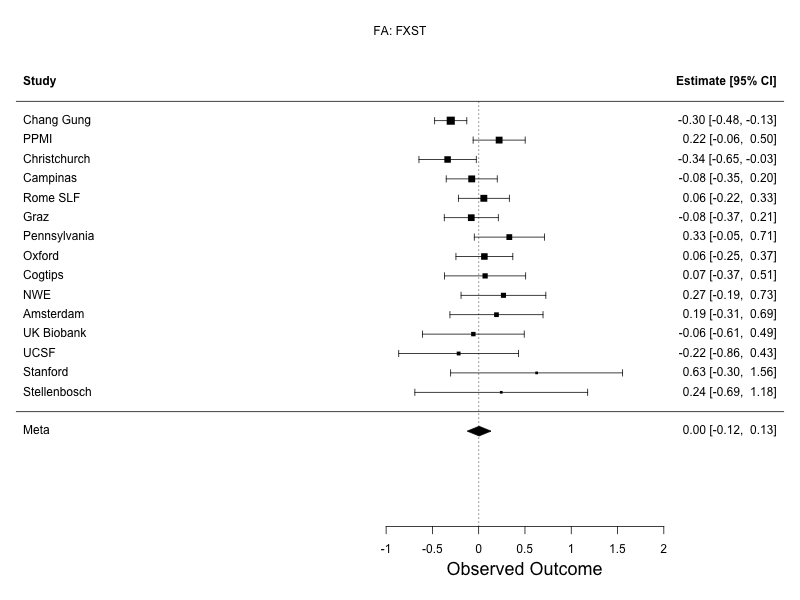 | 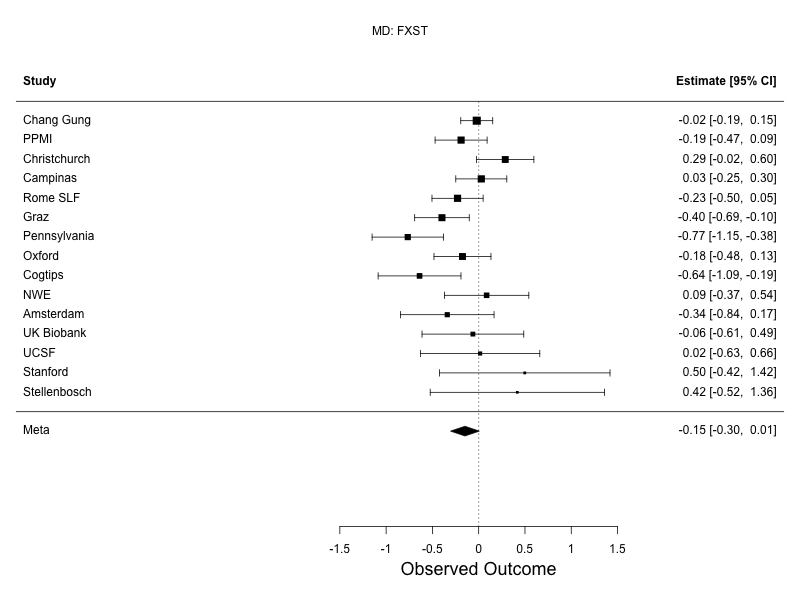 |
| --- | --- |
| 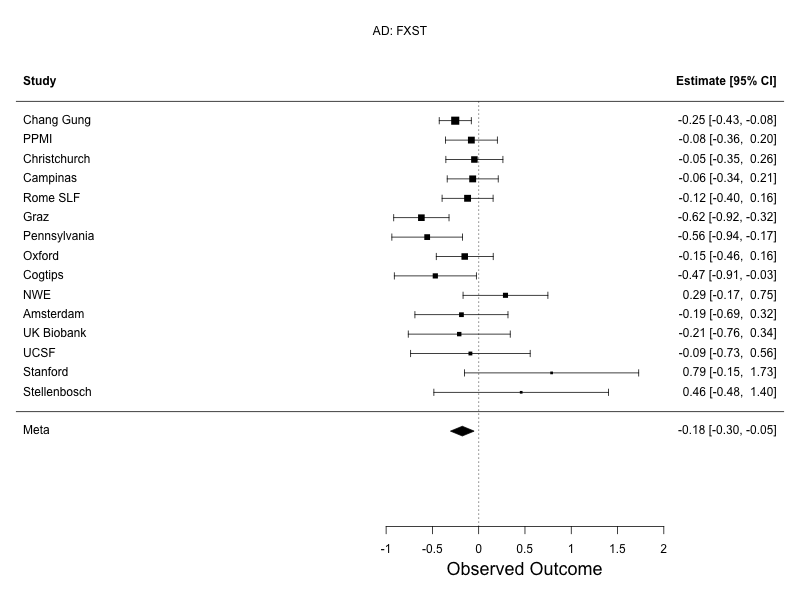 | 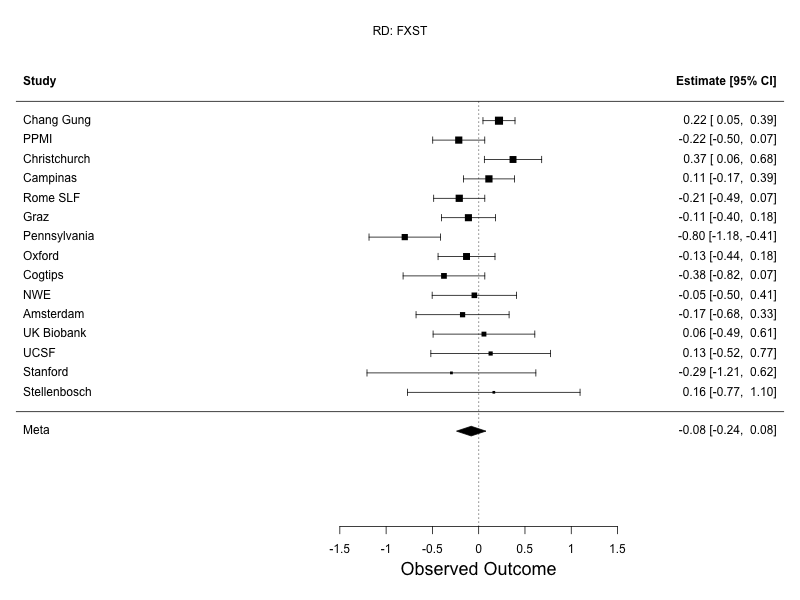 |
| **Supplementary Figure 11: Mixed effects meta analysis forest plots for the FXST.** Differences in DTI measures between PD participants and controls, assessed using random-effects meta-analysis. Sites listed in descending order based on sample size. FA, top left; MD, top right; AD, bottom left; RD, bottom right. | |

| 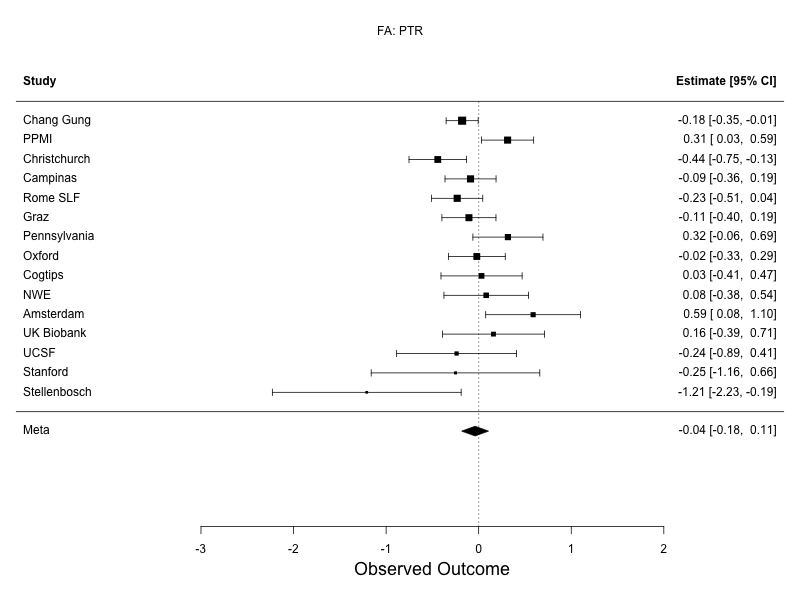 | 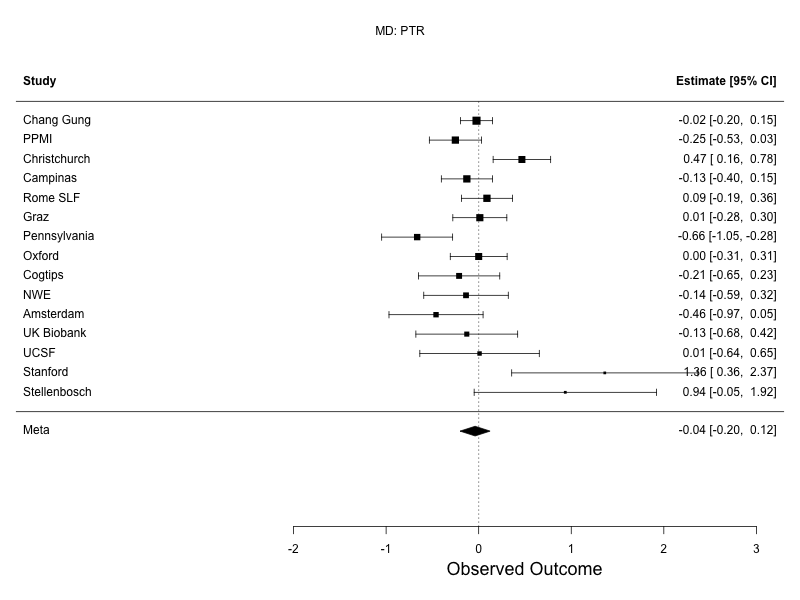 |
| --- | --- |
| 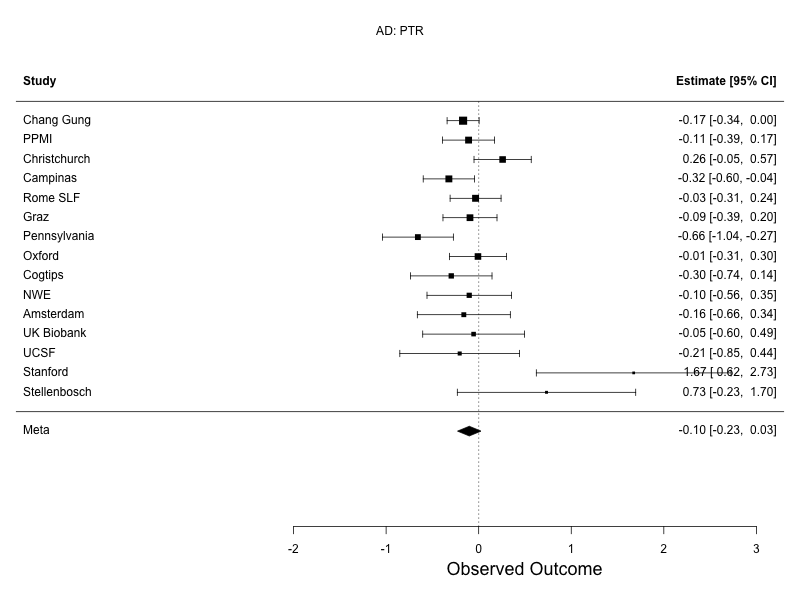 | 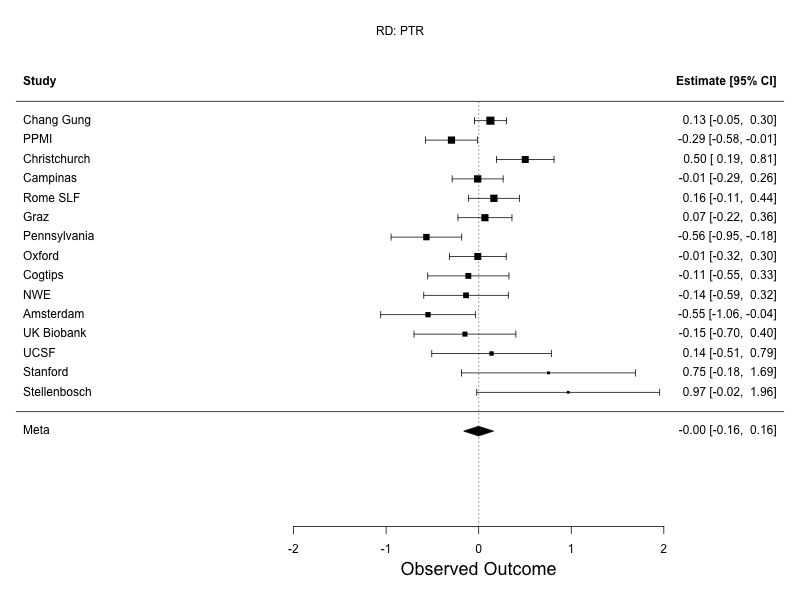 |
| **Supplementary Figure 12: Mixed effects meta analysis forest plots for the PTR.** Differences in DTI measures between PD participants and controls, assessed using random-effects meta-analysis. Sites listed in descending order based on sample size. FA, top left; MD, top right; AD, bottom left; RD, bottom right. | |

| 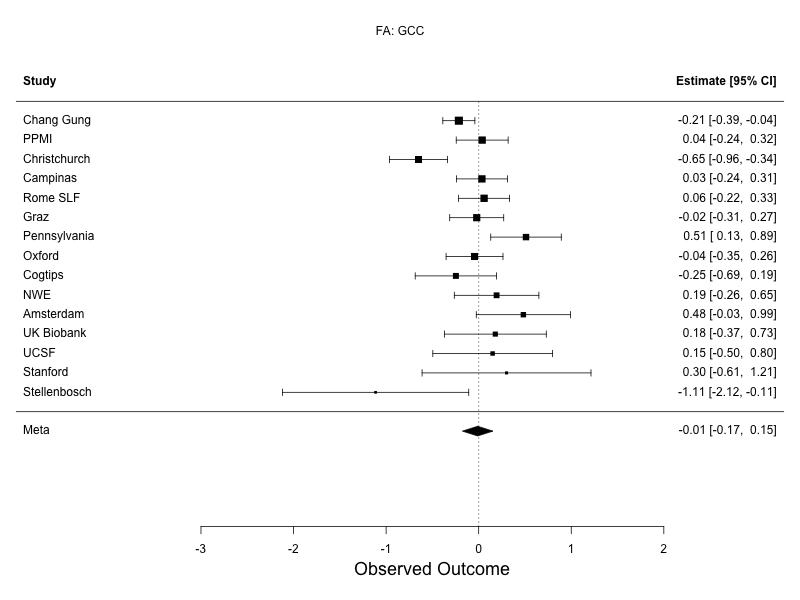 | 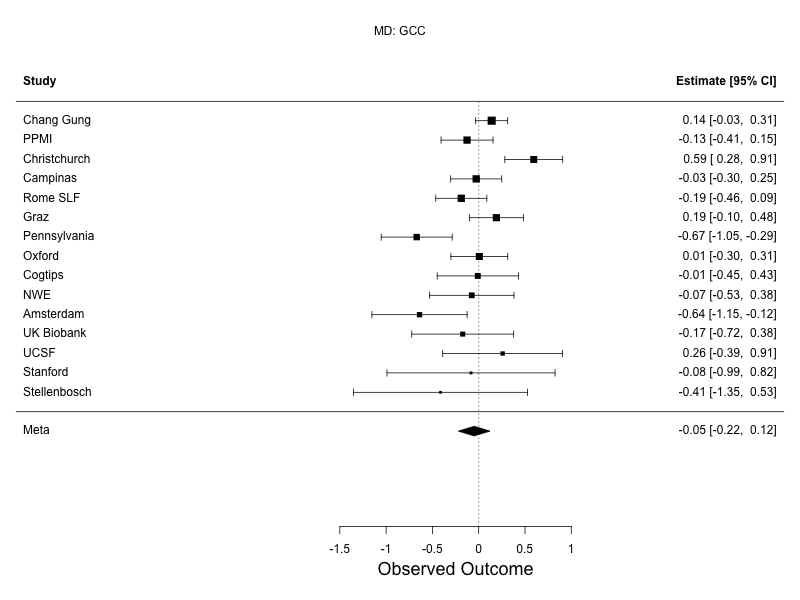 |
| --- | --- |
| 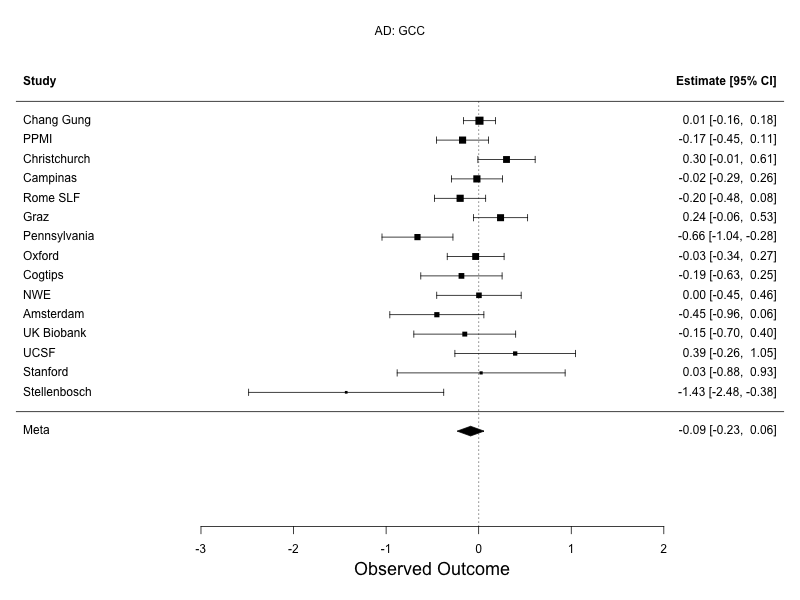 | 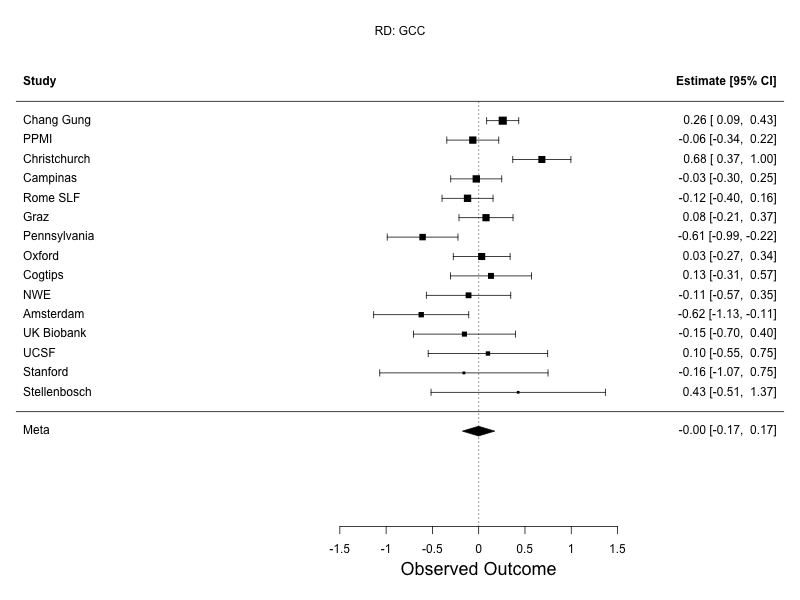 |
| **Supplementary Figure 13: Mixed effects meta analysis forest plots for the GCC.** Differences in DTI measures between PD participants and controls, assessed using random-effects meta-analysis. Sites listed in descending order based on sample size. FA, top left; MD, top right; AD, bottom left; RD, bottom right. | |

| 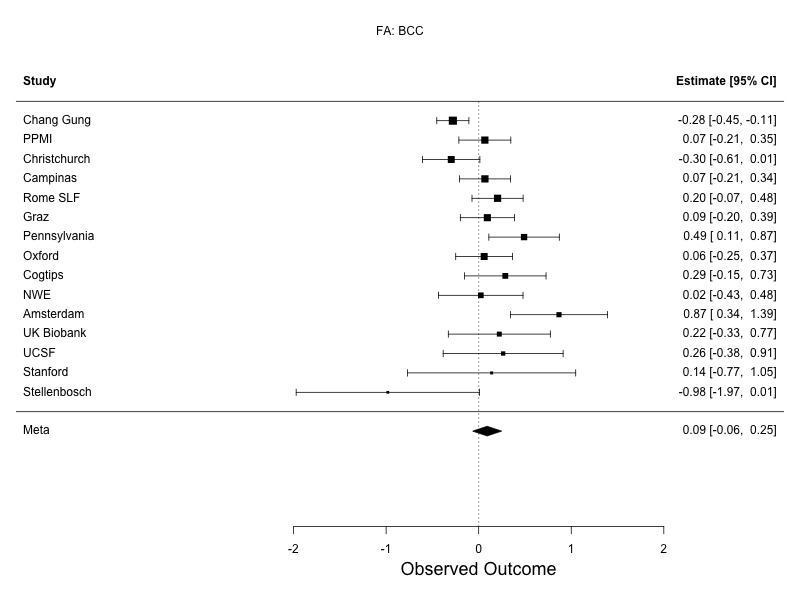 | 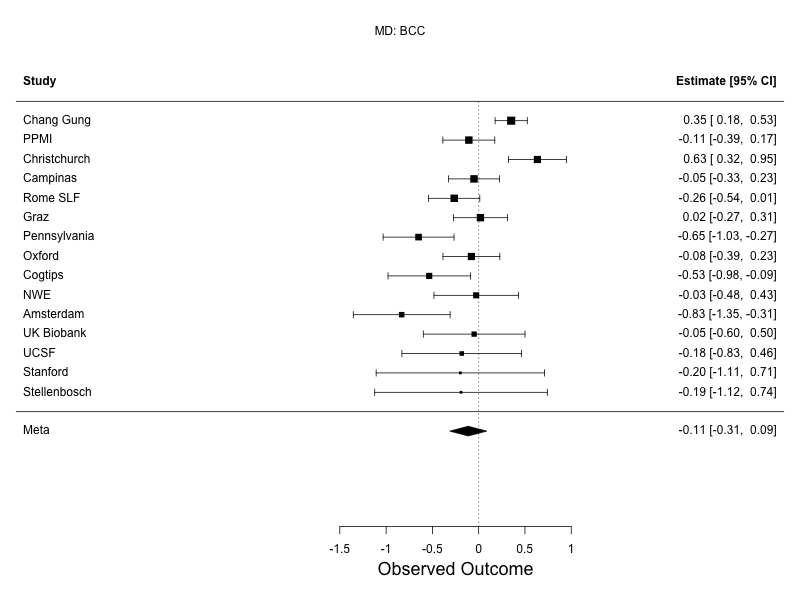 |
| --- | --- |
| 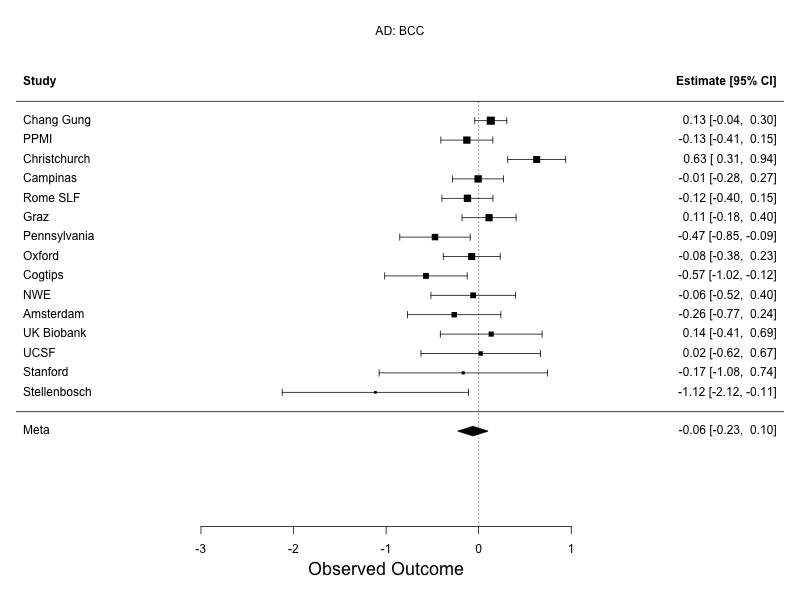 | 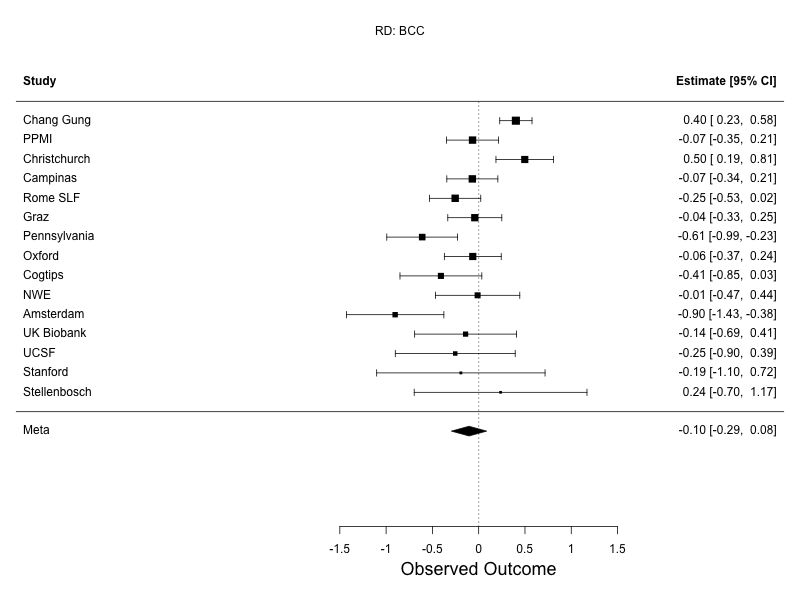 |
| **Supplementary Figure 14: Mixed effects meta analysis forest plots for the BCC.** Differences in DTI measures between PD participants and controls, assessed using random-effects meta-analysis. Sites listed in descending order based on sample size. FA, top left; MD, top right; AD, bottom left; RD, bottom right. | |

| 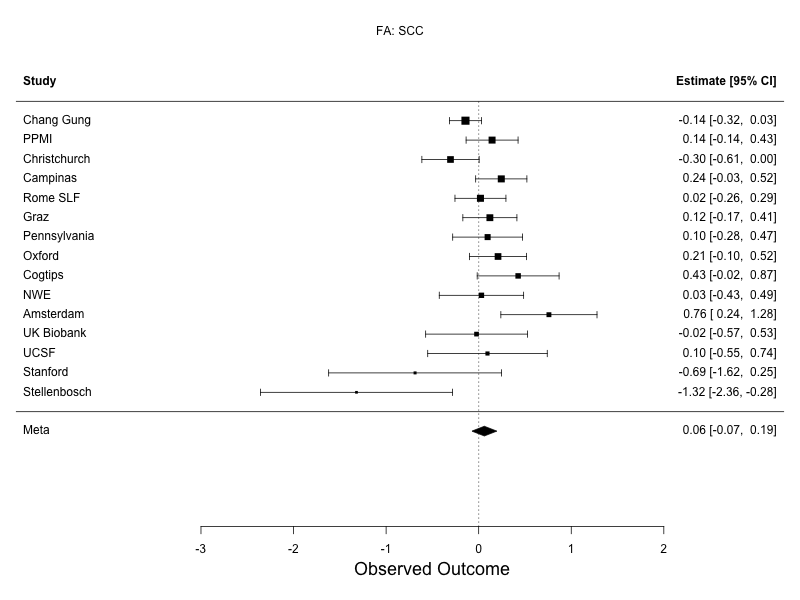 | 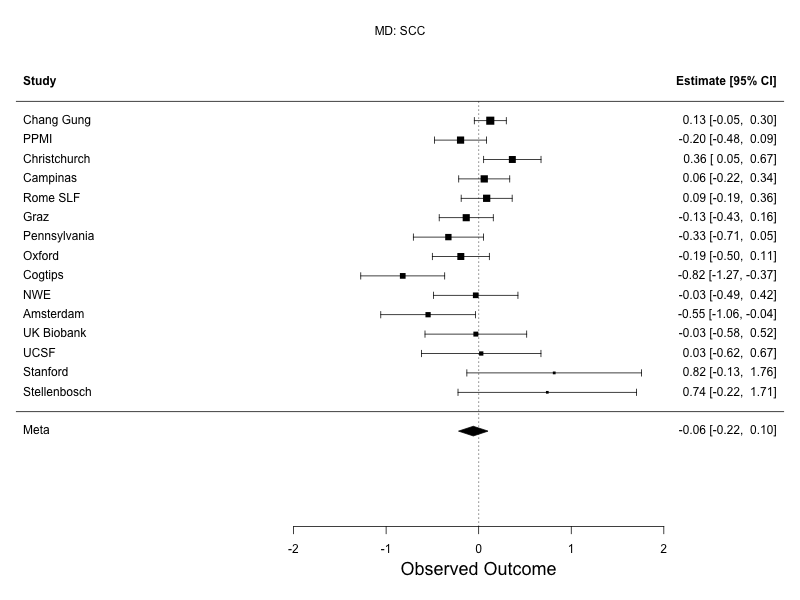 |
| --- | --- |
| 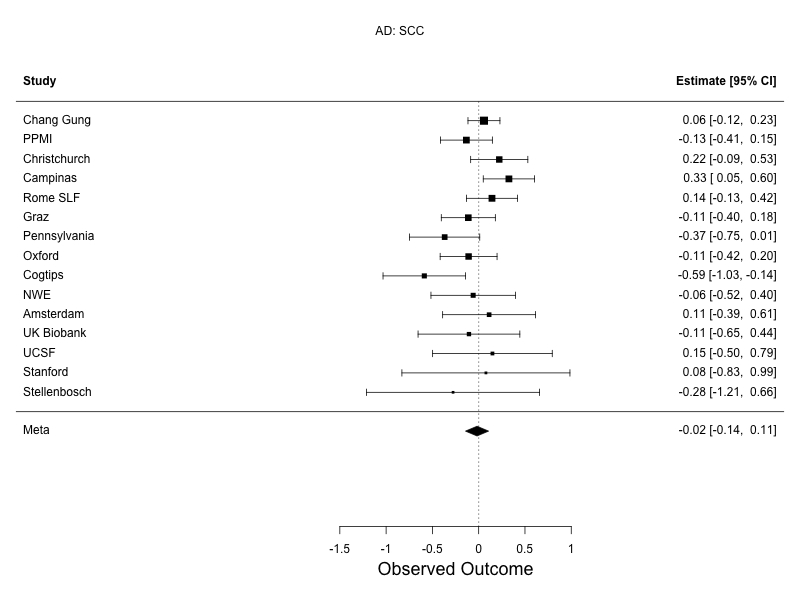 | 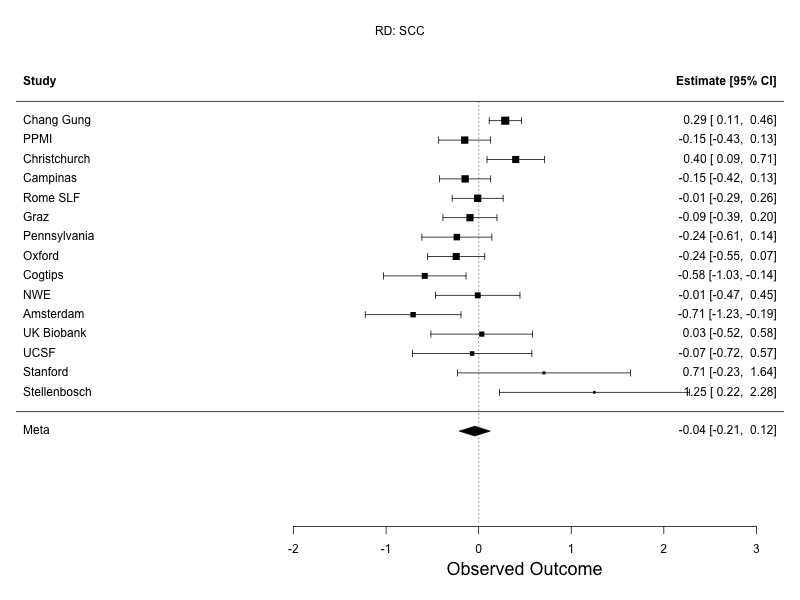 |
| **Supplementary Figure 15: Mixed effects meta analysis forest plots for the SCC.** Differences in DTI measures between PD participants and controls, assessed using random-effects meta-analysis. Sites listed in descending order based on sample size. FA, top left; MD, top right; AD, bottom left; RD, bottom right. | |

| 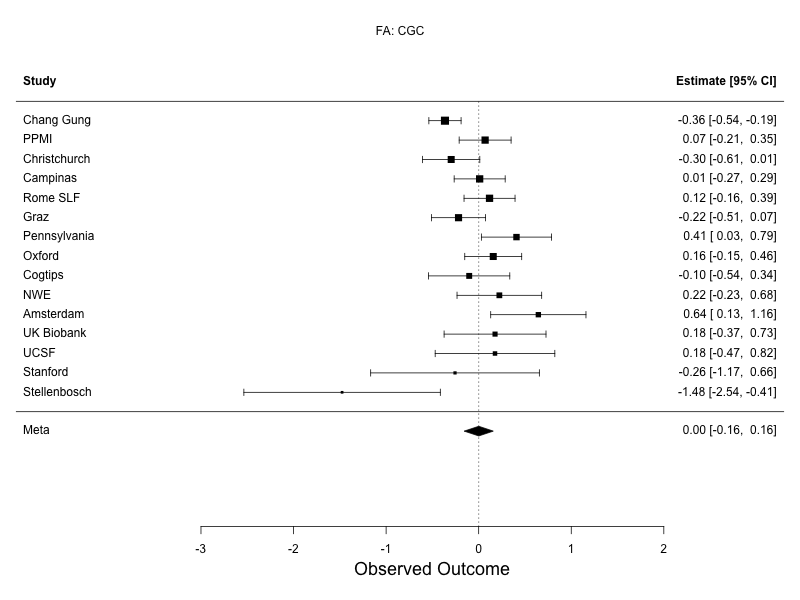 | 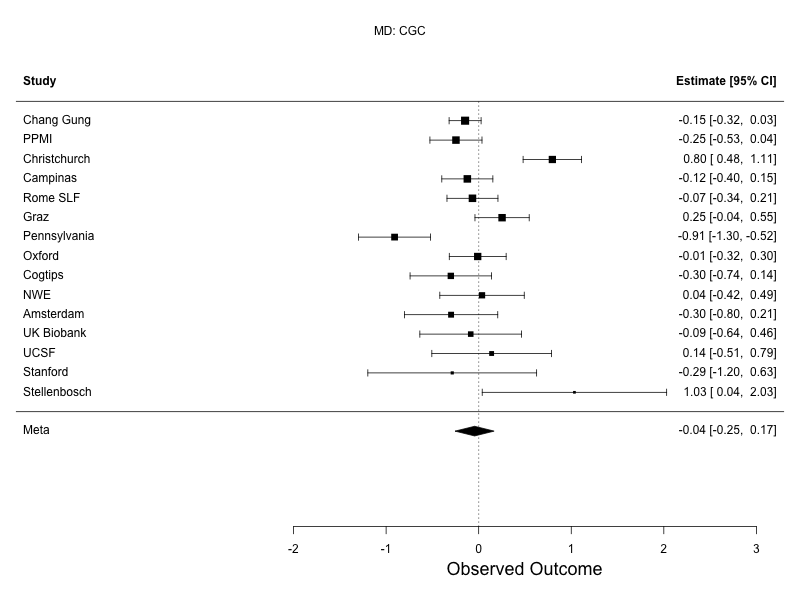 |
| --- | --- |
| 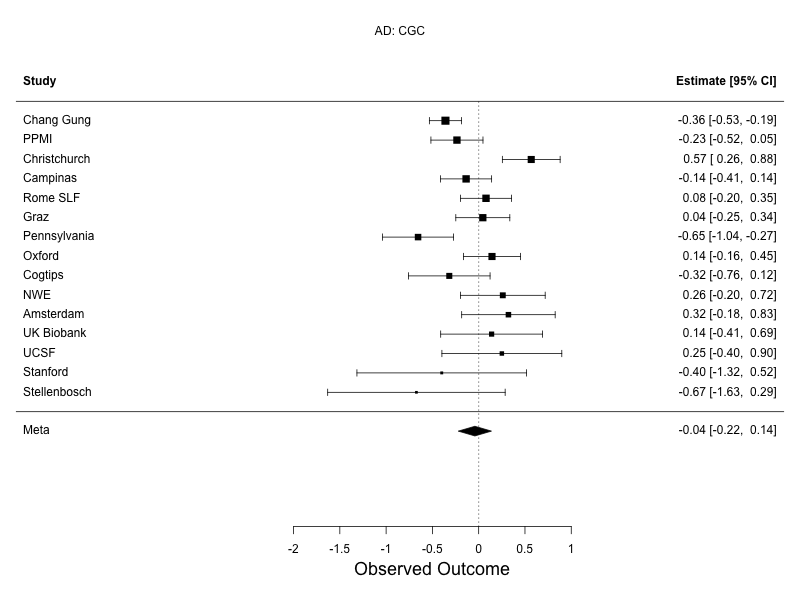 | 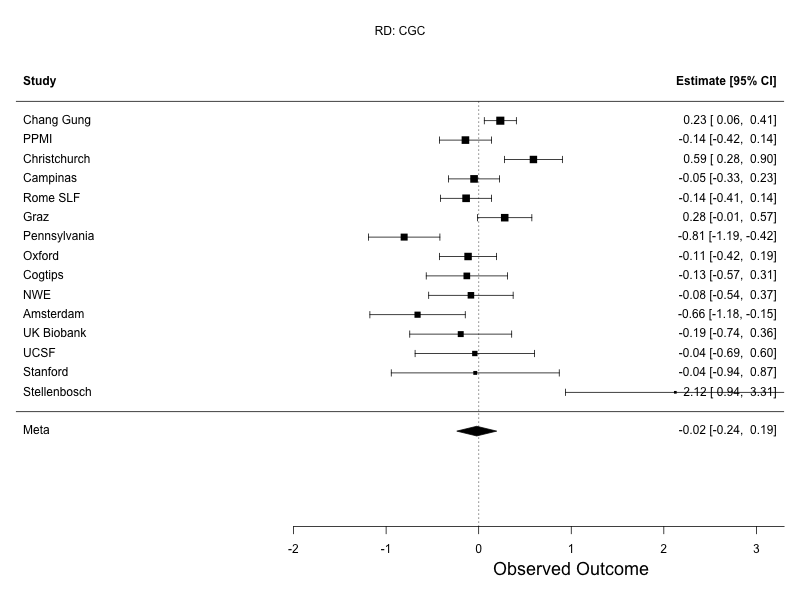 |
| **Supplementary Figure 16: Mixed effects meta analysis forest plots for the CGC.** Differences in DTI measures between PD participants and controls, assessed using random-effects meta-analysis. Sites listed in descending order based on sample size. FA, top left; MD, top right; AD, bottom left; RD, bottom right. | |

| 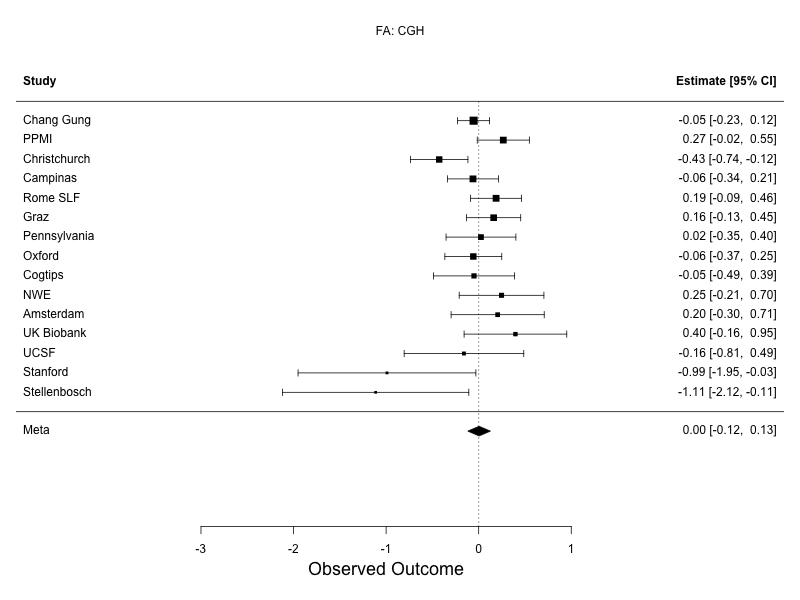 | 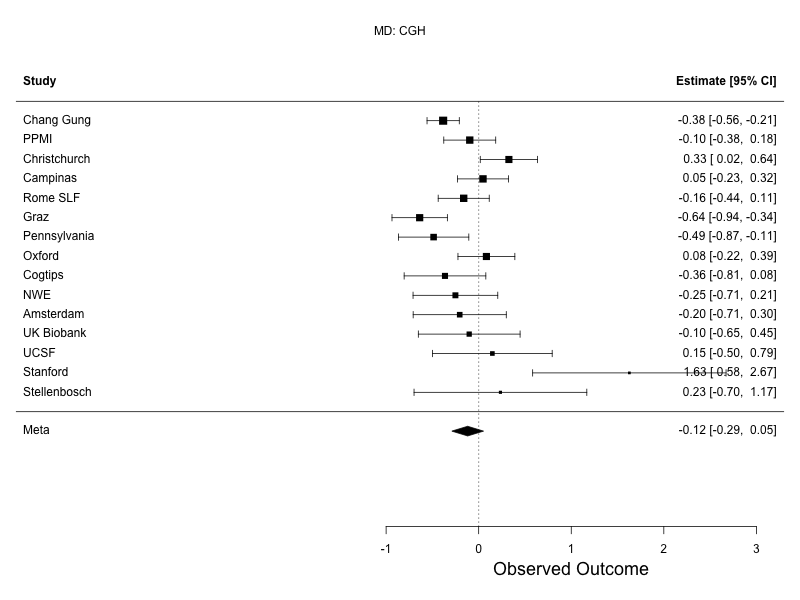 |
| --- | --- |
| 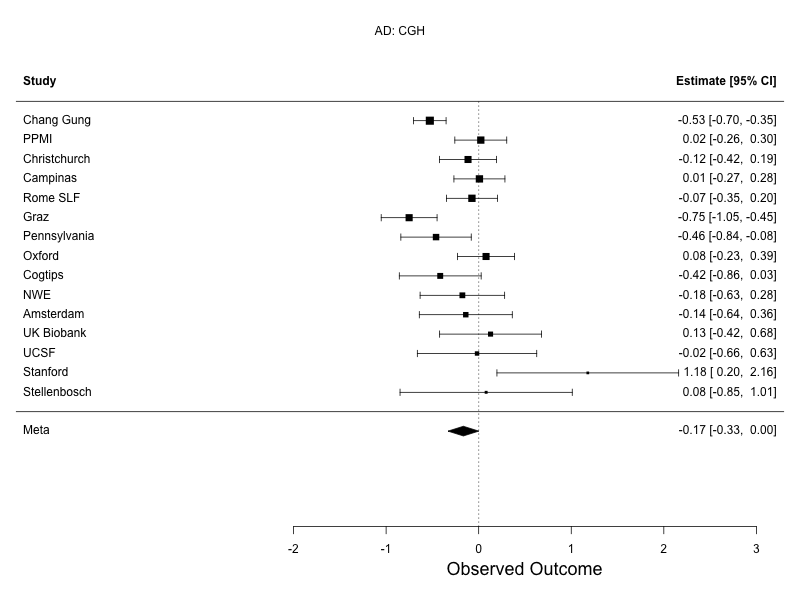 | 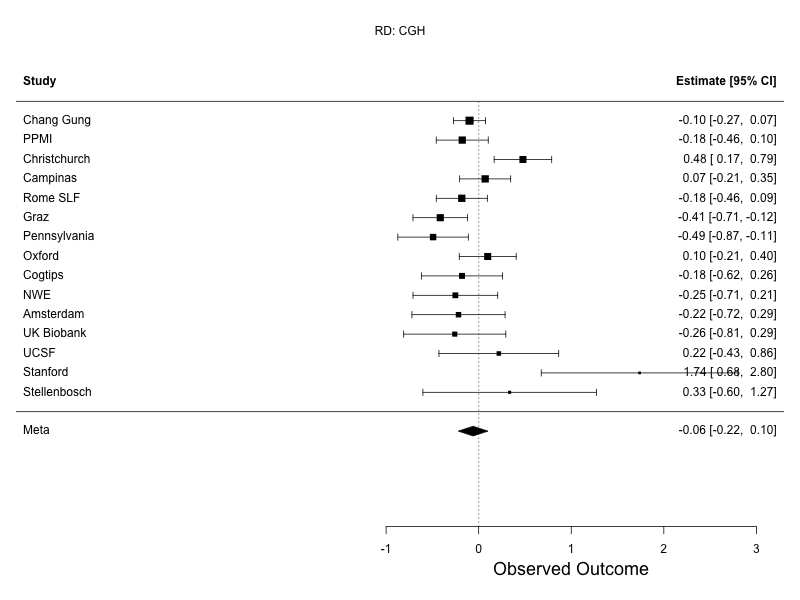 |
| **Supplementary Figure 17: Mixed effects meta analysis forest plots for the CGH.** Differences in DTI measures between PD participants and controls, assessed using random-effects meta-analysis. Sites listed in descending order based on sample size. FA, top left; MD, top right; AD, bottom left; RD, bottom right. | |

| 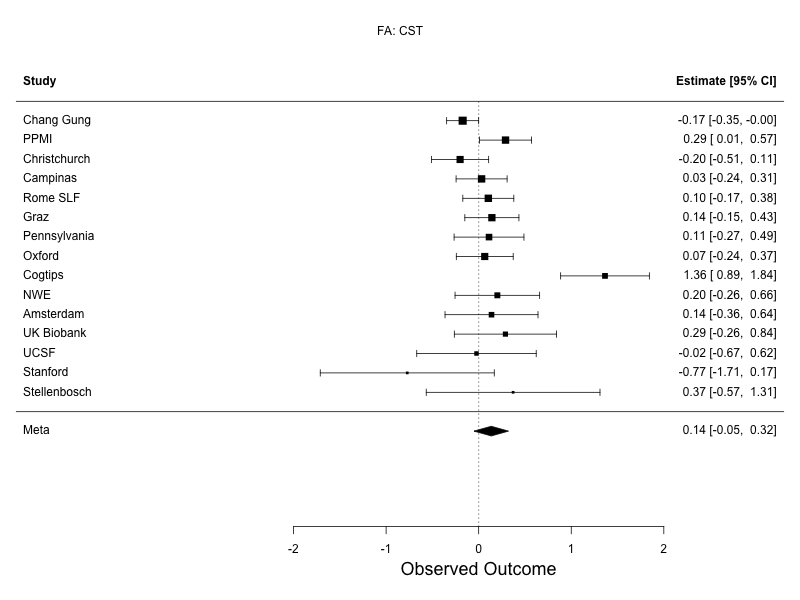 | 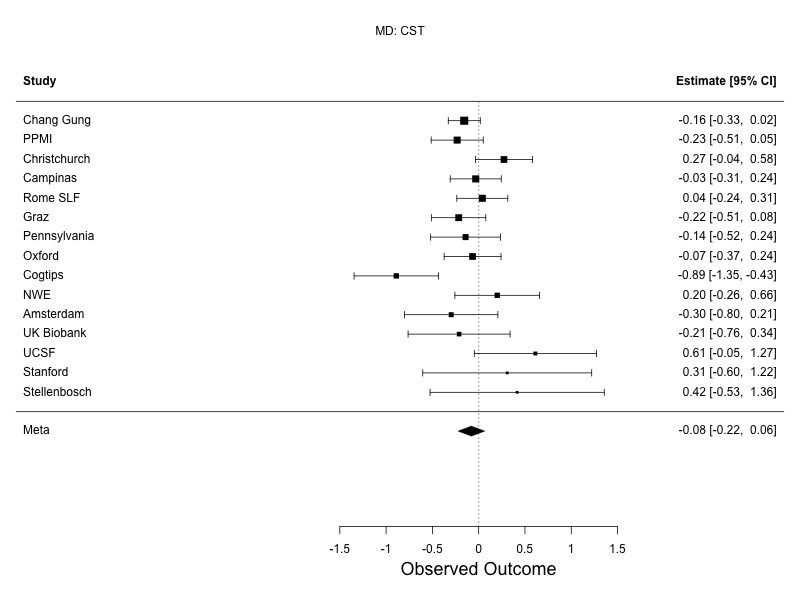 |
| --- | --- |
| 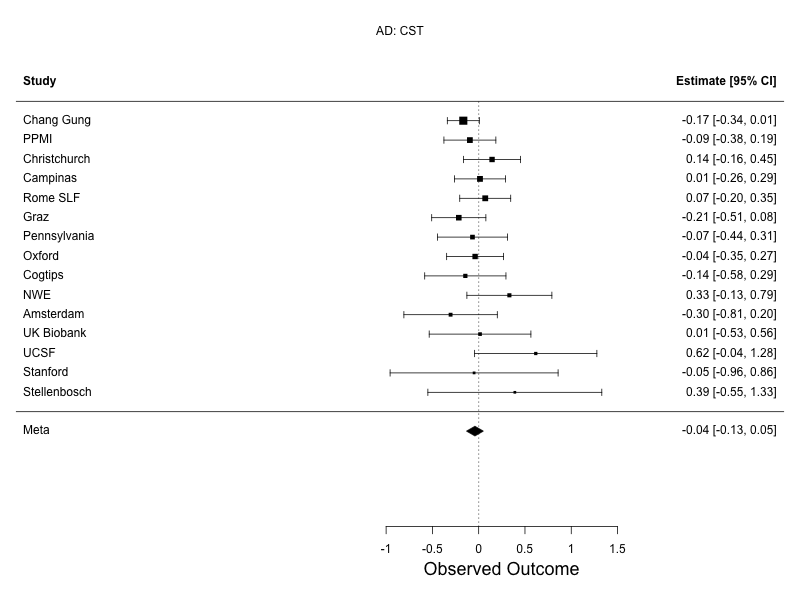 | 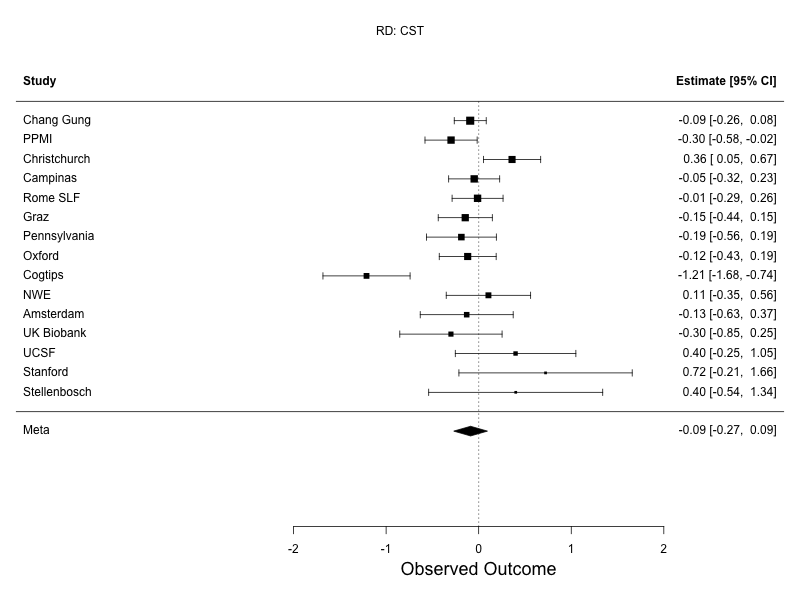 |
| **Supplementary Figure 18: Mixed effects meta analysis forest plots for the CST.** Differences in DTI measures between PD participants and controls, assessed using random-effects meta-analysis. Sites listed in descending order based on sample size. FA, top left; MD, top right; AD, bottom left; RD, bottom right. | |

| 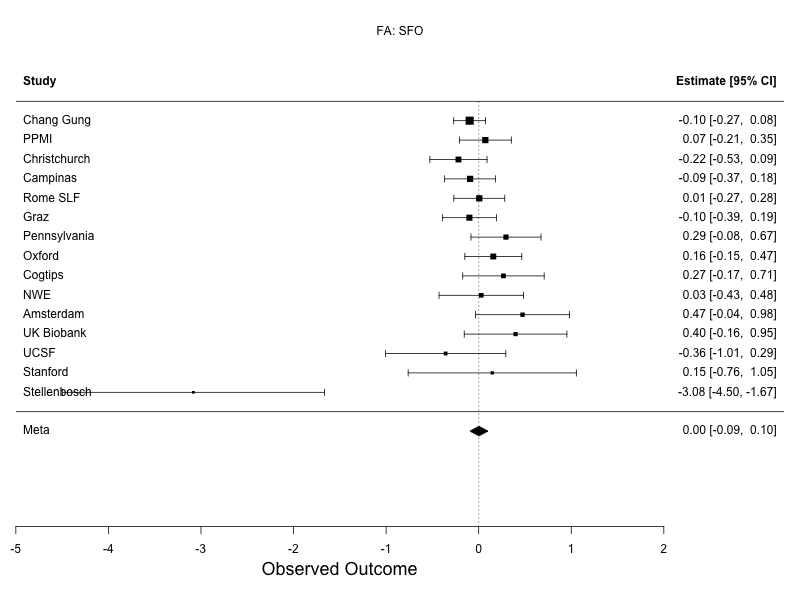 | 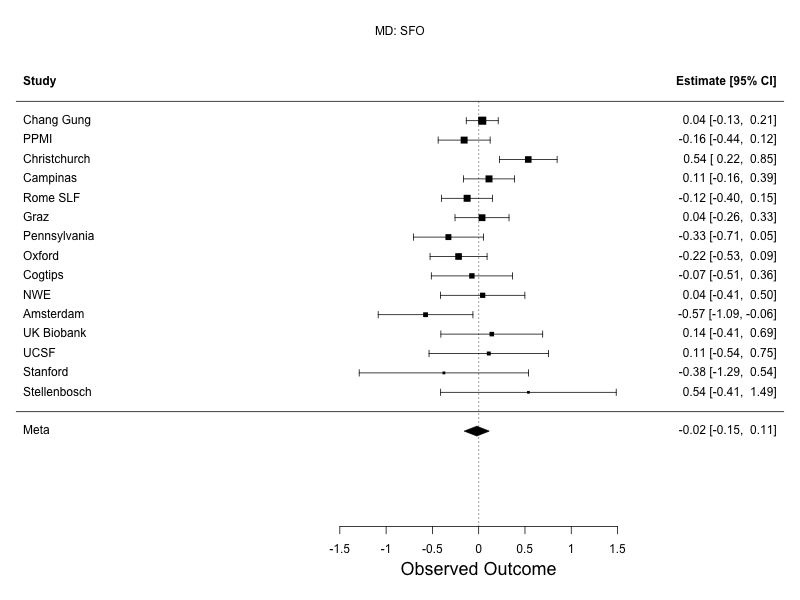 |
| --- | --- |
| 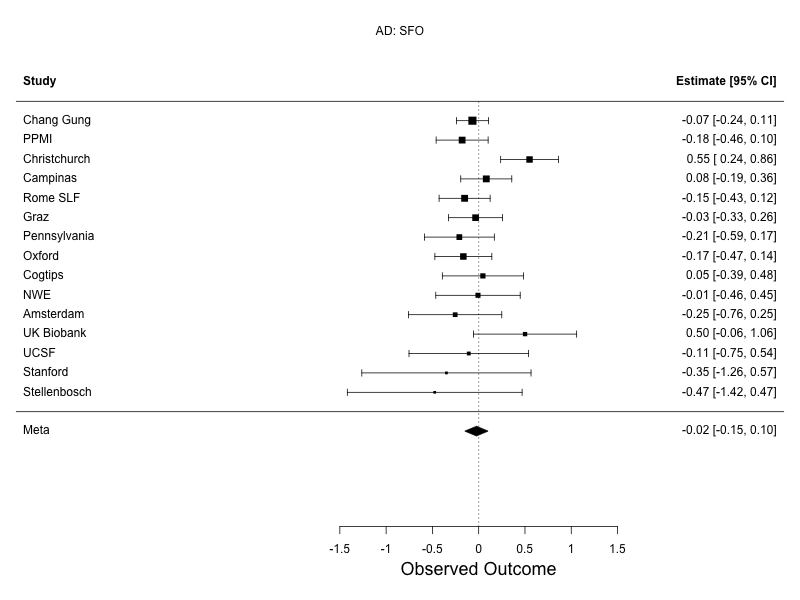 | 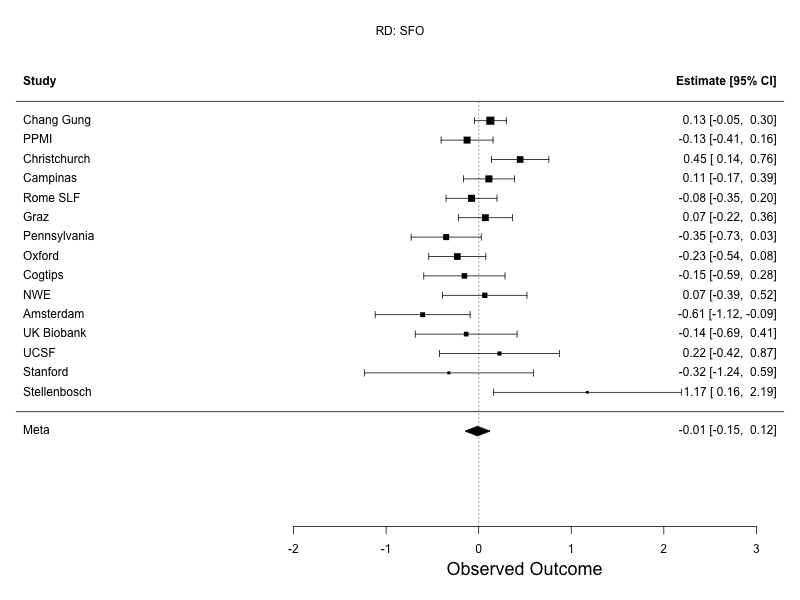 |
| **Supplementary Figure 19: Mixed effects meta analysis forest plots for the SFO.** Differences in DTI measures between PD participants and controls, assessed using random-effects meta-analysis. Sites listed in descending order based on sample size. FA, top left; MD, top right; AD, bottom left; RD, bottom right. | |

| 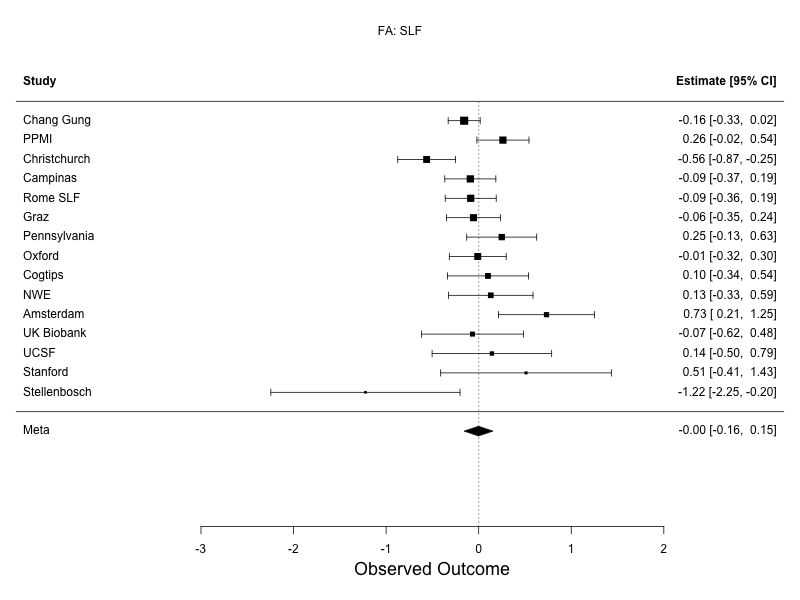 | 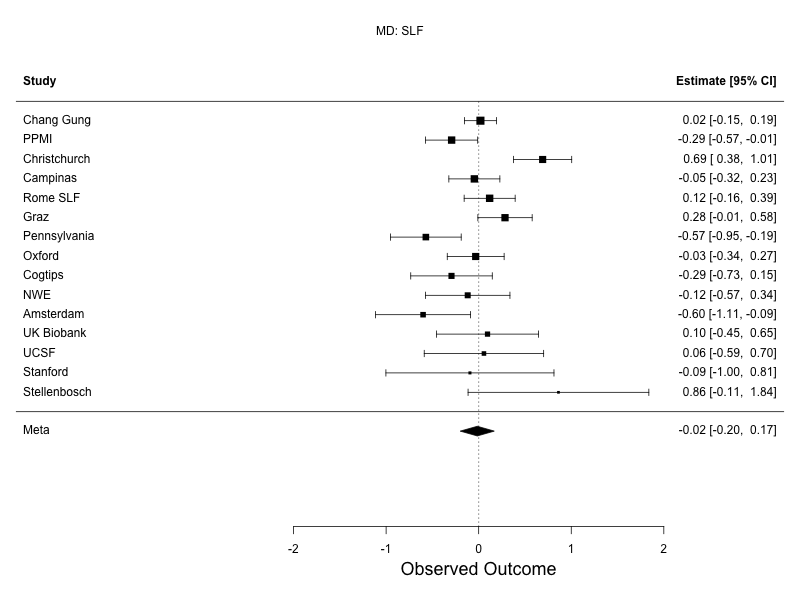 |
| --- | --- |
| 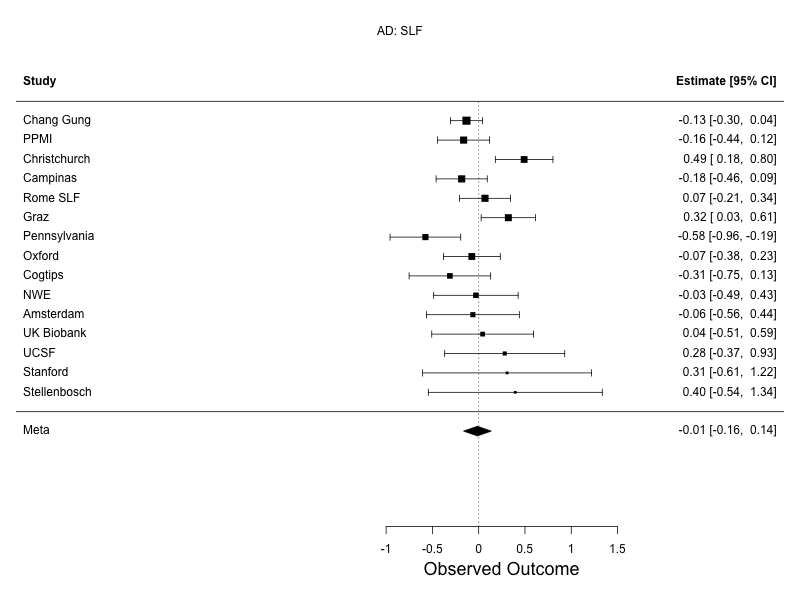 | 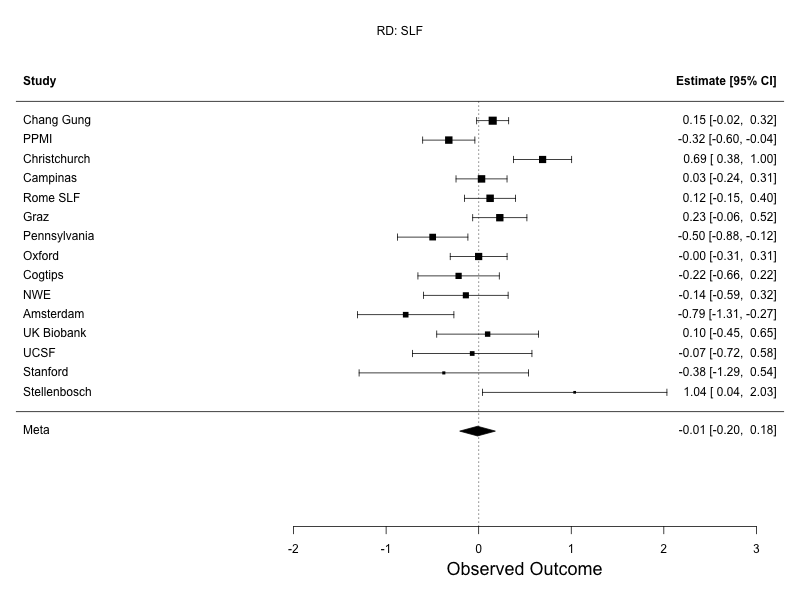 |
| **Supplementary Figure 20: Mixed effects meta analysis forest plots for the SLF.** Differences in DTI measures between PD participants and controls, assessed using random-effects meta-analysis. Sites listed in descending order based on sample size. FA, top left; MD, top right; AD, bottom left; RD, bottom right. | |

| 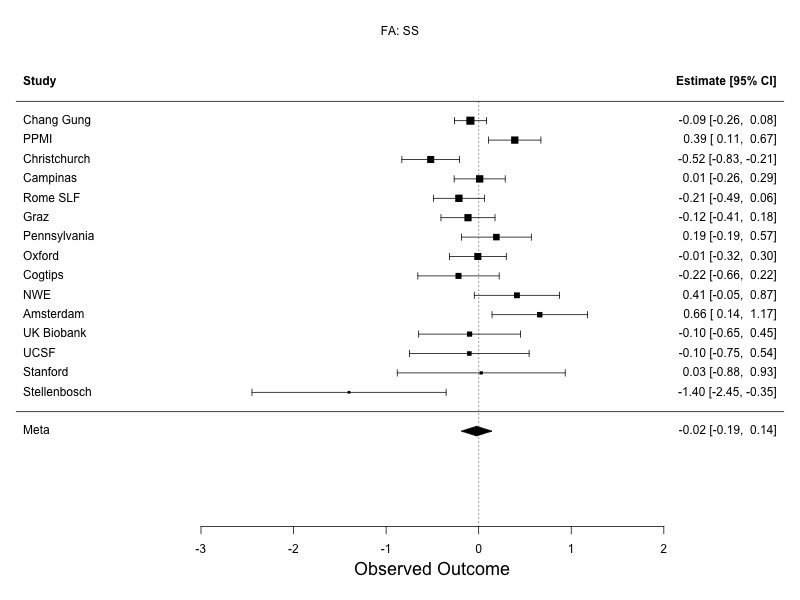 | 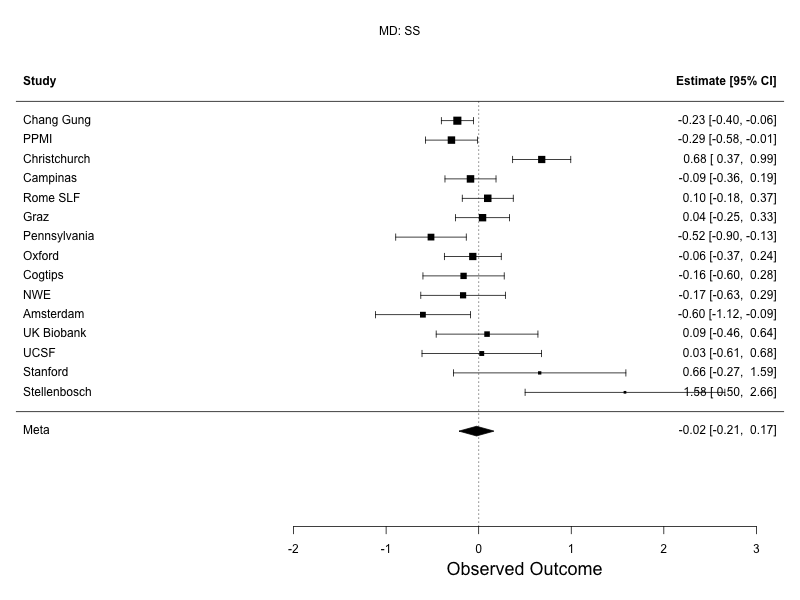 |
| --- | --- |
| 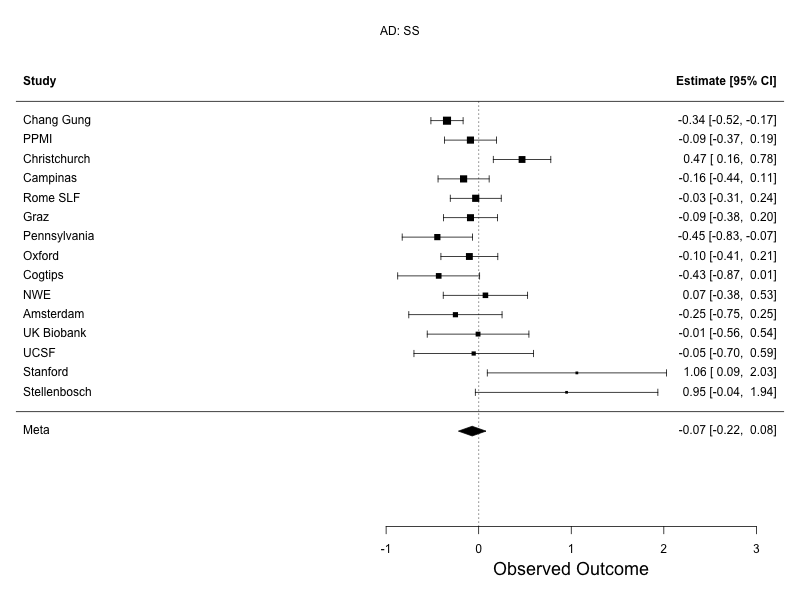 | 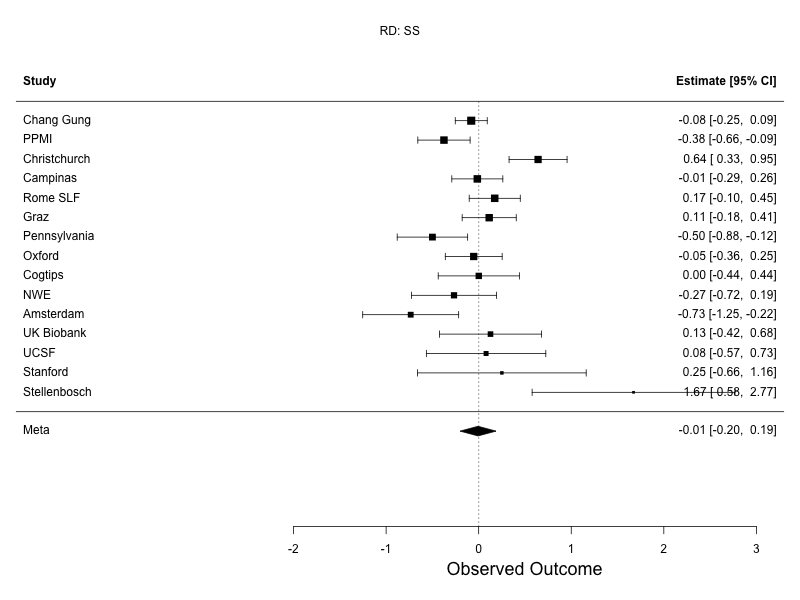 |
| **Supplementary Figure 21: Mixed effects meta analysis forest plots for the SS.** Differences in DTI measures between PD participants and controls, assessed using random-effects meta-analysis. Sites listed in descending order based on sample size. FA, top left; MD, top right; AD, bottom left; RD, bottom right. | |

| 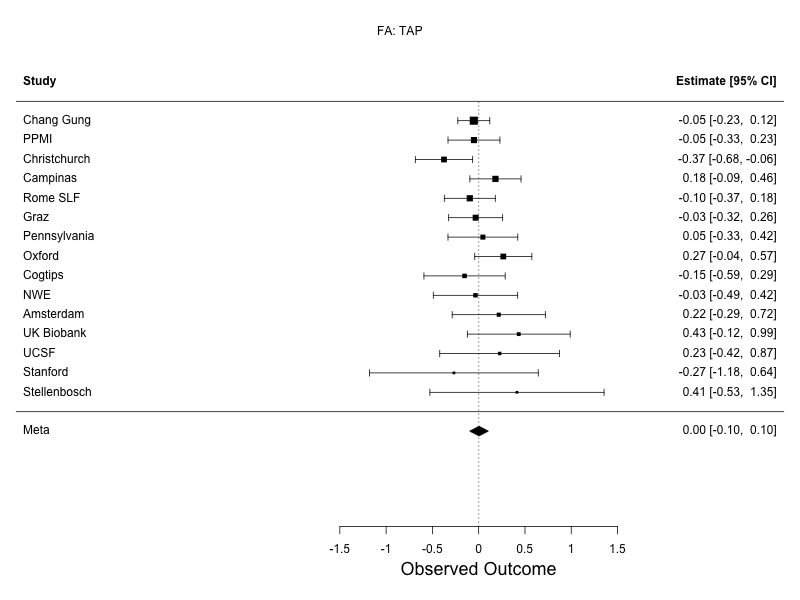 | 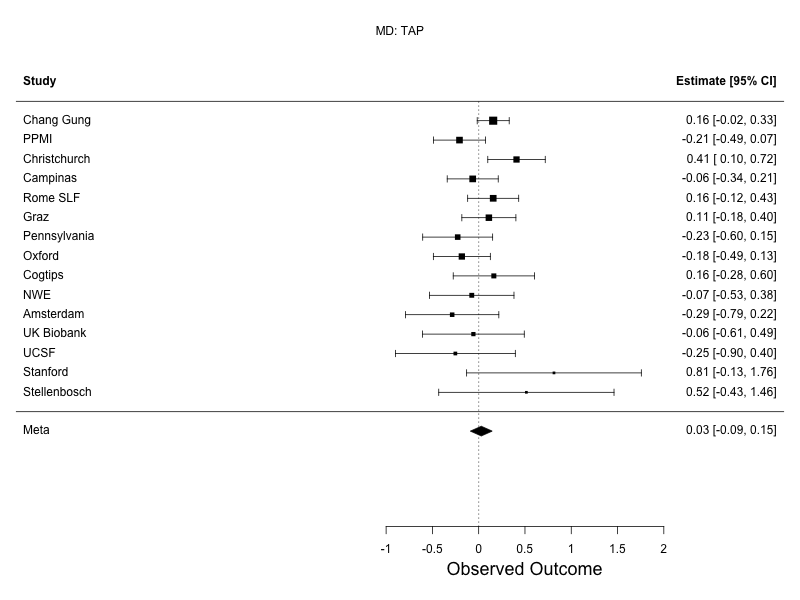 |
| --- | --- |
| 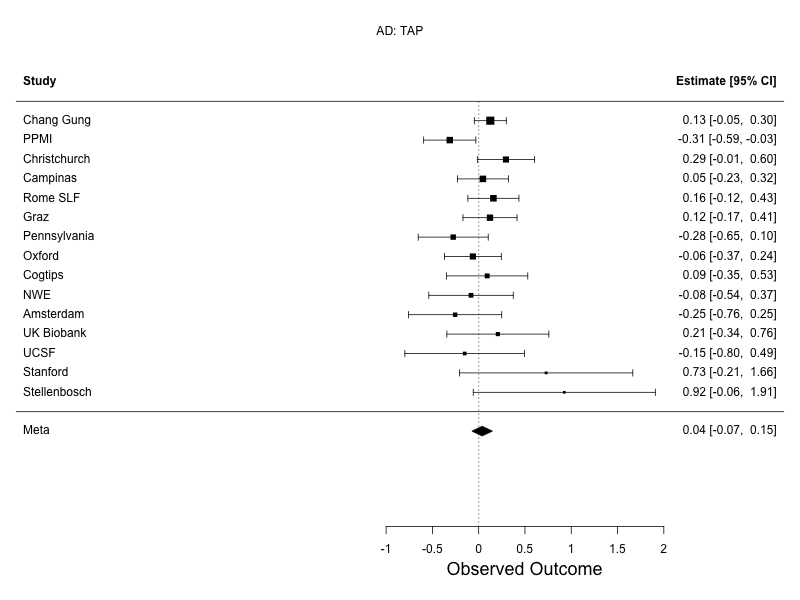 | 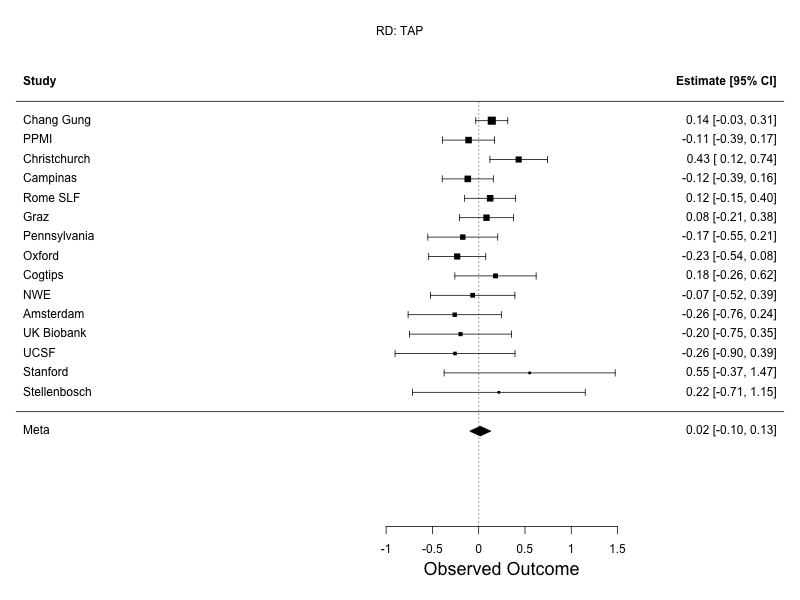 |
| **Supplementary Figure 22: Mixed effects meta analysis forest plots for the TAP.** Differences in DTI measures between PD participants and controls, assessed using random-effects meta-analysis. Sites listed in descending order based on sample size. FA, top left; MD, top right; AD, bottom left; RD, bottom right. | |

| 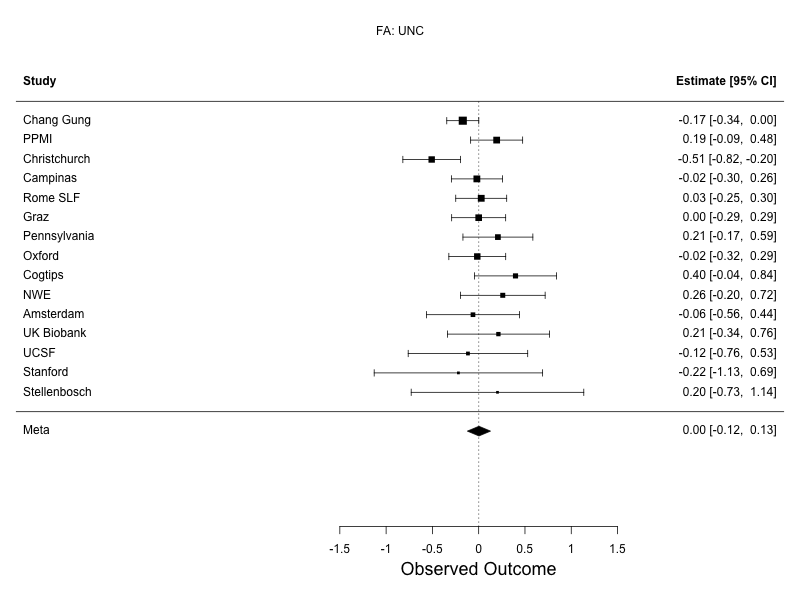 | 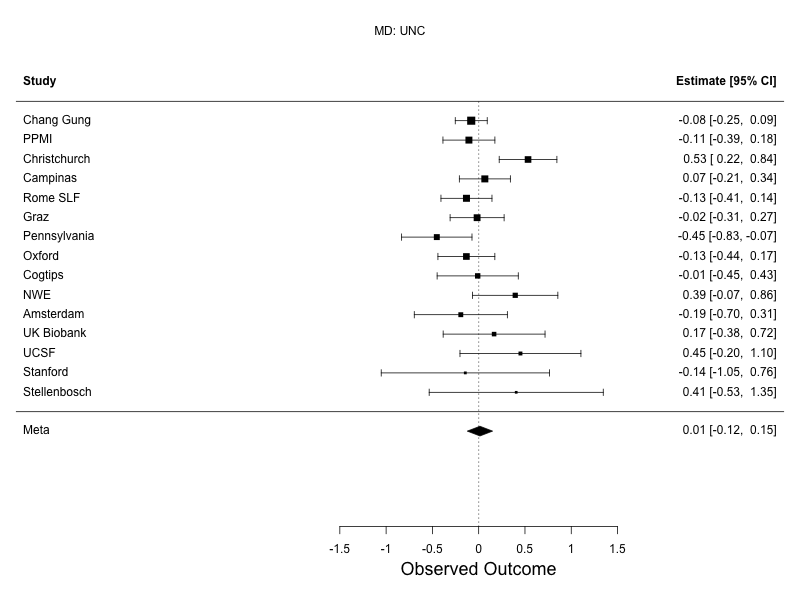 |
| --- | --- |
| 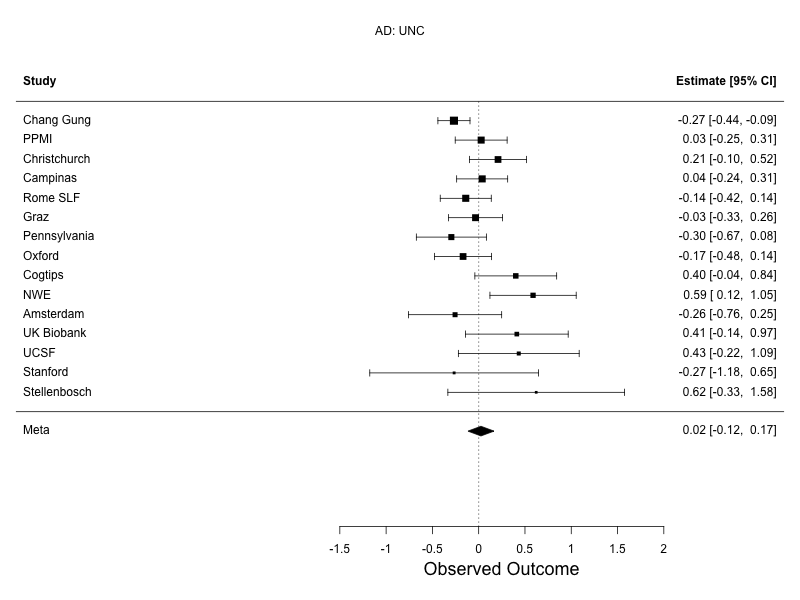 | 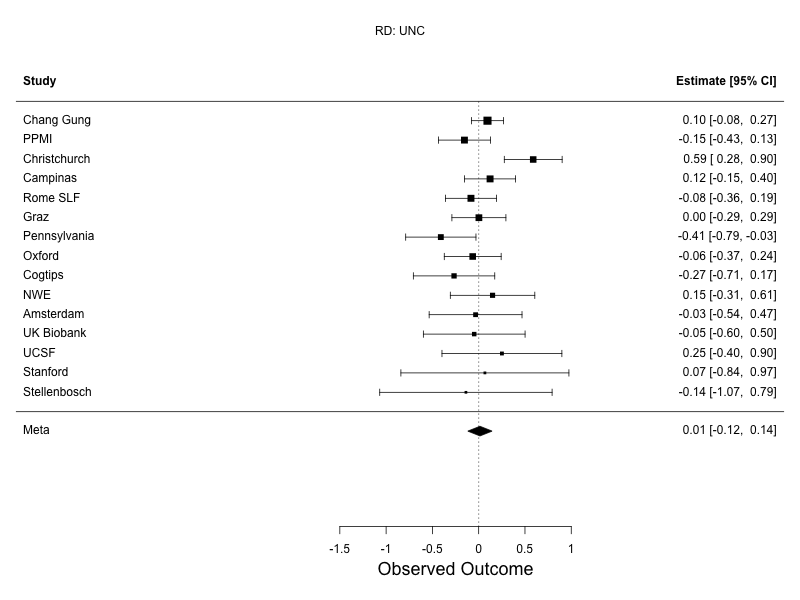 |
| **Supplementary Figure 23: Mixed effects meta analysis forest plots for the UNC.** Differences in DTI measures between PD participants and controls, assessed using random-effects meta-analysis. Sites listed in descending order based on sample size. FA, top left; MD, top right; AD, bottom left; RD, bottom right. | |

#### 2.4 Correlation between Effect Sizes

| 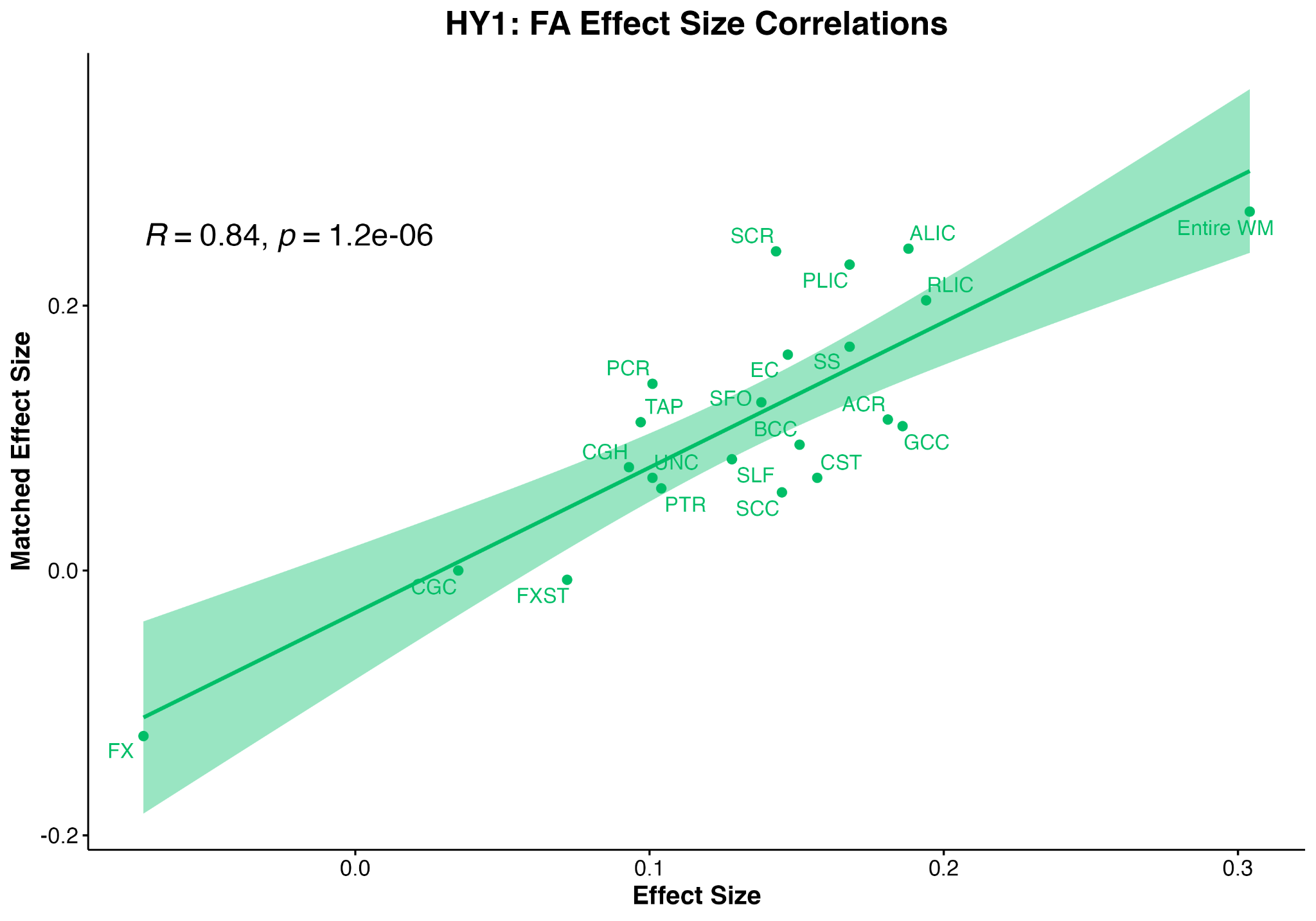 | 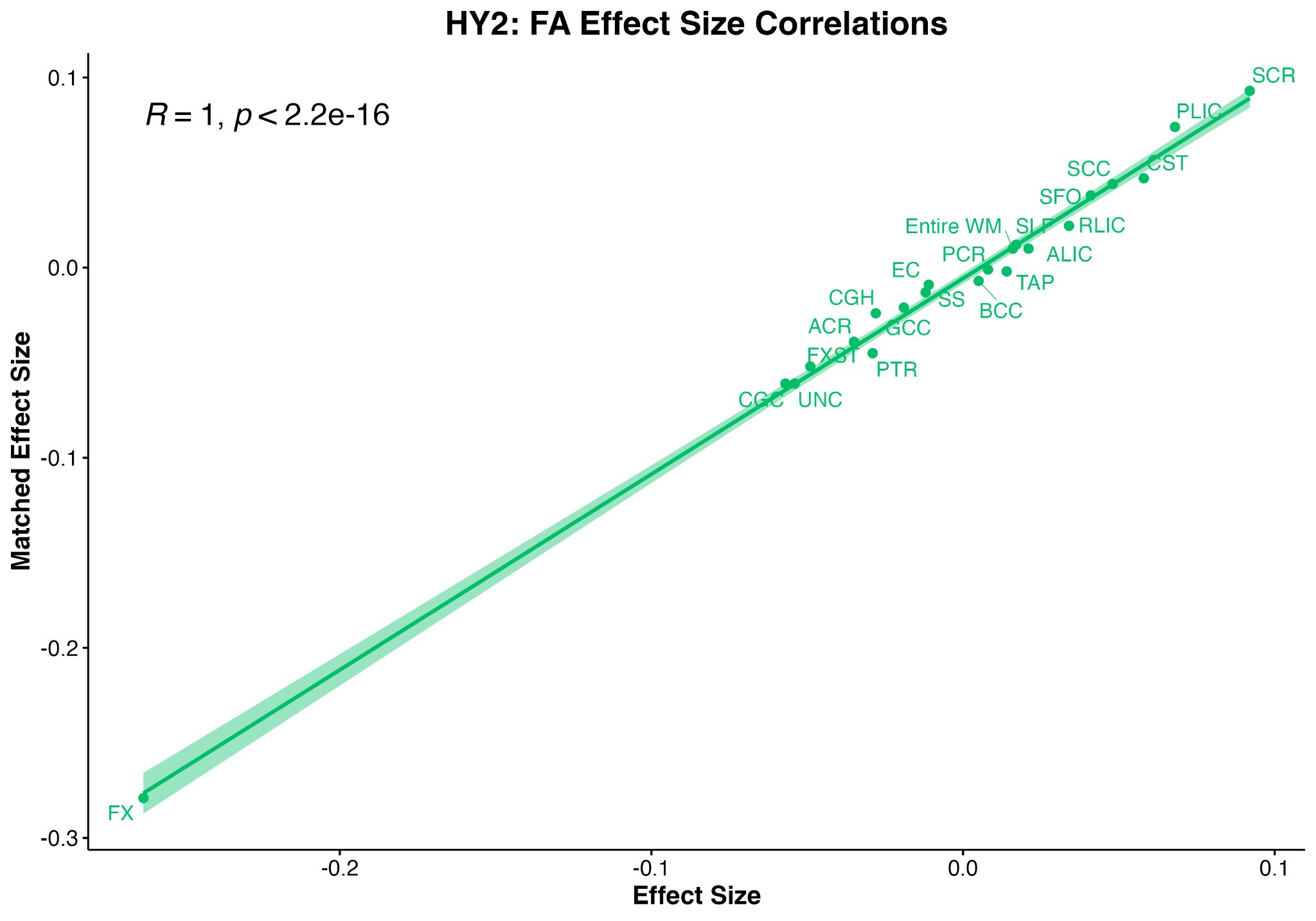 |
| --- | --- |
| 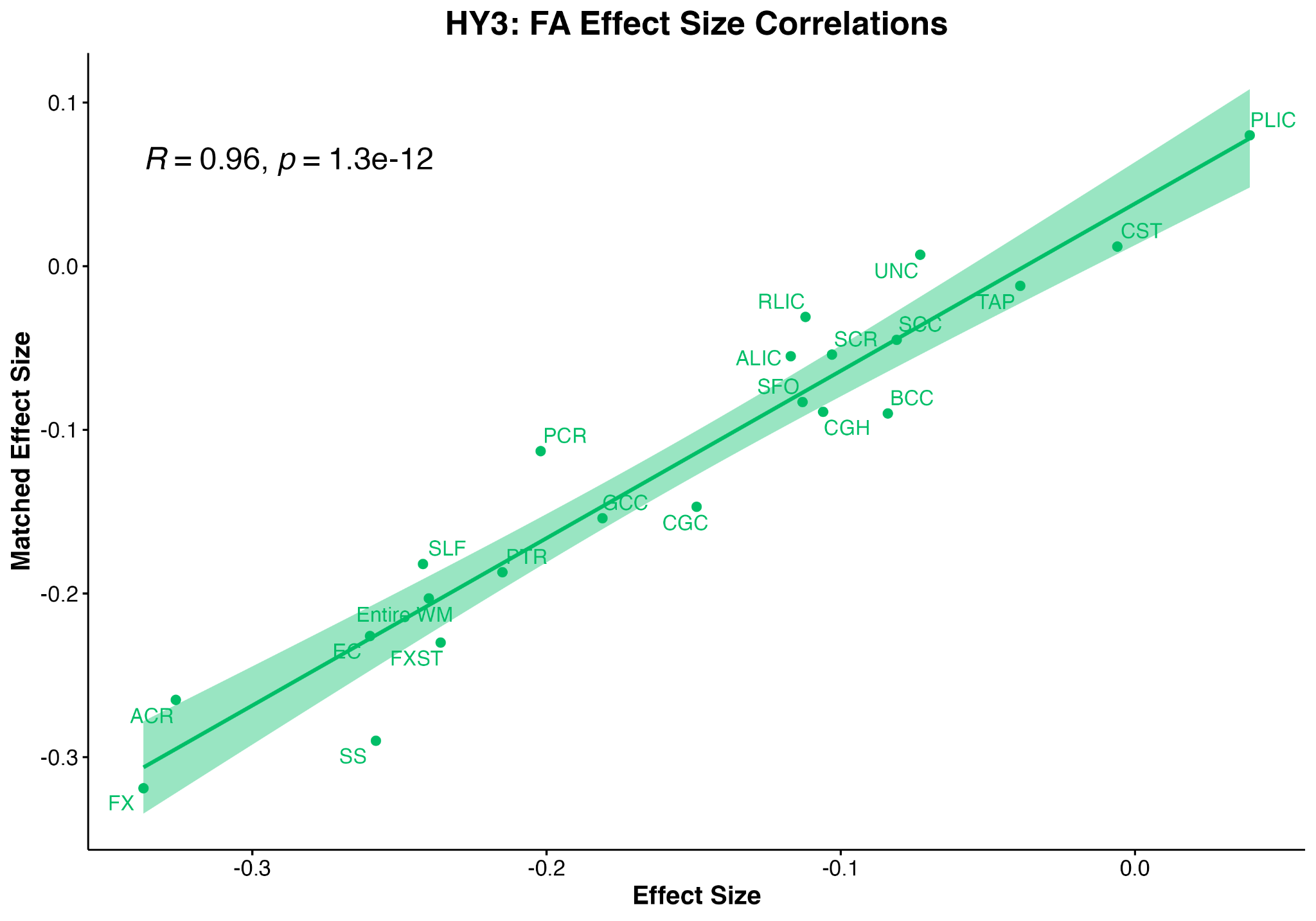 | 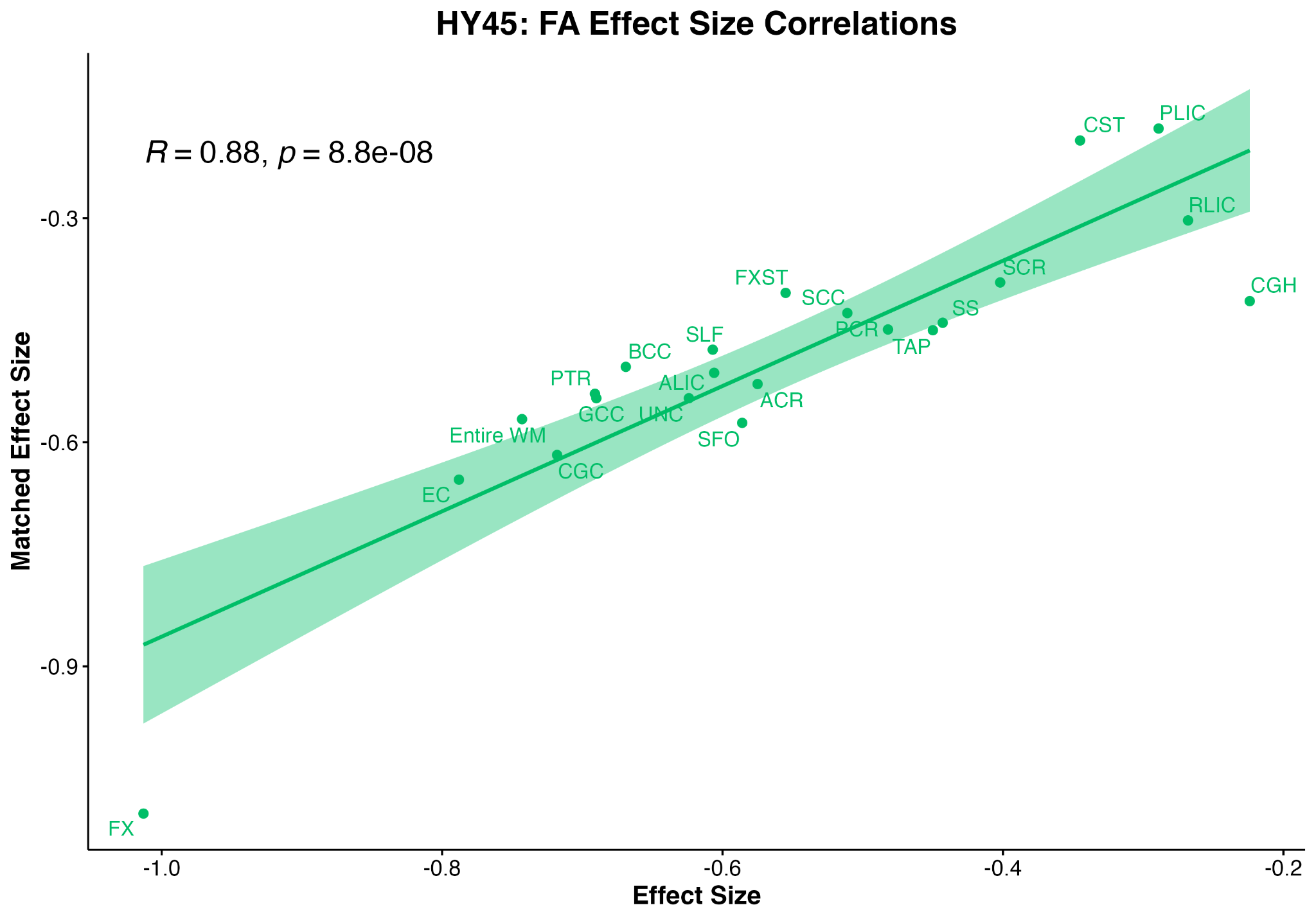 |
| **Supplementary Figure 24:**  **Correlations between effect sizes generated when comparing FA between PD HY subgroups and Controls versus comparing data to matched Control participants.** **Abbreviations**: CR/ACR/PCR/SCR, corona radiata (anterior/posterior/superior); IC/ALIC/PLIC/RLIC, internal capsule (anterior/posterior/retrolenticular limb); EC, external capsule; PTR, posterior thalamic radiation; CC/BCC/GCC/SCC, corpus callosum (body/genu/splenium); CGC, cingulum (cingulate gyrus part); CGH, cingulum (hippocampal portion); CST, corticospinal tract; SFO, superior fronto-occipital fasciculus; SLF, superior longitudinal fasciculus; SS, sagittal stratum; TAP, tapetum; UNC, uncinate fasciculus; FXST, fornix/*stria terminalis.* | |

| 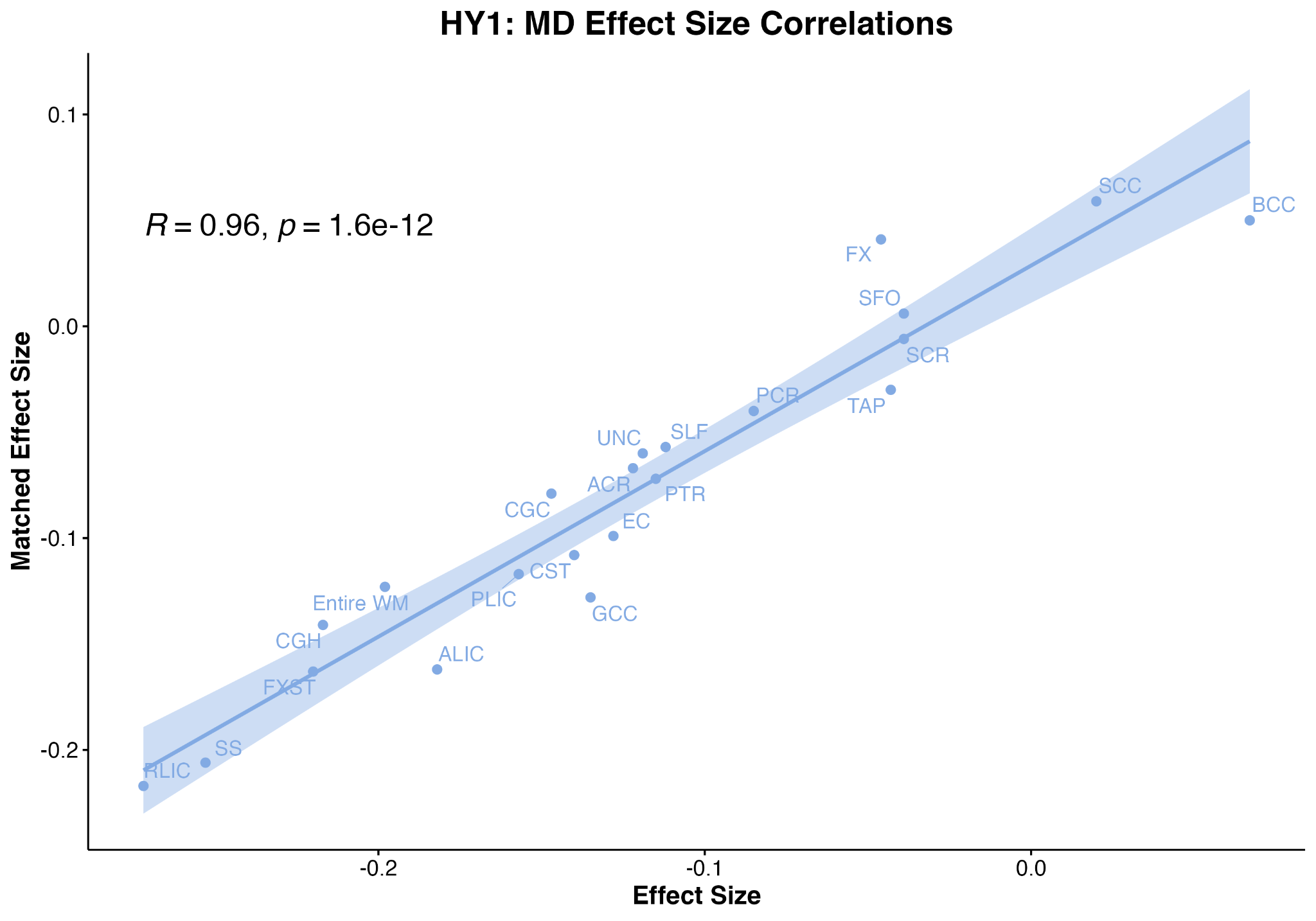 | 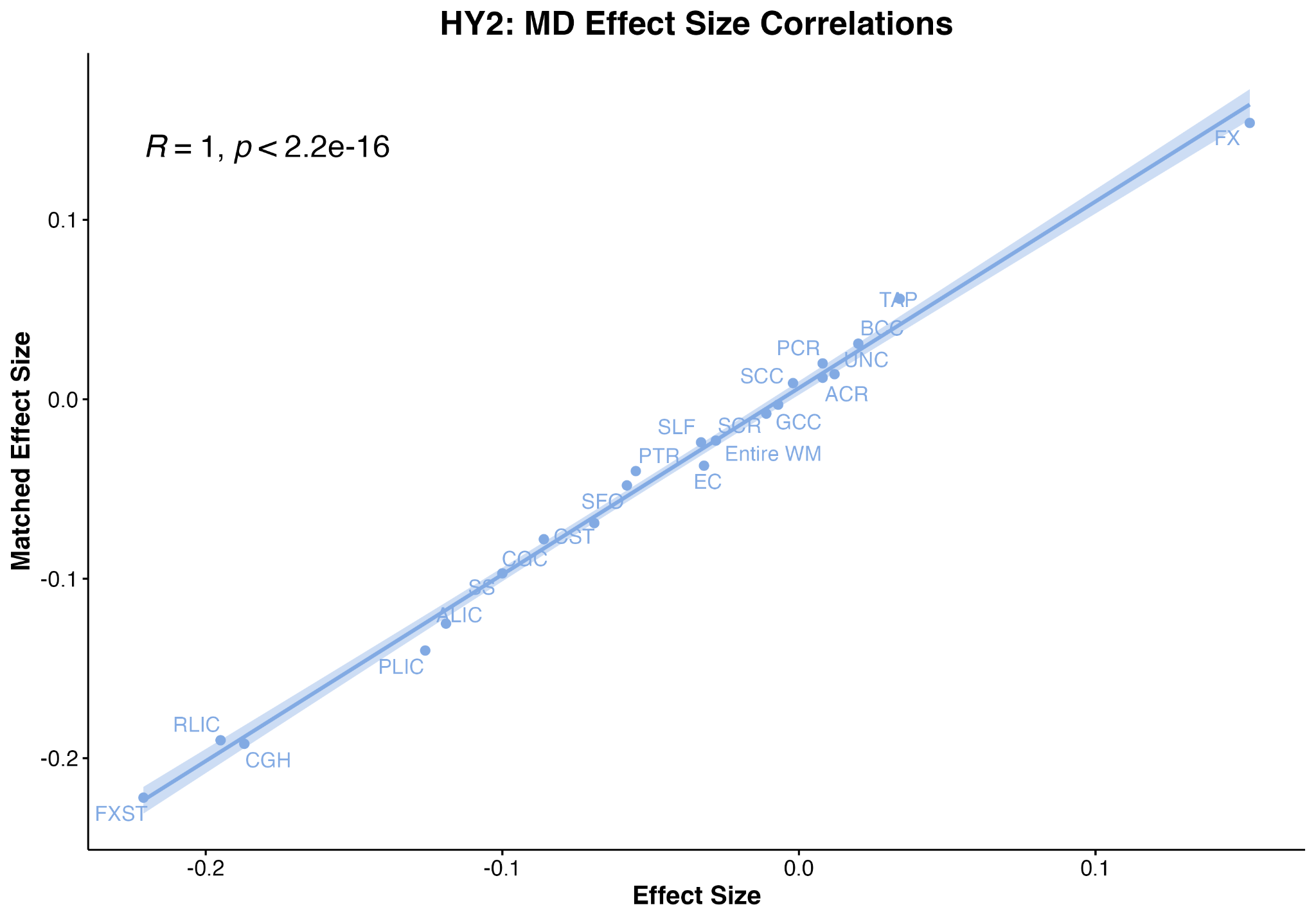 |
| --- | --- |
| 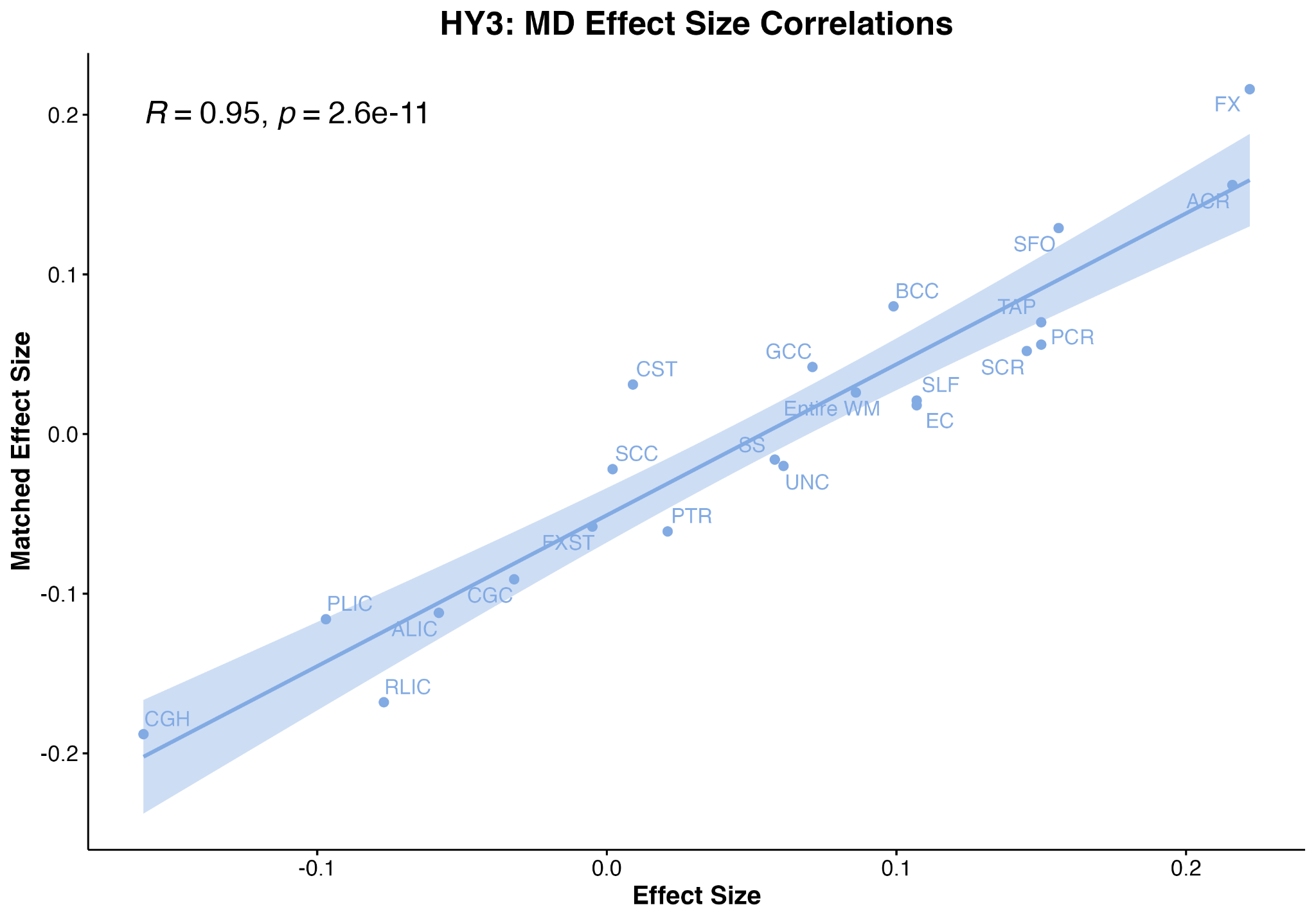 | 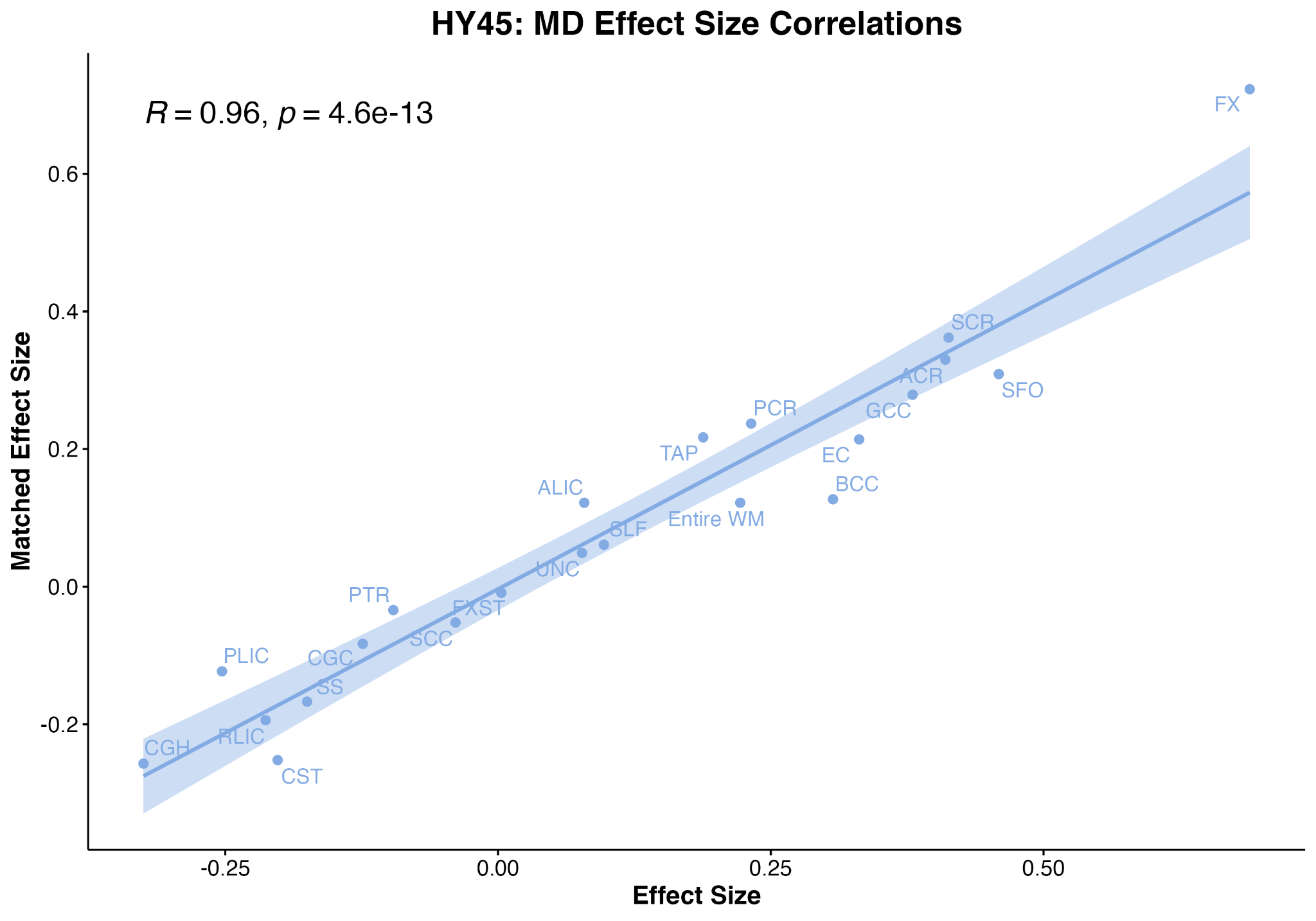 |
| **Supplementary Figure 25: Correlations between effect sizes generated when comparing MD between PD HY subgroups and Controls versus comparing data to matched Control participants.** **Abbreviations**: CR/ACR/PCR/SCR, corona radiata (anterior/posterior/superior); IC/ALIC/PLIC/RLIC, internal capsule (anterior/posterior/retrolenticular limb); EC, external capsule; PTR, posterior thalamic radiation; CC/BCC/GCC/SCC, corpus callosum (body/genu/splenium); CGC, cingulum (cingulate gyrus part); CGH, cingulum (hippocampal portion); CST, corticospinal tract; SFO, superior fronto-occipital fasciculus; SLF, superior longitudinal fasciculus; SS, sagittal stratum; TAP, tapetum; UNC, uncinate fasciculus; FXST, fornix/stria-terminalis; | |

| 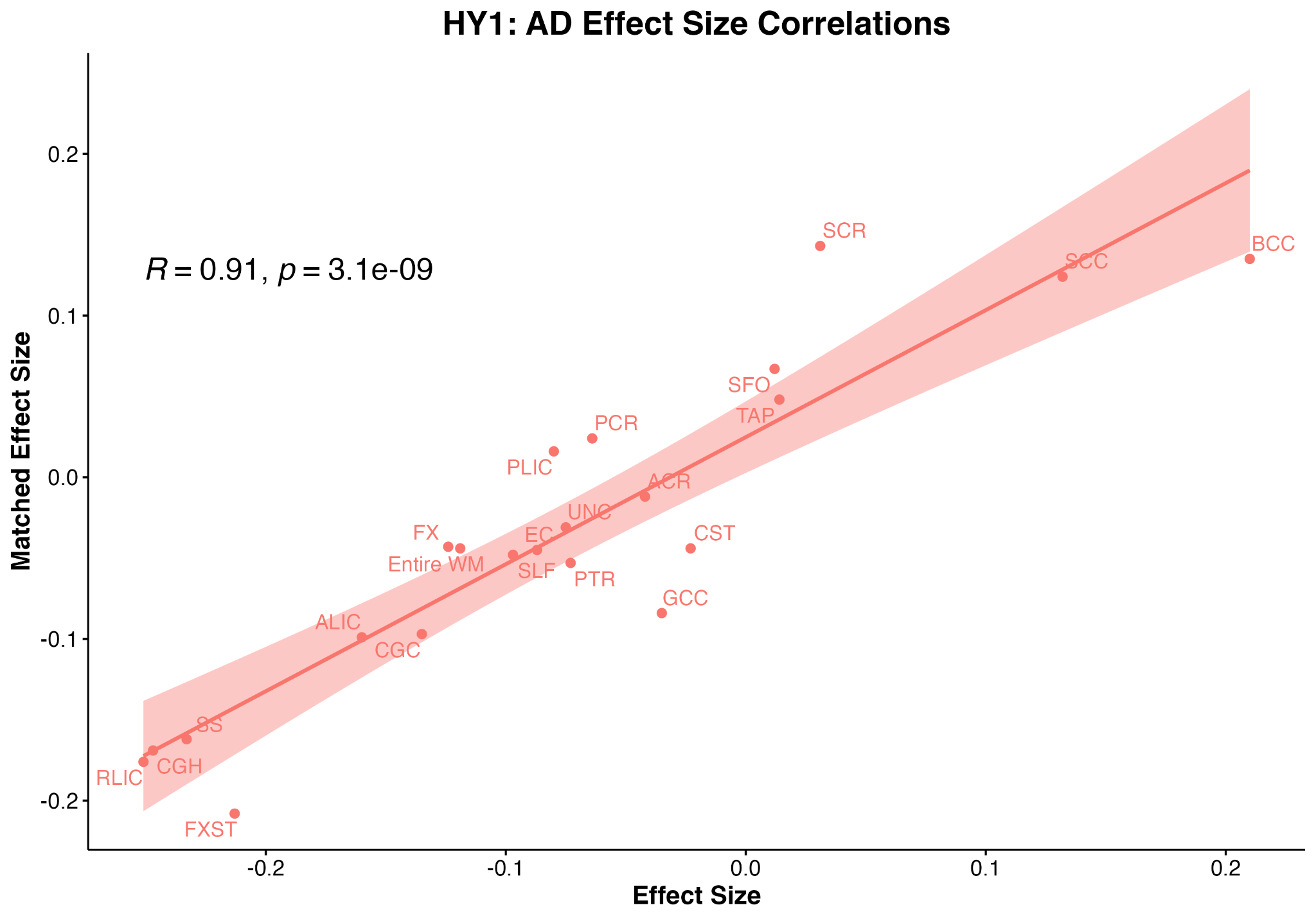 | 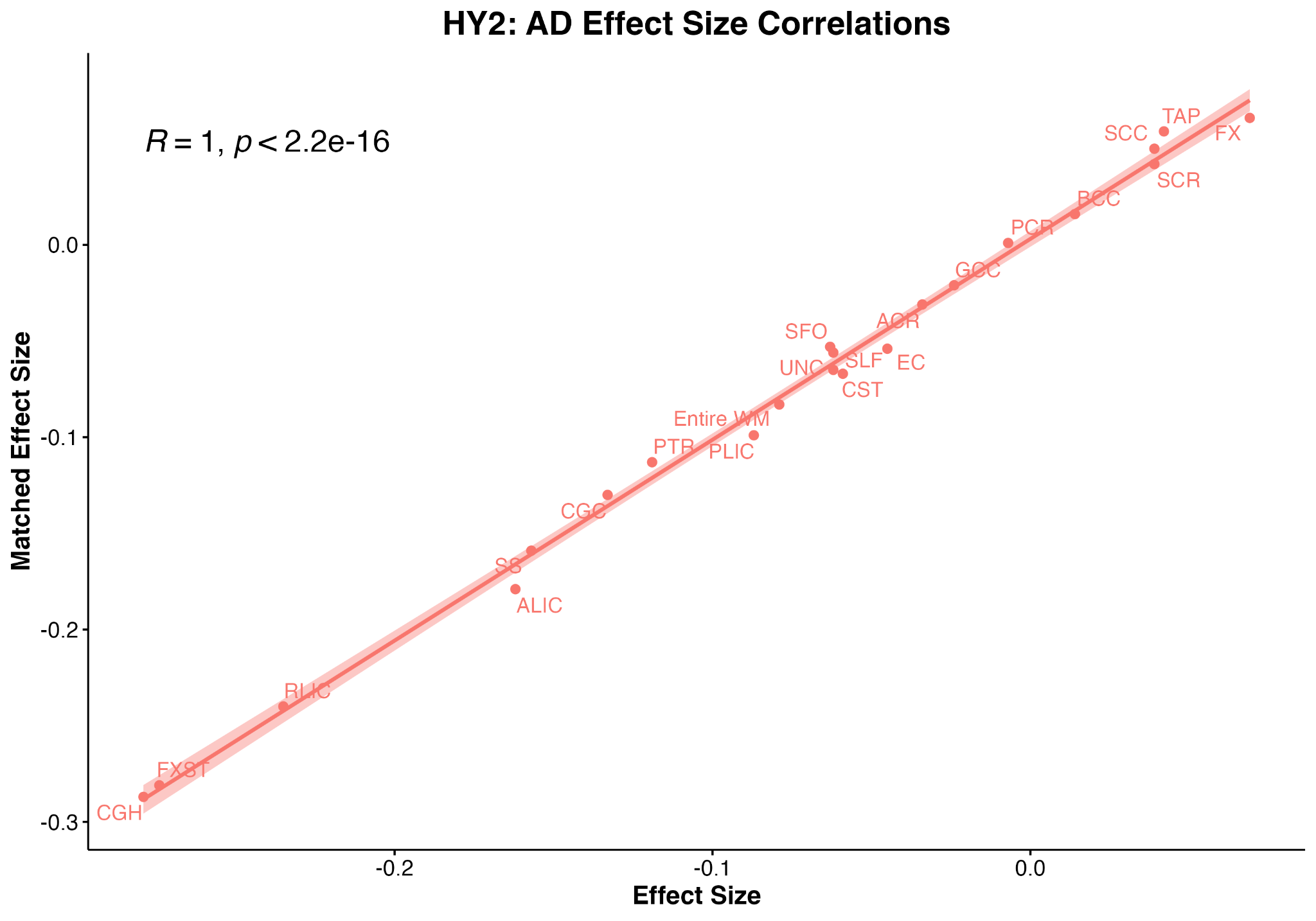 |
| --- | --- |
| 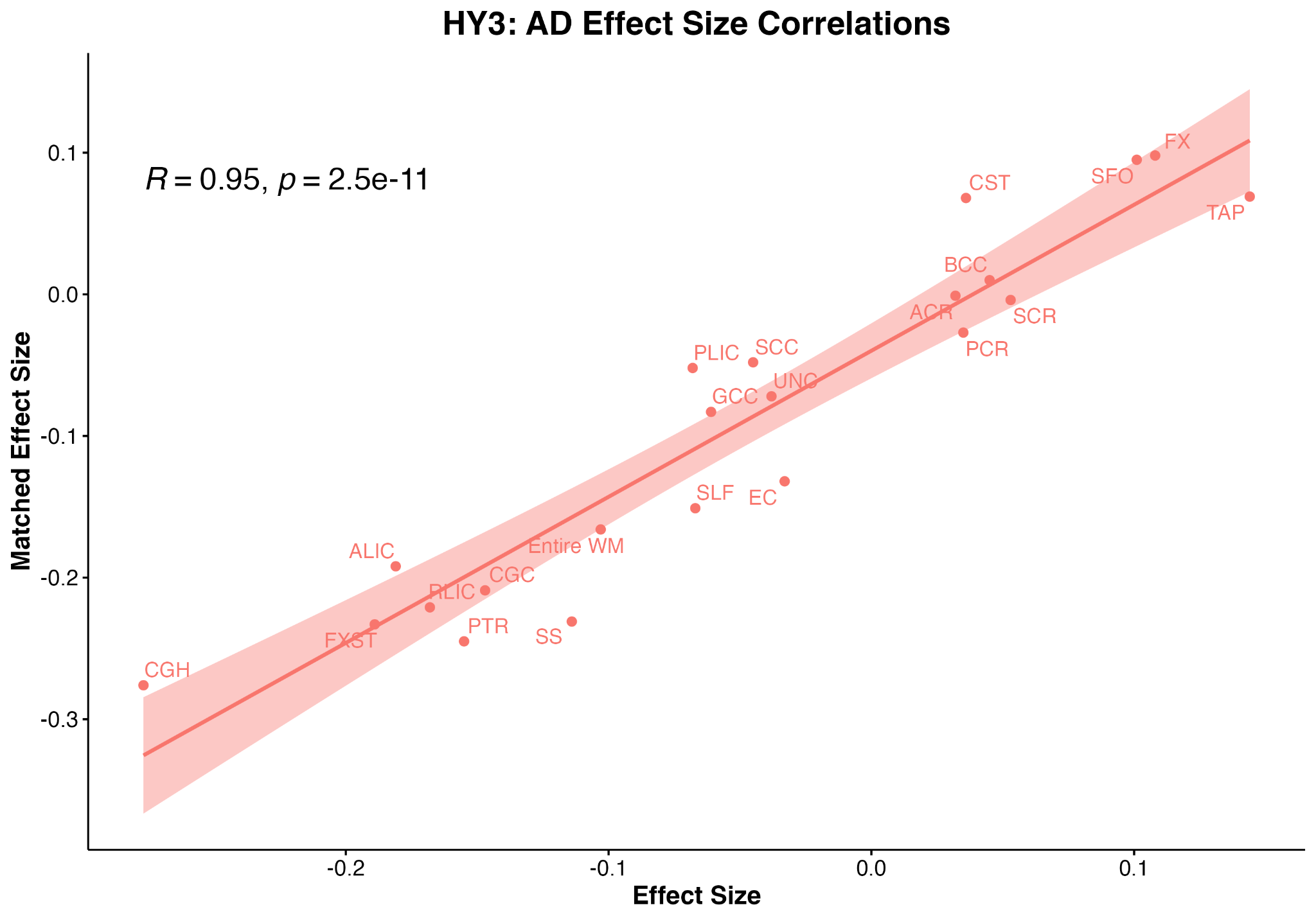 |  |
| **Supplementary Figure 26: Correlations between effect sizes generated when comparing AD between PD HY subgroups and Controls versus comparing data to matched Control groups. Abbreviations**: CR/ACR/PCR/SCR, corona radiata (anterior/posterior/superior); IC/ALIC/PLIC/RLIC, internal capsule (anterior/posterior/retrolenticular limb); EC, external capsule; PTR, posterior thalamic radiation; CC/BCC/GCC/SCC, corpus callosum (body/genu/splenium); CGC, cingulum (cingulate gyrus part); CGH, cingulum (hippocampal portion); CST, corticospinal tract; SFO, superior fronto-occipital fasciculus; SLF, superior longitudinal fasciculus; SS, sagittal stratum; TAP, tapetum; UNC, uncinate fasciculus; FXST, fornix/stria-terminalis; | |

|  |
| --- |
| **Supplementary Figure 27: Correlations between effect sizes generated when comparing RD between PD HY subgroups and Controls versus comparing data to matched Control groups. Abbreviations**: CR/ACR/PCR/SCR, corona radiata (anterior/posterior/superior); IC/ALIC/PLIC/RLIC, internal capsule (anterior/posterior/retrolenticular limb); EC, external capsule; PTR, posterior thalamic radiation; CC/BCC/GCC/SCC, corpus callosum (body/genu/splenium); CGC, cingulum (cingulate gyrus part); CGH, cingulum (hippocampal portion); CST, corticospinal tract; SFO, superior fronto-occipital fasciculus; SLF, superior longitudinal fasciculus; SS, sagittal stratum; TAP, tapetum; UNC, uncinate fasciculus; FXST, fornix/stria-terminalis; |

#### 2.5 Partial Correlations: Within-group investigations between DTI metrics and clinical variables

##### 2.5.1 PD participants: Partial correlation between DTI metrics and Montreal Cognitive Assessment scores

| **Total PD & Controls** | ***p*-value** | **Degrees of Freedom** | **Pearson’s Correlation Coefficient *r*** | **Standard Error** | **Lower CI** | **Upper CI** | ***p*_FDR_** |
| --- | --- | --- | --- | --- | --- | --- | --- |
| **Entire WM FA** | 0.00 | 890 | 0.11 | 0.03 | 0.042 | 0.171 | 1.57 𝗑 10^2^ |
| **ACR FA** | 0.04 | 890 | 0.07 | 0.03 | 0.003 | 0.133 | 8.21 𝗑 10^2^ |
| **SCR FA** | 0.90 | 890 | 0.00 | 0.03 | -0.061 | 0.069 | 9.47 𝗑 10^1^ |
| **PCR FA** | 0.63 | 890 | 0.02 | 0.03 | -0.049 | 0.081 | 7.31 𝗑 10^1^ |
| **ALIC FA** | 0.01 | 890 | 0.09 | 0.03 | 0.028 | 0.158 | 2.49 𝗑 10^2^ |
| **PLIC FA** | 0.52 | 890 | 0.02 | 0.03 | -0.044 | 0.087 | 6.40 𝗑 10^1^ |
| **RLIC FA** | 0.36 | 890 | 0.03 | 0.03 | -0.034 | 0.096 | 4.60 𝗑 10^1^ |
| **EC FA** | 0.00 | 890 | 0.10 | 0.03 | 0.038 | 0.167 | 1.58 𝗑 10^2^ |
| **FX FA** | 0.00 | 890 | 0.13 | 0.03 | 0.062 | 0.192 | 3.13 𝗑 10^3^ |
| **FXST FA** | 0.07 | 890 | 0.06 | 0.03 | -0.004 | 0.126 | 1.08 𝗑 10^1^ |
| **PTR FA** | 0.01 | 890 | 0.09 | 0.03 | 0.028 | 0.157 | 2.49 𝗑 10^2^ |
| **GCC FA** | 0.02 | 890 | 0.08 | 0.03 | 0.015 | 0.145 | 5.34 𝗑 10^2^ |
| **BCC FA** | 0.09 | 890 | 0.06 | 0.03 | -0.008 | 0.122 | 1.26 𝗑 10^1^ |
| **SCC FA** | 0.73 | 890 | 0.01 | 0.03 | -0.054 | 0.077 | 8.06 𝗑 10^1^ |
| **CGC FA** | 0.09 | 890 | 0.06 | 0.03 | -0.009 | 0.122 | 1.26 𝗑 10^1^ |
| **CGH FA** | 0.04 | 890 | 0.07 | 0.03 | 0.002 | 0.132 | 8.21 𝗑 10^2^ |
| **CST FA** | 0.04 | 890 | 0.07 | 0.03 | 0.003 | 0.133 | 8.21 𝗑 10^2^ |
| **SFO FA** | 0.07 | 890 | 0.06 | 0.03 | -0.004 | 0.126 | 1.08 𝗑 10^1^ |
| **SLF FA** | 0.03 | 890 | 0.07 | 0.03 | 0.009 | 0.138 | 7.74 𝗑 10^2^ |
| **SS FA** | 0.01 | 890 | 0.09 | 0.03 | 0.026 | 0.155 | 2.49 𝗑 10^2^ |
| **TAP FA** | 1.00 | 890 | 0.00 | 0.03 | -0.065 | 0.065 | 9.96 𝗑 10^1^ |
| **UNC FA** | 0.04 | 890 | 0.07 | 0.03 | 0.004 | 0.134 | 8.21 𝗑 10^2^ |

| **Total PD & Controls** | ***p*-value** | **Degrees of Freedom** | **Pearson’s Correlation Coefficient *r*** | **Standard Error** | **Lower CI** | **Upper CI** | ***p*_FDR_** |
| --- | --- | --- | --- | --- | --- | --- | --- |
| **Entire WM MD** | 0.00 | 890 | -0.13 | 0.03 | -0.19 | -0.06 | 5.29 𝗑 10^4^ |
| **ACR MD** | 0.00 | 890 | -0.15 | 0.03 | -0.21 | -0.09 | 4.02 𝗑 10^5^ |
| **SCR MD** | 0.00 | 890 | -0.16 | 0.03 | -0.22 | -0.09 | 2.90 𝗑 10^5^ |
| **PCR MD** | 0.05 | 890 | -0.07 | 0.03 | -0.13 | 0.00 | 7.36 𝗑 10^2^ |
| **ALIC MD** | 0.00 | 890 | -0.13 | 0.03 | -0.19 | -0.06 | 5.13 𝗑 10^4^ |
| **PLIC MD** | 0.03 | 890 | -0.07 | 0.03 | -0.14 | -0.01 | 5.37 𝗑 10^2^ |
| **RLIC MD** | 0.07 | 890 | -0.06 | 0.03 | -0.12 | 0.01 | 1.02 𝗑 10^1^ |
| **EC MD** | 0.00 | 890 | -0.13 | 0.03 | -0.20 | -0.07 | 3.43 𝗑 10^4^ |
| **FX MD** | 0.00 | 890 | -0.11 | 0.03 | -0.17 | -0.04 | 3.91 𝗑 10^3^ |
| **FXST MD** | 0.53 | 890 | -0.02 | 0.03 | -0.09 | 0.04 | 5.50 𝗑 10^1^ |
| **PTR MD** | 0.12 | 890 | -0.05 | 0.03 | -0.12 | 0.01 | 1.51 𝗑 10^1^ |
| **GCC MD** | 0.00 | 890 | -0.15 | 0.03 | -0.21 | -0.09 | 4.02 𝗑 10^5^ |
| **BCC MD** | 0.00 | 890 | -0.11 | 0.03 | -0.18 | -0.05 | 1.73 𝗑 10^3^ |
| **SCC MD** | 0.81 | 890 | 0.01 | 0.03 | -0.06 | 0.07 | 8.08 𝗑 10^1^ |
| **CGC MD** | 0.00 | 890 | -0.10 | 0.03 | -0.16 | -0.03 | 8.91 𝗑 10^3^ |
| **CGH MD** | 0.46 | 890 | -0.02 | 0.03 | -0.09 | 0.04 | 5.33 𝗑 10^1^ |
| **CST MD** | 0.53 | 890 | -0.02 | 0.03 | -0.09 | 0.04 | 5.50 𝗑 10^1^ |
| **SFO MD** | 0.00 | 890 | -0.17 | 0.03 | -0.23 | -0.10 | 1.35 𝗑 10^5^ |
| **SLF MD** | 0.00 | 890 | -0.10 | 0.03 | -0.17 | -0.04 | 5.44 𝗑 10^3^ |
| **SS MD** | 0.03 | 890 | -0.07 | 0.03 | -0.14 | -0.01 | 4.88 𝗑 10^2^ |
| **TAP MD** | 0.10 | 890 | -0.05 | 0.03 | -0.12 | 0.01 | 1.32 𝗑 10^1^ |
| **UNC MD** | 0.02 | 890 | -0.08 | 0.03 | -0.14 | -0.01 | 4.45 𝗑 10^2^ |

| **Total PD & Controls** | ***p*-value** | **Degrees of Freedom** | **Pearson’s Correlation Coefficient *r*** | **Standard Error** | **Lower CI** | **Upper CI** | ***p*_FDR_** |
| --- | --- | --- | --- | --- | --- | --- | --- |
| **Entire WM AD** | 0.02 | 890 | -0.08 | 0.033 | -0.146 | -0.016 | 3.75 𝗑 10^2^ |
| **ACR AD** | 0.00 | 890 | -0.14 | 0.033 | -0.208 | -0.079 | 9.56 𝗑 10^5^ |
| **SCR AD** | 0.00 | 890 | -0.16 | 0.033 | -0.220 | -0.091 | 6.53 𝗑 10^5^ |
| **PCR AD** | 0.05 | 890 | -0.06 | 0.033 | -0.129 | 0.001 | 1.21 𝗑 10^1^ |
| **ALIC AD** | 0.00 | 890 | -0.10 | 0.033 | -0.163 | -0.033 | 1.04 𝗑 10^2^ |
| **PLIC AD** | 0.11 | 890 | -0.05 | 0.033 | -0.118 | 0.012 | 1.78 𝗑 10^1^ |
| **RLIC AD** | 0.24 | 890 | -0.04 | 0.033 | -0.105 | 0.026 | 3.51 𝗑 10^1^ |
| **EC AD** | 0.00 | 890 | -0.10 | 0.033 | -0.166 | -0.036 | 9.46 𝗑 10^3^ |
| **FX AD** | 0.00 | 890 | -0.10 | 0.033 | -0.161 | -0.031 | 1.12 𝗑 10^2^ |
| **FXST AD** | 0.42 | 890 | 0.03 | 0.033 | -0.038 | 0.092 | 5.73 𝗑 10^1^ |
| **PTR AD** | 0.70 | 890 | 0.01 | 0.033 | -0.052 | 0.078 | 7.37 𝗑 10^1^ |
| **GCC AD** | 0.00 | 890 | -0.15 | 0.033 | -0.211 | -0.082 | 9.56 𝗑 10^5^ |
| **BCC AD** | 0.00 | 890 | -0.10 | 0.033 | -0.166 | -0.036 | 9.46 𝗑 10^3^ |
| **SCC AD** | 0.51 | 890 | 0.02 | 0.033 | -0.043 | 0.087 | 6.29 𝗑 10^1^ |
| **CGC AD** | 0.09 | 890 | -0.06 | 0.033 | -0.121 | 0.009 | 1.56 𝗑 10^1^ |
| **CGH AD** | 0.70 | 890 | 0.01 | 0.033 | -0.052 | 0.078 | 7.37 𝗑 10^1^ |
| **CST AD** | 0.55 | 890 | 0.02 | 0.033 | -0.045 | 0.085 | 6.35 𝗑 10^1^ |
| **SFO AD** | 0.00 | 890 | -0.14 | 0.033 | -0.208 | -0.079 | 9.56 𝗑 10^5^ |
| **SLF AD** | 0.08 | 890 | -0.06 | 0.033 | -0.125 | 0.006 | 1.39 𝗑 10^1^ |
| **SS AD** | 0.76 | 890 | -0.01 | 0.033 | -0.075 | 0.055 | 7.58 𝗑 10^1^ |
| **TAP AD** | 0.06 | 890 | -0.06 | 0.033 | -0.128 | 0.002 | 1.21 𝗑 10^1^ |
| **UNC AD** | 0.48 | 890 | -0.02 | 0.033 | -0.089 | 0.041 | 6.16 𝗑 10^1^ |

| **Total PD & Controls** | ***p*-value** | **Degrees of Freedom** | **Pearson’s Correlation Coefficient *r*** | **Standard Error** | **Lower CI** | **Upper CI** | ***p*_FDR_** |
| --- | --- | --- | --- | --- | --- | --- | --- |
| **Entire WM RD** | 0.00 | 890 | -0.14 | 0.03 | -0.20 | -0.07 | 2.31 𝗑 10^4^ |
| **ACR RD** | 0.00 | 890 | -0.14 | 0.03 | -0.20 | -0.07 | 2.31 𝗑 10^4^ |
| **SCR RD** | 0.00 | 890 | -0.12 | 0.03 | -0.19 | -0.06 | 6.06 𝗑 10^4^ |
| **PCR RD** | 0.08 | 890 | -0.06 | 0.03 | -0.12 | 0.01 | 1.06 𝗑 10^1^ |
| **ALIC RD** | 0.00 | 890 | -0.13 | 0.03 | -0.19 | -0.07 | 4.15 𝗑 10^4^ |
| **PLIC RD** | 0.04 | 890 | -0.07 | 0.03 | -0.13 | 0.00 | 6.06 𝗑 10^2^ |
| **RLIC RD** | 0.08 | 890 | -0.06 | 0.03 | -0.12 | 0.01 | 1.06 𝗑 10^1^ |
| **EC RD** | 0.00 | 890 | -0.13 | 0.03 | -0.20 | -0.07 | 3.21 𝗑 10^4^ |
| **FX RD** | 0.00 | 890 | -0.11 | 0.03 | -0.17 | -0.04 | 3.70 𝗑 10^3^ |
| **FXST RD** | 0.11 | 890 | -0.05 | 0.03 | -0.12 | 0.01 | 1.36 𝗑 10^1^ |
| **PTR RD** | 0.01 | 890 | -0.08 | 0.03 | -0.15 | -0.02 | 2.10 𝗑 10^2^ |
| **GCC RD** | 0.00 | 890 | -0.13 | 0.03 | -0.19 | -0.06 | 4.21 𝗑 10^4^ |
| **BCC RD** | 0.00 | 890 | -0.10 | 0.03 | -0.17 | -0.04 | 3.77 𝗑 10^3^ |
| **SCC RD** | 0.72 | 890 | -0.01 | 0.03 | -0.08 | 0.05 | 7.23 𝗑 10^1^ |
| **CGC RD** | 0.01 | 890 | -0.09 | 0.03 | -0.16 | -0.03 | 1.08 𝗑 10^2^ |
| **CGH RD** | 0.14 | 890 | -0.05 | 0.03 | -0.11 | 0.02 | 1.57 𝗑 10^1^ |
| **CST RD** | 0.14 | 890 | -0.05 | 0.03 | -0.11 | 0.02 | 1.57 𝗑 10^1^ |
| **SFO RD** | 0.00 | 890 | -0.16 | 0.03 | -0.22 | -0.10 | 3.64 𝗑 10^5^ |
| **SLF RD** | 0.00 | 890 | -0.11 | 0.03 | -0.17 | -0.04 | 2.91 𝗑 10^3^ |
| **SS RD** | 0.00 | 890 | -0.10 | 0.03 | -0.16 | -0.03 | 8.43 𝗑 10^3^ |
| **TAP RD** | 0.19 | 890 | -0.04 | 0.03 | -0.11 | 0.02 | 1.97 𝗑 10^1^ |
| **UNC RD** | 0.01 | 890 | -0.09 | 0.03 | -0.15 | -0.02 | 1.67 𝗑 10^2^ |

##### 2.5.2 PD participants: Partial correlation between DTI metrics and MDS-UPDRS Part-III (OFF) scores

| **Total PD & Controls** | ***p*-value** | **Degrees of Freedom** | **Pearson’s Correlation Coefficient *r*** | **Standard Error** | **Lower CI** | **Upper CI** | ***p*_FDR_** |
| --- | --- | --- | --- | --- | --- | --- | --- |
| **Entire WM FA** | 0.00 | 526 | -0.15 | 0.04 | -0.24 | -0.07 | 3.05 𝗑 10^3^ |
| **ACR FA** | 0.01 | 526 | -0.12 | 0.04 | -0.20 | -0.03 | 2.01 𝗑 10^2^ |
| **SCR FA** | 0.02 | 526 | -0.10 | 0.04 | -0.18 | -0.01 | 3.73 𝗑 10^2^ |
| **PCR FA** | 0.02 | 526 | -0.10 | 0.04 | -0.19 | -0.02 | 3.32 𝗑 10^2^ |
| **ALIC FA** | 0.10 | 526 | -0.07 | 0.04 | -0.16 | 0.01 | 1.18 𝗑 10^1^ |
| **PLIC FA** | 0.14 | 526 | -0.06 | 0.04 | -0.15 | 0.02 | 1.55 𝗑 10^1^ |
| **RLIC FA** | 0.03 | 526 | -0.10 | 0.04 | -0.18 | -0.01 | 3.81 𝗑 10^2^ |
| **EC FA** | 0.00 | 526 | -0.15 | 0.04 | -0.23 | -0.07 | 3.05 𝗑 10^3^ |
| **FX FA** | 0.00 | 526 | -0.15 | 0.04 | -0.23 | -0.06 | 3.05 𝗑 10^3^ |
| **FXST FA** | 0.11 | 526 | -0.07 | 0.04 | -0.15 | 0.01 | 1.25 𝗑 10^1^ |
| **PTR FA** | 0.00 | 526 | -0.15 | 0.04 | -0.23 | -0.07 | 3.05 𝗑 10^3^ |
| **GCC FA** | 0.00 | 526 | -0.16 | 0.04 | -0.24 | -0.07 | 3.05 𝗑 10^3^ |
| **BCC FA** | 0.00 | 526 | -0.13 | 0.04 | -0.22 | -0.05 | 8.08 𝗑 10^3^ |
| **SCC FA** | 0.02 | 526 | -0.10 | 0.04 | -0.19 | -0.02 | 3.32 𝗑 10^2^ |
| **CGC FA** | 0.00 | 526 | -0.13 | 0.04 | -0.21 | -0.05 | 8.12 𝗑 10^3^ |
| **CGH FA** | 0.79 | 526 | -0.01 | 0.04 | -0.10 | 0.07 | 7.90 𝗑 10^1^ |
| **CST FA** | 0.74 | 526 | -0.01 | 0.04 | -0.10 | 0.07 | 7.77 𝗑 10^1^ |
| **SFO FA** | 0.05 | 526 | -0.09 | 0.04 | -0.17 | 0.00 | 5.89 𝗑 10^2^ |
| **SLF FA** | 0.04 | 526 | -0.09 | 0.04 | -0.18 | -0.01 | 5.00 𝗑 10^2^ |
| **SS FA** | 0.01 | 526 | -0.11 | 0.04 | -0.19 | -0.03 | 2.77 𝗑 10^2^ |
| **TAP FA** | 0.02 | 526 | -0.10 | 0.04 | -0.19 | -0.02 | 3.32 𝗑 10^2^ |
| **UNC FA** | 0.02 | 526 | -0.10 | 0.04 | -0.19 | -0.02 | 3.32 𝗑 10^2^ |

| **Total PD & Controls** | ***p*-value** | **Degrees of Freedom** | **Pearson’s Correlation Coefficient *r*** | **Standard Error** | **Lower CI** | **Upper CI** | ***p*_FDR_** |
| --- | --- | --- | --- | --- | --- | --- | --- |
| **Entire WM MD** | 0.70 | 526 | 0.02 | 0.04 | -0.07 | 0.10 | 9.65 𝗑 10^1^ |
| **ACR MD** | 0.08 | 526 | 0.08 | 0.04 | -0.01 | 0.16 | 3.33 𝗑 10^1^ |
| **SCR MD** | 0.84 | 526 | -0.01 | 0.04 | -0.09 | 0.08 | 9.70 𝗑 10^1^ |
| **PCR MD** | 0.66 | 526 | 0.02 | 0.04 | -0.07 | 0.10 | 9.65 𝗑 10^1^ |
| **ALIC MD** | 0.31 | 526 | -0.04 | 0.04 | -0.13 | 0.04 | 6.89 𝗑 10^1^ |
| **PLIC MD** | 0.05 | 526 | -0.08 | 0.04 | -0.17 | 0.00 | 3.33 𝗑 10^1^ |
| **RLIC MD** | 0.83 | 526 | -0.01 | 0.04 | -0.09 | 0.08 | 9.70 𝗑 10^1^ |
| **EC MD** | 0.61 | 526 | 0.02 | 0.04 | -0.06 | 0.11 | 9.65 𝗑 10^1^ |
| **FX MD** | 0.06 | 526 | 0.08 | 0.04 | 0.00 | 0.16 | 3.33 𝗑 10^1^ |
| **FXST MD** | 0.97 | 526 | 0.00 | 0.04 | -0.08 | 0.09 | 9.70 𝗑 10^1^ |
| **PTR MD** | 0.92 | 526 | 0.00 | 0.04 | -0.08 | 0.09 | 9.70 𝗑 10^1^ |
| **GCC MD** | 0.15 | 526 | 0.06 | 0.04 | -0.02 | 0.15 | 4.26 𝗑 10^1^ |
| **BCC MD** | 0.96 | 526 | 0.00 | 0.04 | -0.09 | 0.08 | 9.70 𝗑 10^1^ |
| **SCC MD** | 0.07 | 526 | -0.08 | 0.04 | -0.16 | 0.01 | 3.33 𝗑 10^1^ |
| **CGC MD** | 0.24 | 526 | -0.05 | 0.04 | -0.14 | 0.03 | 5.94 𝗑 10^1^ |
| **CGH MD** | 0.12 | 526 | -0.07 | 0.04 | -0.15 | 0.02 | 4.24 𝗑 10^1^ |
| **CST MD** | 0.02 | 526 | -0.10 | 0.04 | -0.18 | -0.02 | 3.33 𝗑 10^1^ |
| **SFO MD** | 0.42 | 526 | 0.04 | 0.04 | -0.05 | 0.12 | 8.39 𝗑 10^1^ |
| **SLF MD** | 0.92 | 526 | 0.00 | 0.04 | -0.08 | 0.09 | 9.70 𝗑 10^1^ |
| **SS MD** | 0.62 | 526 | 0.02 | 0.04 | -0.06 | 0.11 | 9.65 𝗑 10^1^ |
| **TAP MD** | 0.14 | 526 | 0.06 | 0.04 | -0.02 | 0.15 | 4.26 𝗑 10^1^ |
| **UNC MD** | 0.48 | 526 | -0.03 | 0.04 | -0.12 | 0.05 | 8.76 𝗑 10^1^ |

| **Total PD & Controls** | ***p*-value** | **Degrees of Freedom** | **Pearson’s Correlation Coefficient *r*** | **Standard Error** | **Lower CI** | **Upper CI** | ***p*_FDR_** |
| --- | --- | --- | --- | --- | --- | --- | --- |
| **Entire WM AD** | 0.02 | 526 | -0.10 | 0.04 | -0.18 | -0.01 | 7.76 𝗑 10^2^ |
| **ACR AD** | 0.89 | 526 | 0.01 | 0.04 | -0.08 | 0.09 | 8.90 𝗑 10^1^ |
| **SCR AD** | 0.21 | 526 | -0.05 | 0.04 | -0.14 | 0.03 | 3.54 𝗑 10^1^ |
| **PCR AD** | 0.39 | 526 | -0.04 | 0.04 | -0.12 | 0.05 | 4.71 𝗑 10^1^ |
| **ALIC AD** | 0.06 | 526 | -0.08 | 0.04 | -0.17 | 0.00 | 1.38 𝗑 10^1^ |
| **PLIC AD** | 0.02 | 526 | -0.10 | 0.04 | -0.19 | -0.02 | 6.96 𝗑 10^2^ |
| **RLIC AD** | 0.17 | 526 | -0.06 | 0.04 | -0.14 | 0.02 | 3.03 𝗑 10^1^ |
| **EC AD** | 0.08 | 526 | -0.08 | 0.04 | -0.16 | 0.01 | 1.62 𝗑 10^1^ |
| **FX AD** | 0.37 | 526 | 0.04 | 0.04 | -0.05 | 0.12 | 4.71 𝗑 10^1^ |
| **FXST AD** | 0.28 | 526 | -0.05 | 0.04 | -0.13 | 0.04 | 4.18 𝗑 10^1^ |
| **PTR AD** | 0.02 | 526 | -0.10 | 0.04 | -0.18 | -0.01 | 7.76 𝗑 10^2^ |
| **GCC AD** | 0.37 | 526 | -0.04 | 0.04 | -0.12 | 0.05 | 4.71 𝗑 10^1^ |
| **BCC AD** | 0.03 | 526 | -0.09 | 0.04 | -0.18 | -0.01 | 8.02 𝗑 10^2^ |
| **SCC AD** | 0.00 | 526 | -0.12 | 0.04 | -0.21 | -0.04 | 4.11 𝗑 10^2^ |
| **CGC AD** | 0.00 | 526 | -0.12 | 0.04 | -0.21 | -0.04 | 4.11 𝗑 10^2^ |
| **CGH AD** | 0.04 | 526 | -0.09 | 0.04 | -0.18 | -0.01 | 8.91 𝗑 10^2^ |
| **CST AD** | 0.01 | 526 | -0.12 | 0.04 | -0.20 | -0.04 | 4.11 𝗑 10^2^ |
| **SFO AD** | 0.81 | 526 | -0.01 | 0.04 | -0.10 | 0.07 | 8.88 𝗑 10^1^ |
| **SLF AD** | 0.29 | 526 | -0.05 | 0.04 | -0.13 | 0.04 | 4.18 𝗑 10^1^ |
| **SS AD** | 0.41 | 526 | -0.04 | 0.04 | -0.12 | 0.05 | 4.71 𝗑 10^1^ |
| **TAP AD** | 0.88 | 526 | 0.01 | 0.04 | -0.08 | 0.09 | 8.90 𝗑 10^1^ |
| **UNC AD** | 0.01 | 526 | -0.11 | 0.04 | -0.19 | -0.02 | 6.96 𝗑 10^2^ |

| **Total PD & Controls** | ***p*-value** | **Degrees of Freedom** | **Pearson’s Correlation Coefficient *r*** | **Standard Error** | **Lower CI** | **Upper CI** | ***p*_FDR_** |
| --- | --- | --- | --- | --- | --- | --- | --- |
| **Entire WM RD** | 0.02 | 526 | 0.10 | 0.04 | 0.02 | 0.19 | 9.89 𝗑 10^2^ |
| **ACR RD** | 0.01 | 526 | 0.11 | 0.04 | 0.03 | 0.20 | 9.89 𝗑 10^2^ |
| **SCR RD** | 0.37 | 526 | 0.04 | 0.04 | -0.05 | 0.12 | 4.27 𝗑 10^1^ |
| **PCR RD** | 0.18 | 526 | 0.06 | 0.04 | -0.03 | 0.14 | 3.23 𝗑 10^1^ |
| **ALIC RD** | 0.86 | 526 | 0.01 | 0.04 | -0.08 | 0.09 | 9.01 𝗑 10^1^ |
| **PLIC RD** | 0.90 | 526 | -0.01 | 0.04 | -0.09 | 0.08 | 9.01 𝗑 10^1^ |
| **RLIC RD** | 0.30 | 526 | 0.05 | 0.04 | -0.04 | 0.13 | 4.13 𝗑 10^1^ |
| **EC RD** | 0.04 | 526 | 0.09 | 0.04 | 0.01 | 0.18 | 9.89 𝗑 10^2^ |
| **FX RD** | 0.03 | 526 | 0.09 | 0.04 | 0.01 | 0.18 | 9.89 𝗑 10^2^ |
| **FXST RD** | 0.22 | 526 | 0.05 | 0.04 | -0.03 | 0.14 | 3.41 𝗑 10^1^ |
| **PTR RD** | 0.03 | 526 | 0.09 | 0.04 | 0.01 | 0.18 | 9.89 𝗑 10^2^ |
| **GCC RD** | 0.00 | 526 | 0.14 | 0.04 | 0.05 | 0.22 | 3.38 𝗑 10^2^ |
| **BCC RD** | 0.03 | 526 | 0.09 | 0.04 | 0.01 | 0.18 | 9.89 𝗑 10^2^ |
| **SCC RD** | 0.23 | 526 | 0.05 | 0.04 | -0.03 | 0.14 | 3.41 𝗑 10^1^ |
| **CGC RD** | 0.16 | 526 | 0.06 | 0.04 | -0.02 | 0.14 | 3.23 𝗑 10^1^ |
| **CGH RD** | 0.55 | 526 | -0.03 | 0.04 | -0.11 | 0.06 | 6.07 𝗑 10^1^ |
| **CST RD** | 0.22 | 526 | -0.05 | 0.04 | -0.14 | 0.03 | 3.41 𝗑 10^1^ |
| **SFO RD** | 0.14 | 526 | 0.06 | 0.04 | -0.02 | 0.15 | 3.01 𝗑 10^1^ |
| **SLF RD** | 0.33 | 526 | 0.04 | 0.04 | -0.04 | 0.13 | 4.26 𝗑 10^1^ |
| **SS RD** | 0.11 | 526 | 0.07 | 0.04 | -0.02 | 0.15 | 2.80 𝗑 10^1^ |
| **TAP RD** | 0.04 | 526 | 0.09 | 0.04 | 0.01 | 0.18 | 9.89 𝗑 10^2^ |
| **UNC RD** | 0.35 | 526 | 0.04 | 0.04 | -0.04 | 0.12 | 4.27E 𝗑 10^1^ |

**References:**

Goetz, Christopher G, Glenn T Stebbins, and Barbara C Tilley. 2012. “Calibration of Unified Parkinson’s Disease Rating Scale Scores to Movement Disorder Society‐unified Parkinson’s Disease Rating Scale Scores.” *Movement Disorders* 27 (10): 1239–42.
